# Hedged allocation of committed measles doses clears more US schools of transmission risk

**DOI:** 10.64898/2026.08.20.26360961

**Authors:** Laura W. Alexander, Aakash Pandey, Nathaniel Hupert, Emily A. Serman, Lior Rennert, Ana I. Bento

## Abstract

US measles elimination is in question, and the protective doses are already committed; which schools receive them first is unresolved. Of 36,031 US schools, 12,196 hold enough susceptible children to sustain transmission once a case arrives. One 77,292-dose budget would clear 2,497 of them under a lightly hedged rule against 1,511 under the lowest-coverage-first rule now in use, leaving 112,733 fewer children where a chain can persist at no additional dose. The gain comes from finishing schools near the herd-immunity threshold rather than starting schools far below it. Allocation theory’s point-optimal form of that rule assumes every school is equally likely to see a case; pre-outbreak records from a 2025-26 South Carolina outbreak show exposed schools over-represented 5.0-fold in susceptible headcount, unlike enrollment. Under non-uniform risk, whose strength one outbreak cannot identify, that optimum is the least robust of seven rules, while the hedge holds roughly 90% of attainable benefit.

## Introduction

US measles elimination status is in question. The Region of the Americas lost elimination status in November 2025, and Canada lost its own status the same year, and the United States has been under review since 2026^1^. Whichever way that review resolves, the allocation question it raises does not resolve with it: elimination requires twelve months without continuous transmission of a single chain^2^, and 2026 has already produced the largest national case count in thirty-five years, with close to 3000 confirmed cases across 47 jurisdictions by August of that year^3^. Transmission is regionally sustained across the Americas^4–6^. Kindergarten measles-mumps-rubella (MMR) vaccination coverage has fallen every year since 2019-20, from 95.2% to 92.5%^7–9^, part of a broader post-pandemic disruption to routine immunization^10^. The national figure understates the problem: transmission is determined by how unvaccinated children cluster in the settings a case reaches, such as individual schools, not by the average coverage of a state or county^11–13^. Coverage itself is slow to move, since reversing the exemption trends behind it takes years, whereas the order in which an already-committed dose budget is spent can change within a single procurement cycle. Allocation is therefore one of the few levers that acts on the timescale of a single school year, and a school-level coverage ranking retains only about half of its lowest decile after one year, so the window in which it acts is the year in which the coverage was measured.

The policy response follows a rule so intuitive it is rarely stated as a choice: find where coverage is lowest and raise it. Federal and state programs rank jurisdictions by coverage, and outbreak response concentrates on the lowest-coverage communities^14,15^. The rule is defensible on equity grounds and aligns with the existing surveillance infrastructure, which reports coverage percentage as its headline metric^16,17^. It sits on one side of a tension familiar from early COVID-19 vaccine rollout, a debate that has proceeded without an estimate of what the choice costs. Supplying one for measles is this paper’s purpose.

Lowest-coverage-first is not dose-optimal: it does not avert the most infections a fixed number of doses could avert, and the reason is structural rather than practical. A handful of doses at a school at 50% coverage leaves the school far short of the threshold where herd protection begins, so almost every dose buys individual immunity alone, whereas the same handful at 90% can carry it over that threshold and protect the unvaccinated children who remain. Finishing a job that can be finished buys more than starting one that cannot.

Measles vaccination reduces cases by two mechanisms: it directly defends immunized individuals from infection, and it indirectly averts infections in unvaccinated individuals by reducing the overall infection pressure they face, the effect known as herd immunity. Formally, the indirect benefit of vaccinating a population, the infections per vaccinated individual averted beyond the recipients, is single-peaked in its coverage: nearly flat at low coverage, steep near the herd-immunity threshold, maximal at that threshold and falling above it^18^, and so is convex, curving upward, below an inflection point. Total infections prevented, which adds the directly immunized recipients, rises close to one-for-one from the origin and shares the same inflection (Supplementary Fig. 1, Supplementary Table 1); the classification is insensitive to the assumed introduction size and across the parameter grid (Supplementary Figs. 3 and 4). The indirect protection component is the one allocation can move, because every dose immunizes the same number of recipients wherever it lands. In the convex region the marginal return to a dose is increasing, so the same dose at a better-covered population indirectly averts more infections. Duijzer and colleagues proved that a dose-optimal allocation must therefore avoid spreading doses thinly into the convex region, and that treating the lowest-coverage populations first can always be outperformed^19^. The result is not isolated: optimal prophylactic distribution is generally inequitable, equalizing strategies can be improved upon in segregated populations, an influenza heuristic reached the same reversal independently, and at high reproductive numbers the optimum can prioritize groups least likely to be infected^20–23^. This literature has been established analytically on synthetic or aggregate populations and lacks a test against population records. Whether real populations sit in the convex region, how many, and whether the gap between the optimal and intuitive rules matters are open empirical questions. Real programs are hybrids of the two approaches, so we treat them as endpoints of a range and locate the question inside it.

Getting the ordering right matters for measles specifically. Its basic reproduction number near 15 puts the herd-immunity threshold near 95%, and the inflection of the herd-effect function sits just below it: a school at 90% coverage, which no surveillance system would flag, can still be convex^24^. Coverage is also reported at the wrong scale: aggregation conceals the fine-scale clustering of non-vaccination that governs outbreak risk rather than the mean^14,15,25–29^, transmission scales with the number of susceptible children rather than the percentage vaccinated, and measles dynamics are spatially structured at every scale examined11,30-32.

We take the descriptive groundwork as established. Our previous work^11–13^ assembled a multiscale US vaccination database, showed risk resolves at the school scale, and validated the school-level *R_v_* used here^12^ (reproduced exactly, Supplementary Table 2); this paper inherits that database and transmission formalism unchanged (Supplementary Note 6) and builds a prescriptive layer on top, asking which schools a fixed dose budget should be sent to and whether that answer survives not knowing where an outbreak begins. It does not: the efficiency gain rests on an assumption the theorem makes, and US surveillance cannot check that every school faces the same introduction probability.

We assemble coverage for 36,031 schools in 27 states and the District of Columbia, classify each against the herd-effect inflection (H), compare a marginal-efficiency rule with lowest-coverage-first at identical budgets, validate the risk model against the 2025 to 2026 outbreaks including a temporally external school-scale test, price the difference against published costings^33^. Of seven state outbreak records assessed, two carry an investigation register with pre-outbreak coverage and a comparison set. Because the strength of that risk scaling cannot be estimated from existing data, we choose the rule whose worst case across its plausible range is least bad, the minimax-regret criterion of decision theory^34–36^. That criterion is standard for choosing between actions when a parameter is only partially identified; what has been missing for this allocation problem is an uncertainty set for introduction risk built from outbreak records rather than assumed, which these registers supply.

The result is a reversal, and it is not specific to measles: wherever a targeting variable is measured well and the variable governing exposure is measured badly, the point-estimate optimum is the fragile choice. The theoretically optimal rule is the least robust of the seven we evaluate, and that ordering holds across three structurally different constructions of the uncertainty set, while a light hedge on measured risk recovers most of the attainable benefit from the two columns every state already publishes.

## Results

### Most susceptible children attend schools where spending order matters

We assembled school-level kindergarten MMR coverage from state open-data releases spanning 2013-14 to 2024-25, giving 224,353 school-years across 33 states. Five states were excluded for reporting an implausible share of schools at exactly 100% coverage, reflecting censoring rather than saturation, from 44.3% of schools in North Dakota to 89.5% in Maine (Supplementary Fig. 2, Supplementary Note 6, Supplementary Table 3). The retained latest-year set comprises 36,031 schools in 28 jurisdictions (27 states and DC) with 309,167 susceptible kindergarteners and an enrollment-weighted mean coverage of 93.1%. The latest available year is 2024-25 for 21 states and 2023-24 for seven, a one-year spread. At measles parameters and *i*_0_ = 10^−3^ the herd-effect function H inflects from convex to concave at *f̄*= 94.4% coverage, with dose-optimal fraction *f̄* = 96.2% (Methods, Supplementary Tables 5 and 6).

Classifying every school against the inflection of the herd effect function H, *f̄*= 94.4%, not the dose-optimal *f̄* = 96.2%, gives the central result (Fig. 1a, b). 16,589 schools (46.0%) are in the convex region, below the inflection where the marginal return to a dose is still increasing, and they hold 234,259 susceptible kindergarteners, 75.8% of the total. That classification is conditional on the assumed introduction; it is not a statement about which schools lack herd immunity. Concave-region schools (19,442, 54.0%) outnumber convex but hold only 74,908 susceptibles (24.2%).

**Fig. 1.**
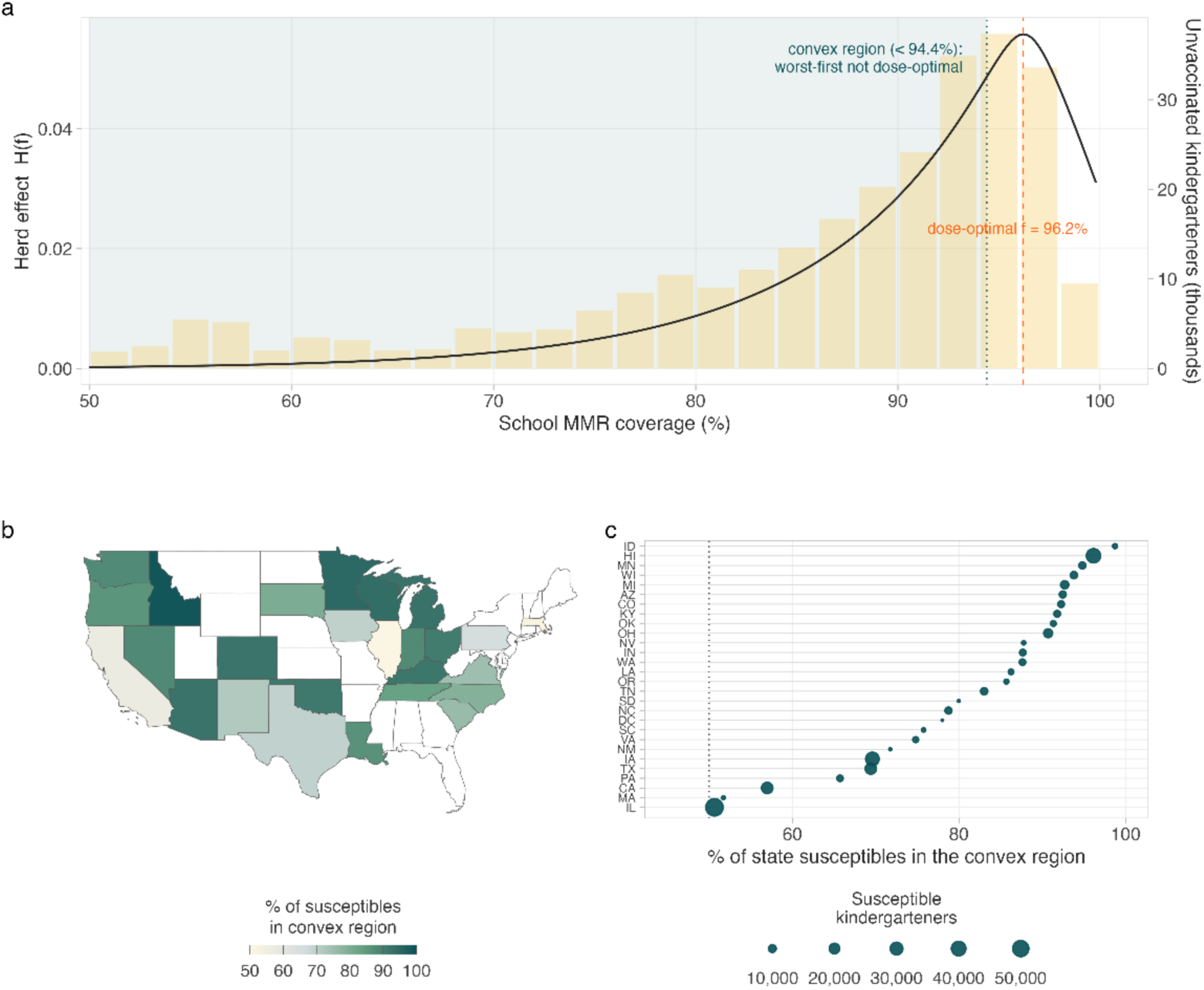
Under uniform introduction risk, most susceptible children attend schools where lowest-coverage-first is not optimal. **a**, The herd-effect function *H*(*f*) (black curve, left axis) against the distribution of unvaccinated kindergarteners by school coverage (bars, right axis). Shading marks the convex region below the inflection *f̄* = 94.4%; the dashed line marks the dose-optimal target *f̄* = 96.2%. 75.8% of national susceptibles fall in the shaded region. **b**, Percentage of each state’s susceptible kindergarteners attending schools in the convex region (coverage below *f̄*). All 28 reporting jurisdictions exceed 50%. White indicates states excluded for implausible reporting or with no school-level data; Hawaii (96%) and Alaska (no data) are not shown. **c**, Every state ranked by convex share, point area proportional to the number of susceptible kindergarteners. The dotted line marks 50%. n = 36,031 schools across 28 jurisdictions.

Replacing *i*_0_ = 10^−3^ with a single index case per school makes the inflection school-specific (88.4% to 95.4%) and moves the convex share to 68.5% of susceptibles in 33.0% of schools, 25 rather than 28 jurisdictions above half (Fig. 2, Supplementary Note 1, Supplementary Fig. 27a, Supplementary Table 7). The majority claim survives, the exact share does not. The asymmetry arises because low-coverage schools contribute disproportionately to the susceptible pool while being a minority of institutions. Under the fixed specification the share of susceptibles attending convex-region schools exceeds half in all 28 jurisdictions, ranging from 50.6% (Illinois) to 98.7% (Idaho) (Fig. 1b,c). The share is robust to sample and parameters: leave-one-state-out gives 72.9-81.6%, and across the grid of *R*_0_ and initial infected fraction it stays between 54.0% and 89.0% (Supplementary Note 9, Supplementary Tables 3, 8 and 9, Supplementary Figs. 3 and 4).

**Fig. 2.**
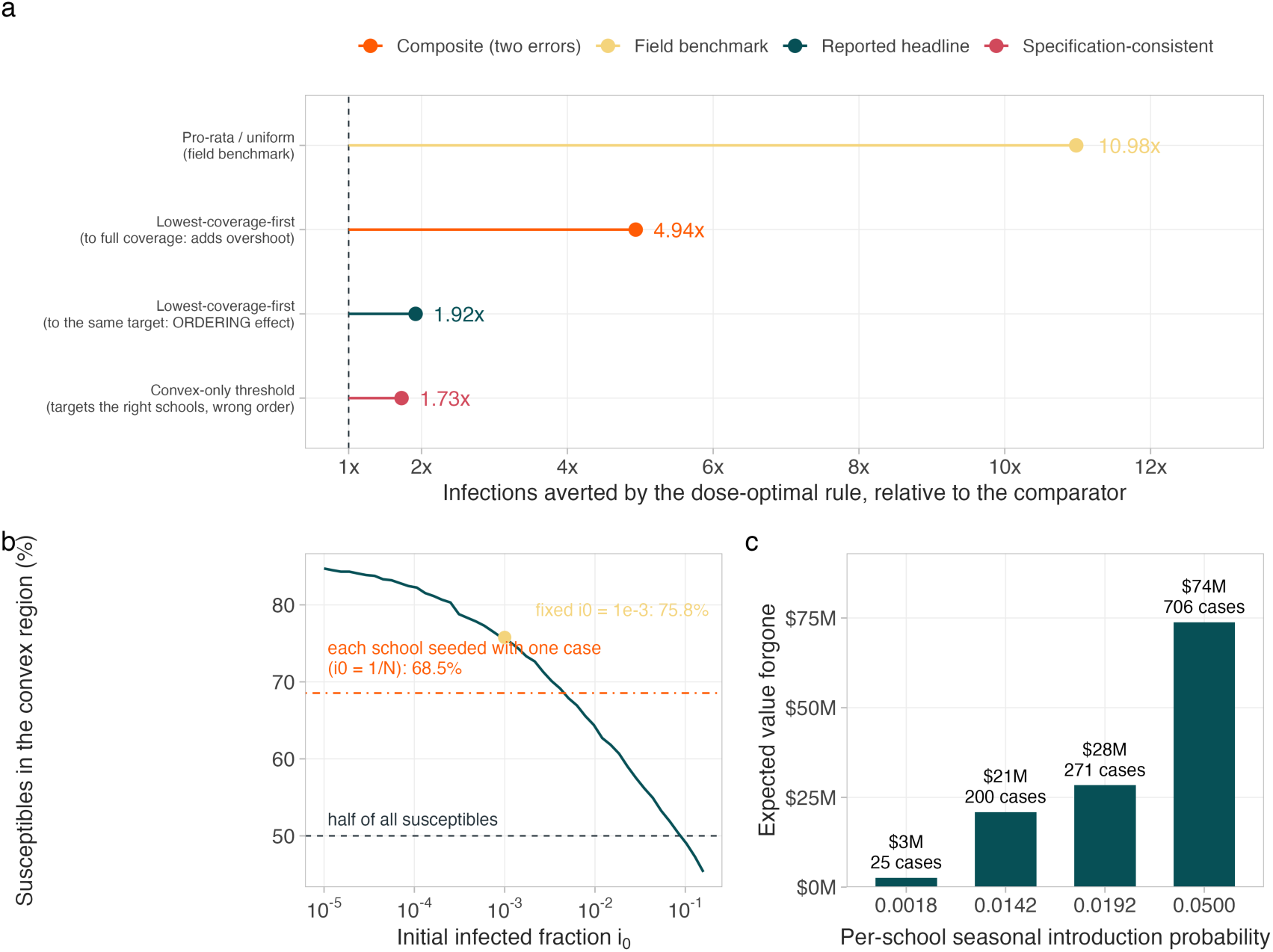
The size of the allocation gain is set by the comparator, and the economics is an expectation. **a**, Infections averted by the dose-optimal rule relative to four baselines, at an identical 77,292-dose budget with each state spending its own 25% and the same optimizer in every row. Lowest-coverage-first taken to the same per-school target isolates the effect of ordering (1.92×) and is the figure reported throughout; taken to full coverage it adds a separable stopping-rule error (4.94×); the convex-only threshold rule (1.73×) already knows where the inflection is and is the more demanding test; pro-rata allocation (10.98×) is the benchmark used in recent allocation work. **b**, Percentage of national susceptibles in the convex region against the assumed initial infected fraction *i_0_.* The point marks the fixed *i_0_* = 10⁻³ used throughout (75.8%); the dot-dash line marks seeding each school with a single index case*, i_0_*= 1/N_j_ (68.5%). The median school holds 69 students. The two specifications are the same deterministic herd effect H evaluated at different *i_0_*, not two functions: fixing the seed fraction gives one inflection common to all schools, while one index case per school makes the fraction, and so the inflection, school-specific. This panel plots the consequence for the convex share; Supplementary Fig. 27a plots the underlying curves. **c**, Expected value forgone by coverage-rank allocation under the societal cost per case, at four per-school seasonal introduction probabilities: three derived from the outbreak record and a fourth pessimistic upper anchor (Supplementary Table 20). Labels give expected excess infections. The fold-gain in a is invariant to this probability because it cancels in the ratio-a per-school probability is constant across schools and so cancels exactly; an expectation scaling with enrollment does not.

### Ordering a fixed dose budget by marginal return doubles indirect protection

We compared two rules at an identical budget of 77,292 doses, 25% of the pool of susceptible individuals. Lowest-coverage-first sorts by coverage and brings the worst up to *f̄*; the dose-optimal rule ranks by the average herd benefit per dose of that move^19^, its marginal efficiency. They fund largely different sets, overlapping in only 582 of the 12,160 and 3,371 schools each funds (Fig. 3a): the intuitive rule spends 62.3% of its budget below 75% coverage and none above the inflection, while the dose-optimal rule concentrates 76.4% of its budget on the 85-94.4% band and sends 2,564 doses (3.3%) to schools already above *f̄*.

**Fig. 3.**
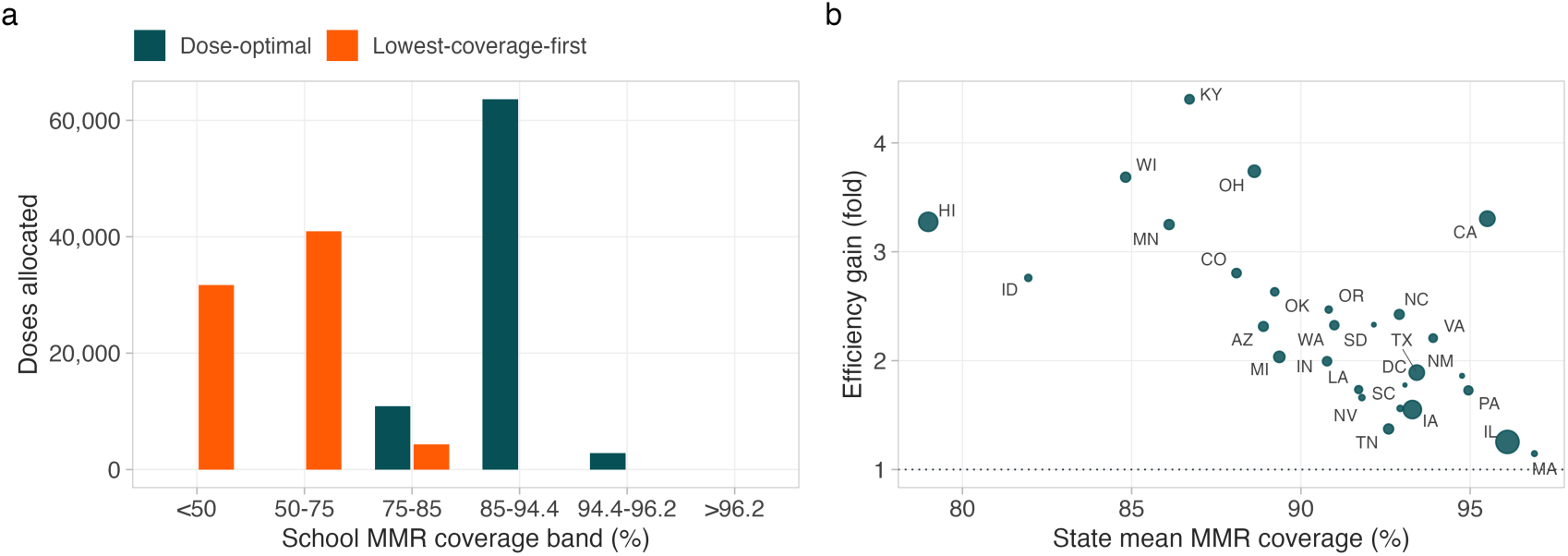
Dose-optimal allocation funds a different set of schools. **a**, Doses allocated by school coverage band under the two rules, at a national budget of 77,292 doses (25% of susceptibles). The lowest-coverage-first rule spends 62.3% of its budget below 75% and none above *f̄*; the dose-optimal rule concentrates on the 85-94.4% band and directs 3.3% of the budget above *f̄*. **b**, Efficiency gain (fold increase in infections averted) against state mean coverage, point area proportional to susceptible pool. Gains are largest where coverage is lowest (Spearman ρ = −0.70, n = 28 jurisdictions).

**Fig. 4.**
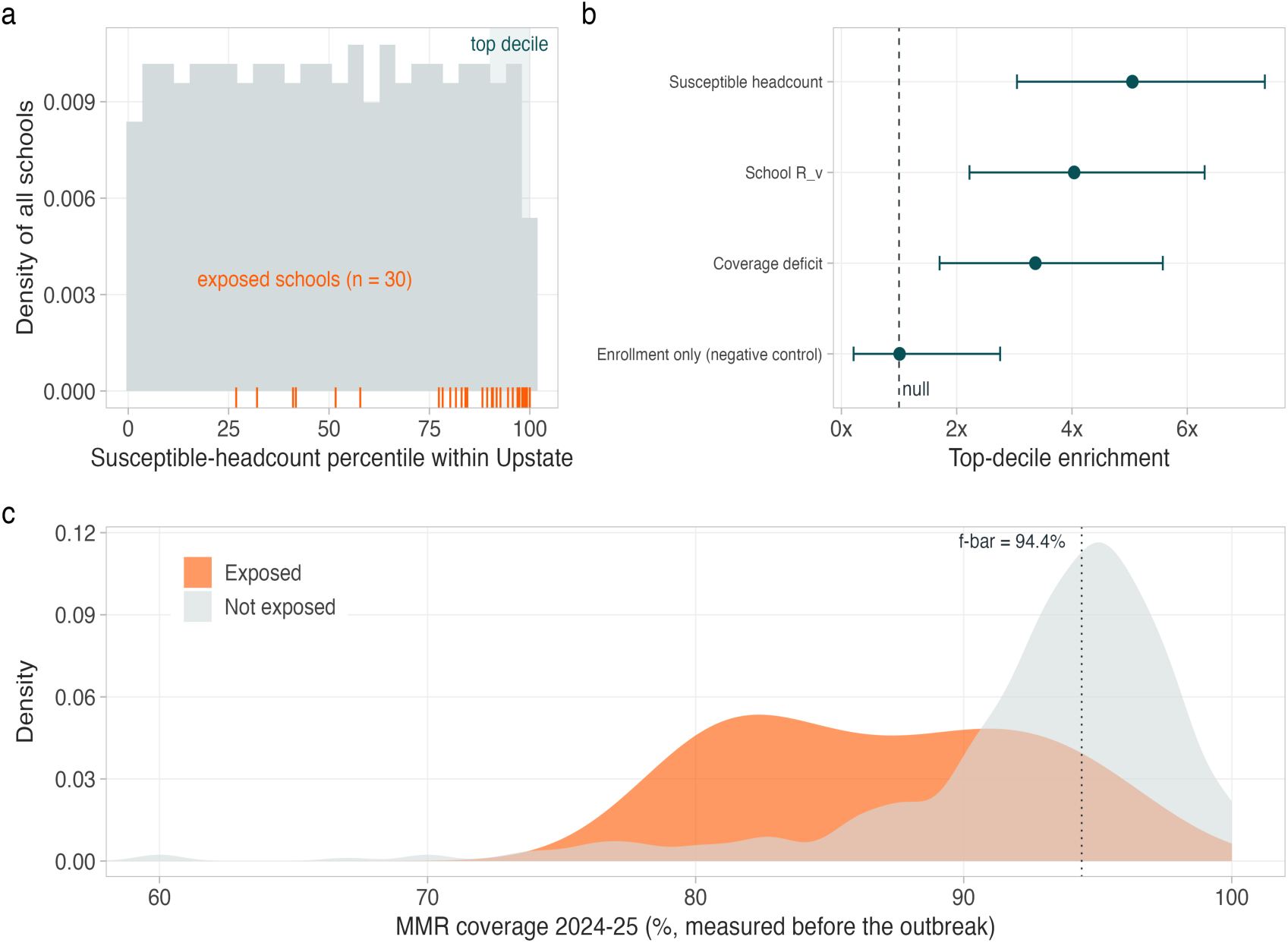
A temporally external test at school scale, where school-level vaccination records are available. South Carolina, 2025-26 Upstate outbreak. Coverage is measured in 2024-25, the school year before exposure. All figures are within the Upstate region, where all 30 exposed schools fall; n = 424 schools with pre-outbreak coverage, 30 exposed. **a**, Distribution of all Upstate schools by susceptible-headcount percentile (bars), with exposed schools marked as a rug. Shading marks the top decile, which contains 15 of the 30 exposed schools. **b**, Over-representation of exposed schools in the top decile for three risk metrics and one negative control, with 95% Clopper-Pearson intervals on the top-decile exposure proportion, rescaled by the region-wide exposure rate held fixed, so they do not propagate uncertainty in that rate; quoted p values are one-sided Fisher exact tests on the corresponding 2 x 2 table. Enrollment used alone does not discriminate (1.01×, p = 0.59), so the result is not an artifact of school size. **c**, Pre-outbreak coverage distributions for exposed and unexposed schools. The dotted line marks *f̄*= 94.4%. Exposed schools averaged 84.7% against 92.1%.

The dose-optimal rule averts 29,479 infections against 15,366 for the intuitive rule, a 1.92-fold gain of indirect protection at identical cost. That is the ceiling of what ordering alone can buy, and it holds only under the uniform-risk premise the theorem assumes; we test that premise below and it does not survive. Every state shows the effect, from 1.15-fold (Massachusetts) to 4.40-fold (Kentucky), median 2.26 (Fig. 3b, Supplementary Table 10); per-school allocations are in Supplementary Table 11. The gain is larger where coverage is lower (Spearman ρ = −0.70), the opposite of a diminishing-returns intuition.

How large that gain looks depends on the comparator, so we report the full ladder rather than the most favorable rung (Fig. 2, Supplementary Table 12). We report 1.92-fold throughout because it isolates ordering: both rules bring schools to the same per-school target and differ only in which schools they fund. A convex-region rule that already knows where the inflection is, the more demanding test, gives 1.73-fold; weaker comparators give larger numbers for reasons unrelated to ordering: lowest-coverage-first taken to full coverage overshoots *f̄* = 96.2% and gives 4.94-fold, and pro-rata allocation gives 10.98-fold; the decomposition of both is in Supplementary Note 2 (Supplementary Fig. 5, Supplementary Tables 12 and 13)^37^.

The allocation result does not depend on the spatial risk model, though it does depend on the final-size function defining the objective (Supplementary Note 2, Supplementary Table 14). Two checks show the theory rather than the classification does the work. Scored on a quantity the optimizer does not maximize, schools left above *R_v_* = 1, the dose-optimal rule still wins in 26 of 28 jurisdictions (22.1% versus 12.4% cleared), and against a convex-region rule ordering by pool size it averts 29,479 against 17,084 (Supplementary Figs. 3c, d, Supplementary Tables 12, 15 and 16).

### The rule choice changes how many schools can sustain a transmission chain

What matters for sustained transmission is how many settings can support a chain: of 36,031 schools, 12,196 have a school-level reproduction number above one, meaning transmission can persist there once a case arrives (*R_v_* > 1) under current coverage. At the 25% budget the dose-optimal rule leaves 9,498 such schools against 10,685 under lowest-coverage-first, clearing 1,187 more and removing 153,555 more children from schools above that threshold (Supplementary Fig. 6, Supplementary Table 17). Neither rule brings every one of those schools below one at this budget, and the margin between the rules peaks at intermediate budgets (683 schools at 10%, 1,187 at 25%, 836 at 50%), because a large budget reaches the biggest pools either way: the rule matters most in the budget-constrained regime that describes current programs.

Health departments are often required to prioritize the lowest-coverage jurisdictions, so we evaluated a hybrid rule that ring-fences a fixed fraction of the budget for those schools. Reserving half still averts 1.43-fold more infections than spending all of it lowest-coverage-first, retaining 47% of the unconstrained gain; a quarter retains 71% (Supplementary Fig. 7, Supplementary Table 18). The efficiency argument does not require abandoning an equity constraint, it prices one.

### Pricing the ordering gap: 25 to 271 expected infections forgone per season

Both rules use the same doses and delivery, so the difference is a pure allocation loss. The 14,113-infection gap is conditional on an introduction reaching each school, so summing over 36,031 schools prices a season in which every school is seeded at once; the probability cancels in the ratio, leaving the 1.92-fold gain unaffected, and reporting the gap in dollars overstates the expected loss by 1⁄p. Counting the directly immunized recipients as well, a constant 72,503 across rules, gives a total-protection gain of 1.16-fold, the figure for a program choosing on total cases prevented (Supplementary Note 2, Supplementary Table 19).

Two decompositions bracket the introduction probability, p = 0.0018 and p = 0.019 (Supplementary Table 20), across which the coverage-rank rule fails to avert 25 to 271 expected infections per season, worth US$2.6 to 28.4 million societally, a fourth pessimistic anchor at p = 0.05 gives 706 expected infections (US$74 million, Fig. 2c, Supplementary Tables 20 and 21), reported for scale only33,38,39; sensitivity to the assumed cost per case and to the cost perspective is in Supplementary Figs. 8 and 9. The range spans an order of magnitude because p rests on one outbreak, so the dollar figures indicate scale rather than estimate it; expected net benefit of the funded allocation against no program, a different comparison from the rule-versus-rule gap priced here, is positive in 14 of 16 probability-by-perspective combinations, negative in two at the lowest probability under the narrowest perspectives (Supplementary Figs. 8 and 9, Supplementary Table 21). In those two the doses do not pay for themselves within a single season, which understates a multi-season return we do not model; the allocation comparison is unaffected. These figures price the unweighted ceiling against coverage rank; because the efficiency ratio is free of both the cost per case and the introduction probability (Methods), whatever fraction of the ceiling survives the risk test below is worth the same fraction of this range.

### Exposed schools concentrate in susceptible headcount, not enrollment

Our implementation of the school-level gravity-kernel reproduction number reproduces the published estimates across all 26 usable state-years (Supplementary Table 2). Such models estimate amplification given an introduction, not where importations land, so we restricted attention to the 81 counties with a demonstrated 2025-26 introduction and asked whether pre-outbreak risk predicts secondary cases after the index report, ranking counties within states^12^ (Supplementary Fig. 10a).

Of the 18 counties that amplified (≥5 secondary cases), six fall in the top decile of their state’s susceptible pool, a 3.3-fold enrichment of amplifying counties in the top decile (p = 0.0064; Supplementary Fig. 10b, Table 1). The *R_v_*-based metrics point the same way without significance (1.67-fold, p = 0.27; Supplementary Fig. 11), and overall discrimination is near chance, so the signal sits in the extreme tail.

School-scale enrichment here was 1.67-fold (3 of 18, p = 0.27) against 2.0-fold across seven earlier outbreaks^12^, a power limit not a replication failure: the exact interval (0.36 to 4.14-fold) contains both the null and the published value (Supplementary Tables 22 and 23). The epicenters are also where the database is thinnest. A county cannot generate more secondary cases than its recorded susceptible pool unless the records are incomplete; 8 of 81 fail that filter, including every Texas and New Mexico epicenter (Supplementary Fig. 10c). Among the 73 that pass, school-scale enrichment rises to 3.0-fold (p = 0.070) and county-scale to 6.0-fold (p = 0.0001), monotonically in data richness (Supplementary Note 5, Supplementary Tables 24, 25 and 26): a statement about surveillance, not the risk model.

As a parameter check we fitted exponential growth rates in three outbreak settings, including Utah, which contributes nothing to the risk model; *R*_0_ = 15 falls inside the 95% interval in 8 of 15 windows and all seven misses fall below it, the direction predicted where within-group transmission deflates a reproduction number inferred from between-group growth^40,41^ (Supplementary Fig. 10d, Table 1, Supplementary Methods).

**Table 1.** Validation of the risk ranking against the 2025-26 outbreaks. **a.** School scale, South Carolina Upstate. Coverage measured in 2024-25, the school year before the outbreak; exposure during 2025-26. Within-region analysis, n = 424 schools with pre-outbreak coverage, 30 exposed. Top decile defined by rank so the comparison set is identical across metrics (42 schools). Enrichment is the exposure rate in the top decile divided by the rate across all 424 schools; intervals are Clopper-Pearson; p from one-sided Fisher exact test; AUC is Mann-Whitney. **b.** County scale, five outbreak states. Conditional on introduction: 81 counties with a recorded index case; 18 amplified (≥5 secondary cases). Metrics are percentile ranks within each state. One-sided binomial test against a null of 0.10.

| Ranking metric | Exposed in top decile | Enrichment | 95% CI | p | AUC |
| --- | --- | --- | --- | --- | --- |
| Susceptible headcount | 15 / 42 | 5.05× | 3.05-7.35 | $3.8 \times 10^{-9}$ | 0.84 |
| School $R_v$ | 12 / 42 | 4.04× | 2.22-6.30 | $4.4 \times 10^{-6}$ | 0.80 |
| Coverage deficit | 10 / 42 | 3.37× | 1.70-5.58 | $2.2 \times 10^{-4}$ | 0.77 |
| Enrollment only (negative control) | 3 / 42 | 1.01× | 0.21-2.75 | 0.59 | 0.62 |

| Risk metric | Amplifying counties in top decile | Enrichment | p |
| --- | --- | --- | --- |
| Susceptible pool | 6 / 18 (33.3%) | 3.33× | 0.0064 |
| Maximum school | 3 / 18 (16.7%) | 1.67× | 0.27 |
| Schools above $R_v = 1$ | 3 / 18 (16.7%) | 1.67× | 0.27 |
| Top school percentile | 3 / 18 (16.7%) | 1.67× | 0.27 |

*Note: Growth-rate calibration: $R_0 = 15$ falls within the 95% confidence interval in 8 of 15 exponential growth windows across three independent outbreak settings (Gaines County TX, Upstate SC, Utah statewide); generation time 11.7 days; all seven misses fall below the interval, the direction structured-population theory predicts.*

### The premise the theorem needs fails where it can be tested

The county-scale test cannot reach the unit the rule acts on, because the states that amplified in 2025-26 have the thinnest school records. South Carolina is the exception. A file compiled from the department’s public releases covers its 2025-26 Upstate outbreak: 1,564 schools, coverage for both years, and 31 K-12 schools flagged measles-exposed (Supplementary Note 6, Supplementary Tables 4 and 27). Coverage precedes the outbreak, so the predictor cannot be contaminated by the outcome. The 30 analyzed exclude one school lacking 2024-25 coverage, and all 31 fall in the Upstate, so we report within-Upstate figures throughout (n = 424 schools); statewide gives a region-confounded 5.66-fold enrichment (Supplementary Table 28). The release reports whole-school enrollment and an all-grades rate, not the kindergarten units as in the national panel, so this test transfers a ranking, not a headcount (Methods). Ranking by susceptible headcount, 15 of the 30 exposed schools fall in the top decile, over-represented 5.05-fold against the region-wide rate (95% CI 3.05-7.35, p = 3.8 × 10⁻⁹), with AUC 0.84 (Fig. 4 a, b, Table 1).

Restricting to the schools with a kindergarten grade, 269 total with 19 exposed, leaves that estimate at 4.72-fold (95% CI 2.34 to 7.64) with the negative control still failing, so it is not an artifact of grade composition (Supplementary Fig. 12, Supplementary Table 29). The school *R_v_* gives 4.04-fold and coverage deficit 3.37-fold, all discriminating with overlapping intervals. This pattern is not an artifact of school size. Enrollment carries the size component of susceptible headcount while stripping out coverage, so if exposure merely tracked school size it would reproduce the enrichment. It does not: enrollment alone gives 1.01-fold (95% CI 0.21-2.75, p = 0.59), a negative control that fails to discriminate (Fig. 4b), while percentage unvaccinated adds strongly beyond log-enrollment (0.114 per point, likelihood-ratio χ² = 26.7 on 1 *df*, p = 2.4 × 10⁻⁷; Supplementary Table 30). Exposed schools averaged 84.7% coverage in 2024-25 against 92.1% among the unexposed (Fig. 4c). This is the only out-of-sample test at the scale the allocation acts on, the ranking fixed before these exposures occurred. It survives heavy undercounting, including undercounting that tracks coverage: significance is lost only beyond 62 missed schools, a 67% non-detection rate, and assigning missed exposures preferentially to above-median-coverage schools, the direction that would manufacture the enrichment from investigation intensity, leaves it at 1.94-fold with the same bound (Supplementary Fig. 13, Supplementary Tables 31).

The quantity the rule depends on moves independently of mean coverage: across the decade spanning California’s Senate Bill 277 mean coverage rose while the share of susceptibles held by the lowest 5% of schools rose from 32% to 52%, and South Carolina shows the same independence over two consecutive years with no mandate change (Supplementary Note 7, Supplementary Tables 32 and 33).

The risk scaling is therefore a bounded unknown rather than a transferable estimate, which is why the rule comparison below is posed as a decision under uncertainty.

### Non-uniform introduction risk inverts the ranking of allocation rules

The allocation objective weights every school equally, pricing a season in which each faces an introduction with equal probability; the South Carolina validation refutes that premise on our own data, and the consequence shows in where the doses go once it is dropped. In Upstate South Carolina 26 of 30 exposed schools sat below *f̄* and 14 at or below 85% coverage, with 62% of the susceptibles in exposed schools sitting at or below 85% coverage (Supplementary Tables 34 and 35). The dose-optimal rule sends nothing into that band: 430 of 4,510 doses (9.5%) reach later-exposed schools against 1,942 (43.1%) under lowest-coverage-first (Supplementary Fig. 16a, Supplementary Table 36).

One caveat bounds that risk model: fitted inside one reached region, it estimates exposure risk given an introduction reached the county. Between counties it is the within-county spread of school coverage, not the mean, that predicts which counties were reached (p = 0.019 against p = 0.79), the same clustering mechanism at county scale, and the concentration appears nationally in exemptions^15^ as in transmission12. The between-county model, exemption trends and state-by-state replication diagnostics are in Supplementary Note 3 (Supplementary Fig. 14, Supplementary Tables 37 and 69).

The school-level scaling replicates once outside South Carolina: Clark County, Washington published school MMR exemption rates before its 2018-19 outbreak, and the same specification on 131 schools with 12 exposed gives +0.232 log-odds per point unvaccinated (SE 0.074) against +0.114 (SE 0.024) here, enrollment again failing as a control. That is one replicate; the remaining registers publish exposed schools without a usable denominator, a visit list rather than an investigation register, or too few exposed schools to support a coefficient (Supplementary Table 38). These two fitted coefficients anchor the uncertainty range analyzed below rather than a scaling chosen for convenience (Supplementary Fig. 15a).

Re-scoring the objective by introduction risk separates two claims that are easy to conflate. Fitting the risk model to the South Carolina exposures and applying it nationally with a tempering exponent *α*, where 0 means introduction risk is assumed uniform across schools and 1 means it scales at the full strength the South Carolina fit implies, the unweighted ranking falls below lowest-coverage-first beyond *α* ≈ 0.33, reaching 0.20-fold untempered. But re-deriving the ranking on the weighted objective beats lowest-coverage-first at every weighting strength tested, 1.37-fold to 1.92-fold (Supplementary Fig. 16b, Supplementary Table 39). The advantage is preserved when the exposure expectation scales with school size instead (1.95- to 2.02-fold). The first says an optimizer tuned for one objective underperforms on another, true of any optimizer; the second says marginal efficiency still says what to do once the objective is corrected.

### A light hedge is the most robust rule, and clears 986 more schools than coverage rank

A program cannot condition on an *α* it does not know, so the operational question is which single rule to field. We evaluated six candidate rules plus the minimax-regret design, seven in total, against the attainable benefit at each *α*, re-deriving the ranking at that *α* rather than scoring against the best rule on a menu, and took each rule’s worst case over *α* ∈ [0,1] (Fig. 5a, b; Table 2). Worst cases are relative regret, the share of attainable benefit a rule retains rather than an absolute shortfall, so they stay comparable where the attainable total differs; and the seven are a restricted family, six fixed rules plus the one-parameter blend, so the selected rule is least-bad within that family rather than over all allocation rules. The ordering is the result: the unweighted dose-optimal rule is worst of the seven at 14.2% of attainable and lowest-coverage-first second worst at 52.1%, while four of the seven candidates hold above 67%. Best is a rule spending a fraction *λ* of its objective on measured risk and the rest uniformly, written λ to keep it distinct from the risk-scaling exponent α. The blend is linear, so it floors every school’s weight at 1 − *λ* rather than shrinking all weights multiplicatively, and a small *λ* therefore buys a large change in the worst case: at *λ*^∗^ = 0.02 its worst-case regret is 10.3%, against 21.3% for the next-best candidate and 85.8% for the unweighted rule (Fig. 5b, Supplementary Tables 40 and 41). The minimum is flat: every weight from 0.01 to 0.03 sits within two percentage points of it (Supplementary Table 40).

**Fig. 5.**
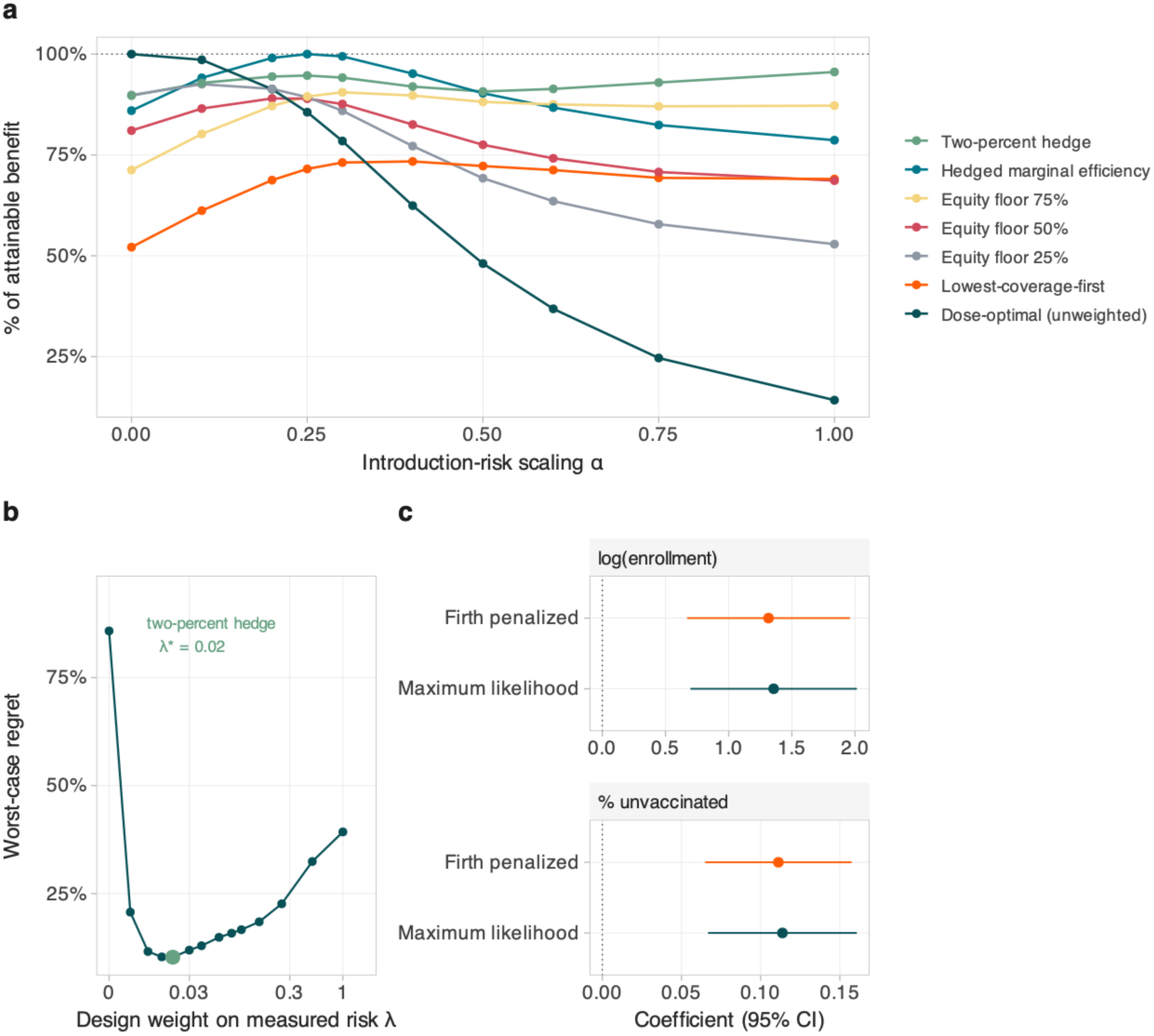
Rule performance under uncertainty about the introduction-risk scaling. Numerical values for every candidate rule are in Table 2. **a**, Infections averted by each of seven candidate rules as a percentage of the attainable benefit at each *α*, meaning the marginal-efficiency ranking re-derived at that *α*. The denominator is therefore the best attainable benefit at that value, not the best of the seven rules; scoring against the best rule on a fixed menu would flatter every rule on the menu. **b**, Worst-case regret of a design that spends a fraction *λ* of its objective on measured risk and the remainder uniformly, (1 − *λ*) + *λw*, as a function of *λ*. The curve has a flat interior minimum, within two percentage points across 0.01 to 0.03, at *λ*^∗^ = 0.02, marked and hereafter the two-percent hedge, where worst-case regret is 10.3% and the plateau rather than the point is the result; *λ* = 0 is the unweighted dose-optimal rule at 85.8% and *λ* = 1 the fully calibrated one at 39.3%, so both extremes are worse than a light hedge. The x-axis is on a pseudo-log scale so the minimum is resolvable. **c,** Maximum-likelihood and Firth-penalized coefficients of the exposure model, faceted on free scales because the two differ by an order of magnitude. Penalization shrinks the coverage slope by 2.3% and leaves the ranking in Table 2 unchanged, so the imprecision reflects the design, 30 exposed schools in one region, rather than estimator bias.

**Table 2.** Worst-case performance of each candidate rule under uncertainty about the introduction-risk scaling. Six candidate rules plus the minimax-regret design, seven in total, scored against the attainable benefit at each value of the introduction-risk scaling exponent α, meaning the marginal-efficiency ranking re-derived at that α. Worst case is the minimum of that ratio over the α grid and regret its complement. The unweighted dose-optimal rule has the worst case of any candidate; a light hedge at λ = 0.02 has the best. Firth column repeats the worst case under Firth-penalized exposure coefficients. Source values: Supplementary Tables 78 and 40.

| Rule | Worst case (% of attainable) | Worst-case regret (%) | $\alpha$ at worst case | Mean across $\alpha$ (%) | Worst case, Firth (%) |
| --- | --- | --- | --- | --- | --- |
| Dose-optimal (unweighted) | 14.2 | 85.8 | 1 | 64.0 | 14.7 |
| Two-percent hedge (minimax-regret design, $\lambda = 0.02$) | 89.7 | 10.3 | n/a | 92.8 | 89.7 |
| Hedged marginal efficiency (ranking derived at $\alpha = 0.25$) | 78.7 | 21.3 | 1 | 91.2 | 77.8 |
| Equity floor 75% | 71.2 | 28.8 | 0 | 85.8 | 71.2 |
| Equity floor 50% | 68.6 | 31.4 | 1 | 80.7 | 68.3 |
| Equity floor 25% | 52.9 | 47.1 | 1 | 77.0 | 52.9 |
| Lowest-coverage-first | 52.1 | 47.9 | 0 | 68.2 | 52.1 |

Worst-case regret varies by 1.6 percentage points across that range (Supplementary Table 40).

Both fitted coefficients sit inside the widened set, while the lower end is the uniform-risk null rather than an estimate: *α* = 1 is the South Carolina coefficient, *α* = 2 the Washington replicate. Widening the set to *α* ∈ [0,2] worsens the unweighted rule rather than rescuing it, 14.2% to 10.6%, while the two-percent hedge holds at 89.7%, unchanged from the narrower set because its worst case falls at the uniform-risk end rather than at the fitted coefficient (Supplementary Table 42). It holds at 10% and 50% budgets and under Firth penalization (Supplementary Note 4, Supplementary Tables 43 and 44).

At the same 25% budget the hedged rule brings 2,497 of those 12,196 schools below one secondary case, against 1,511 under lowest-coverage-first and 2,698 under the unweighted dose-optimal rule: 986 more schools than coverage rank, and 112,733 fewer children left in a school where a chain can persist (Fig. 6a). It does so while still directing 21% of the budget to schools below 75% coverage, against 62% under coverage rank (Fig. 6b). Neither count depends on the risk-scaling exponent, because the school-level reproduction number is a function of post-allocation coverage alone and dose placement is a property of the rule; the hedge forgoes 7.4% of the unweighted rule’s clearing while retaining 89.7% rather than 14.2% of attainable benefit in the worst case.

**Fig. 6.**
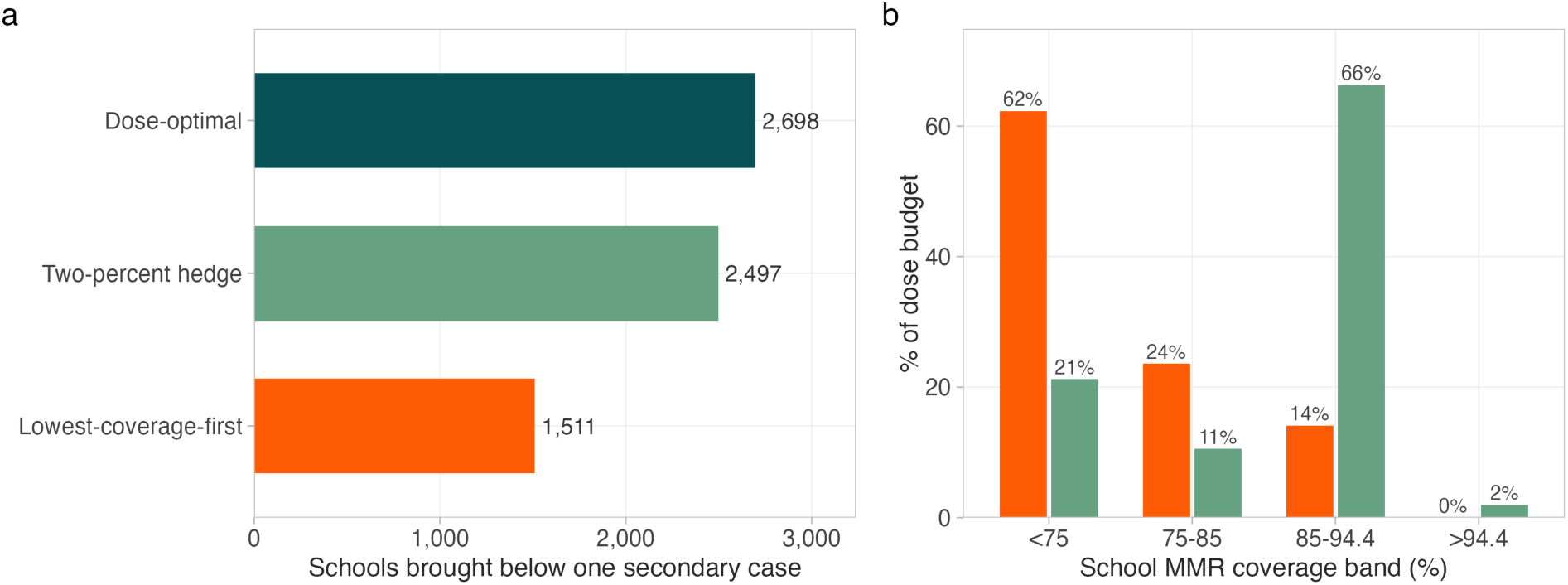
What the rule choice changes in program units. Both panels are computed at an identical 77,292-dose budget, 25% of the national susceptible pool, with each state spending its own 25%. **a,** Schools brought below one secondary case under the lowest-coverage-first rule in current use, the two-percent hedge and the unweighted dose-optimal rule. Of the 12,196 schools that can currently sustain a chain, the hedge clears 2,497 against 1,511 under coverage rank, 986 more schools, leaving 112,733 fewer children in a school where transmission can persist. **b,** Share of the same budget directed to each coverage band under lowest-coverage-first (orange) and the two-percent hedge (green). The hedge still sends 21% of the budget below 75% coverage against 62% under coverage rank, and 2% above the 94.4% inflection. Neither panel depends on the risk-scaling exponent: the school-level reproduction number is a function of post-allocation coverage alone and dose placement is a property of the rule.

The negative result is not an artifact of how the uncertainty set is defined, which is the strongest form the claim can take while α is unidentified. Defining the set three structurally different ways, over the coefficient’s own 95% confidence interval with no tempering, over four functional forms for the weight, and over a coverage-only refit dropping enrollment, leaves the unweighted rule worst in every instance (12.1%, 12.1% and 34.1% of attainable). Which hedge wins depends on the set, and under the coverage-only refit lowest-coverage-first attains the benchmark while the unweighted rule remains worst; what is invariant is the negative result, not the choice among hedges (Supplementary Table 45).

The defensible statement is narrow: the advantage of unweighted marginal-efficiency allocation is not robust to plausible introduction-risk weighting, while a partially weighted ranking or moderate equity floor performs better in the worst case. That is not an argument for lowest-coverage-first, which every hedge beats on the primary uncertainty set.

*α* is the central unmeasured quantity and penalization does not measure it: a Firth fit shrinks the slope 2.3% and leaves the ranking intact (Fig. 5c, Supplementary Table 44). Nor is the weight cap correcting extrapolation: 99.6% of national schools fall inside the fitted linear-predictor range (Supplementary Table 46). The imprecision is a property of the design, one outbreak in one region with 30 exposed schools, not of the estimator: the coefficient is 0.114 per point (95% CI 0.067 to 0.161; Supplementary Table 30), so the direction is firm and the magnitude is not. The 30 exposures are also not independent draws, since 28 fall in one county, but the slope survives that: county-clustered errors give a design effect of 1.67 and p = 0.0032, against the 5.87 that would be needed to push the coefficient to p = 0.05, so the slope tolerates roughly three and a half times the clustering it carries; a county random intercept gives p = 0.0026, and restriction to the outbreak county alone, which removes between-county variation by construction, strengthens it to p = 0.0004, with Moran’s I on the model residuals 0.064 (permutation p = 0.035). What would resolve it is exposure records linked to pre-outbreak coverage in more than one jurisdiction. The two-percent hedge inherits that imprecision without depending on it: 0.02 is an interior optimum of the uncertainty set we analyze rather than a calibrated parameter, and what transfers is that a light hedge outperforms both extremes, not the value.

## Discussion

At an identical 77,292-dose budget, the hedged rule brings 2,497 of the 12,196 schools that can currently sustain a chain below one secondary case, against 1,511 under the coverage ranking programs use now: 986 more schools, and 112,733 fewer children left where transmission can persist, with no additional doses and no new data collection (Fig. 6). It does this while still directing 21% of the budget to schools below 75% coverage. Neither count depends on how steeply introduction risk scales, because the school-level reproduction number is a function of post-allocation coverage alone. The reason the hedge beats both the intuitive rule and the theoretically optimal one is a reversal. The theorem’s binding case is the empirically common one: two thirds to three quarters of US susceptible children attend schools where ranking by marginal efficiency favors completing higher-coverage schools first^19^. But the uniform-introduction assumption it rests on fails in the one place it can be tested, and once it is dropped the theoretically optimal rule is the worst of the seven candidates we evaluate while a light hedge holds 89.7% of attainable benefit. Minimax regret is the right tool for a parameter two outbreaks cannot identify, and not a substitute for estimating it; the ranking it produces is stable where the parameter is not. The operational recommendation is therefore the hedged rule, not the dose-optimal one; a program that reads this result as “send doses to high-coverage schools” has inverted it, because no hedge we test sends doses to the schools that need them least.

One boundary deserves stating plainly. Our findings concern the allocation of a fixed vaccination budget across schools, not its size: every rule compared here delivers the same total doses, and all rely on maintaining and expanding the childhood vaccination schedule as the foundation of measles control. Nothing here argues for withholding doses from any school, and the gains we report are unavailable to a program whose budget or coverage base is shrinking. Allocation under supply constraints is familiar from COVID-19 and yellow fever^42,43^; for MMR the binding constraint is less dose availability than the staff able to convert a dose delivered into a dose accepted^44,45^.

Three cases cover what a program should run. The requirement is light: both rules are functions of reported coverage and enrollment alone, and deleting school coordinates leaves every state’s allocation identical (Supplementary Note 2, Supplementary Table 14). What constrains deployment is not computation but which schools the rule selects. Where no linked records exist, almost everywhere, field a hedged rule: the two-percent hedge holds 89.7% of attainable in its worst case, and a moderate risk weighting or 75% equity floor holds above 68%, against 14.2% unweighted (Table 2). Where a jurisdiction holds linked records, refit the model and re-derive the ranking on the weighted objective, which beats coverage rank at every weighting. Where statute binds allocation to the lowest-coverage settings, ring-fence the mandated share and spend the rest by marginal efficiency, retaining 71% of the gain at a quarter, 47% at a half. In none is the unweighted theorem-optimal rule right to field, nor is coverage rank. One requirement is temporal rather than informational. A school-level coverage ranking retains only 52% of its lowest decile after a year and 43% after three, so this rule should be fielded on current-year or prior-year coverage rather than on the most recent file a program happens to hold (Supplementary Note 3, Supplementary Fig. 26, Supplementary Tables 79, 80).

A floor on coverage is not a floor on sector. A dose at a 50%-coverage school cannot approach the herd-immunity threshold, while at 90% it can cross it, so the rule sequences rather than abandons. One consequence deserves naming: total infections fall under the dose-optimal rule, but it raises the vaccinated share of cases while cutting their number, which is arithmetic of a falling case count rather than any change in vaccine performance, and is harder to defend publicly (Supplementary Tables 49, 50). Coverage and concentration are separate, so a policy aimed at the mean will not address the clustering driving risk46-50 (Supplementary Figs. 20-24, Supplementary Note 7).

The saturation target is not a free parameter of the herd-effect function. Differentiating the final-size relation shows that the coverage at which the herd effect stops repaying doses solves *R*_0_*s*_0_ = 1, which up to the seeded fraction is the classical vaccine-adjusted herd-immunity threshold (1 − 1/*R*_0_)/*VE* at 96.2% so the objective is anchored to the textbook quantity by construction rather than by agreement (Supplementary Fig. 16c, Supplementary Fig. 17, Supplementary Table 47)^51^.

Where sector is reported with enrollment the rules swap places: the dose-optimal rule over-represents private schools where lowest-coverage-first does not, and which rule lands harder on the private sector differs by state, because the sector mix follows which part of the coverage distribution each rule spends in (Supplementary Figs. 18 and 19, Supplementary Note 3, Supplementary Tables 51-57, Supplementary Figs. 17-19). Fewer states can compel private-school reporting, so jurisdictions should compute the split locally first.

What this paper adds is the prescription. A school-level allocation problem is only well posed because risk resolves below the county scale, our previous study’s diagnostic result, which we take as given rather than re-argue^12^. Prior school-targeting studies compare arms consuming different dose counts or rank schools by risk rather than by marginal return^27,52,53^, and our South Carolina and Clark County estimates agree with them on where introductions land. The distinction this paper draws is that arrival and return are different quantities: low coverage marks where a case is likely to arrive, but it does not fix the order in which doses buy the most protection, and conflating the two is what the intuitive rule costs.

Several limitations bound these conclusions (Supplementary Note 10). First, and most consequentially, school-level exposure linkage exists in only two jurisdictions. Texas and New Mexico schools are in the allocation database and receive doses under every rule, and 2,142 of the 2,628 cases in the five-outbreak series we assemble, 81.5%, fall in jurisdictions the database covers; but their epicenter counties are among the 8 of 81 that fail the susceptible-pool consistency filter, so the ranking cannot be validated where most 2025-26 spread occurred^54^. South Carolina and Clark County, Washington are the exceptions. Second, three states publish whole-school rather than kindergarten enrollment and hold 42.4% of the susceptible headcount; dropping them raises both headline figures (Methods). Third, the risk model behind exposure-coverage relationship *α* is fitted to 30 exposed schools in one region and to whole schools, so it transfers as a ranking, not a headcount; the sign replicates in one independent county, but the magnitude does not, and the conclusion holds across the range the two estimates span. Fourth, structural simplifications bound scope: separability prevents the risk model representing between-school herd protection^55,56^, the herd effect *H* is a single-population final-size calculation. Under school-specific seeding a per-school allocation is not resolvable on the integer coverage lattice, and the deterministic final-size relation has no extinction, so at kindergarten enrollments it places the inflection above an exact stochastic treatment seeded with one index case (Supplementary Note 1, Supplementary Fig. 28).

Five further limitations are set out with their evidence in Supplementary Note 10 (Supplementary Figs. 20-23, Supplementary Tables 58-72). Whether these results generalize to lower-*R*_0_ pathogens or outside the United States is untested. Two extensions sit outside an allocation paper: school-to-school mobility would permit a network-aware rule^56^, and genomic data would confirm which schools were in fact sustaining transmission.

This is a modeling result about ordering a dose budget, not clinical or programmatic guidance. Any change to allocation policy would need review by the responsible public health authority, and the constraints such a review weighs, from statutory equity to delivery capacity and consent, lie outside what a final-size model represents.

Raising national coverage, or reversing the exemption trends behind it^15,57,58^, takes years; reordering an already-committed dose budget takes one procurement cycle. This is a result about what can be done inside a fixed budget, with no new doses, no new data collection and no new authority.

Which of these rules is best for a given jurisdiction turns on *α*, unidentified anywhere in the United States today, and the constraint is definitional rather than one of collection: the records that would resolve it are already held and simply not published. Departments name the public places a case visited; few name the schools an investigation linked to a case. A visit list records where an infectious person was; an investigation register records which schools a department judged exposed and acted on, which is what a weighting requires. South Carolina and Clark County published the second kind, which is why estimates exist for two regions only. One change in reporting practice would resolve the central uncertainty at no collection cost: publishing which schools an investigation touched, alongside their coverage the year before, would convert this paper’s central uncertainty into an estimate. The finding also travels beyond measles: in any targeted-vaccination, testing or screening problem where the targeting variable is measured well and the exposure variable badly, the point-estimate optimum is the fragile choice, and a light hedge retains most of its value. Until then, for measles, a light hedge is the rule to field.

## Methods

### Data

#### Primary panel

School-level kindergarten MMR coverage was assembled from state open-data releases, giving 224,353 school-years across 33 states from 2013-14 to 2024-25. Each record carries school identifier, enrollment, percent MMR coverage, county and geocoded coordinates; records were retained where enrollment ≥ 10 and coverage fell in [0, 1] or [0,100]. The enrollment field is taken as each state publishes it, and three states publish a denominator that is not the kindergarten cohort: median per-school enrollment is 393 in Hawaii, 318 in Illinois and 313 in Iowa, against 34 to 83 across the other 25 states, and their panel totals exceed published statewide kindergarten enrollment by roughly twelve- to fourteen-fold, so these three report whole-school or whole-district counts. The three flagged states contain 6,006 schools (16.7% of the panel) and 42.4% of the national susceptible total. We retain them because the analysis is a within-state ranking and the herd-effect function is evaluated on coverage, which is unaffected by the denominator, but the national susceptible headcount is inflated in consequence, and the leave-one-state-out interval bounds the effect on every reported share (Supplementary Table 8). Dropping all three raises the convex share from 75.8% to 80.8% and the efficiency gain from 1.92-fold to 2.29-fold, so the reported figures are the conservative ones. County-day measles case counts for 2025-26 are from a public outbreak tracker; boundaries are the 2023 US Census cartographic files^59^. The meaning of the enrollment field was verified against NCES Common Core of Data grade-0 counts, which reproduce the classification independently (Supplementary Note 6, Supplementary Table 81).

#### South Carolina exposure file

The validation panel is a 2024-25 and 2025-26 school file with per-school exposure flags, address-geocoded by us, compiled from South Carolina Department of Public Health releases published at https://dph.sc.gov and obtained before school-level rates were consolidated into the current aggregated format. Coverage is a school-wide rate across all grades, from the department’s 45-day school reports. The report records the share of enrolled students holding a valid South Carolina Certificate of Immunization, which certifies compliance with the full required school schedule rather than MMR receipt alone; because two MMR doses are required for kindergarten through grade 12, this rate is a lower bound on MMR-specific coverage and the susceptible headcounts derived from it are correspondingly upper bounds. The validation is a rank-based top-decile test, so this affects it only insofar as the gap between certificate completeness and MMR-specific coverage varies across schools. The exposure flags are not a departmental data product: they were reconstructed from the department’s public outbreak communications, enumerating the release series from the first school announcement on 8 October 2025 to the declaration that the outbreak had ended, recording the date of first announcement as the exposure event and treating later appearances as continuations. The reconciliation of that register against the deposited panel is documented in Supplementary Note 6 and Supplementary Table 73. Every school with reports leading to an “exposed” classification was public. The exposure definition and quarantine records are described in the published report on this outbreak^13^.

#### Multistate exposure linkages

Seven state outbreak records were assessed for whether they allow an independent estimate of *α*; two do (Supplementary Note 6, Supplementary Table 74). The binding requirement is an investigation register, meaning a departmental list of schools judged exposed, rather than a list of locations a case visited, together with pre-outbreak coverage for those schools and a comparison set of unexposed schools from the same release. Retrieval sources, dates, exact pages and the verification procedure for each state, and the reason Arizona is deposited but not fitted, are in Supplementary Note 6. **Supplementary sources**. Three further files support only supplementary state-level analyses and no main-text result; they are described, with provenance and license status, in Supplementary Note 6 and the replication package manifest.

### Reporting-quality exclusion

States were excluded where more than 40% of schools reported exactly 100% coverage, indicating censoring rather than saturation. Five met this criterion (North Dakota, Connecticut, Arkansas, Maryland, Maine; Maine reports 89.5% of schools at exactly 100%). The threshold was varied from 20% to no exclusion (Supplementary Table 3).

### Persistence of the coverage ranking

Both rules rank schools on coverage measured before the allocation, so how long a ranking remains valid bounds how stale a coverage file may be. The primary panel carries no stable school identifier across years, so schools were keyed on state and coordinates rounded to five decimal places; keys resolving to more than one record in any year were dropped rather than aggregated, because a shared coordinate is generally a campus with several reporting units and averaging them would create a school that does not exist. That leaves 153,879 school-years across 27 states, a mean of 10,149 schools per one-year comparison. For every ordered pair of years we computed the Spearman rank correlation of coverage and the retention of the lowest decile, defined as the share of the 10% of schools with the lowest coverage in the earlier year that are still in the lowest 10% in the later year, and averaged both over all pairs at each lag. Louisiana was excluded because its coverage field is a single snapshot repeated across the four years it contributes; the diagnosis, and the cross-year identical-value audit for every state, are in Supplementary Note 3 (Supplementary Fig. 26, Supplementary Tables 79, 80 and 82). No reported result depends on the exclusion, because main-text quantities are computed within a single year.

### Herd-effect function and critical fractions

For a single population with coverage *f*, vaccine efficacy *VE* and initial infected fraction *i*_0_, the susceptible fraction is *s*_0_ = 1 − *fVE*. The final size of a Kermack-McKendrick SIR epidemic^18^ solves the implicit equation

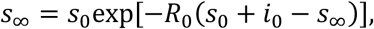

with (*f*) = *s*_0_(*f*) − *s*_∞_(*f*) the fraction of the population infected at coverage *f*. Three quantities follow, and the paper uses them consistently. Two symbols are used deliberately: *H* denotes the herd effect, the deterministic expected benefit on continuous coverage, which approaches the classical threshold from below as the assumed introduction vanishes; G denotes the same expected benefit where children come in integers, on the achievable coverage lattice and under exact stochastic seeding (used in Supplementary Note 1, Supplementary Fig. 28). Total protection is *T*(*f*) = *Z*(0) − *Z*(*f*), the fraction of the population spared infection. The directly protected fraction is *fVE*, the vaccinated who are immunized whatever anyone else’s coverage. The indirect benefit is the difference,

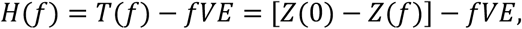

and it is *H*, not *T*, that the allocation analysis optimizes. The direct credit is netted out scaled by the seeded susceptible fraction, *f* V_E_(1 - *i_0_*) rather than *f* V_E_, which is what makes H non-negative at every coverage level; the correction is linear in f, so it leaves the curvature, both critical fractions and the allocation ordering unchanged. The distinction is not cosmetic. *fVE* does not depend on how doses are distributed across schools, so it is a constant added to every rule’s total, and it cancels from no ratio: subtracting it changes both the location of the inflection and the size of any reported gain. We therefore report the indirect ratio and the total-cases ratio separately throughout, and never a ratio of one against the other. The indirect ratio is the primary figure because it isolates the mechanism the allocation theorem speaks to, the herd effect that depends on where doses land; a program evaluating total burden should use the smaller total-protection figure, and both are reported side by side in the Abstract and Results for that reason. *H* is a fraction of the school’s enrollment, not of *Z*(0); multiplying by enrollment gives infections.

Following Duijzer et al.^19^, three critical fractions follow: *f̄*, the inflection at which *H* changes from convex to concave, where *H*″(*f̄*) = 0; *f̄*, the dose-optimal fraction maximizing the average return *H*(*f*)/*f*; and *f*^∗^, the critical coverage above which the outbreak does not take off. The three depend on different premises: *f*^∗^ is governed by R_0_ and *VE* and coincides with the classical vaccine-adjusted herd-immunity threshold (1-1/R0)/VE, denoted pc in Supplementary Fig. 16c, in the small-introduction limit, whereas the inflection *f̄* also depends on the assumed introduction *i_0_*, because it is a property of the marginal return to a dose rather than of herd immunity (Supplementary Note 1, Supplementary Fig. 27). For measles *f*^∗^ and *f̄* coincide to three decimals because the coverage maximising the herd effect solves *R*_0_*s*_0_ = 1 up to the seeded fraction, a closed form rather than a numerical accident, which is why the overshoot penalty below is stated against *f̄*. The convex region is a property of the assumed introduction, and it is widest when that introduction is small: the inflection rises toward the critical fraction as *i*_0_ → 0 and the concave shoulder above it disappears, so *i*_0_ is specified explicitly. For measles (*R*_0_ = 15, *VE* = 0.97, *i*_0_ = 10^−3^) we obtain *f̄* = 0.944, *f̄* = 0.962 and *f*^∗^ = 0.962. All three were computed across a 3 × 3 grid of *R*_0_ and *i*_0_ (Supplementary Table 6).

### School-level reproduction number

The school-level effective reproduction number follows the gravity-kernel next-generation matrix of Chen and Bento^12^, which is the sole source of the deposited *R_v_* estimates and decay parameter replicated here. References 11 and 13 are earlier, narrower studies from our group, a school-level clustering analysis and a single-state outbreak case report respectively, and supply no parameters or estimates used in this work. For schools *i* and *j* separated by great-circle distance *d_i_*_j_, the spatial kernel is

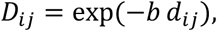

and the transmission scaling *β* is set so the dominant eigenvalue of the next-generation matrix *K_i_*_j_ = *β D_i_*_j_ *N*_j_ equals *R*_0_, where the subscripts run over schools and the matrix is calibrated on a fully susceptible population. The school-level reproduction number is then

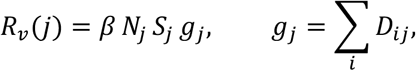

with *N*_j_ the enrollment and *S*_j_ = 1 − *f*_j_*VE* the susceptible fraction at school *j*.

Our implementation reproduces the published estimates at the deposited decay parameter *b* = 10^−3^ km^−1^ to 5.4 × 10^−13^ (Supplementary Table 2). Two implementation details matter. The published pipeline defines susceptibility as *S* = 1 − *f*, folding vaccine efficacy into reported coverage, so the replication uses *VE* = 1 while all other analyses use *VE* = 0.97 explicitly. And *g*_j_ sums over every school in the state-year, so rows must not be filtered before it is computed.

The deposited *b* = 10^−3^ km^−1^ is a national-scale parameter, suited to the question it was estimated for^12^. Our problem is posed within states, so for all analyses other than the replication we set *b* = 0.02 *km*^−1^ (50 km), consistent with measured school-age contact structure^60,61^. This is a change of spatial scope, not a correction, so the *R_v_* magnitudes here are not comparable with the deposited values^12^; only the replication in Supplementary Table 2 uses the published value. At the deposited national value the characteristic length exceeds most states’ width and *R_v_* ranks schools almost identically to a count of unvaccinated children (Spearman ρ = 0.984); at the within-state value used for every other analysis the geometry term varies substantially (CV 0.55, ρ = 0.49), so the ranking is not a rescaled headcount, which makes it the more informative comparison for susceptible headcount outperforming the school-level reproduction number as an exposure discriminator in Table 1 (5.05-fold against 4.04-fold enrichment, Supplementary Fig. 25, Supplementary Table 75). The separable form of *R_v_*(*j*) makes school-level allocation tractable, the marginal effect of a dose at *j* being independent of the allocation elsewhere, which is the condition under which the greedy rule is exact; the same property means the diagnostic does not capture between-school herd protection, tested separately below.

### Allocation rules

Rules were compared at an identical budget *B* of 25% of a state’s susceptible pool, with 10% and 50% as sensitivity analyses. The direct component reported in the Results counts susceptibles actually removed from the pool, which is the budget times efficacy scaled by the seeded susceptible fraction, less a small remainder withheld by the 0.999 coverage cap. Netting out that scaled credit rather than the unscaled product makes the indirect benefit non-negative at every coverage level. Lowest-coverage-first sorts ascending by coverage and brings each school to *f̄* until *B* is exhausted. Dose-optimal ranks by the average herd benefit per dose of that same move, [*H*(*f*) − *H*(*f*_j_)]*N*_j_/*x*_j_, which is the ratio Duijzer’s theorem is stated on^19^. Convex-only threshold targets convex-region schools by descending susceptible count, and uniform distributes in proportion to enrollment. An equity-floor variant ring-fences a share of *B* for lowest-coverage-first and applies the dose-optimal rule to the remainder. Doses convert to coverage as

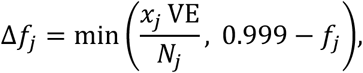

for *x*_j_ doses allocated to school *j*, and the total benefit of an allocation *x* subject to ∑_j_ *x*_j_ ≤ *B* is

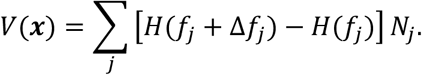

### Cost model

The economic quantity is the difference between two allocations at an identical budget, not the burden of measles; the cost model, its equations and the per-case cost perspectives are specified in Supplementary Methods.

The efficiency-gain ratio is free of both the cost per case and the introduction probability, so it is invariant to either. Conditional net benefit is positive in all 45 cost-by-dose-price-by-budget scenarios, at US$70 to US$412 per infection averted; expected net benefit is negative in 2 of 16 probability-by-perspective combinations, both at the smallest introduction probability under the two narrowest perspectives (Supplementary Note 8, Supplementary Tables 20, 21 and 76).

### Outbreak validation

Analysis was conditioned on introduction: only counties recording a case were retained, the outcome was secondary cases after the index report, and a county was amplifying at ≥5 secondary cases. Risk metrics were converted to percentile ranks *within each state*, so between-state differences in coverage and reporting do not contaminate the signal. Top-decile enrichment was tested with a one-sided binomial test against 0.10 at county scale. For the South Carolina school-scale test the top decile is defined by rank so that the comparison set size is identical across ranking metrics, enrichment is the observed exposure rate in that decile divided by the rate across all schools in the region, intervals are Clopper-Pearson, and significance is by one-sided Fisher exact test on the 2 × 2 table; discrimination over the whole ranking is summarized by the Mann-Whitney AUC. Exponential growth rates *r* were fitted to cumulative case curves over windows of 30-120 days from the first reported case. The realized reproduction number is *R_v_* = *e^rTg^* with generation time *T*_g_ = 11.7 days, and *R*_0_ = *R_v_*/*S* with *S* = 1 − *f*_local_ VE the local susceptible fraction. We use the exponential rather than the linear approximation 1 + *rT*_g_ because the latter is accurate only for *rT*_g_ ≪ 1 and returns *R*_0_ ≈ 2 for every window here, which is incompatible with measles under any calibration.

### Test for between-school herd protection

Because the separable school-level reproduction number cannot express a neighbor effect, we tested for one with a two-level stochastic metapopulation (specification in Supplementary Methods); own coverage dominated and the kernel-weighted neighbor term was not significant, consistent with school-household network analyses of measles spread in high-uptake settings^62^.

### South Carolina temporally external validation

School-level immunization coverage for South Carolina was obtained for 2024-25 and 2025-26 from state releases compiled before school-level rates were consolidated into the current aggregated format; school addresses were geocoded to obtain coordinates for the gravity kernel. Coverage for 2024-25 is the pre-outbreak predictor and exposure during the 2025-26 Upstate outbreak is the outcome, so the predictor is measured in the prior school year. Susceptible headcount is formed the same way as in the national panel, enrollment times one minus coverage, with vaccine efficacy carried in the transmission model rather than the exposure definition, but the units differ and the distinction matters for interpretation. The national panel is kindergarten enrollment and kindergarten MMR coverage; the South Carolina release reports total enrollment and a school-wide immunization rate, and its schools span all grade ranges, so 155 of the 424 Upstate schools (36.6%), one with an unrecorded grade range, and 11 of 30 exposed schools contain no kindergarten grade. Median enrollment is therefore 511 in the Upstate panel against 69 nationally, an offset of 2.0 on the log scale. The exposure model is fitted and interpreted at the unit the South Carolina data provide, the whole school, and the risk ranking it induces is applied to national schools through the linear predictor rather than through absolute headcounts; 99.6% of national schools fall inside the fitted linear-predictor range (Supplementary Table 46). The validation therefore tests whether coverage structure predicts which schools measles reached, at the scale each dataset measures, and not whether a kindergarten headcount transfers between them. Because all exposed schools fall in one of the four state regions (Upstate), every reported figure is computed within that region; the statewide figure is reported alongside only to quantify the regional confounding it would introduce. Enrollment used alone as a ranking score serves as a negative control, and a logistic model of exposure on log-enrollment plus percentage unvaccinated tests whether susceptibility adds information beyond school size by likelihood-ratio test.

### Introduction probability and expected value

The herd effect H prices infections averted conditional on an introduction reaching a school, so the ratio of two rules is invariant to the introduction probability. The cancellation holds because the probability is estimated per school; an introduction probability that scaled with enrollment would not cancel, and that sensitivity is bounded separately. The two estimators of the per-school seasonal introduction probability, their decompositions and the treatment of dose cost are specified in Supplementary Methods.

### Minimax rule selection

Because that risk scaling is not identified from a single outbreak, we treated rule choice as a decision under uncertainty: seven candidate rules, the unweighted dose-optimal rule, lowest-coverage-first, three equity floors and two hedged designs, were evaluated across the grid of introduction-risk scalings, scored against the attainable benefit at each value, and compared on worst-case regret; the candidate set, the blended design family and the scoring convention are specified in Supplementary Methods. The ranking was repeated at 10% and 50% budgets and under Firth-penalized coefficients.

The exposure model was refitted by Firth’s penalized likelihood, which adds 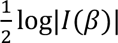 to the log-likelihood and is the standard correction for rare-event bias with 30 events. We also tested whether the weight cap is correcting extrapolation by comparing the national linear-predictor range against the fitted range.

### Interrupted time series

For California we fitted a segmented regression to the annual state series; the model specification, robustness alternatives, placebo fits, permutation inference and the state-level trend fits are given in Supplementary Methods.

### Introduction-risk weighting

To test the uniform-risk assumption we fitted a logistic model of exposure on log-enrollment and percentage unvaccinated to the 424 Upstate South Carolina schools with pre-outbreak coverage and used its linear predictor to form per-school weights *w*_j_ ∝ exp(*η*_j_)*^α^*, normalized to mean one. The tempering exponent *α* regularizes the weights rather than correcting an out-of-support projection: only 0.44% of national schools fall outside the fitted linear-predictor range, and the spread the untempered weights imply is already present within the fitted sample itself (Supplementary Table 46). The weighted objective replaces 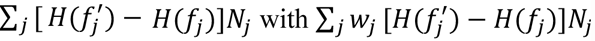. We evaluated two quantities at each *α*: the benefit of the ranking derived on the *unweighted* objective, and the benefit of the ranking re-derived on the weighted objective, both against lowest-coverage-first at the same per-state budget. Reporting only the first conflates a mis-specified optimizer with a failure of the criterion.

### Flag completeness and sector

The exposure flag records schools that a health department investigated and linked to a case, which is not a census; the incompleteness bound and the sector classification rule are specified in Supplementary Methods.

### Statistics and reproducibility

Analyses were run in R^63^. Intervals are 95%, tests two-sided unless stated, and bootstraps use 1,000 resamples. No data were excluded other than by the pre-specified quality rules above. Apart from the stochastic simulation and bootstraps, for which seeds are set in the code, the analysis is deterministic and reproduces from raw inputs.

## Data availability

Sources are in Supplementary Table 77. The school-level vaccination data analyzed here derive from public state open-data releases and are deposited at https://github.com/BentoLab-Cornell/measles-allocation, with a versioned archive DOI to be minted on acceptance. The South Carolina school file (1,564 schools with 2024-25 and 2025-26 school-wide immunization-certificate completeness, geocoded addresses and outbreak exposure flags) was compiled from South Carolina Department of Public Health releases published at https://dph.sc.gov and obtained before school-level rates were consolidated into the current aggregated format; it is included in the deposit because the aggregated version now published cannot reproduce the analysis in Fig. 4. The exposure register reconstructed from the department’s public outbreak communications is deposited alongside it, so the flags can be regenerated from those releases; the geocoding is ours. California kindergarten immunization files are available from the California Health and Human Services open-data portal. County and state boundary files are the 2023 US Census cartographic boundary files. County-day measles case counts are from the public outbreak tracker cited in Methods. Source data for all main figures are provided with this paper.

## Code availability

All analysis code is available at https://github.com/BentoLab-Cornell/measles-allocation, with a versioned archive DOI to be minted on acceptance. The pipeline consists of 44 numbered R scripts with a single driver (run_all.R). A README documents the pipeline, color system, data provenance and limitations.

## Acknowledgements

We thank colleagues in the Bento lab for helpful comments on the manuscript, and colleagues at the Cornell Atkinson Center for Sustainability for discussion and support. We are deeply grateful for the extraordinary public stewardship of these national data systems, which made this work possible. We thank Marco Tori (CDC) for his helpful comments and suggestions. A.I.B. was supported by NIH AWD00009424.

## Author contributions

A.I.B. conceived and designed the study, curated the data, conducted the analyses and wrote the first draft manuscript. A.I.B. and L.W.A., visualized the data and iterated on the early drafts of the manuscript. L.R., A.P., E.A.S curated and provided context to the South Carolina data. N.H., A.P., E.A.S. and L.R. contributed to interpretation of the findings, commented and provided edits to the manuscript. A.I.B. was responsible for funding acquisition. All authors approved the final version.

## Ethics declarations

This study analyzed aggregate, school-level vaccination coverage and outbreak-exposure counts. No individual-level records, personal identifiers or protected health information were obtained, accessed or analyzed at any stage, and no human participants were enrolled or contacted. The work therefore does not constitute human-subjects research and did not require institutional review board approval or informed consent. All coverage data, including the South Carolina school file, are aggregate public records published by state education and health agencies under their routine open-data programs. The South Carolina coverage figures are the department’s 45-day school reports, published at https://dph.sc.gov, and the exposure flags were reconstructed from the department’s public outbreak communications; both are public records, and the geocoding is ours. Exposure flags record whether a school was named in a public health investigation, not which individuals were involved.

## Supplementary Information

## Supplementary Methods

This section specifies the model, the estimators and the implementation. It is separated from the Supplementary Notes, which defend design choices and report sensitivity analyses, so that every quantity needed to reproduce the analysis is in one place. Nothing here is a robustness check; nothing in the Notes is needed to define the analysis.

### The allocation unit

The allocation problem presumes a unit at which a dose can be targeted and at which susceptibility aggregates into transmission risk. Three lines of evidence support the school.

Household contacts cannot outnumber school-based contacts, whatever their vaccination pattern. Vaccination decisions are made per household, so an unvaccinated child’s siblings are more likely than average to be unvaccinated as well, and a household with one susceptible child may contain others. That correlation does not disturb the ordering the allocation relies on, because it sets a handful of contacts against a much larger number. In this panel the median susceptible kindergartener shares a grade with 18 other susceptible children, weighting each school by the susceptibles it holds (quartiles 7 and 65; mean 112, inflated by a small number of very large schools), and 82% share a grade with at least five. No household holds that many children, so for essentially every susceptible child in the panel the school is the larger susceptible aggregation whatever the vaccination status of that child’s siblings. Sibling contacts are also largely nested within the school, because enrollment follows residence and similarly-aged siblings are therefore drawn into the same catchment, so household transmission among them occurs inside the unit doses are delivered to. This parallels pre-vaccination patterns, for a reason specific to the sibling layer: a susceptible child’s similarly-aged siblings were usually susceptible too, though adult immunity was the result of near-universal childhood infection^1,2^.

Schools concentrate risk. The 5-to-17-year cohort holds the highest concentration of unvaccinated individuals in the contemporary United States, because compliance is operationalized at school entry and hesitancy is geographically correlated within catchment areas. Within a school, susceptible children are in prolonged contact, of the order of six hours a day for roughly 180 days a year, with a peer group drawn from that same catchment and therefore sharing its coverage. High susceptible density, long contact duration and age-homogeneous mixing make the school the unit at which susceptibility is most likely to exceed the local epidemic threshold. The age distribution of recent cases is consistent: in the 2025-26 South Carolina Upstate outbreak 64% of cases were school-age (5 to 17 years) and 26% were under 5, and nationally in 2025, 38% were school-age (5 to 19 years) and 31% were under 5^3^.

Contact structure within schools is heterogeneous but does not undermine the unit. Sensor-based measurement of a US high school found a non-uniform contact degree distribution with coefficient of variation 0.118, which under the standard heterogeneous-mixing correction R = R_homogeneous_ (1 + CV²) raises the reproduction number by about 1.4% above the homogeneous-mixing estimate, together with high modularity by classroom and grade^4^. Analysis of influenza in Japanese primary schools found that within-class and within-grade transmission jointly set the within-school reproduction number, with class size having little influence^5^. Sub-school structure therefore modulates the within-school final size we compute but does not displace the school as the unit at which a dose is delivered and at which coverage is recorded.

### Spatial scope of the distance-decay parameter

The published school-level *R_v_* uses a gravity kernel with decay *b* = 10^−3^ km^−1^, a characteristic length of 1,000 km. That is the appropriate scale for the question it was estimated for, which concerns coupling between communities, districts and counties across the United States^6^. Our allocation problem is posed within a state, and at that scope the same parameter behaves differently, in two ways we quantify here.

First, the geometric term *g*_j_ is nearly constant within a state: its coefficient of variation is 0.064, so *R_v_* becomes close to a rescaling of the raw susceptible headcount, ranking schools at Spearman *ρ* = 0.984 against a simple count of unvaccinated children. Second, the form *R_v_*(*j*) = *βN*_j_*S*_j_*g*_j_ is *separable*: a school’s *R_v_* depends on its own enrollment and susceptibility and on a purely geometric weight, not on its neighbors’ coverage. For the allocation problem this is an advantage, since it is the condition under which the marginal value of a dose at one school can be evaluated independently of the allocation elsewhere and the greedy rule is exact. It does mean the diagnostic does not by itself capture between-school herd protection, which we therefore test separately.

For the within-state allocation analyses we set the decay to *b* = 0.02 km^−1^ (50 km), a scale closer to that on which school-age contact operates. This is a change of spatial scope to match the question, not a correction of the published estimate, which we reproduce exactly at its own parameter value (Supplementary Methods). Supplementary Fig. 25 shows how the geometric variation and the rank correlation against raw headcount change with *b*.

### Kernel form and growth-rate calibration

#### Exponential rather than power-law decay^4^

The two common choices for a spatial transmission kernel are exponential, exp(−*b d*), and power-law, *d*^−*γ*^. Power-law kernels arise from mobility models with heavy-tailed displacement and suit continental-scale models. The exponential kernel corresponds to contact probability decaying at a constant fractional rate with distance, and is the appropriate choice here for three reasons: at the 1 to 50 km scales relevant to school-to-school transmission, school-entry contact data are well described by exponential decay; the exponential kernel gives a finite, analytically tractable spectral radius for the next-generation matrix, whereas an unnormalized power-law on a finite lattice yields a spectral radius that depends sensitively on the domain boundary; and it is consistent with the Haversine metric used for inter-school distances^7,8^.

#### Population-size exponents

The general gravity form is 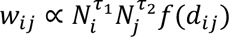. We use *τ*_1_ = 0 and *τ*_2_ = 1. The linear scaling in the source population is not an imposed assumption but a consequence of the density-dependent formulation: the force of infection at *i* is *βS_i_* ∑_j_ exp (−*b d_i_*_j_)*I*_j_ with *I*_j_ an absolute infective count, and in a fully susceptible population *I*_j_ ∝ *N*_j_, so contact volume from *j* scales exactly as 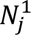.

#### Growth-rate estimates are a lower bound on *R*_0_, not a point estimate

In populations with structured within-group transmission, the reproduction number inferred from an observed exponential growth rate via the Euler-Lotka equation does not equal the true *R*_0_; it systematically underestimates it when within-group transmission is intense relative to between-group transmission^7^, because early growth is partly driven by rapid within-group spread rather than by independent chains of between-group transmission. Applying naive mass-action estimates in populations with strong household or school structure biases both *R*_0_ and the implied critical vaccination fraction^8^. This is the correct reading of Supplementary Fig. 10d. The growth windows bracket *R*_0_ = 15 in 8 of 15 cases and fall below it in the remaining seven; none falls above it. That one-sided pattern is the signature the bias predicts: within-school clustering deflates the observed between-school growth rate *r*, so the naive estimate 1 + *rT*_g_ sits below the true *R*_0_. Fits consistent with *R*_0_ ≈ 6 to 15 in these data are therefore consistent with a true *R*_0_ ≥ 15, and the windows bound the calibration from below rather than contradicting it.

### Replication of the published risk model

We re-derived the school-level reproduction number from the deposited coverage files using our own implementation and compared against the deposited estimates. For California 2019-20, the state-year with the most schools in the deposit and the row reported in Supplementary Table 2, the maximum absolute deviation is 5.4 × 10^−13^. The deposit covers 26 states, a set that overlaps but does not coincide with the 28 jurisdictions of our analysis panel, and we take one state-year per state, the one with the most schools. Across all 26 of those usable state-years the minimum correlation is 0.99945 (Hawaii 2014-15) and the median ratio of deposited to reproduced values is 1.000000 to six decimal places in all 26 (the largest deviation from unity across the 26 state-years is 3 × 10⁻¹⁵, at floating-point precision).

Residual differences of up to approximately 0.09 concentrate almost entirely at schools reporting exactly 100% coverage, whose deposited *R_t_* is not identically zero, consistent with coordinate and coverage rounding in the deposit rather than a difference in the formula.

Two implementation details are recorded for anyone reproducing this. The deposited pipeline sets *S* = 1 − *f*, folding vaccine efficacy into reported coverage, so the replication passes *VE* = 1 while all other analyses use VE = 0.97 explicitly. And *g*_j_ sums over every school in the state-year, so rows must not be filtered before it is computed, dropping one school perturbs *g* for all the others.

### Risk weights: construction, normalization and capping

#### What the weight cap does and does not do

Capping relative risk at 20-fold is often motivated as preventing extrapolation beyond the fitted sample. That motivation does not survive checking. Of 36,031 national schools, 99.6% fall inside the South Carolina linear-predictor range, and only 1.4% of susceptibles sit below the fitted coverage range. The relative risk implied within the 424 fitted Upstate schools spans roughly 780,000-fold, and the national spread is smaller at about 90,000-fold (Supplementary Table 46). The cap therefore regularizes a spread the fit itself implies from 30 events; it does not correct an out-of-support projection. We retain the cap and say what it is doing.

Note also that the cap is applied to the raw relative risk before normalizing to unit mean. If exp(*η*)*^α^* mostly sits far below the cap, dividing by the mean can return normalized weights well above it, so a nominal 20-fold cap does not bound the weights used. We cap after normalization as well, which is what makes the bound bind.

### Scoring rules under uncertainty: the choice of denominator

#### The denominator matters

A natural approach scores each candidate against the best rule on a fixed menu. That is misleading, because it rescales every rule by whatever the menu happens to contain: adding or removing one candidate changes every reported percentage. We score instead against the attainable benefit, the marginal-efficiency ranking re-derived at the modeled *α*. Under the menu denominator, a half-budget equity floor reads as 78% of the best menu rule at *α* = 1; against the attainable optimum it is 68%. No rule on the menu exceeds 72% of attainable at *α* = 1, which is the honest statement of how much a fixed rule gives up.

### Test for between-school herd protection

Because the separable *R_v_* cannot express a neighbor effect, we tested for one with a two-level stochastic metapopulation: within each school an outbreak runs to its SIR final size at *R*_0_*S*_j_, and onward seeding between schools occurs with probability proportional to *D_i_*_j_ and the source infection count. Seeding 1,200 random California schools (15 replicates each) and regressing *log*(1 + *outbreak size*) (case count, not enrollment) on own and kernel-weighted neighbor coverage, controlling for enrollment and density, own coverage dominated and the neighbor term was not significant.

### Interrupted time series

For California we fitted a segmented regression to the annual state series,

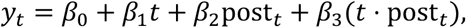

where *post_t_* indicates years from 2016-17 onward, *β*_1_ is the pre-policy trend, *β*_2_ the level change at the interruption and *β*_3_ the change in trend. Note that in this analysis *β*_0_, *β*_1_, *β*_2_, and *β*_3_ represent components of the regression model and are not the same *β* as is in the context of force of infection. SB277 took effect in 2016-17, the interruption point; the signing year and excluding the 2020-21 pandemic year were robustness alternatives. Standard errors are Newey-West with lag 1. Outcomes were enrollment-weighted mean coverage, the enrollment-weighted Gini of susceptibility, the share of susceptibles in the bottom 5% of schools, and the percentage below 95%. Placebo fits used the same model at the same breakpoint in the three other states with ≥3 pre-2016-17 years and no mandate change. Because ten annual observations and four parameters leave six residual degrees of freedom, Newey-West standard errors are anticonservative here, so we also report an exact permutation p-value obtained by refitting at every admissible interior breakpoint and comparing the observed level change against that null distribution. With seven admissible breakpoints the smallest attainable one-sided p-value is 1/7, which bounds what any single-series design can establish.

For the 18 states with ≥4 reported years we fitted ordinary least squares annual trends with Newey-West standard errors.

### Flag completeness and sector

The exposure flag records schools that a health department investigated and linked to a case, which is not a census. We bounded incompleteness by adding hypothetical missed exposures and assigning every one outside the top decile, the assumption that reduces enrichment fastest, and recorded where the Clopper-Pearson lower bound reaches 1. For sector we classified schools as private by equality against the reported private category rather than by inequality against public, because the file also contains a state-run special-education category that an inequality test would fold into the private group. Expected private exposures were taken as the sum of fitted probabilities from the size-and-coverage model over private schools.

### Cost model

The economic quantity is the difference between two allocations at an identical budget, not the burden of measles. Writing *c* for cost per case, the conditional value of the difference between the optimal strategy and lowest-coverage-first is

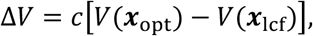

which inherits the conditioning of *H*: it prices infections averted given that an introduction reaches a school. Multiplying by the per-school seasonal introduction probability *p* gives the expected value, and subtracting dose cost at price *κ* gives expected net benefit (NB),

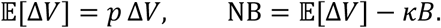

Three consequences follow. The ratio *V*(***x***_opt_)/*V*(***x***_lcf_) is free of both *c* and *p*, so the efficiency gain is invariant to either; Δ*V* is linear in *c*, which is the limitation a fixed-plus-incremental cost structure would remove; and *κB* enters unconditionally while benefits are conditional, so NB can be negative at small *p* even where Δ*V* is large. Conditional net benefit is positive in all 45 cost-by-dose-price-by-budget scenarios, at US$70 to US$412 per infection averted; expected net benefit is negative in 2 of 16 probability-by-perspective combinations, both at the smallest *p* under the two narrowest perspectives (Supplementary Note 8, Supplementary Tables 20, 21 and 76).

Four per-case perspectives are reported: incremental response cost (US$16,197), mean observed response cost (US$43,203), societal cost including hospitalization, productivity loss and mortality (US$104,629)^9^, and the largest observed (US$243,614). The sensitivity factorial adds a fifth, lowest-published level (US$6,973) and crosses all five with three dose prices (US$30, US$60, US$120) and three budget fractions, giving 45 scenarios (Supplementary Fig. 8).

### Introduction probability and expected value

The herd effect H estimates infections averted conditional on the pathogen circulating in the population-in this context, an introduction reaching a school, so the summed gap across all schools is a conditional quantity and the ratio of two rules is invariant to the introduction probability. To report expected rather than conditional costs we multiply the gap by a per-school seasonal introduction probability estimated two ways. The first decomposes into the probability a school is exposed given an introduction in its county, taken from the South Carolina within-region attack rate, and the rate of coded introductions per county; we report this with both a national county denominator and one restricted to states that reported introductions, since the restriction double-counts conditioning already present in the attack rate. The second decomposes into the statewide school attack rate and the fraction of states reporting cases. Neither is a precise estimate, and they differ by an order of magnitude, which is the resolution a single outbreak with adequate school records supports; all expected-value figures are reported across the full range. Dose cost is incurred regardless of whether an introduction occurs, so expected net benefit subtracts it in full.

### Minimax rule selection

Because *α* is not identified from a single outbreak, we treated rule choice as a decision under uncertainty. Seven candidates were evaluated at each *α* ∈ 0,0.1,0.2,0.25,0.3,0.4,0.5,0.6,0.75,1: the unweighted dose-optimal rule, lowest-coverage-first, equity floors reserving 25%, 50% and 75% of the budget for schools below *f̄* (largest susceptible pool first, remainder spent by marginal efficiency on residual need a hedged rule whose ranking is derived once at *α* = 0.25 and then fielded unchanged, and the minimax-regret design selected below. Each rule’s benefit at a given *α* was divided by the attainable benefit, the ranking re-derived at that same *α*, and we report both the minimum of that ratio and its complement, worst-case regret, over the grid. The minimax-regret design was selected by searching a one-parameter design family that blends the uniform and risk-weighted objectives linearly, (1 − *λ*) + *λw*, over *λ* ∈ [0,1]; a linear blend places a floor of (1 − *λ*) on every school, whereas a tempering exponent *w^λ^* shrinks all weights multiplicatively, and the blend dominates the exponent on worst-case regret at every *λ* tested. Scoring against the best rule on the menu rather than the attainable optimum inflates every rule by an amount that depends on what was omitted from the menu. The ranking was repeated at 10% and 50% budgets and under Firth-penalized coefficients.

## Supplementary Note 1 | The convex-region classification

### Sensitivity of the convex-region result

In this work, we use two symbols to describe related metrics of the indirect benefits of vaccination. H denotes the herd effect, the deterministic expected benefit on continuous coverage, which approaches the classical threshold from below as the assumed introduction vanishes; G denotes the same expected benefit where children come in integers, on the achievable coverage lattice and under exact stochastic seeding. For both, the convex to concave inflection *f̄* depends on the basic reproduction number and on the initial infected fraction. The dependence on *i*_0_ is qualitative rather than incidental: as *i*_0_ → 0 the convex region collapses entirely, because the herd effect H of a population with no active outbreak is concave everywhere. It is worth being precise about what i_0_ represents, because it does two jobs at once and only one of them affects the shape of G. The first is the conditioning event: whether a case arrives at all. That is a probability, it multiplies G, and because it does not vary with coverage it cannot move the inflection-scaling G by any constant from 1 down to 10^-6^ leaves the inflection unmoved. The second is the seed mass: how large the introduction is relative to the school, which enters the final-size relation and therefore does bend the curve. So the inflection is common to all schools in our primary specification not because exposure is assumed present, but because we hold the seed fraction fixed rather than the seed count. Fixing the fraction at 10^-3^ is arithmetically one index case in a school of 1,000; the median school in this panel has 69 children, so seeding one index case per school makes the seed fraction school-specific and the inflection with it. The convex region is a property of a population facing an introduction, which is the situation the allocation problem addresses.

Across a 3 × 3 grid of *R*_0_ (12, 15, 18) and *i*_0_ (10^−4^, 10^−3^, 10^−2^), *f̄* ranges from 0.898 to 0.966 and the national convex share ranges from 54.0% to 89.0%. It never falls below half. No second function is involved in any of this: H takes no school-size argument, so the school-specific inflection is this same H evaluated at a different i_0_, and there is no other convex-to-concave object in the model. Fig. 2b of the main text plots the consequence for the convex share, with the *i_0_* = 1/N_j_ result as a reference line; Supplementary Fig. 27a plots the curves themselves, which is where the moving bend and the nearly fixed peak are visible together.

### Fixed versus school-specific seeding

The herd effect H depends on the initial infected fraction *i*_0_, which is not observed. The main analysis fixes *i*_0_ = 10^−3^ for every school. This value is our own choice rather than one carried over from Duijzer et al.: their worked example uses three populations of 10,000, 20,000 and 40,000 with initial infected fractions of 0.015, 0.012 and 0.010, whereas what we take from that work is the qualitative condition that a convex region exists only under an active outbreak. The sensitivity range in Supplementary Table 5 brackets our fixed value and spans their example. An alternative is to seed each school with a single index case, *i*_0_ = 1/*N*_j_, which is arguably more faithful: an introduction is one child, and the median school here holds 69 kindergarteners, the grade the coverage records cover, so a K-5 campus around it holds roughly six times that, so 1/*N*_j_ = 0.014 at the median, an order of magnitude above the fixed value.

Under school-specific seeding (H) the inflection *f̄*_j_ becomes school-specific, ranging from 88.4% to 95.4%, though that range is the deterministic one and understates the effect of small school size, and the convex share falls from 75.8% to 68.5% of susceptibles and from 46.0% to 33.0% of schools (Supplementary Table 7). The qualitative claim is unchanged: a clear majority of susceptible children still sit where a marginal dose is in the convex region, and 25 of 28 states remain above half rather than 28 of 28. A separate question is the scale of the exposure expectation, which changes the expected return per school without touching the inflection at all. The per-school seasonal probability estimated in the expected-value analysis is constant across schools, so it cancels from the efficiency ratio and the reported gain is insensitive to it, while the risk weights examined elsewhere vary with susceptibility. Neither varies the exposure expectation with school size in terms of enrollment, and a larger school is a larger target. We therefore report a third sensitivity family: introductions arriving per school, per child (in proportion to enrollment), and an intermediate square-root scale. Re-deriving the ranking on each, the advantage over lowest-coverage-first is preserved and rises slightly, from 1.92-fold under a per-school expectation to 1.95-fold under the square-root scale and 2.02-fold under a per-child expectation. What changes is which schools are funded: at a 25% budget the per-child scale funds 7,118 schools against 13,036 under a per-school expectation, of larger mean enrollment (183 against 112), and the two funded sets overlap by only 41.0%. Applying the unweighted ranking to a per-child objective attains 66.3% of what re-derivation achieves, the same failure mode as an unweighted ranking under a susceptibility weight. We do not estimate which scale is correct: exposure is observed at county level and school size correlates with almost everything, so the outbreak record cannot separate them. This family is therefore a sensitivity bound rather than a calibration, in the same sense as the tempering exponent α in Supplementary Note 5: introductions arriving per school and per child are the two ends of the plausible range, so across that family the gain lies between 1.92- and 2.02-fold and the unweighted ranking retains at least 66.3% of attainable benefit. We retain the fixed seed fraction *i_0_* = 10⁻³ as primary and report the school-specific figure alongside it in the abstract and in Fig. 2b, because the fixed value is the one the source theory is stated with and the more conservative choice would be arbitrary in the other direction. One caveat on that specification. Substituting *i_0_* = 1/N_j_ into the deterministic final-size relation is not the same object as seeding one stochastic index case in a school of N_j_: the deterministic relation has no extinction, so it cannot represent the chance that an introduction fails to establish, which at kindergarten enrollments is the dominant term. Solved exactly, the inflection under a single index case falls to 78.4% at N = 100 and 72.2% at N = 50 (Supplementary Fig. 28), between 9.2 and 28.4 points below the deterministic school-specific value. The expectation is solved by first-step analysis on the (s, i) chain, cross-checked against exhaustive path enumeration at small N and against a Sellke-construction Monte Carlo; the inflection is unchanged if the direct credit is defined differently, because subtracting a linear function of coverage cannot alter a second derivative. A second consequence of small N is arithmetic rather than epidemiological. The integer nature of humans means that the continuous herd effect H is not so continuous, and G is what a school faces, and expected benefit depends on the introduction regime. Coverage at these enrollments is an integer lattice: one child is worth 100/(VE x N_j_) percentage points, 1.49 points at the median school of 69, so in 39.2% of schools, holding 16.5% of susceptibles, a single child moves coverage further than the whole 1.80-point interval between *f̄* and *f̃*. The lattice does not threaten the population-level result: re-classifying every school at its nearest achievable coverage moves the convex share of susceptibles from 75.8% to 76.0%. What it limits is the per-school split, which is therefore reported as a population summary rather than as a resolvable classification of individual schools (Supplementary Fig. 27b, d).

One property of *H* prevents school-specific seeding being adopted wholesale. *H* is defined net of the directly immunized, *H*(*f*) = [*Z*(0) − *Z*(*f*)] − *fVE*(1 − *i*₀), so it measures indirect protection only. This is the definition used throughout, including in the main-text Methods, where the (1 - i0) factor on the direct term was omitted in error; it is linear in f and leaves both critical fractions and the allocation ordering unchanged. The direct credit is netted out scaled by the seeded susceptible fraction rather than as the unscaled product, which keeps the indirect benefit non-negative at every coverage level and leaves the dose-optimal target well defined at every enrollment in the panel: its argmax does not fall below 0.9485 for any distinct enrollment between 10 and 60 pupils. Under the unscaled definition, the target collapsed to zero at *i*_0_ above 0.026, schools of 38 pupils or fewer, affecting 7,646 schools (21.2% of the sample, 7.6% of susceptibles); the scaled credit removes that degeneracy. The underlying magnitude is real even so: at those enrollments the average return *H*(*f*)/*f* is nearly flat, because in a 20-pupil school seeded with one case an *R*_0_ = 15 outbreak infects essentially every susceptible whatever the coverage, so the herd effect there is small. What limits a per-school allocation under this specification is therefore the integer lattice rather than the objective: every school of 38 pupils or fewer has a one-child coverage step of at least 2.71 points against an interval of 1.80 points between the two thresholds, so the per-school split is not resolvable at any enrollment where the concern would arise. That is why the classification, not the allocation, is what we report here.

### Gini on susceptibility, not on coverage

The interrupted time series reports an enrollment-weighted Gini coefficient of susceptibility, 1 − *f*, not of coverage. The distinction matters when comparing with the previous study^6^, which reports coverage Gini values of approximately 0.046 at school level. Our susceptibility Gini for California ranges from 0.51 to 0.63 across the series (Supplementary Table 60). The two are not in conflict; they are different quantities.

Coverage in the United States is compressed near the ceiling, roughly between 0.85 and 1.00, so a Gini computed on coverage is bounded far below the conventional income-inequality range and its absolute magnitude is not interpretable on that scale. Normalizing by mean susceptibility recovers the informative quantity: the previous study reports a relative Gini of 0.62 at school level, 0.49 at district level and 0.36 at county level. Our susceptibility Gini is the same construction applied directly, which is why our values sit near their school-level relative figure.

For the allocation problem susceptibility is the correct base, because the quantity being allocated against is the susceptible headcount rather than the coverage percentage.

### Which curve is convex where allocation has leverage, and which is sigmoid

Three quantities are easy to conflate, and only one of them is the objective this paper optimizes. Two symbols carry the expected benefit, deliberately: *H* is the deterministic herd effect on continuous coverage, whose inflection approaches the classical threshold *f*^∗^ from below as the assumed introduction vanishes; *G* is the same expected benefit where children come in integers, on the achievable coverage lattice and under exact stochastic seeding, and it is *G* that Supplementary Figs. 26d, 27 and 28 report. The direct component is the vaccinated fraction times efficacy, *f* VE: it is linear by construction, with numerical second derivative at floating-point zero. Total infections prevented, *T*(*f*) = *Z*(0) − *Z*(*f*), is the whole reduction in final size. The indirect benefit is the difference, *H*(*f*) = *T*(*f*) − *f* VE(1 − *i*₀), the infections averted beyond the recipients themselves, and it is *H* that the allocation objective sums over schools.

None of these three is a sigmoid in the strict sense of a monotone, bounded curve with a single convex-to-concave bend, and the allocation objective is the furthest from one. It is single-peaked: convex below its inflection at *f̄* = 94.4%, concave above it, reaching its maximum of 0.0558 of the population at the critical fraction *f*^∗^ = 96.2%, and declining above that *f*^∗^ is the herd-immunity threshold proper and lies above the inflection, so further coverage buys no additional indirect protection while the direct term keeps growing, and the difference declines. Total prevented shares that same inflection, since it is the indirect benefit plus a linear term; the two second derivatives agree to 5.6 × 10⁻⁹. A genuinely sigmoidal quantity exists in this system, and it is none of the three: the probability that a remaining susceptible escapes infection, 1 - Z(*f*)/s_0_(*f*), is monotone increasing, bounded in [0, 1], and has a single convex-to-concave bend at *f* = 95.4%, which is the textbook shape. It is not the allocation objective, because a program allocating doses cares about infections averted per dose, not about the escape probability of whoever remains unvaccinated. The distinction matters for wording only: every result in this paper rests on the allocation objective being convex below its inflection, which it is, and not on its being sigmoidal, which it is not.

A reader may expect *T* to rise roughly one-for-one at low coverage, on the reasoning that each early dose removes a susceptible who would otherwise have been infected. That expectation is correct, and it is why total prevented is the curve that looks nearly linear at low coverage while the indirect part is nearly flat. At *R*_0_ = 15, the marginal return on total prevented is 0.9691 at *f* = 0.25, close to vaccine efficacy, and 0.9719 at *f* = 0.50, rising only to 1.4778 at *f* = 0.95 as herd effects begin to repay more than the recipient alone (Supplementary Fig. 1, Supplementary Table 1). What is nearly flat at low coverage is the indirect part, whose marginal return is 0.0001 at *f* = 0.25 against 0.5087 at *f* = 0.95.

The distinction is not cosmetic. Because *H* subtracts a term linear in *f*, and because that term is identical across allocations at a fixed budget, the fold-gain between two rules measured on *H* exceeds the fold-gain measured on total cases: 1.918-fold against 1.161-fold in this analysis. Both are reported in the main text and Note 2 gives the arithmetic. The convex-region argument runs on *H* because that is where allocation has leverage; the total-cases figure is what a program would observe.

## Supplementary Note 2 | Allocation rules, comparators and objectives

### The allocation result does not use the risk model

The convex classification and both allocation rules are functions of reported coverage and enrollment only. Neither invokes the gravity kernel, the distance matrix or *R_v_*. To demonstrate this rather than assert it, we recomputed every state’s allocation after deleting the coordinate columns entirely, so that no kernel can be formed: the benefit of each rule is identical to machine precision in all 28 states, and the national efficiency gain is 1.918 either way (Supplementary Table 14).

This bounds the reach of the risk-model limitations reported in Notes 5 and 8. The gravity-kernel *R_v_* is used for two things only: the outbreak validation, and the count of transmission-capable schools. The headline result, that 75.8% of susceptibles sit in the convex region and that marginal-efficiency allocation averts 1.92 times as many infections, does not depend on it.

### Two comparators: ordering versus stopping rule

A “lowest-coverage-first” program makes two choices: which schools to fund (the ordering) and how far to raise each funded school (the stopping rule). The two can fail independently and conflating them inflates any reported gain.

The herd effect *H* is single-peaked, with a maximizer *f*^∗^ = 96.2% under *R*_0_ = 15, *VE* = 97% and *i*_0_ = 10^−3^. Doses delivered above *f*^∗^ therefore buy strictly less benefit than the same doses delivered at *f*^∗^: a school taken to *f* = 99.9% receives more doses and gains less than the same school taken to *f*^∗^. A program that funds the lowest-coverage schools *and* takes each to full coverage makes both errors at once.

We report the two separately (Supplementary Table 13, Supplementary Fig. 5). Holding the target fixed at *f*^x^ for both rules, ordering alone accounts for a 1.92-fold difference nationally. Letting the comparator overshoot to full coverage costs a further 2.57-fold, and the two compose exactly: 1.92 × 2.57 = 4.94. The headline result in the main text is the ordering effect, because that is what Duijzer’s theorem speaks to and because it is the smaller of the two. The overshoot penalty is reported as a separate quantity, since a real program choosing to level schools all the way up would incur it in addition.

The two rules must share a target for the comparison to isolate ordering.

### Two objectives: indirect benefit and total cases averted

The allocation objective in this paper is the indirect benefit of a dose: infections averted beyond those prevented by immunizing the recipients themselves. That is the quantity allocation theory speaks to, because the direct effect of a dose does not depend on where it lands, and it is the quantity the herd effect *H* measures, with the directly immunized fraction subtracted.

A reader could reasonably ask about total cases prevented instead. Because every dose immunizes children wherever it goes, the direct component is a constant across rules: at the national 25% budget it is 72,503 children directly immunized under every rule tested, identical to machine precision. They are counted as protected, not as infections averted, since not all of them would have been infected. The two objectives therefore give different ratios from the same allocations:

| Rule | Indirect only | Total cases prevented |
| --- | --- | --- |
| Dose-optimal | 29,479 | 101,983 |
| Equity floor 50% | 23,889 | 96,393 |
| Lowest-coverage-first | 15,366 | 87,870 |
| Dose-optimal / lowest-coverage-first | 1.92-fold | 1.16-fold |

Subtracting a constant *c* from both sides of a ratio increases it whenever the numerator exceeds the denominator, since (*A* − *c*)/(*B* − *c*) > *A*/*B* for *A* > *B* > *c*. Roughly a third of the 1.92-fold figure is this arithmetic rather than a difference in what the rules achieve in total. Neither number is wrong: the indirect ratio answers “how much more herd protection does the ordering buy”, the total ratio answers “how many fewer children are infected”. We report the indirect figure as the headline because it is the theorem’s object and because the rules are being compared on the mechanism, they differ in, and we report the total figure here so a program evaluating total burden uses the smaller one. The direction is the same under both, and the choice of objective does not affect the classification result (75.8% convex) or the validation, neither of which involves a ratio of benefits.

### An equity floor retains most of the gain

Health departments are frequently constrained, by statute or by policy, to prioritize the lowest-coverage jurisdictions. We therefore evaluated a hybrid rule: ring-fence a fixed share of the budget for the lowest-coverage schools, allocate it lowest-coverage-first, and apply the marginal-efficiency rule only to the remainder.

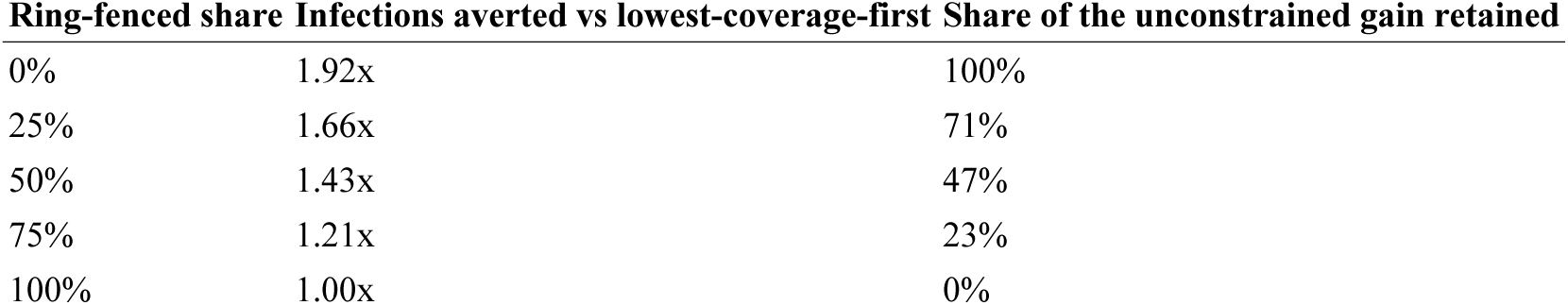

| Ring-fenced share | Infections averted vs lowest-coverage-first | Share of the unconstrained gain retained |
| --- | --- | --- |
| 0% | 1.92x | 100% |
| 25% | 1.66x | 71% |
| 50% | 1.43x | 47% |
| 75% | 1.21x | 23% |
| 100% | 1.00x | 0% |

Half the budget can be reserved for equity-driven allocation, and the discretionary remainder still averts 1.43 times as many infections as spending the whole budget lowest-coverage-first, retaining 47% of the unconstrained gain (Supplementary Fig. 7, Supplementary Table 18). The efficiency argument therefore does not require abandoning an equity constraint; it quantifies what each increment of that constraint costs, so the trade-off can be made deliberately.

## Supplementary Note 3 | Heterogeneous introduction risk

### The uniform-introduction-risk assumption

The allocation objective sums a per-school herd benefit, which prices a season in which every school faces an introduction with equal probability. The South Carolina validation contradicts that premise on our own data, and the contradiction deserves a stated bound rather than silence.

#### Two claims must be kept apart

Scoring the unweighted ranking against a risk-weighted objective makes it fall below coverage rank beyond a tempering exponent of about 0.33. That is not evidence against the criterion: any optimizer underperforms when scored on an objective it was not derived for. Re-deriving the ranking on the weighted objective, which changes only the benefit each school contributes and not the ranking criterion, beats coverage rank at every strength tested, from 1.37-fold to 1.92-fold (Supplementary Table 39). The correct reading is a scope condition: marginal-efficiency allocation is dose-optimal for whichever objective a program adopts, and a program that believes introduction risk tracks susceptibility should re-derive the ranking under that belief.

#### The magnitudes are a bound, not a calibration

The risk model is fitted on 424 schools in one region, mostly between 57% and 100% coverage, and the untempered weights span roughly a 90,000-fold range when applied nationally, against 780,000-fold within the fitted sample itself (Supplementary Table 46). We therefore report the whole tempering path and draw conclusions only from its shape.

Independent support for the underlying premise comes from Harris et al.^10^, whose assortativity estimator reproduces on this panel across 17 states they did not have, median 0.35 against their 0.37 (Supplementary Fig. 17a, Supplementary Table 48). If unvaccinated children cluster, contacts are not random and introduction risk is not uniform, which is exactly the condition under which the re-derived ranking rather than the unweighted one is the operative recommendation. Under their closed form the dose-optimal rule raises the enrollment-weighted breakthrough share more than lowest-coverage-first, 0.490 against 0.457 from a baseline of 0.434 (Supplementary Tables 49 and 50); total infections fall under both, but a rule that raises the vaccinated share of cases while cutting their number is harder to communicate, which is a presentational cost rather than an epidemiological one.

That work is a preprint and has not been peer reviewed, so nothing in the main text rests on it. It is reported here as corroboration of a premise the paper establishes independently from its own exposure data, and the two quantities we take from it, the assortativity estimator and the breakthrough closed form, are recomputed on our panel rather than quoted. The threshold correspondence in Supplementary Fig. 16c is a separate matter and does not depend on it: *p_c_* there is the classical vaccine-adjusted herd-immunity threshold (1 − 1/*R*_0_)/*VE* of Anderson and May^1^, which the preprint also uses and attributes to the same source.

We computed the enrollment-weighted assortativity of Harris et al.^10^ on each state panel as *φ*_j_ = (*p*_j_ − *P*)/(1 − *P*) where *p*_j_ ≥ *P* and (*P* − *p*_j_)/*P* otherwise, with *P* the enrollment-weighted state coverage. Their breakthrough fraction *f_V_* = *pε*/(1 − *p*(1 − *ε*)) is algebraic in coverage and was evaluated at post-allocation school coverage under each rule. Their unimodal total-breakthrough curve is a whole-population equilibrium result and was not applied per school, since that would treat each school as closed and contradict the between-school coupling both analyses rest on.

### The introduction gradient is a within-county effect

The exposure model behind *α* is fitted to schools inside one reached region, so it estimates the probability that a school is exposed *given that an introduction reached its county*. Applying it to absolute national coverage implicitly assumes the same gradient operates between counties. That assumption is testable, and it fails.

Joining the school panel to county boundaries gives 1,476 counties with at least three schools across 28 states, of which 154 recorded 2025-26 cases. In a logistic model with state fixed effects and log enrollment, mean county coverage does not predict which counties were reached (−0.0075 log-odds per percentage point unvaccinated, *p* = 0.79). What does predict it is the within-county spread of school coverage (+0.059 per percentage point of standard deviation, *p* = 0.019, likelihood-ratio *p* = 0.020). Clustering, not the average, is the between-county risk factor (Supplementary Table 37). The same concentration is visible in the national exemption trends: the post-2020 rise in exemptions exceeded one percentage point in 53.5% of 2,842 counties and five points in 5.3%, a gradient invisible at state level.

This has two consequences. First, this is an independent empirical instance of the assortativity mechanism Harris and colleagues describe at the contact level^10^, appearing here at the county level: what matters is how unevenly susceptibility is distributed, not its mean. It is also what the metapopulation literature on spatiotemporal heterogeneity would predict, where dispersal combined with variance in local conditions inflates regional growth above the average local rate^11,12^. Second, the within-county gradient and the between-county gradient are different quantities, and our national weights use the within-county one. This is a limitation of the weighting rather than of the classification result, and it is a further reason to treat the *α* analysis as a bounded sensitivity: the unweighted school mean coverage of our panel is 91.5% (enrollment-weighted, 93.1% as reported in the main text) against 91.6% in the fitted Upstate sample, so the two are close in level, but nothing in the design establishes that the slope transfers across counties.

### State-level replication and the limits of the clustering signal

The preceding subsection reports the asymmetry nationally: between counties, mean coverage does not predict which counties were reached, but the within-county spread of school coverage does. A single national fit is one test. This note asks whether the same asymmetry appears inside individual states, at what spatial resolution it survives, and whether the signal is stable enough over time for a targeting rule to rely on. All four analyses use the same outcome as the national fit, at least one case in our own county series, so the coefficients are comparable.

#### Texas replicates; New Mexico is underpowered

In Texas alone (137 counties with three or more schools, 36 with cases), the within-county spread carries +0.301 log-odds per percentage point (*p* = 0.011) while mean coverage is null (−0.015, *p* = 0.85), the same asymmetry as nationally with a coefficient about five times as large. New Mexico returns nothing in either direction (+0.094, *p* = 0.76). That is a power limitation rather than a contradiction, and it is diagnosable: the entire New Mexico school panel holds 969 susceptible children across 419 schools, a third of its counties have cases, and log enrollment alone gives AUC 0.805 for being affected, so county size already accounts for the pattern and little is left for spread to explain (Supplementary Table 67).

California returns the same null for the same reason, and a third state makes the pattern legible^3^. In 53 California counties with 16 affected, the within-county spread carries −0.099 log-odds per percentage point (*p* = 0.41) and mean coverage is null; log enrollment is the only significant covariate (+1.34, *p* = 0.004) and separates affected from unaffected counties far better than the spread does, AUC 0.80 against 0.42 (Supplementary Tables 68 and 69, Supplementary Fig. 20). The three single-state fits are not a random scatter of successes and failures. What separates them is whether county size already accounts for the outcome, and the diagnostic is the gap between the two AUCs rather than either alone: enrollment out-predicts the spread by 0.48 in New Mexico and 0.38 in California, both states where measles reached the large metropolitan counties so that being reached is close to being populous, but by −0.02 in Texas, where the two are level and the spread coefficient is largest. Clustering is detectable where size does not already determine reach. This is a power and confounding account rather than a contradiction, and it is checkable rather than asserted. Two further features of the California test work against detection and are stated rather than buried: its most recently available coverage is 2022-23, more than two years before the 2025-26 cases, whereas the Texas fit uses the pre-outbreak year, and the California release censors coverage at 99%.

The same California file answers a second question, and the answer bounds a claim rather than supporting one. The private-school position reported for Texas does not generalize. Texas is monotone, private schools concentrating in the low-coverage tail and thinning in the band the dose-optimal rule targets; California shows no such concentration (25.2% of schools below 85% against 23.5% in the targeted band, odds ratio 1.09, *p* = 0.64), and on susceptibles the direction reverses (7.8% against 16.4%). Censoring does not explain the difference, because the ceiling can only move schools into the top band, which is where California private schools are rarest, while the comparison at issue involves only schools far below it. One state with a 34.8% private sector behaves one way and one with 13.8% behaves differently, so the sector position is a state-level property to compute locally, not a national regularity.

#### The signal does not survive aggregation to districts

Repeating the Texas fit on districts rather than schools attenuates the coefficient to +0.099 (*p* = 0.15). Units follow the reporting frame rather than geography: the named Texas release reports public provision at the level of the independent school district, so each ISD is already one row, and private schools and open-enrollment charters, which belong to no ISD, each remain their own unit. No private school is placed inside a public district, and no unit mixes the two sectors. The aggregation is therefore the one a jurisdiction would itself publish, and it is slight: of the 1,093 public rows, 966 are already whole ISDs resolving to 956 units and 127 are charters that each stand alone, while the 585 private schools resolve to 575 units. Only 19 units of 1,658 combine more than one row, so the attenuation reflects the coarser reporting resolution itself rather than an averaging step imposed here. This is expected rather than disappointing: a district average is computed over exactly the within-county variation the test is looking for, so aggregating to district scale removes the predictor. It matters because district resolution is what most states publish. The finding therefore has a resolution requirement, and jurisdictions reporting only district aggregates cannot evaluate it on their own data.

#### Significance depends on how many counties count as affected

The sign is positive at all six case-count thresholds tested, from at least one case to at least fifty, but the coefficient reaches *p* < 0.05 only where enough counties are affected to support the fit, failing below about ten (Supplementary Table 70). We report the threshold that matches the national definition and deposit the rest.

#### The Texas effect is specific to the pre-outbreak year

Six years of Texas coverage data are available, and the effect is significant in one of them, 2024-25, at *p* = 0.011 nominally and not after Bonferroni correction for six looks (threshold 0.0083); the other five range from +0.096 to +0.255, all positive and none were significant (Supplementary Table 70). The year we use was chosen on epidemiological grounds, being the last measurement before the outbreak, rather than selected on its result. But the honest statement is that this is a test of pre-outbreak clustering, not a time-invariant property of Texas coverage data, and that the single-state result is nominal.

#### A Texas replication of the private-school pattern

The 2023-24 named Texas file carries a sector field the panel does not. Private schools are 72.4% of campuses below 75% coverage and 47.8% of those between 75% and 85%, against 24.8% in the 85 to 94.4% band the dose-optimal rule targets and 34.8% overall (Fisher OR 4.26 for private in the sub-85% tail versus the targeted band, *p* = 9.3 × 10⁻¹⁹; Supplementary Table 71). This is an independent single-state replication of the national sector pattern, with a different source and year. It is a statement about where private schools sit in the coverage distribution, not about where transmission occurred.

#### Clustering is much less persistent than mean coverage

Matching California schools by name within county across the SB277 boundary (Supplementary Fig. 22) gives 5,393 schools in 42 counties with at least ten matches (Supplementary Table 58). County mean coverage is strongly persistent over the eight years (Spearman *ρ* = 0.54, *p* = 0.0003), but the within-county spread is only weakly so (*ρ* = 0.31, *p* = 0.044; enrollment-weighted *ρ* = 0.41, *p* = 0.007). A rule that targets on clustering therefore needs recent measurements: coverage means from several years ago remain informative about a county’s level, while its internal distribution does not. This is a practical constraint on the surveillance recommendation, and it cuts against, not for, the risk model.

#### The same question can be asked at the scale the rule acts on

The allocation rule ranks schools, not counties, and the primary panel tracks schools across twelve years, so the persistence of a school-level ranking is directly measurable. The keying, the exclusion of ambiguous coordinates and the definition of worst-decile retention are given in Methods. A one-year-old coverage ranking retains 52% of its worst decile (Spearman *ρ* = 0.57); at three years it retains 43% (*ρ* = 0.47) and at five years 39% (*ρ* = 0.43). The pattern is not driven by a few states: 16 of 17 states with enough consecutive years to test fall below 60% one-year retention, median 45% (Supplementary Fig. 26, Supplementary Tables 79 and 80). School ranks therefore decay faster than county means, which is the direction the county result already implied but could not show at this scale. The temporally external South Carolina test uses coverage measured one year before the outbreak and recovers a 5.05-fold enrichment, so the lag the paper actually validates is the one the data supports; a ranking built on a three- or five-year-old coverage file is a materially weaker instrument, and this bounds the surveillance recommendation rather than supporting it.

One state is excluded from that measurement, for a reason that is itself a reporting-quality finding. Averaged over consecutive-year comparisons, 99.5% of Louisiana schools carry identical coverage values while only 2.8% carry identical enrollment: the enrollment field updates and the coverage field does not, which is the signature of one coverage snapshot repeated across the four years the state contributes rather than of genuinely stable coverage. Left in, it returns a Spearman *ρ* of 0.99 and dominates the average; excluded, the one-year national figure moves from 0.576 to 0.568. The reporting-quality screen cannot detect this because every metric it computes is within a single year, and on all of them Louisiana is unremarkable (median coverage 0.929, 12.0% of schools at exactly 100%, 383 distinct values, comparable to Washington). California’s 47.6% identical fraction is a different and benign phenomenon, integer-rounded percentages on small denominators producing genuine ties and is not flagged because its enrollment field turns over normally. No reported result depends on this: the convex share, the fold gain and the cost figures are computed within a single year, where repetition across years cannot enter, and dropping Louisiana altogether moves the convex share from 75.8% to 75.6% and the fold gain from 1.918 to 1.922. The cross-year identical-value fraction is deposited for every state (Supplementary Table 82) so the defect is detectable rather than incidental.

The 2022-23 California file is capped at 99% with no school reported at 100%, a ceiling absent from the 2014-15 release, which has 1,273 matched schools at exactly 100%. Capping the earlier year at 99 to match moves the mean coverage change from +3.97 to +4.13 percentage points, so the ceiling accounts for 0.16 points and does not affect the persistence comparison, which is computed within each year (Supplementary Table 59).

## Supplementary Note 4 | Robust rule choice under uncertain risk scaling

### Choosing a rule when the risk scaling is unknown

Note 2 establishes that the allocation objective assumes uniform introduction risk and that our own data refute it. This note addresses the decision that follows: *α* is not identified from one outbreak, so which rule should a program field?

Both hedges must be on the menu. A program uncertain about *α* can hedge in two ways: fix a moderate equity floor or derive the marginal-efficiency ranking once at a central *α* and field it unchanged. Omitting the second makes the first look uniquely robust. With both included they are statistically indistinguishable at the top, 71.2% for a three-quarter floor and 78.7% for a ranking derived at *α* = 0.25, and both dominate the two extremes, 14.2% for the unweighted dose-optimal rule and 52.1% for lowest-coverage-first (Supplementary Table 78). The ranking is unchanged at 10% and 50% budgets, where the two-percent hedge holds 80.4% and 96.9% of attainable against 10.0% and 71.2% for the unweighted rule (Supplementary Table 43).

Penalization does not resolve it. The robustness is structural, not incidental. The two extremes fail for opposite reasons, and the reason is where each rule’s worst case falls inside the uncertainty set. The unweighted dose-optimal rule is at its best at α = 0 and degrades as the gradient steepens, so its worst case falls near the upper end of whatever set is assumed and every widening makes it worse: 14.2% of attainable over α ∈ [0,1], 10.6% over α ∈ [0,2], with the minimum at α = 1.5. The two-percent hedge is the mirror image: its worst case falls at α = 0 and its performance rises with α 89.7% at α = 0, 94.7% at 0.25, 95.6% at 1.0, 96.6% at 1.5, so its worst case is 89.7% on both [0,1] and [0,2] (Supplementary Table 42). A rule whose worst case is pinned to a fixed endpoint of the set is robust to how that set is drawn; a rule whose worst case chases the set’s far edge is not. Firth’s penalized likelihood, the standard correction for rare-event bias, shrinks the coefficient on percentage unvaccinated by 2.3%, from 0.1137 to 0.1112, and leaves the worst-case ranking intact, with the two-percent hedge at 89.7% of attainable under either estimator (Supplementary Tables 44 and 78). The uncertainty is not estimator bias that a penalty can remove; it is design imprecision from one outbreak in one region. A hierarchical fit would be the right tool given two or more outbreaks with linked school-level exposure and pre-outbreak coverage, and no such second dataset is currently public. That is why *α* is reported as an unmeasured quantity with a stated measurement path rather than calibrated away. The minimum is flat: worst-case regret is 11.6% at λ = 0.01, 10.4% at 0.015, 10.3% at 0.02 and 11.9% at 0.03, so every design weight from 0.01 to 0.03 sits within two percentage points of the optimum (Supplementary Table 40). What transfers is that a light hedge beats both extremes, not the value.

## Supplementary Note 5 | Outbreak validation

### Outbreak validation: design and interpretation

The school-level reproduction number estimates *amplification given an introduction*. It carries no information about where importations occur, which depends on travel, community connections to endemic regions, and chance. A regression of case counts on risk would therefore conflate two distinct processes and could show an association driven entirely by importation pressure.

We conditioned on introduction: only counties recording at least one case enter the analysis, and the outcome is secondary cases after the index report. We additionally ranked each county within its own state distribution, so that between-state differences in coverage levels and reporting practice do not contaminate the within-state comparison. A pooled analysis without this ranking returns null for every metric.

Only the susceptible-pool metric shows significant top-decile enrichment (3.3-fold, p = 0.0064). Overall discrimination between amplifying and fizzling counties is near chance, indicating that the signal is confined to the extreme tail. We report this rather than presenting the enrichment alone.

School-scale enrichment was 1.67-fold in our 2025-26 set (3 of 18 amplifying counties) against the 2.0-fold reported in^6^. This is not a replication failure. With 18 amplifying counties the one-sided binomial test against a null of 0.10 has a critical value of 5, so the design has only 28% power against a 2.0-fold effect, and the exact 95% interval on our estimate (0.36 to 4.14-fold) contains both the null and the published value (Supplementary Tables 22 and 23). The measurement gap documented in Supplementary Fig. 10c compounds this: the epicenter counties are where school-level records are sparsest.

### Enrichment stratified by data richness

The school-scale enrichment failure in the 2025-26 set could reflect the risk model or the records. We separated the two with a filter on reporting scope rather than a tuned threshold. Counties differ in how much of their observed spread their school records can resolve, and the ratio of secondary cases to recorded susceptibles measures that: the numerator counts cases at all ages while the denominator counts kindergarten records only, which is the reporting standard every state in the panel meets, so the two differ by roughly a grade span before any question of completeness arises.

Eight of 81 counties with an introduction sit above one on that ratio, including every Texas and New Mexico epicenter (Supplementary Table 25). Rescaling to a K-5 pool puts five of the eight below one and leaves three above it (Gaines and Terry marginally, at 1.08 and 1.06, and Hudspeth at 5.17); against a K-8 pool only Hudspeth remains, and its denominator is a single school. The ordering is therefore dominated by grade span rather than by missing records, and Supplementary Table 25 reports both scales. Gaines County, Texas is the extreme case, with 408 secondary cases against three recorded schools and 63 recorded susceptible kindergarteners. Its 408 cases exceed a six-grade rescaling of that pool, 378, by only 8%, which is the point: the ratio is a statement about reporting scope, and at a plausible grade span the county sits at the margin rather than far beyond it.

Among the 73 counties whose records are consistent with the spread they recorded, top-decile enrichment rises on every metric: school-scale from 1.67-fold to 3.0-fold (3 of 10 amplifying counties, p = 0.070) and county-scale susceptible pool from 3.3-fold to 6.0-fold (6 of 10, p = 0.0001; Supplementary Table 21). The direction is monotone in data richness across all thresholds examined (Supplementary Table 26).

Two readings are available and we state both. The favorable one is that where the vaccination data can account for the observed spread, the school-scale ranking works, and the published 2.0-fold enrichment is recovered within uncertainty. The cautious one is that the filter removes 8 of 18 amplifying counties, leaving 10, so the school-scale test is underpowered, and its p-value does not clear 0.05. We therefore report this as evidence that the failure is attributable to surveillance coverage rather than to the model, not as positive confirmation of the model at school scale.

### Non-independence of the exposure flags

The 30 exposed Upstate schools are not independent draws: they arise from one transmission chain, and 28 fall in a single county. Clustering by county inflates the coverage slope’s standard error from 0.0240 to 0.0310, a design effect of 1.67, leaving it at p = 0.0032 (CR1, t on 12 clusters); a county random intercept gives p = 0.0026 with a county standard deviation of 2.58 on the logit scale. The strictest available test restricts to the outbreak county alone, removing between-county variation by construction, and the slope rises from 0.114 to 0.186 (p = 0.00036); clustering further on six spatial neighborhoods within that county leaves it at p = 0.040. Moran’s I on the model’s Pearson residuals is 0.064 (permutation p = 0.035, 4,999 draws), so residual spatial correlation is present and weak, the same fact the design effect states. Cluster-jackknife (CR3) and wild bootstrap procedures are uninformative on this design and reported as such: with 28 of 30 exposures in one county, both effectively delete the only informative cluster (CR3 inflates the standard error 8.5-fold). Widening the interval accordingly moves the implied risk-scaling set from α ∈ [0.59, 1.41] to α ∈ [0.41, 1.59], inside the α ∈ [0, 2] set already analyzed, so no rule ranking changes.

## Supplementary Note 6 | Data, scope and provenance

### Why five states are excluded

School-level coverage is reported by state agencies under differing rounding and suppression conventions. In five states an implausible share of schools report coverage of exactly 100%: Maine (89.5% of schools), Maryland (72.4%), Arkansas (56.2%), Connecticut (50.7%) and North Dakota (44.3%)^3^. A distribution with most of its mass at a single boundary value is a signature of censoring or coarse rounding, not of genuine universal coverage, and it makes the susceptible pool unmeasurable for those schools.

We therefore excluded states where more than 40% of schools report exactly 100% coverage. The threshold is arbitrary, and the analysis does not depend on it: varying it from 20% to no exclusion at all moves the national convex share only between 75.6% and 75.9% (Supplementary Table 3, Supplementary Fig. 3b).

### What carries over from the previous study, and what differs

The multiscale vaccination database, the school-level *R_v_* metric and its validation were established in the previous Nature Medicine study^6^, which assembled records spanning 45 states and Washington DC and over 50,000 schools, 13,000 districts and 3,000 counties for 2013 to 2025. This work does not re-argue that assembly. The table below states what is inherited unchanged and what differs.

| Element | Previous study<sup>6</sup> | This work |
| --- | --- | --- |
| Data source | 45 states + DC, 50,000+ schools, 2013-2025 | Same assembly; 33 states have usable school-level files, 28 retained after the reporting-quality rule |
| Spatial scales | School, district, county | School only (the allocation unit) |
| $R_v$ formalism | Gravity-kernel next-generation matrix | Identical; reproduced to $5.4 \times 10^{-13}$ (Note 6) |
| Distance decay | $b = 10^{-3} \text{ km}^{-1}$ (national coupling) | $b = 0.02 \text{ km}^{-1}$ (within-state allocation); magnitudes not comparable |
| Question | Descriptive: where does risk concentrate? | Prescriptive: which school gets the next dose? |
| Validation outbreaks | Seven, 2013-2025 | 2025-26 set, 81 introductions in five states |

The narrower state footprint is a consequence of the exclusion criteria, not a different data source. Of the 33 states with school-level files, five report an implausible share of schools at exactly 100% coverage (Note 1); because the allocation rule turns on where a school sits relative to a coverage threshold, censored coverage makes those schools unplaceable, and they are dropped. Because the enrollment field is whatever each state publishes, its meaning was checked against an external source collected on one federal definition: NCES Common Core of Data grade-0 counts for the same 164 state-years reproduce the classification independently. The three states flagged in Methods as publishing a non-cohort denominator come out at 15.4, 12.5 and 14.3 times CCD kindergarten enrollment, against a median of 1.03 across the 24 states whose denominator is cohort-scale. Tennessee, the largest recent addition, sits with the latter at 1.56 (median per-school enrollment 65, and a panel total 1.45 times published statewide kindergarten enrollment); it contains 184 exact duplicate records carrying 0.4% of the national susceptible headcount, whose removal moves the convex share from 75.8% to 75.7% and leaves every reported figure unchanged at the stated precision. South Carolina’s 0.63 is expected rather than anomalous: its release reports a school-wide rate and the panel carries the kindergarten subset of those campuses. The extract is deposited (Supplementary Table 81).

### Which schools the rule selects, and whether a program can reach them

The allocation rule is a function of reported coverage and enrollment only. Supplementary Table 14 shows this directly: recomputing every state after deleting the school coordinates, so that no spatial kernel can be formed, leaves every allocation identical to exactly zero difference. Two columns are sufficient, which is what makes the rule deployable from data states already publish.

Deployability also depends on which schools it selects, because state authority differs by sector. We therefore compute the share of allocated doses that goes to private schools, against their share of enrollment, for every rule at the same 25% budget.

The scope is narrow and is not worked around. The national panel carries no sector field: none of the 33 state files has a sector, type or charter column. Sector is available alongside the enrollment needed to form a dose share for exactly two states, California 2022-23 and South Carolina 2024-25 (Supplementary Table 51). Texas flags sector but publishes no enrollment, so only the coverage-band composition is available there (Supplementary Table 70). Three files support these supplementary state-level analyses and no main-text result. The Texas 2023-24 named kindergarten release carries facility name and sector, which the deposited panel does not, and supports the sector analysis only. Two California kindergarten releases, 2014-15 and 2022-23, are matched on school name within county to give a pre- and post-SB277 school pair; the deposited California series begins in 2017-18, entirely post-SB277, so the full series from 2013-14 was obtained separately from the state open-data portal for the interrupted time series (Note 3). The 2022-23 California release is capped at 99%, a ceiling absent from 2014-15; the effect of that censoring is quantified in Note 3 rather than assumed negligible. Provenance, license and redistribution status for every file are in the replication package manifest.

Two states cannot give a national estimate, and the result is that they disagree in sign, which is the reportable finding. In California the dose-optimal rule assigns 16.4% of doses to schools holding 6.9% of enrollment, a 2.4-fold over-representation consistent with their 10.1% share of susceptibles, while lowest-coverage-first assigns 4.1%. In South Carolina the ordering reverses: 7.0% under the dose-optimal rule against 18.7% under lowest-coverage-first, with private schools at 6.4% of enrollment and 8.4% of susceptibles.

The equity floor behaves inconsistently for the same reason. A 75% floor takes the private share from 16.4% to 8.0% in California but from 7.0% to 15.9% in South Carolina. A floor defined on coverage is not a floor on sector, because the two are only loosely correlated and the direction of that correlation differs by state. The Texas coverage-band composition shows the mechanism: private schools are 72.4% of campuses below 75% coverage but 24.8% of the 85 to 94.4% band the dose-optimal rule targets, so a rule’s sector footprint is determined by where in the coverage distribution it spends rather than by any sector preference.

One scale caution for anyone rerunning this. The California release stores coverage as a percentage in [0, 100] and the South Carolina file stores it as a fraction in [0, 1], despite both columns being named for a percentage. Dividing the South Carolina column by 100 gives a mean coverage near 1%, which makes every school eligible and every rule look equivalent. The script detects the scale rather than assuming it.

### The South Carolina data and why the reporting format matters

Three of this paper’s four claims are about the low tail of the coverage distribution, so the reporting practice that removes that tail determines what can be tested. Five states were excluded from the national panel because more than 40% of their schools report exactly 100% coverage, which is not a plausible distribution and truncates precisely the schools the allocation rule funds.

South Carolina’s Department of Public Health has since consolidated high and low school-level rates into an aggregated reporting format. The file analyzed here was compiled before that change. Its distribution is intact: of the 1,512 schools in the 1,564-school file that report a 2025-26 rate, 52 (3.4%) report exactly 100% and 339 distinct coverage values appear (Supplementary Table 27). Under the current aggregated format, the low tail is not resolvable and the enrichment test Fig. 4 could not be run at all, since the exposed schools average 84.3% coverage in 2024-25 across the 29 schools flagged in the deposited file (84.7% across the 30 after the register reconciliation reported in the main text) and a substantial number fall within the aggregated band. We note this because it bears on whether the analysis is repeatable elsewhere: the constraint on this class of work is increasingly disclosure policy rather than method.

Two conditioning choices deserve statement. First, all 31 exposed schools in the register and all 30 of those retained for analysis are in the Upstate region, so a statewide test partly measures distance from the outbreak rather than school susceptibility; the statewide figure is 5.66-fold against 5.05-fold within region (Supplementary Table 28), and we report the smaller. Second, 20 Upstate schools lack a complete 2024-25 record and are dropped: nine lack the coverage rate itself and 11 lack the enrollment denominator (Supplementary Table 72 reports the two together). The nine missing coverage average 76.0% in 2025-26 against 90.7% among those retained, and the one exposed school among the dropped is one of these nine, so the exclusion removes low-coverage exposed schools and makes the test conservative rather than favorable. The 11 missing only enrollment average 95.6% in 2025-26 and include no exposed school, which is why the combined dropped group averages 82.0% (Supplementary Table 72).

### The search for a second exposure-coverage linkage

An independent estimate of *α* requires three things at once, and the third is what decides every case: school-level exposure flags, pre-outbreak coverage for those same schools, and a comparison set of unexposed schools drawn from the same reporting frame. Without the third there is no denominator and so no coefficient, only a description of coverage at schools that happened to be named.

The requirement behind the first is definitional rather than a matter of collection effort, and it is worth stating precisely because it explains why so few linkages exist. Health departments issuing outbreak communications publish two kinds of record. A visit list names the places an infectious person was during the infectious period, so a ranking built on it estimates the geography of an index case’s week. An investigation register names the schools a department judged exposed and acted on, through exclusion orders, quarantine, or targeted clinics; that determination of linkage is the quantity a risk weighting requires. The two are not interchangeable, and only the second identifies which schools an investigation reached.

We sought a second such linkage without success. Among states publishing school-level coverage alongside a 2025-26 outbreak, the exposure record fails the requirement in one of three ways: 1) Transmission did not run through schools, as in New Mexico, where the median case age was 20 years and no school or childcare outbreaks were reported^5,13^, 2)The published record lists locations a case visited rather than schools an investigation linked to a case, as in Utah, Arizona and Colorado^6^, or 3) exclusion was ordered by residence, as in the 2019 New York orders, which applied to every yeshiva in four ZIP codes irrespective of case linkage; the accompanying survey also reports coverage as percentages with no enrollment, so no susceptible headcount can be formed from it. Supplementary Table 74 records every state considered, the kind of record it publishes, and the reason it does or does not admit an estimate, so the exclusions can be checked rather than taken on trust.

#### Retrieval

South Carolina coverage figures are the Department of Public Health 45-day school reports published at https://dph.sc.gov, obtained before school-level rates were consolidated into the current aggregated format; the exposure register was reconstructed from the department’s public outbreak communications, enumerating the release series from the first school announcement on 8 October 2025 to the declaration that the outbreak had ended, recording the date of first announcement as the exposure event and treating later appearances as continuations. Reconciling that register against the deposited panel added one school, the main Dorman High campus, exposed in a separate event from its Freshman Campus; the Freshman Campus has no 2024-25 record and so carries no pre-outbreak predictor. The register also contains a homeschool cooperative that does not report to the 45-day report, so it has no enrollment or coverage and cannot enter either the flags or the denominator (Supplementary Table 73). These reports record the share of enrolled students holding a valid Certificate of Immunization, that is, compliance with the full required school schedule rather than MMR receipt alone, so the coverage used here is a lower bound on MMR-specific coverage (Methods, and item 12 of Supplementary Note 10). Washington’s Clark County school immunization files, covering 2018-19 and 2024-26, come from the Department of Health school immunization data pages, with exposed schools taken from county health department outbreak notices. Utah exposed schools come from Department of Health and Human Services outbreak communications, matched by name to the school immunization report. Colorado school immunization rates come from the Department of Public Health and Environment school dashboard, with exposed schools from departmental outbreak updates. The Arizona Mohave County figures were taken from Department of Health Services Immunization Data Report statistics as reported in state and national coverage and cross-checked across three independent secondary sources citing that department directly; one of the two schools has a coverage and enrollment figure confirmed by all three, the other by two, and its exemption rate is left missing rather than estimated. Rockland County figures come from the New York State school immunization survey. The geocoding of school addresses is ours in every case; no coverage or exposure figure is.

#### Why Arizona is deposited but not fitted

Arizona’s school-level immunization report is served through an interactive query tool requiring per-school form submission, with no bulk file, so the Mohave County roster that would form the denominator could not be verified against a primary source; the department’s county summary confirms 25 K-12 schools reporting and none at or above 95%, but a county-level aggregate cannot substitute for the school-level comparison set. The two verified schools are used for a different purpose. Both sit in Colorado City, the Arizona half of Short Creek; Hildale, Utah is the other half, and its school appears in the Utah file. One town, three exposed schools at 7.7%, 34.0% and 40.0% coverage, ranked in two separate state denominators (Supplementary Table 65). That is the within-state ranking limitation in its clearest observed form, and it is the reason these two schools are in the package.

## Supplementary Note 7 | Coverage mandates and susceptible concentration

### Coverage and concentration trends

Mean coverage and the concentration of susceptibles, measured throughout as the share of a state’s susceptibles held by the 5% of schools with the lowest coverage, move independently. In 15 of 16 retained states with four or more reported years, mean coverage fell over the observation window. Concentration did not track it: some states with falling coverage saw concentration fall as well, others saw it rise (Supplementary Fig. 23).

### Interrupted time series and placebo test

California’s deposited series begins in 2017-18, entirely after SB277 took effect, so a fitted pre-trend is not identifiable from it. We obtained the full 2013-14 onward series from the state open-data portal (Supplementary Table 60), giving three pre-policy years, the minimum for identification, and a real constraint on what the design can support.

Coverage was already rising at 1.09 percentage points per year before the policy. A first-versus-last comparison would attribute that pre-existing trend to the mandate. The fitted level change of 2.26 points is the policy effect after accounting for it (Supplementary Table 61).

Two cautions. First, the pre-trend should not be extrapolated far: projected linearly to 2022-23 it exceeds 100% coverage, so Supplementary Fig. 24a clips the counterfactual at the feasible bound and it should be read as a direction rather than an estimate. Against that counterfactual the net effect is positive for under two years and negative thereafter, crossing zero at 1.7 years (Supplementary Table 62), which is a further reason to read the design as descriptive. Second, there is no comparator state, so the design assumes nothing else changed in California at the 2016-17 boundary.

The placebo test addresses the second concern directly. Massachusetts, Minnesota and Hawaii are the only other states with three or more pre-2016-17 years, and none changed their school mandate. Fitting the identical segmented model at the identical breakpoint, two of the three return a significant level shift somewhere no policy occurred: Minnesota on mean coverage (+1.51 points, p = 0.041) and Hawaii on concentration (−12.8 points, p = 0.0001). Only Massachusetts is null on both outcomes. Of the six state-outcome fits, two are significant at the 0.05 level, against the 0.3 expected by chance (Supplementary Fig. 21, Supplementary Table 63). We report this rather than omitting it. The interpretation differs by outcome, and the distinction matters. For mean coverage, the Minnesota result is a direct caution: the design produces a significant level shift on that outcome where no policy exists, so the California coverage estimate cannot be read causally, which the permutation test below and a refit excluding 2020-21 confirm independently (Supplementary Table 64) (p = 0.43). For concentration, the Hawaii result cuts the other way: it shows the design can detect a large shift in that outcome, and it found none in California (permutation p = 1.00), so the California null on concentration is a finding rather than a failure to detect.

## Supplementary Note 8 | Economic evaluation

### Cost model and sensitivity

The expected-value figures in the main text use the two estimated introduction probabilities, *p* = 0.0018 and *p* = 0.019. A deliberately pessimistic anchor of *p* = 0.05, above anything the outbreak record supports, gives 706 forgone infections and US$73.8 million (at the societal cost of US$104,629 per case); it is reported here as a ceiling rather than as an estimate, and it is the upper bound in Supplementary Table 21. The 77,292 doses cost US$4.6 million whether an introduction occurs, which is why net benefit can be negative at the lowest probability under the narrowest cost perspectives while the allocation comparison, a ratio in which the probability cancels, is unaffected.

The structure of the comparison is given in Methods. Both rules purchase and deliver the same doses, so dose cost cancels from the ratio and enters only the net-benefit and cost-per-averted calculations. The conditional quantity, the value forgone if every school in the panel faced an introduction in one season, is 14,113 excess infections at a fixed budget; Supplementary Fig. 9 shows what that is worth under each per-case perspective, and it is a counterfactual ceiling rather than an expectation.

Four per-case perspectives are used, spanning the range in the published literature: the incremental public-health response cost (US$16,197), the mean observed response cost (US$43,203), the societal cost including hospitalization, productivity loss and mortality (US$104,629), and the largest observed per-case cost (US$243,614). The sensitivity factorial adds a fifth level, the lowest published per-case cost (US$6,973), so it crosses five cost levels rather than the four reported perspectives. The societal figure is taken from Wells et al.^9^ so that our estimate is directly comparable with theirs.

Across the full factorial of per-case cost (5 levels), dose cost (3 levels) and budget fraction (3 levels), 45 scenarios, *conditional* net benefit is positive in every one, and the cost per infection averted ranges from US$70 to US$412. This is not in tension with the two negative expected net benefits in Supplementary Table 21: the conditional figure assumes an introduction, the expected figure multiplies by *p*, and dose cost is subtracted in both.

## Supplementary Note 9 | Robustness of the central result

### Robustness of the central result

Four ways the headline result could be an artifact, each tested.

<u>R1, driven by a few large states?</u> Leave-one-state-out gives 72.9% to 81.6% against a full-sample 75.8%. The most influential state is Illinois and removing it *raises* the estimate.

<u>R2, driven by the exclusion rule?</u> Varying the saturation threshold from 20% to no exclusion at all gives 75.6% to 75.9%.

<u>R3, circular, because the allocation is scored with the objective the optimizer maximizes?</u> Scored instead on the number of schools left above the epidemic threshold *R_v_* = 1, which the Duijzer rule does not maximize, the dose-optimal allocation still wins in 26 of 28 states. The margin is narrower on this metric, 22.1% versus 12.4% of supercritical schools cleared, than the herd-benefit metric suggests.

<u>R4, is the convex classification just a coverage threshold?</u> The classification is a threshold by construction. The allocation rule is not: it ranks by the average herd benefit per dose of the move to. Against a rule that targets convex-region schools directly at the same budget, the marginal-efficiency rule averts 30,939 infections versus 15,875 at a pooled national budget, a further 1.95-fold gain (1.73-fold at the per-state budgets used in the main text, 29,479 versus 17,084), and directs 3.7% of the budget to schools already above, which no threshold rule would do. This is the check that establishes the contribution.*f̄f̄*

## Supplementary Note 10 | Limitations

### Limitations

Most concern what the surveillance record can currently support rather than the estimator or the allocation comparison itself, and several are bounded by analyses reported above.

1. <u>Measurement gap at the epicenters</u>. The 2025-26 outbreak epicenters are systematically under-represented in the vaccination databases. Gaines County, Texas records 408 secondary cases against three schools and 63 susceptible kindergarteners. Several amplifying counties recorded more secondary cases than their entire recorded susceptible pool. A risk model calibrated on recorded schools cannot represent these communities; the allocation comparison itself does not use the risk model (Supplementary Note 2) and is not affected by this gap.
2. <u>Tail-only discrimination</u>. Overall separation between amplifying and fizzling counties is near chance, while the top-decile enrichment is significant: the metric identifies the extreme tail, which is where the susceptible pool concentrates, rather than ranking the middle of the distribution.
3. <u>The school-scale test is underpowered in this outbreak set</u>. School-scale enrichment was 1.67-fold here against 2.0-fold reported in the previous study^6^ across seven earlier validation outbreaks, on a design with 28% power against an effect of that size. The most plausible explanation is limitation 1: the 2025-26 epicenters are precisely the counties where school-level records are sparsest, whereas the published validation outbreaks occurred in better-recorded settings. This is a statement about the 2025-26 data, not about the earlier result.
4. <u>Utah has a denominator but no register</u>. Utah contributes 486 cases to the outbreak analysis. A kindergarten MMR file obtained from the state epidemiology program in August 2026 supplies the comparison set the earlier assessment lacked: 867 schools over twelve school years, 2014-15 to 2025-26 (8806 school-years, of which 10% are suppressed because the kindergarten enrolled fewer than eleven students), including 662 schools with usable 2024-25 coverage measured before the 2025-26 outbreak. What still blocks an independent estimate of the risk scaling is the exposure side, not the denominator: Utah’s public record is a visit list, naming locations a case attended, rather than an investigation register naming the schools a department judged exposed and acted on. A revised release of the same file (11 August 2026) adds the reporting denominator, the number of kindergarteners each school reported on, so a susceptible headcount can now be formed: 45,600 enrolled across the 662 schools with usable 2024-25 coverage, holding 5,099 susceptible children, 93.0% of them in schools below the inflection. Enrollment-weighted mean coverage is within a percentage point of the unweighted mean in every year, and the difference is not distinguishable from zero: −0.62 pp in 2024/25 (bootstrap 95% CI −1.80 to 0.33), with school size and coverage effectively uncorrelated (Spearman *ρ* = −0.03, p = 0.41). School size therefore carries no coverage signal here, and the unweighted series reported above is not misleading. What the file still lacks is the exposure indicator, and its schools carry anonymized identifiers with no name, LEA or coordinates, so it cannot be joined to the national panel either. Utah therefore remains outside the risk-scaling estimate while moving from unlinkable to one field short, and that remaining field is one the department already holds.
5. <u>The SB277 series is descriptive, not causal</u>. Three pre-policy years is the minimum for identification, two of three placebo states return a significant level shift where no policy occurred, and the exact permutation p on the coverage level change is 0.43 against a design floor of 1/7.
6. <u>Separability</u>. The property that makes the allocation problem tractable also means the risk model does not itself represent between-school herd protection. A simulation test controlling for own coverage and local school density returned a null, so we make no claim of a neighbor effect.
7. <u>Single-population herd effect H</u>. *H*(*f*) relies on an SIR final-size calculation for an isolated population. A fully coupled metapopulation treatment would shift the inflection point, though the convex share never falls below half across the parameter grid examined. Separately, because the inflection *f̄* depends strongly on the assumed introduction while the critical fraction *f*^∗^ barely does (7.0 against 0.8 percentage points across the school-size grid), the convex share is a statement about expected return under a stated exposure assumption and not a count of schools lacking herd immunity; at kindergarten enrollments the deterministic final-size relation also overstates *f̄* relative to an exact stochastic treatment with a single index case (Supplementary Note 1, Supplementary Fig. 27, Supplementary Fig. 28).
8. <u>Cost model linearity</u>. Averted infections are assumed to scale with the herd effect. A fixed-plus-incremental structure per averted *outbreak* would be closer to the structure of the underlying costings, which is why the dollar figures are reported as indicating scale rather than estimating it.
9. <u>Settings that report nothing cannot be reached or ranked</u>. The rule allocates against reported school coverage, so a setting that files no enrollment and no coverage is invisible to it. The South Carolina exposure register contains a homeschool cooperative with 17 quarantined, more than at most schools in the analytic panel, which files neither and so can be neither flagged nor dosed. This is the limiting case of the measurement gap in item 1: in Texas and New Mexico the schools exist and the records are thin, whereas here the health department identified the setting and no school-based instrument can represent it.
10. <u>Deliverability is not uniform across the coverage gap, and the rule cannot see it</u>. The allocation objective treats a dose sent as a dose administered, so a school’s non-immune share is taken as addressable. Only one file we hold decomposes it. The Utah kindergarten series separates the gap into exemptions and children conditionally enrolled or out of compliance, and exemptions are 83% of it in 2024-25, rising from 67% in 2014-15 to 91% in 2025-26. The two components are not equally reachable: a documentation gap closes with a dose or a record, whereas a filed exemption reflects a refusal that catch-up delivery does not by itself reverse. Where the gap is mostly exemptions the achievable coverage gain at a school is smaller than its reported deficit implies, which would attenuate the benefit of any rule that spends against that deficit, ours and coverage-rank alike. Whether it attenuates them differentially depends on how the exemption share varies with coverage, which one state’s aggregates cannot establish; we flag it as a bound on interpretation rather than adjust for it and note that no state in the analytic panel reports the split at school level.
11. <u>The ranking is within states, but transmission is not</u>. Coverage reporting and dose budgets are state functions, which is why the ranking is computed within state, but a susceptible community that straddles a state line is split across two denominators. Short Creek spans Hildale, Utah and Colorado City, Arizona as a single town; its three exposed schools, at 7.7%, 34.0% and 40.0% coverage, are ranked separately, and the percentile understates concentration on both sides (Supplementary Table 65).
12. <u>The clustering signal is conditional</u>. The within-county spread of school coverage predicts which counties an introduction reached in Texas, but not in California or New Mexico, where county size already predicts reach. It is also less persistent year to year than mean coverage, Spearman *ρ* 0.54 against 0.31, so a rule built on it needs recent data and a setting where reach is not size-driven.
13. <u>The validation coverage is certificate completeness, not MMR receipt</u>. The South Carolina 45-day school report records the share of enrolled students holding a valid Certificate of Immunization, certifying compliance with the full required school schedule rather than MMR receipt alone. Because two MMR doses are required for kindergarten through grade 12, the rate is a lower bound on MMR-specific coverage and the susceptible headcounts derived from it are upper bounds. The consequence for the validation is limited by its design: the top-decile test ranks schools rather than using coverage levels, so a bias that is common across schools cancels, and only school-to-school variation in the gap between certificate completeness and MMR-specific coverage can move it. The national panel is not affected, as those releases are MMR-specific.

Taken together, these constraints bound the magnitude of the risk scaling and the reach of the school records rather than the ordering result. The unweighted rule is the worst of the seven candidates under all three structurally different uncertainty sets, retaining 12.1%, 12.1% and 34.1% of attainable benefit under the coefficient interval, the weight functional form and the covariate set respectively (Supplementary Table 78; these are the structural sets, distinct from the 14.2% worst case over the fitted α range), at 10% and 50% budgets and under Firth penalization, and the two-percent hedge is best at every budget and under either estimator, though which hedge is best does depend on how the uncertainty set is drawn, and the convex-share result ranges 72.9% to 81.6% under leave-one-state-out. What the limitations above establish is that the strength of the introduction-risk scaling is a bounded unknown, which is why the rule comparison is posed as a decision under uncertainty rather than as an estimate.

## Supplementary Figures

**Supplementary Fig. 1.**
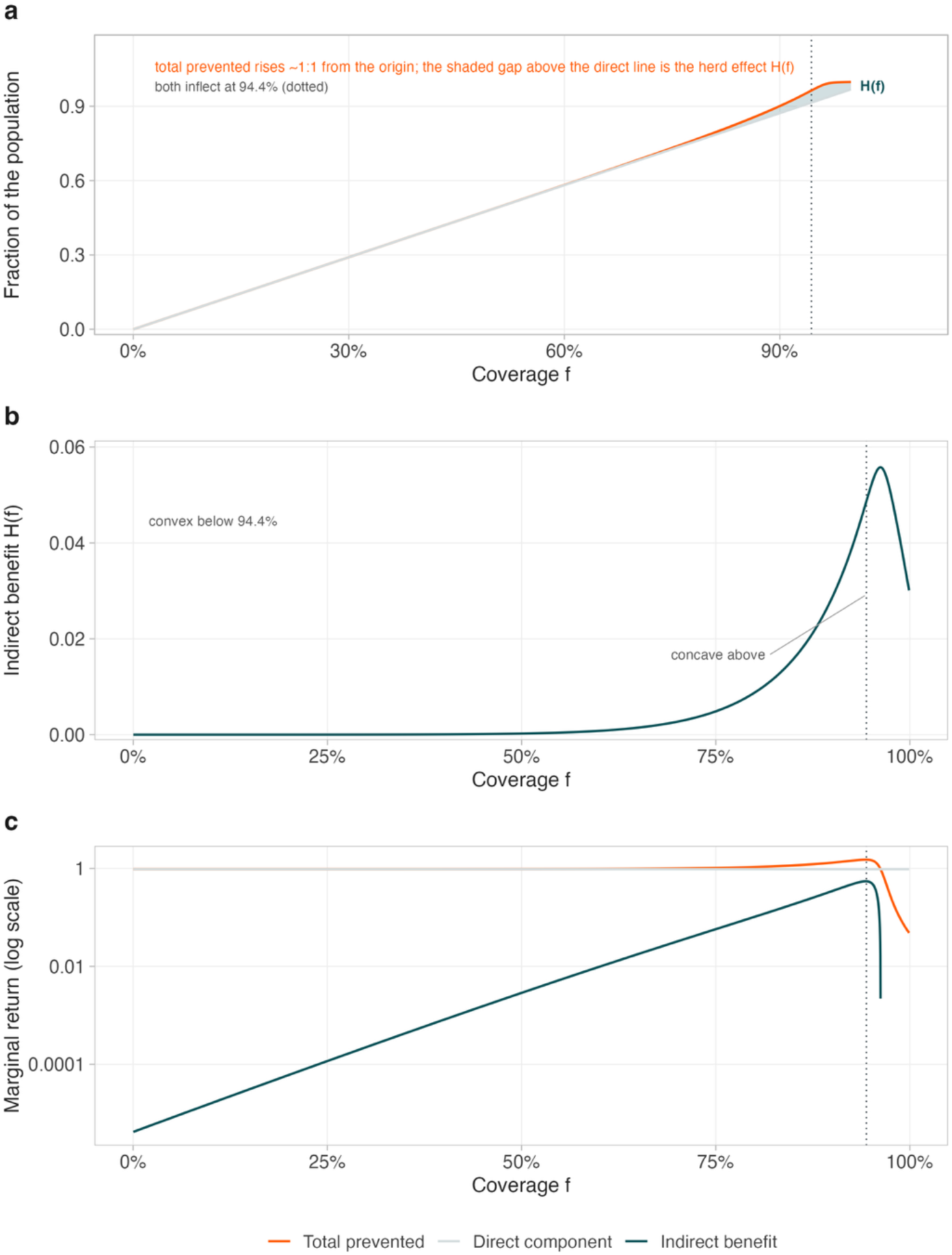
What is sigmoid and what is single-peaked. **a,** Total infections prevented and the directly immunized fraction against coverage, on the shared population-fraction axis. Total averted rises close to one-for-one from the origin and lies just above the direct component throughout; the shaded gap between the two curves is the indirect benefit *H*(*f*), redrawn on its own axis in b. **b**, The indirect benefit *H*(*f*), the paper’s allocation objective; it peaks at 0.0558 of the population, so on a shared axis with total prevented it would be a flat line along the bottom. Convex below the inflection *f̄* = 94.4%, concave above. Total averted shares that inflection, since it is *H*(*f*) plus a linear term. **c**, Marginal return per unit coverage, log axis. The direct component is constant at the vaccine efficacy; total averted runs just above it, from 0.9691 at *f* = 0.25 to 1.4778 at *f* = 0.95, while the indirect benefit rises from below 10^−3^ to 0.5087 over the same range. It is *H*, not total prevented, that is nearly flat at low coverage. Dotted line marks *f̄* in all panels. Values in Supplementary Table 1.

**Supplementary Fig. 2.**
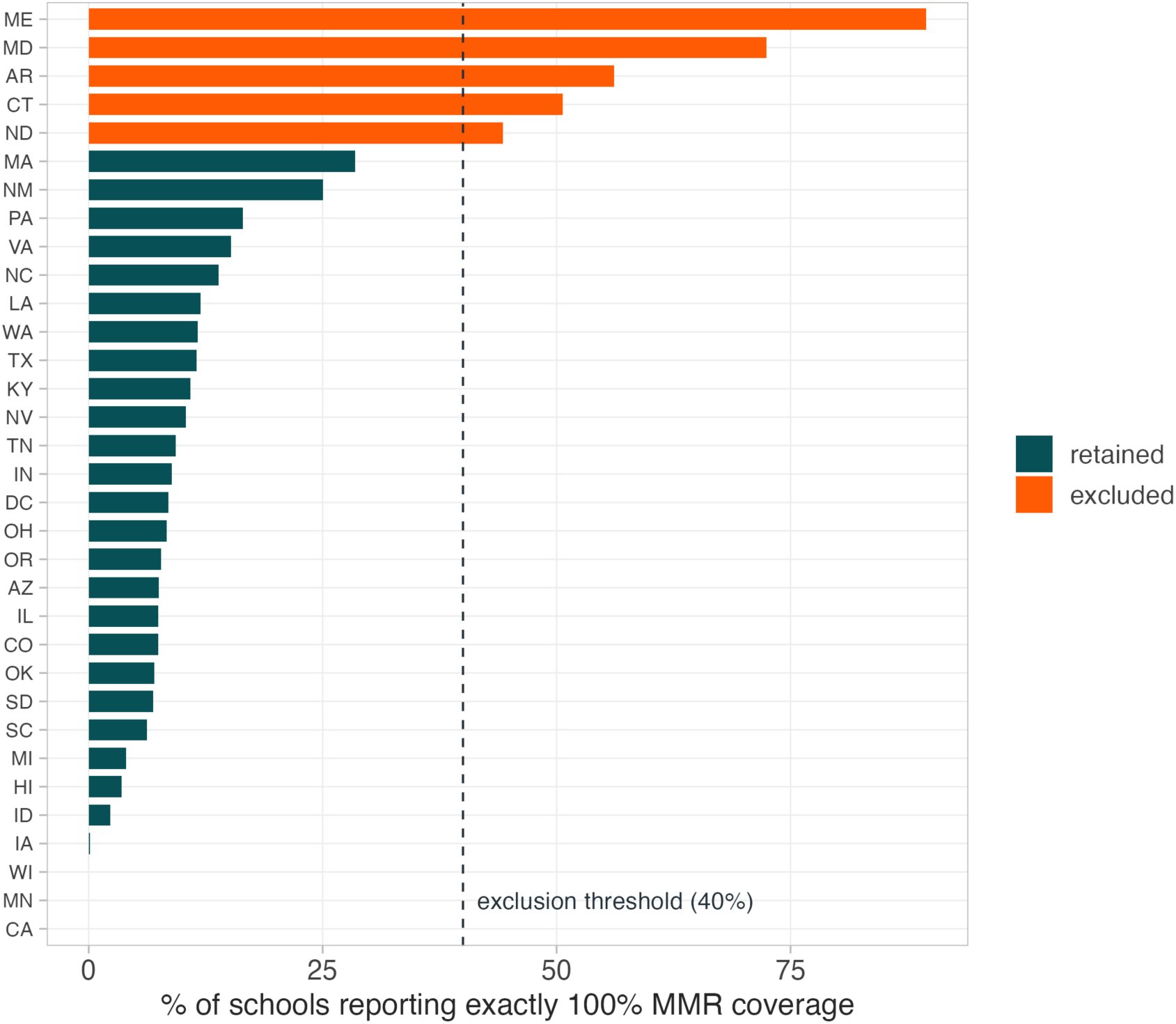
Reporting-quality audit. Percentage of schools reporting exactly 100% MMR coverage, by state. The dashed line marks the 40% exclusion threshold. Excluded states are shown in a contrasting color.

**Supplementary Fig. 3.**
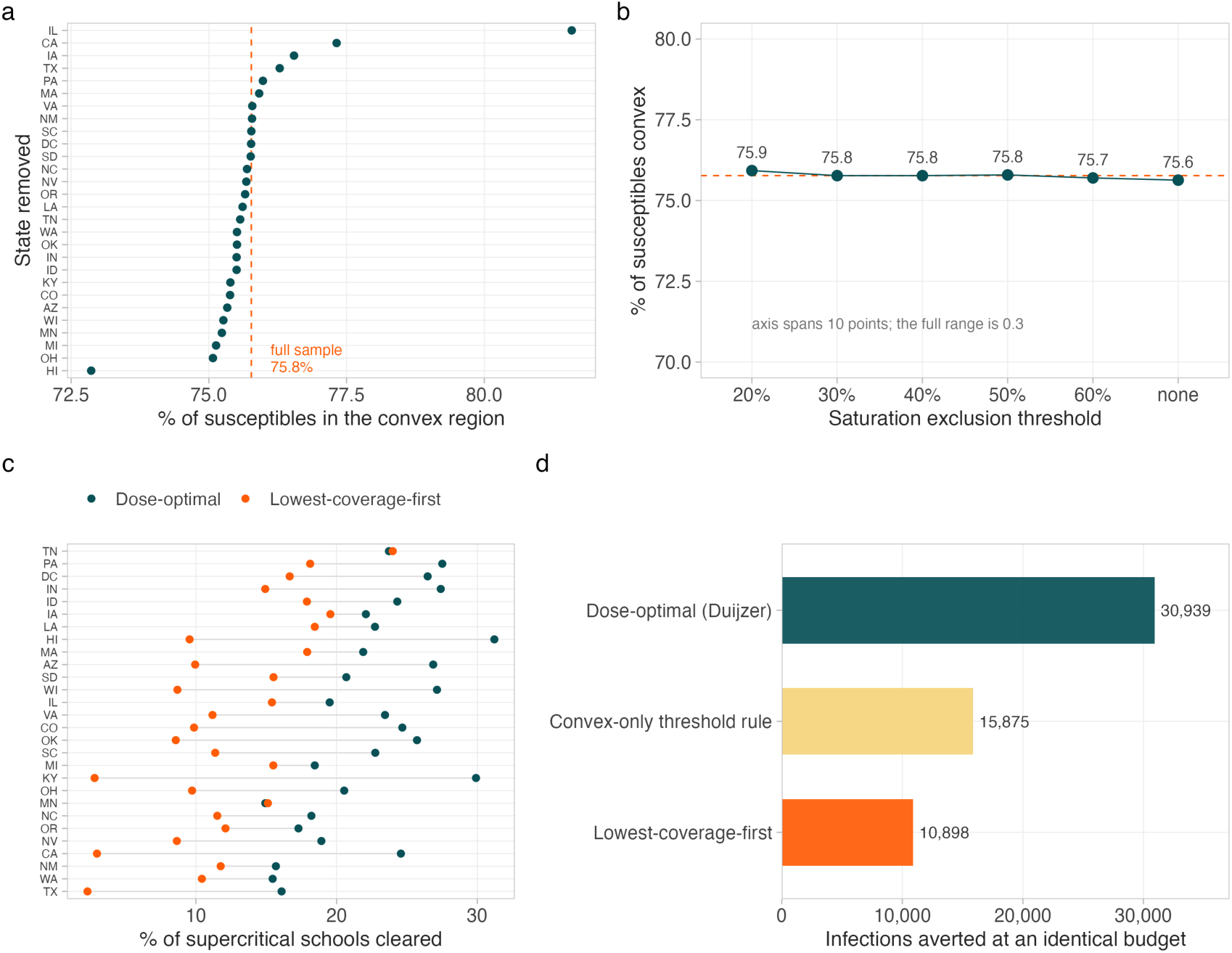
Robustness. **a,** Convex share of national susceptibles (%) recomputed with each state omitted in turn; n = 28 jurisdictions, range 72.9% (dropping HI) to 81.6% (dropping IL) against 75.8% for the full panel. **b,** Convex share against the reporting-quality exclusion threshold (maximum permitted percentage of schools at exactly 100% coverage); note the vertical axis spans 10 points while the full range is 0.3, from 75.6% (no exclusion, 33 states) to 75.9% (20% threshold, 26 states). **c,** Independent scorer: schools left above *R_v_* = 1 in each state under the dose-optimal and lowest-coverage-first rules at a 25% budget, a quantity neither rule maximizes; the dose-optimal rule clears more in 26 of 28 jurisdictions (22.1% against 12.4% of the base). **d,** Allocation rule comparison at a pooled national budget of 77,292 doses: indirect infections averted by the dose-optimal rule (30,939), a convex-only threshold rule targeting the same schools largest-pool-first (15,875, 1.95-fold below) and lowest-coverage-first (10,898, 2.85-fold below).

**Supplementary Fig. 4.**
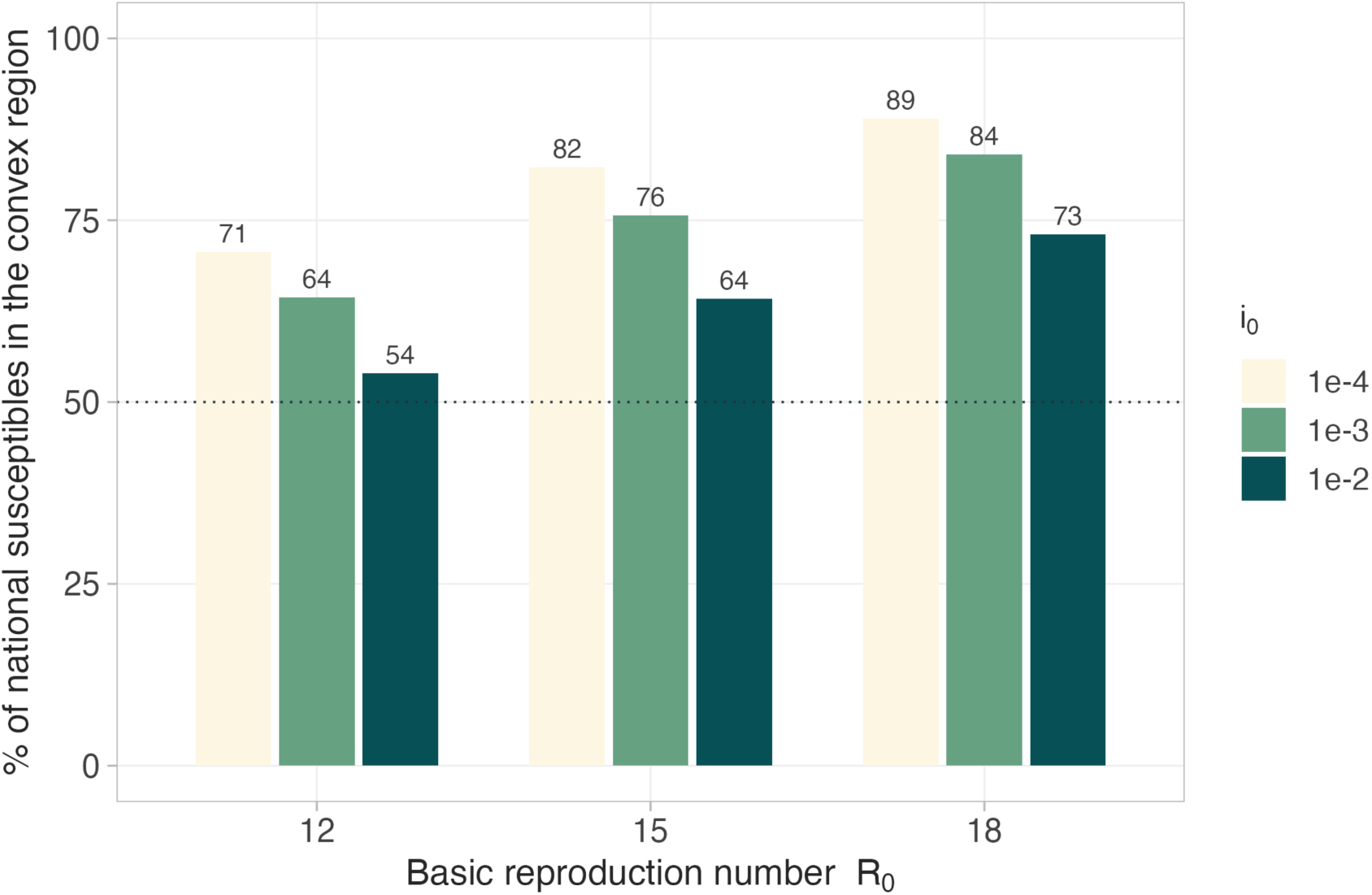
Convex share across the parameter grid. Percentage of national susceptibles in the convex region for each combination of *R*_0_ and *i*_0_. The dotted line marks 50%. The share never falls below 54% in any of the nine scenarios, so the majority claim does not depend on the central parameter choice.

**Supplementary Fig. 5.**
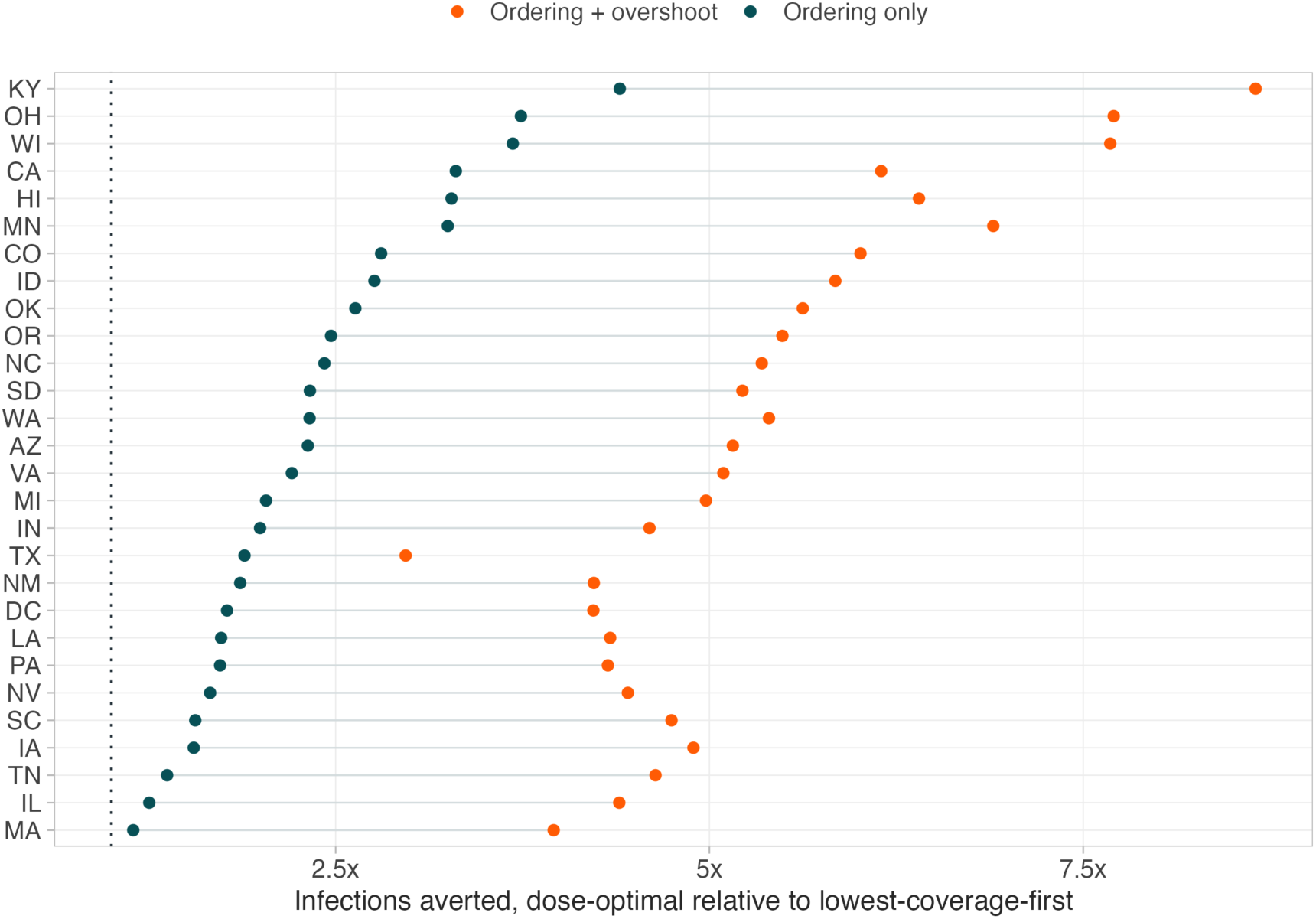
The ordering error and the stopping-rule error are separable. Per-state efficiency gain of the dose-optimal rule over lowest-coverage-first under two definitions of the comparator. “Ordering only” gives both rules the same per-school target *f̄*, so they differ solely in which schools are funded. “Ordering + overshoot” additionally lets the comparator take each funded school to full coverage, past the maximizer *f*^∗^ of *H*. States are ordered by the ordering-only gain.

**Supplementary Fig. 6.**
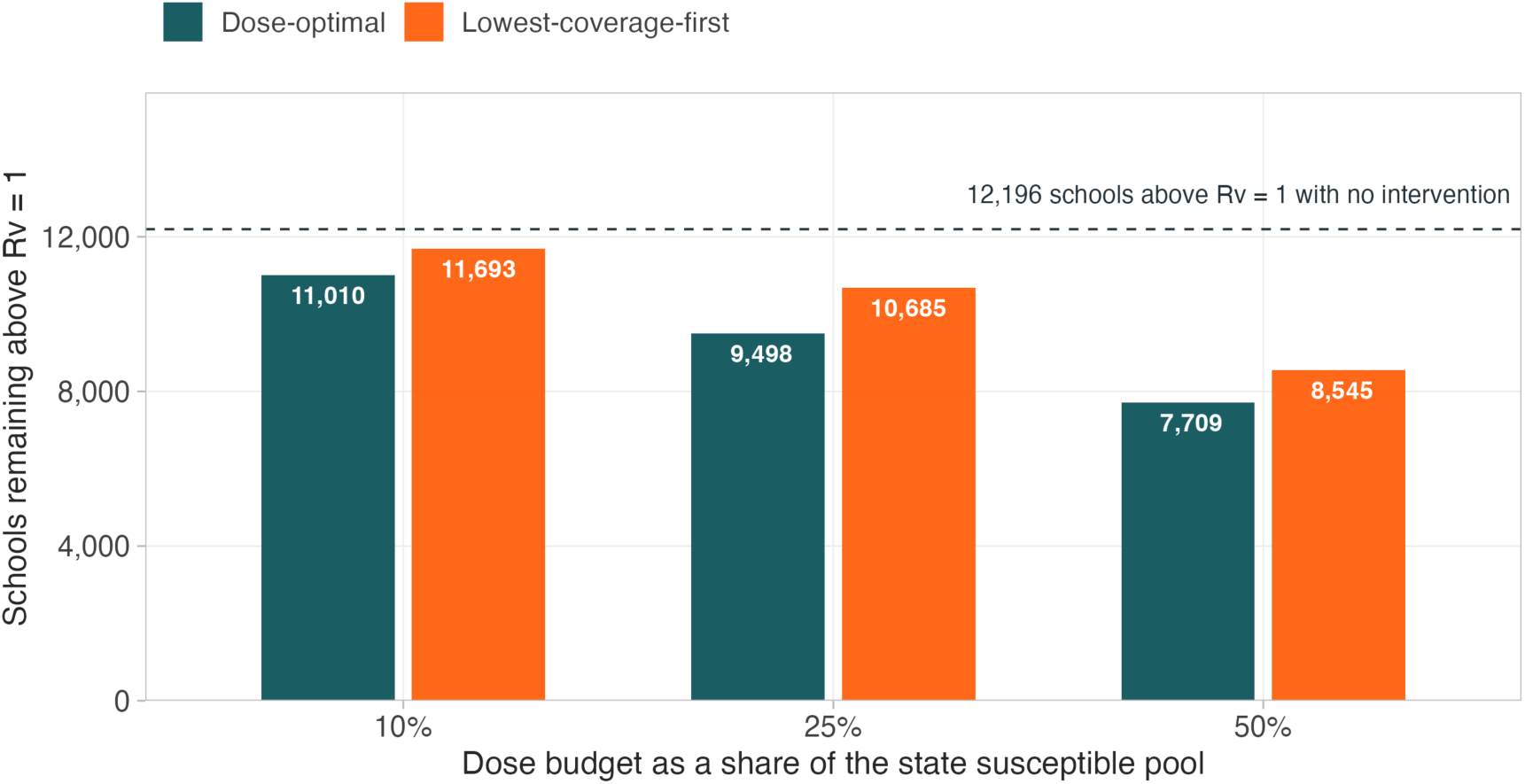
Transmission-capable schools remaining. The two-percent hedge is not drawn here; it clears 2,497 of the 12,196 schools against 1,511 under lowest-coverage-first and 2,698 under the unweighted dose-optimal rule (Fig. 6a of the main text, and figure7_sourcedata_a.csv and TableS83_practice_clearance.csv in the replication package). Schools above *R_v_* = 1 after allocation under each rule, at three budget levels. The dashed line is the 12,196 schools above threshold with no intervention.

**Supplementary Fig. 7.**
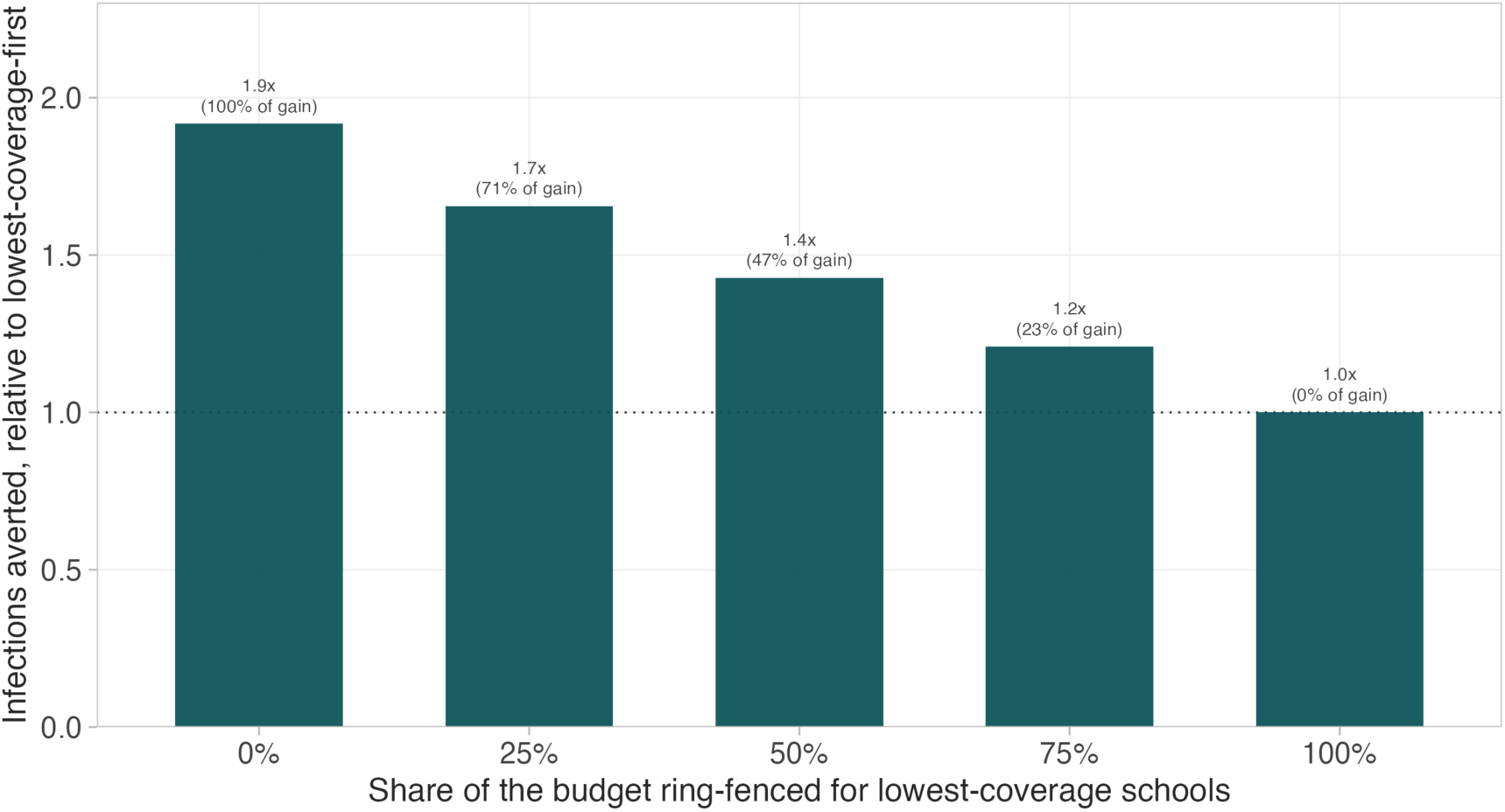
The efficiency gain under an equity floor. Infections averted relative to lowest-coverage-first, as a function of the share of the budget ring-fenced for the lowest-coverage schools. Ring-fenced doses are allocated lowest-coverage-first; the remainder by marginal efficiency.

**Supplementary Fig. 8.**
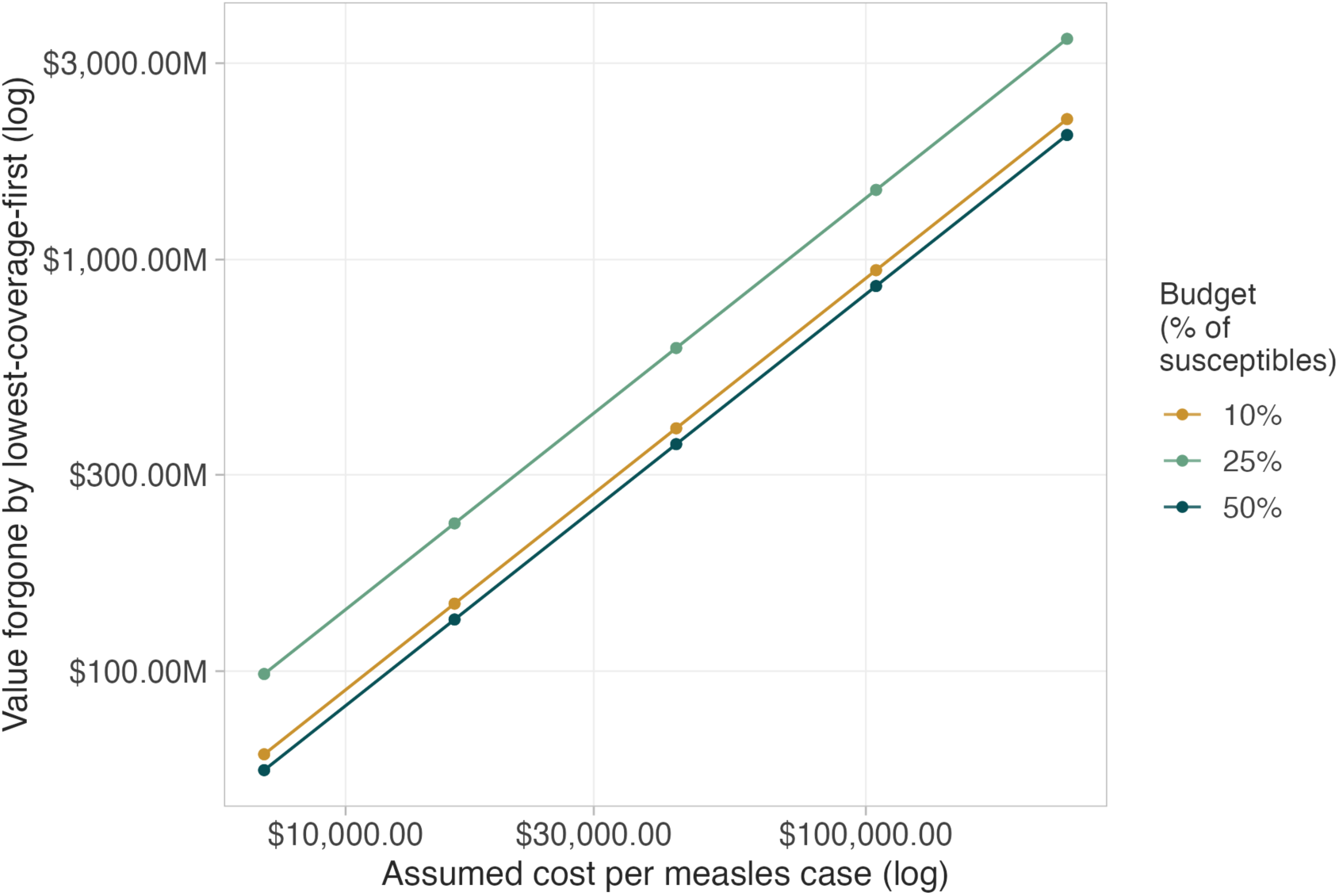
Cost sensitivity. Net benefit and cost per infection averted across all 45 scenarios.

**Supplementary Fig. 9.**
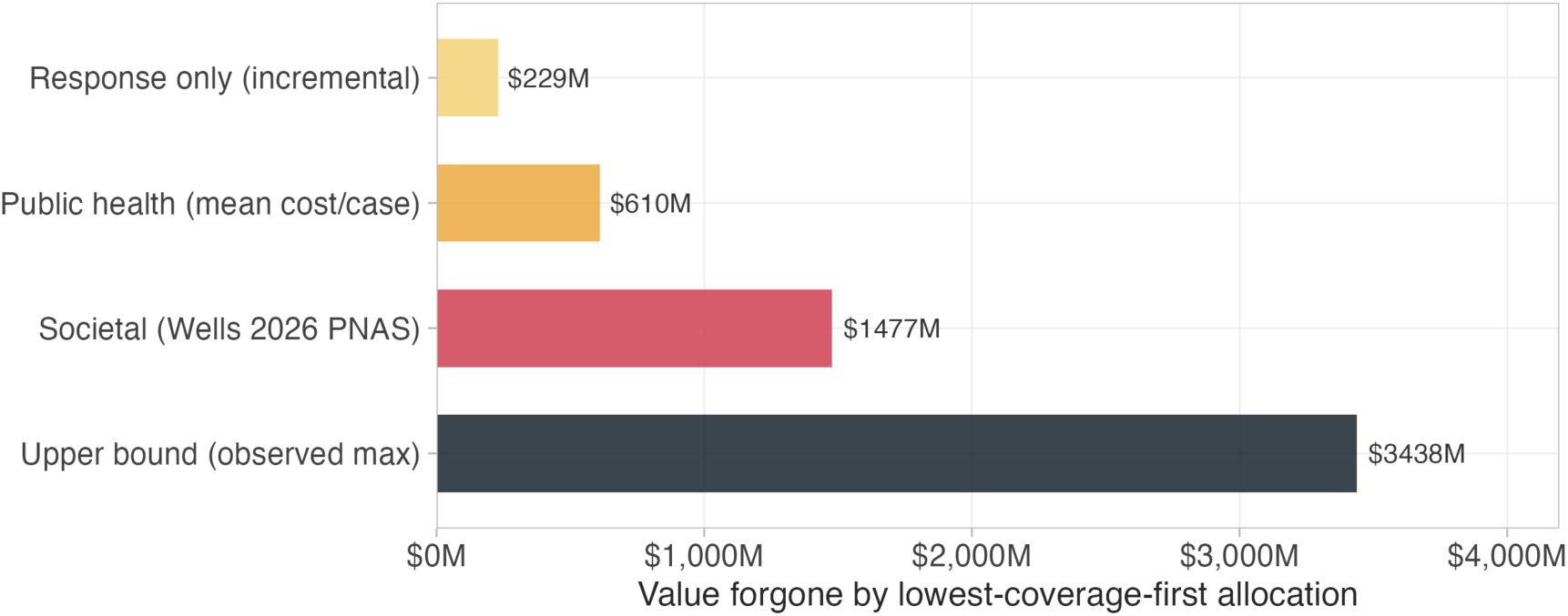
Conditional value forgone, by cost perspective. The counterfactual in which every school in 28 states experiences an introduction in the same season, corresponding to 14,113 excess infections at an identical dose budget. This is the quantity the herd effect *H* computes directly and the one whose ratio gives the 1.92-fold gain; it is not an expected cost, and the expected-value figures in Fig. 2c should be used for any cost comparison. Dose cost is US$4.6 million under all four perspectives.

**Supplementary Fig. 10.**
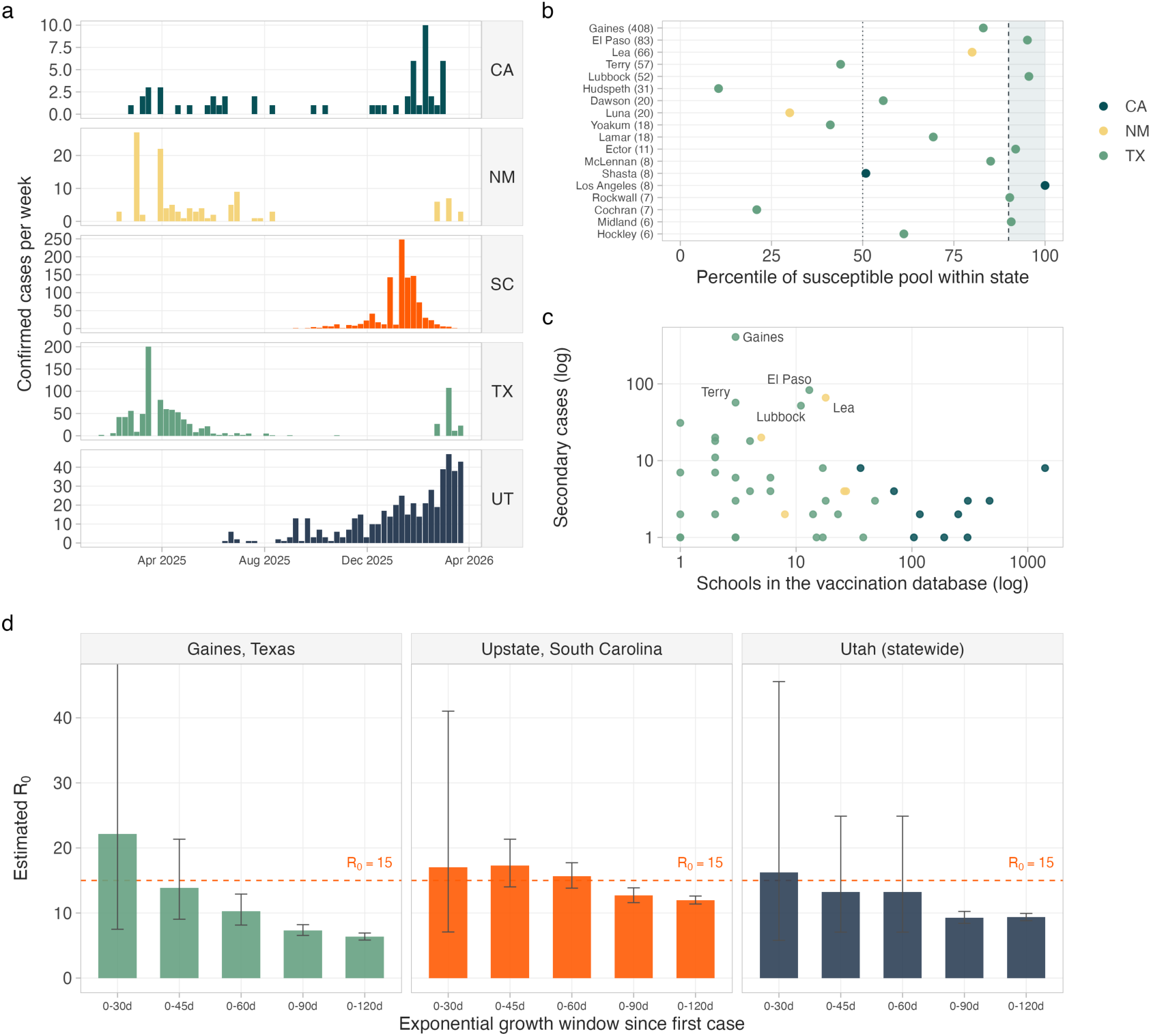
County-scale validation against the 2025-26 outbreaks. The South Carolina test (Fig. 4) reaches the school, which is the unit the allocation rule acts on, whereas this analysis is limited to 18 amplifying counties. **a**, Weekly confirmed measles cases in five outbreak states, county-day surveillance, 2,628 cases total. **b**, The 18 amplifying counties (≥5 secondary cases) ranked by percentile of susceptible pool within their own state. Shading marks the top decile; 6 of 18 fall there (3.3-fold enrichment, p = 0.0064, one-sided binomial). Parenthetical values are secondary case counts. **c**, Secondary cases against the number of schools recorded in the vaccination database, both on log scales. The epicenters are where the data are thinnest: Gaines County, Texas recorded 408 secondary cases with three schools on record. **d**, Exponential growth rate estimates converted to *R*_0_ across five-time windows for three independent outbreaks, including Utah, which contributes no information to the risk model. Error bars are 95% confidence intervals; *R*_0_ = 15 falls within the interval in 8 of 15 windows. Generation time 11.7 days.

**Supplementary Fig. 11.**
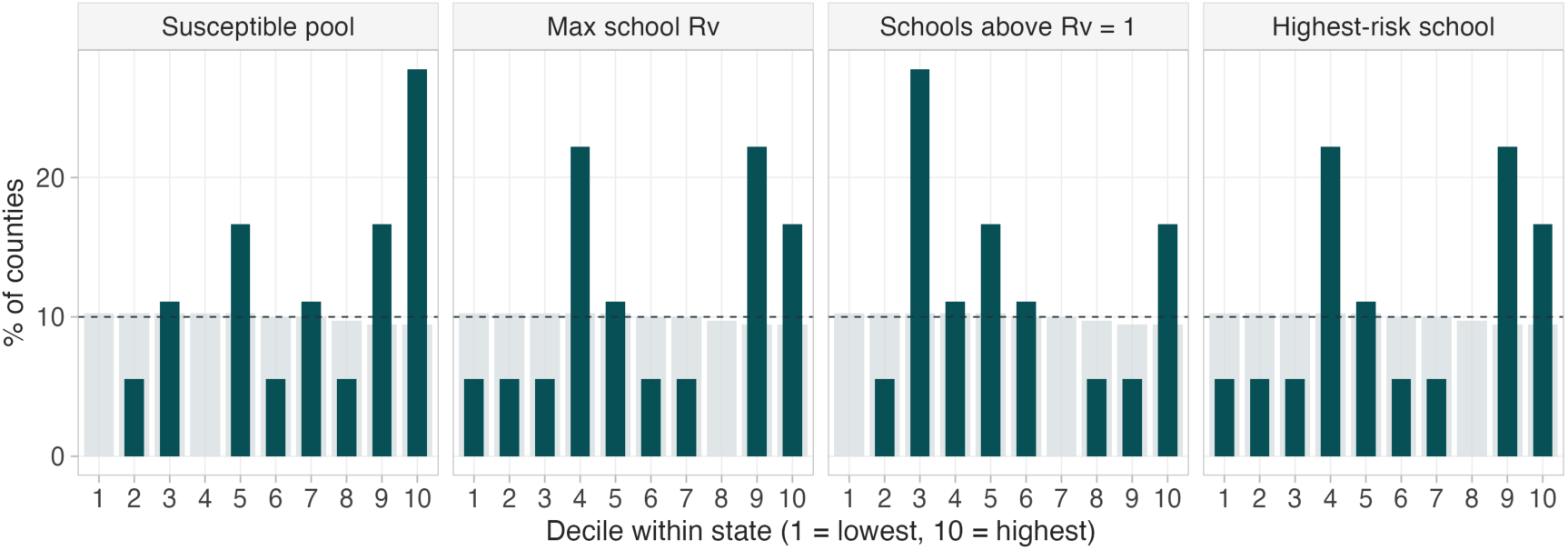
Decile enrichment across all four risk metrics. Distribution of amplifying counties across within-state deciles for the susceptible pool, maximum school *R_v_*, number of schools above *R_v_* = 1, and top school percentile.

**Supplementary Fig. 12.**
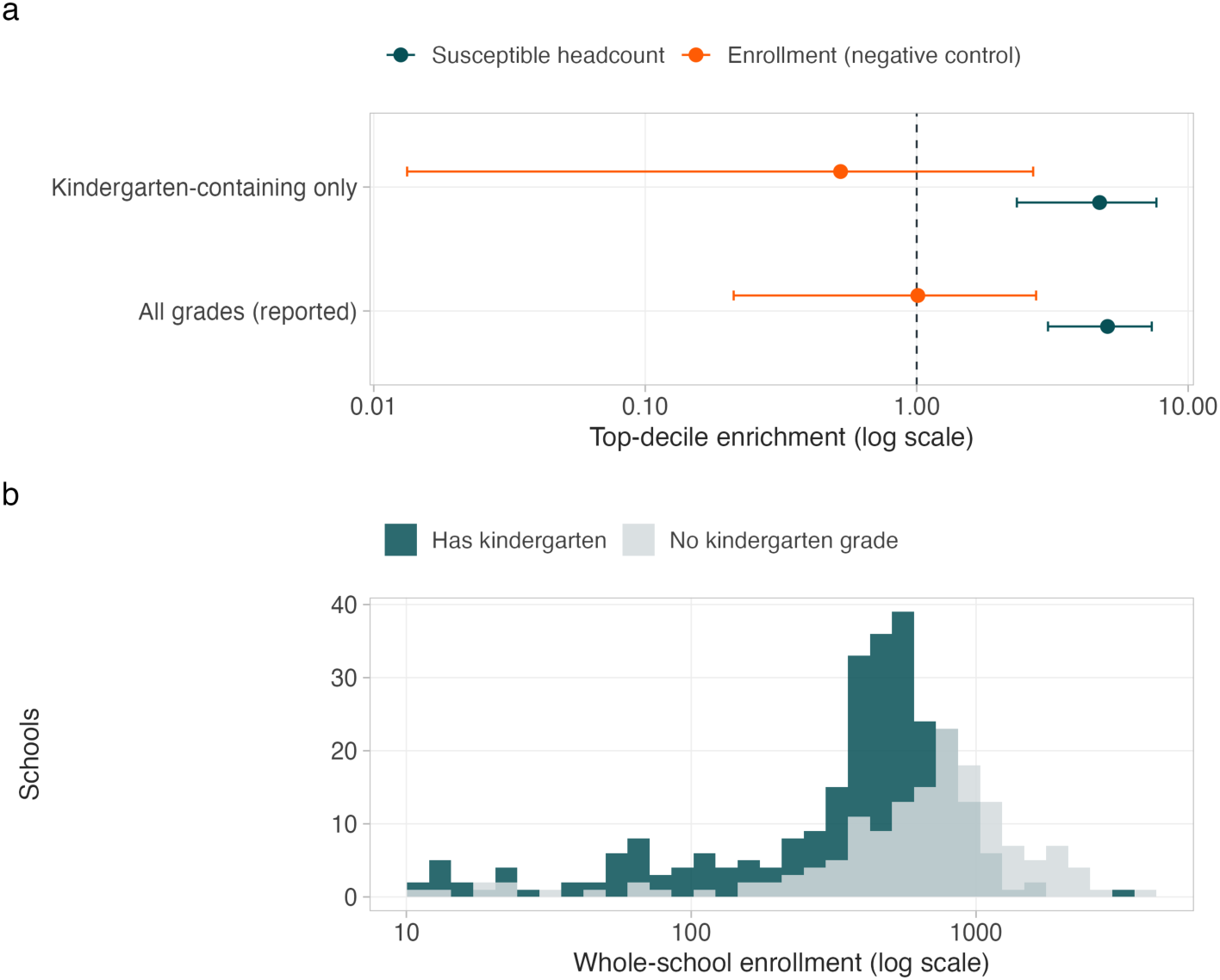
The South Carolina enrichment does not depend on grade composition. **a**, Top-decile enrichment of exposed schools, on all 424 Upstate schools as reported and on the 269 whose grade range contains a kindergarten grade, for the susceptible-headcount ranking and for enrollment alone as a negative control. Bars are exact 95% intervals; the dashed line is no enrichment. Restriction leaves the estimate at 4.72-fold (95% CI 2.34 to 7.64) against 5.05-fold on all grades, and the negative control fails to discriminate in both (0.52-fold restricted). The restricted design retains 83% power against the all-grades effect size, so the agreement is informative rather than a wide interval covering everything. **b**, Whole-school enrollment by whether the grade range includes kindergarten, on a log scale: the restriction removes the larger middle and high schools, which is why it is the relevant test of whether school size drives the ranking. Grade range is recorded before the outbreak and is independent of exposure, so the subset cannot select on the outcome. Values in Supplementary Table 29.

**Supplementary Fig. 13.**
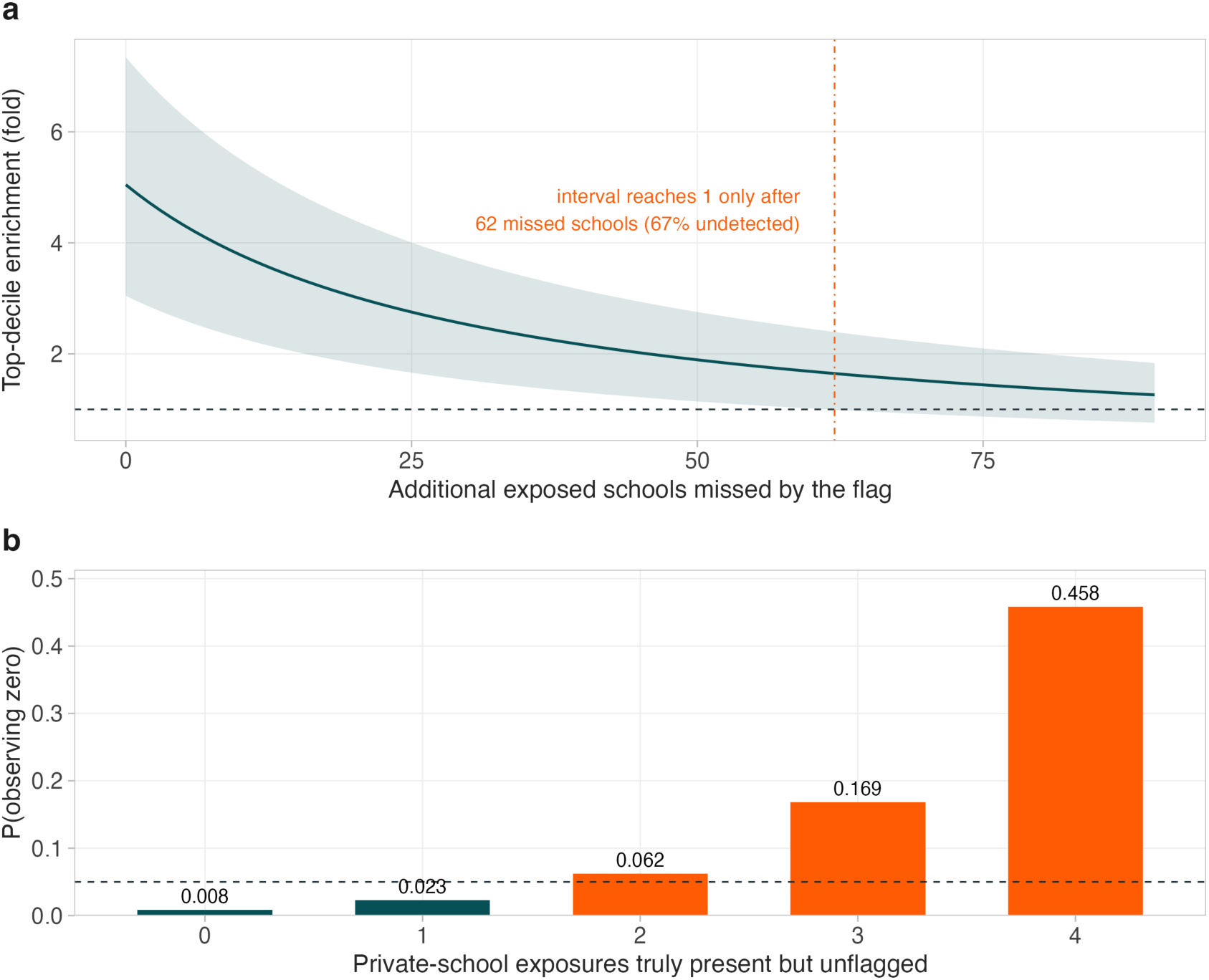
Flag completeness and the private-school zero. **a**, Top-decile enrichment as hypothetical missed exposures are added to the 30 recorded ones, each assigned outside the top decile: the assumption that reduces enrichment as fast as arithmetically possible. Band: 95% Clopper-Pearson interval. The interval reaches 1 only after 62 missed schools, a 67% non-detection rate, and the private-school zero stops being surprising once two exposures go unflagged (Supplementary Table 52). **b**, Probability of observing zero flagged exposures among the 77 Upstate private schools, against the 4.95 expected from enrollment and coverage alone, as a function of how many private exposures are assumed present but unflagged. Two suffice to make the zero unsurprising, so it is reported as uninterpretable rather than as a sector effect. Five state-run special-education schools are classified as public, not private (Supplementary Table 53). The bound also holds when the missing flags are assumed to track coverage. Assigning hypothetical missed exposures preferentially to above-median-coverage schools, the direction that would manufacture the enrichment from investigation intensity rather than exposure, leaves enrichment at 1.94-fold with 62 added (p = 9 × 10⁻⁴); in the rank-ordered worst case, where every missed exposure goes to the highest-coverage unflagged schools, significance holds to 62 (1.65-fold, p = 0.020) and is lost at 80. The enrollment negative control does not address this threat, because it is coverage rather than school size that would drive investigation intensity.

**Supplementary Fig. 14.**
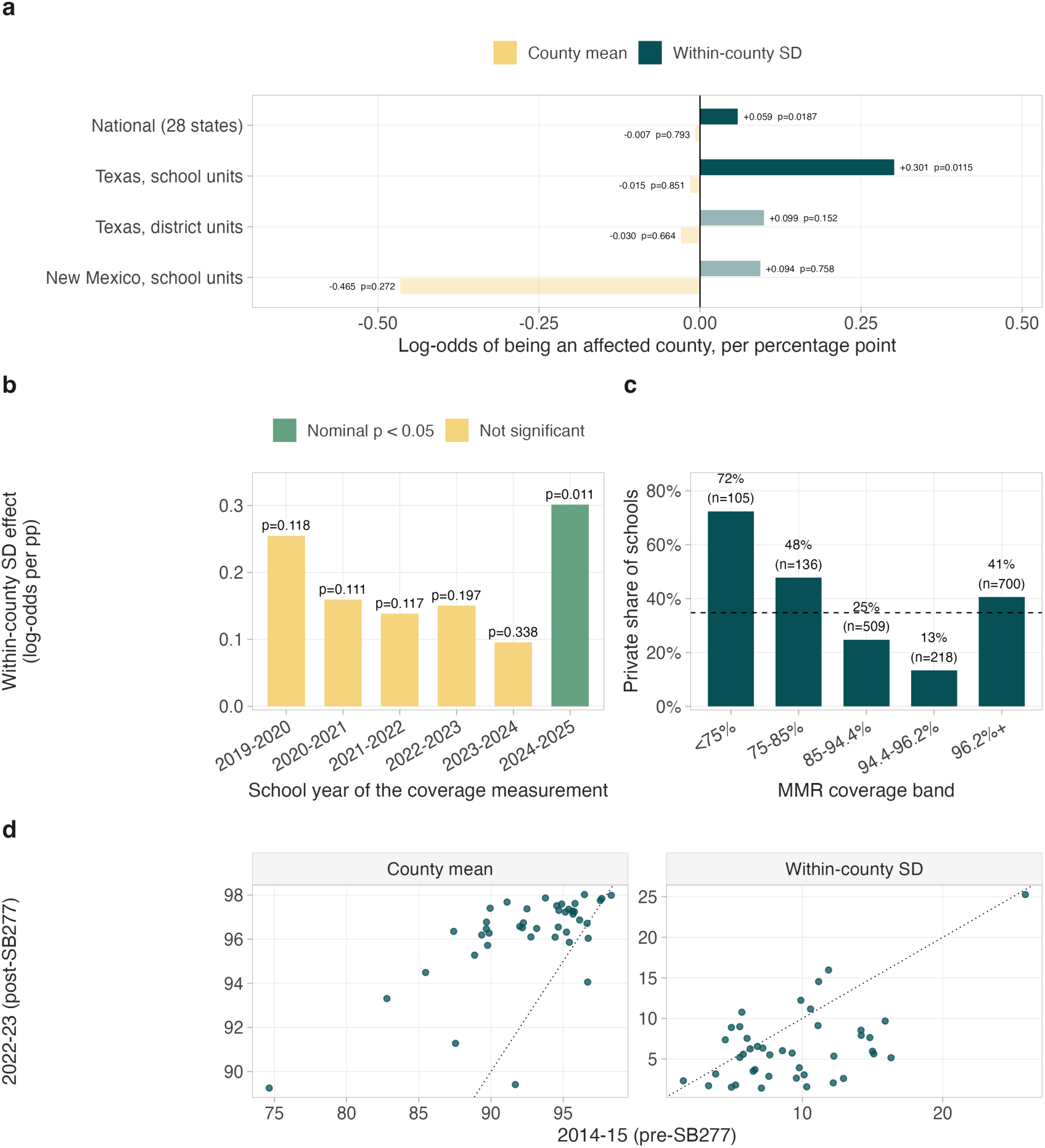
State-level replication of the county clustering model. **a**, Coefficients from the county model fitted separately by panel and unit of analysis, all using the same outcome as the national fit. Faded bars are not significant at *p* < 0.05. Texas at school resolution reproduces the national asymmetry with a larger coefficient; the same fit on district aggregates does not, because a district average is taken over the within-county variation the test is looking for. New Mexico is underpowered rather than contradictory. **b**, The Texas within-county spread coefficient by the school year of the coverage measurement. All six years are positive; only 2024-25, the year before the outbreak, reaches nominal significance, and it does not survive Bonferroni correction for six looks. **c**, Private share of Texas campuses by coverage band, from the 2023-24 named file; the dashed line is the overall private share of 34.8%. **d**, County mean coverage and within-county spread in 2014-15 against 2022-23, for the 42 California counties with at least ten schools matched across the SB277 boundary. Means track the identity line far more closely than spreads do.

**Supplementary Fig. 15.**
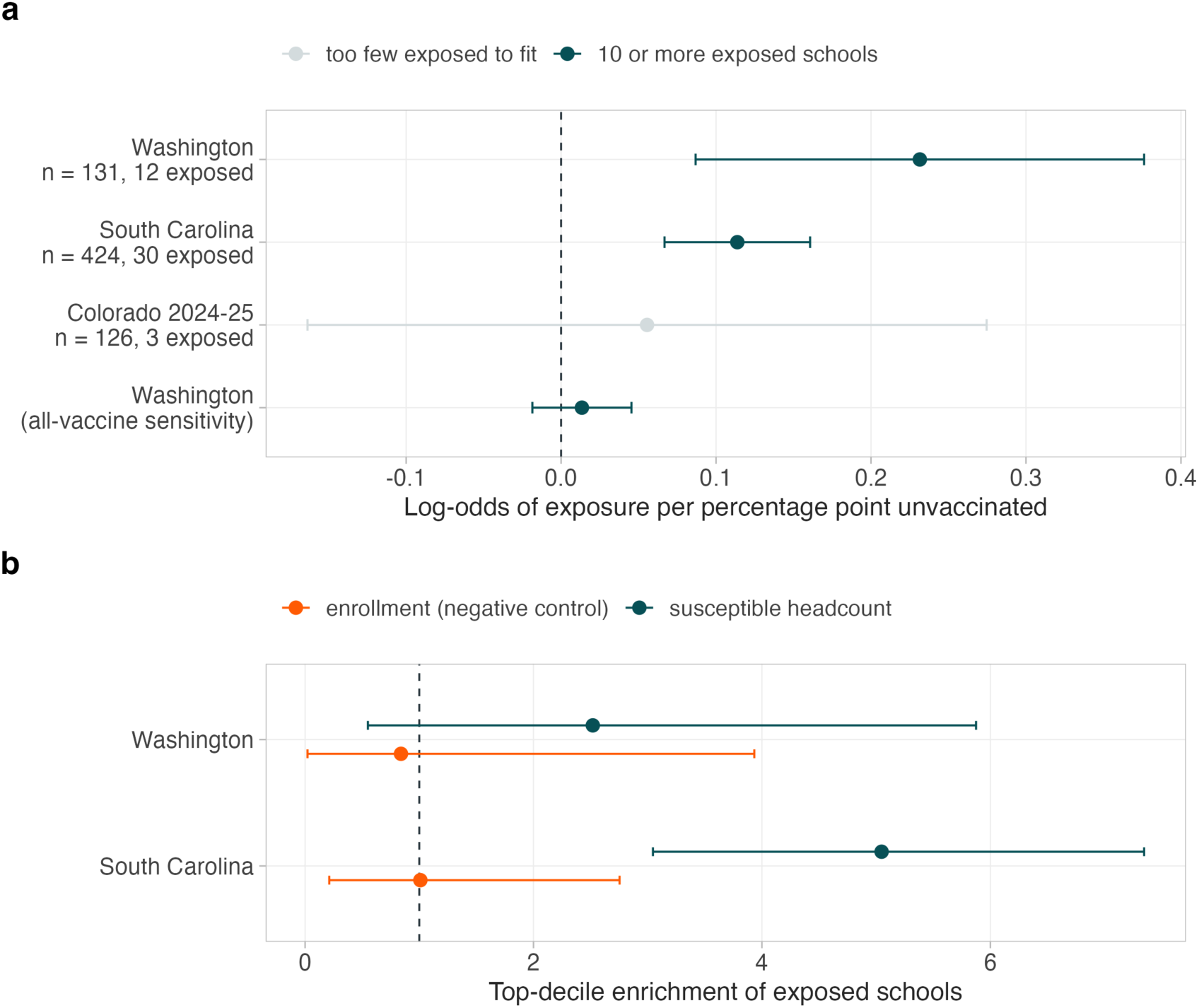
One independent replicate of the exposure-risk scaling, and why there is only one. **a**, Coefficient on percentage points unvaccinated from the paper’s own specification, exposure on log enrollment plus percentage unvaccinated, refit on each dataset that carries exposure flags, pre-outbreak coverage and a comparison set of unexposed schools from the same release. Bars are 95% Wald intervals. Clark County, Washington, 2018-19 gives +0.232 (SE 0.074, *p* = 0.00171) against +0.114 (SE 0.024) in Upstate South Carolina: the sign and significance replicate, the magnitude is roughly double. Colorado is shown for completeness and grayed, because three exposed schools cannot support a coefficient (+0.055, SE 0.112). The Washington all-vaccine series is a sensitivity, not a second replicate: Washington published only the MMR exemption rate at whole-school level in 2018-19, and the all-vaccine Complete series in the same file is a different quantity correlating at 0.16 with it across these schools; it gives +0.013 (SE 0.016), so what replicates is the direction under the MMR-specific measure, not the magnitude under any measure. **b**, Top-decile enrichment of exposed schools, with enrollment alone as a negative control, for the two datasets with ten or more exposed schools. Rockland County, New York, 2018-19 (7 exposed schools) publishes coverage for exposed schools only, with no unexposed denominator from the same reporting frame; Utah 2025-26 (20) now has both a statewide comparison set and an enrollment denominator but publishes a visit list rather than an investigation register, so its exposed schools cannot be identified within that frame; and Ridgefield 2024-26 carries one exposed school; none can yield a coefficient, and they are listed with their reasons in Supplementary Table 38.

**Supplementary Fig. 16.**
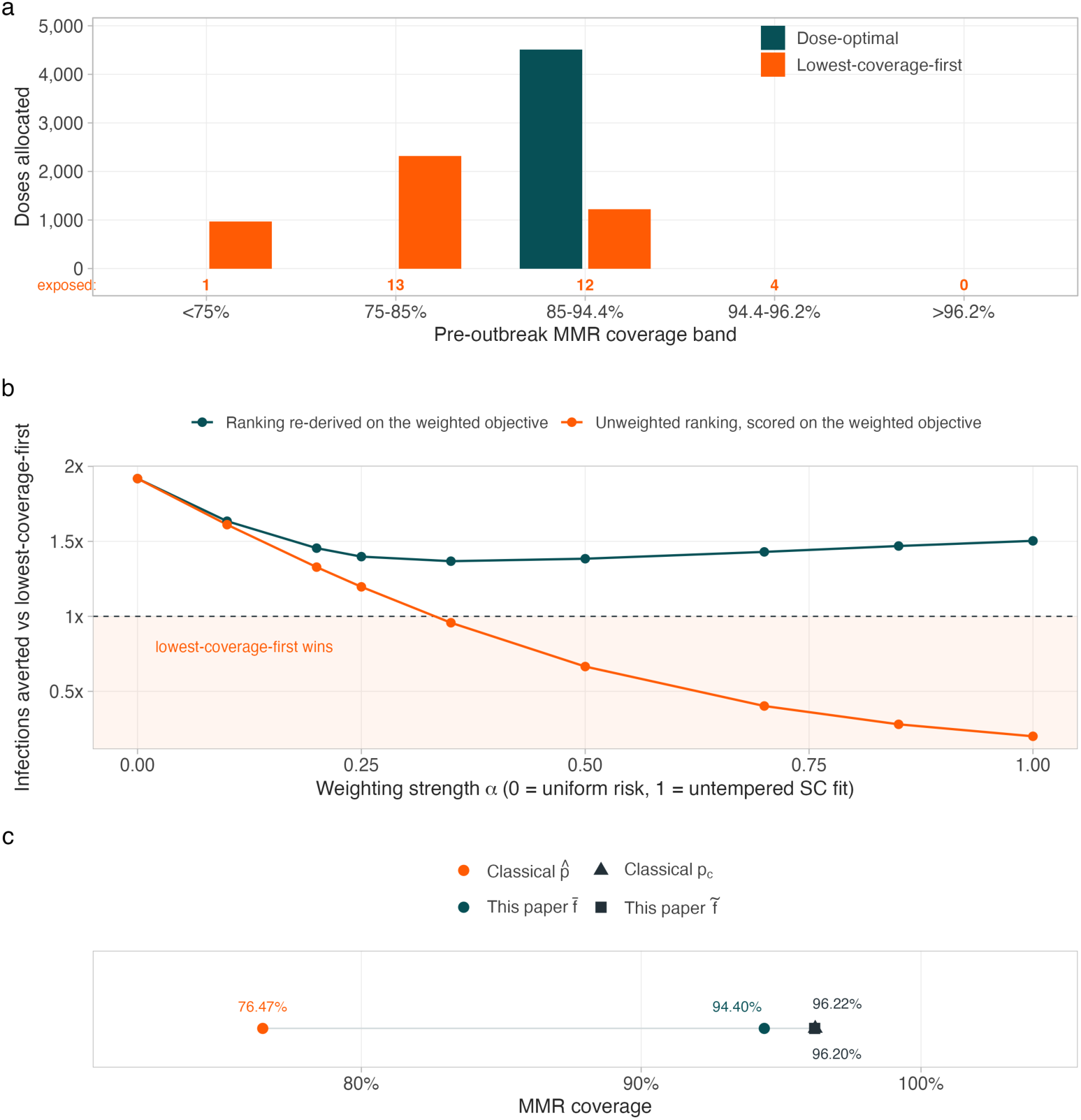
Introduction risk is not uniform at the allocation scale. **a**, Doses allocated by each rule across pre-outbreak coverage bands in Upstate South Carolina at a 4,510-dose budget (25% of susceptibles), with the number of later-exposed schools in each band beneath the axis. The dose-optimal rule allocates nothing at or below 85% coverage, where 14 of the 30 exposed schools sat. **b**, Infections averted relative to lowest-coverage-first as the introduction-risk weighting strengthens. *α* = 0 is the uniform-risk objective used throughout the paper and *α* = 1 the untempered fit to the South Carolina exposures; intermediate values temper the out-of-sample extrapolation. Teal: the marginal-efficiency ranking re-derived on each weighted objective. Orange: the unweighted ranking scored on that same objective. Shading marks where lowest-coverage-first wins. Magnitudes are a bounded sensitivity, not a calibrated national risk surface. **c**, The allocation objective recovers the classical threshold. The vaccine-adjusted herd-immunity threshold *p_c_* = (1 − 1/*R*_0_)/*VE*^1^ (96.22%, triangle) and the saturation target *f̄* at which the herd-effect function stops repaying doses (96.20%, square). The breakthrough-maximizing coverage *p̂* (76.47%) and the convex-concave inflection *f̄* (94.40%) are plotted for scale. The two are the same root of the same equation rather than independent estimates, so the panel fixes the coverage scale the allocation rule acts on rather than corroborating it (Supplementary Note 3, Supplementary Table 47).

**Supplementary Fig. 17.**
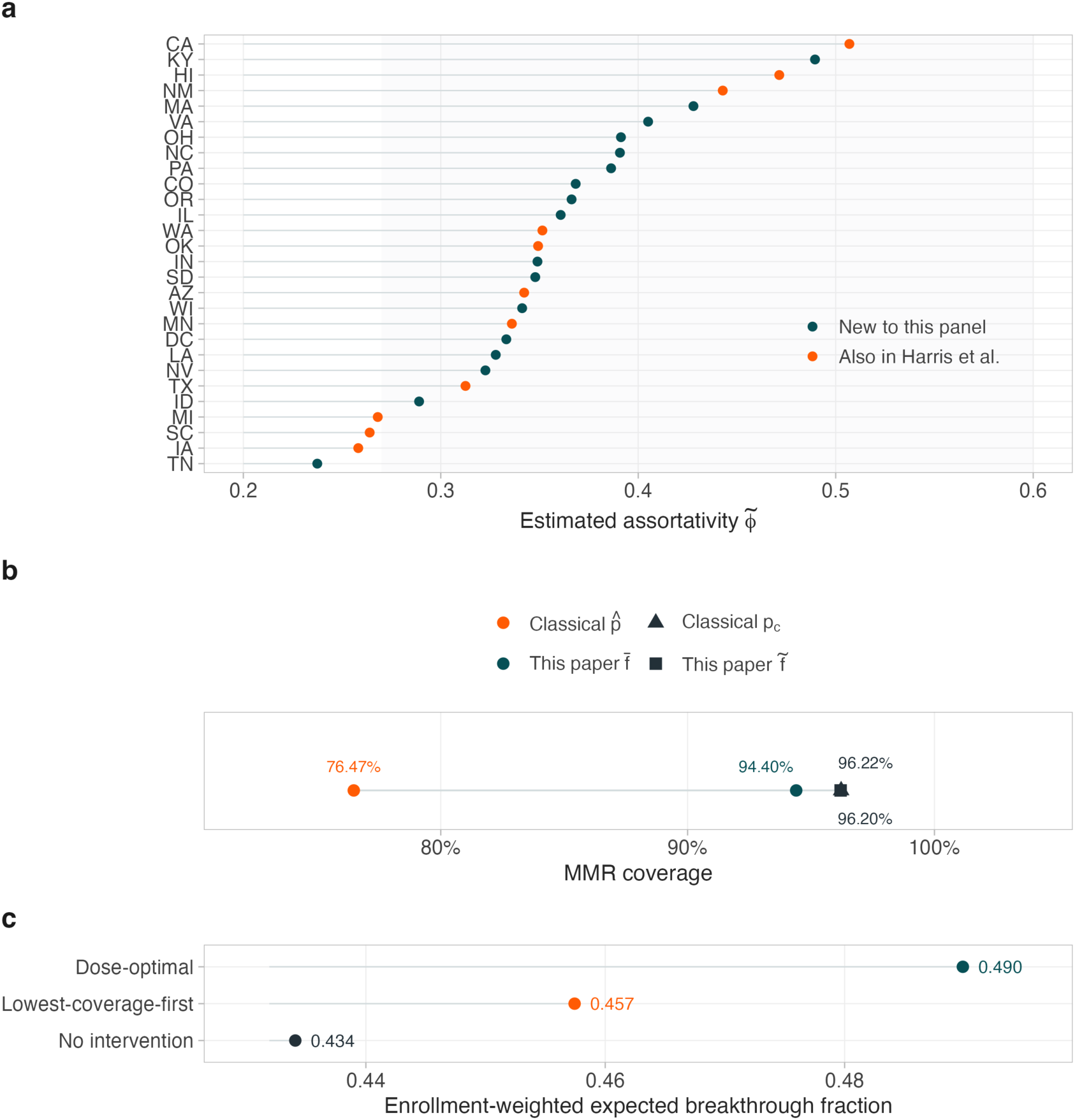
Assortative mixing, threshold correspondence and the breakthrough fraction. **a**, The assortativity estimator of Harris et al. ^10^ computed on each state panel, enrollment-weighted. Every state exceeds 0.2 as they report, with median 0.35 against their 0.37; shading marks their reported range and color distinguishes the 17 states absent from their analysis. **b**, Their vaccine-adjusted herd-immunity threshold and the Duijzer saturation target coincide at 96.2%, two derivations of one quantity. Their breakthrough-maximizing coverage is a whole-population equilibrium result shown for context only. **c**, Enrollment-weighted expected breakthrough fraction at post-allocation coverage under each rule. The dose-optimal rule raises it more, a communication cost of concentrating doses in higher-coverage schools; total infections still fall.

**Supplementary Fig. 18.**
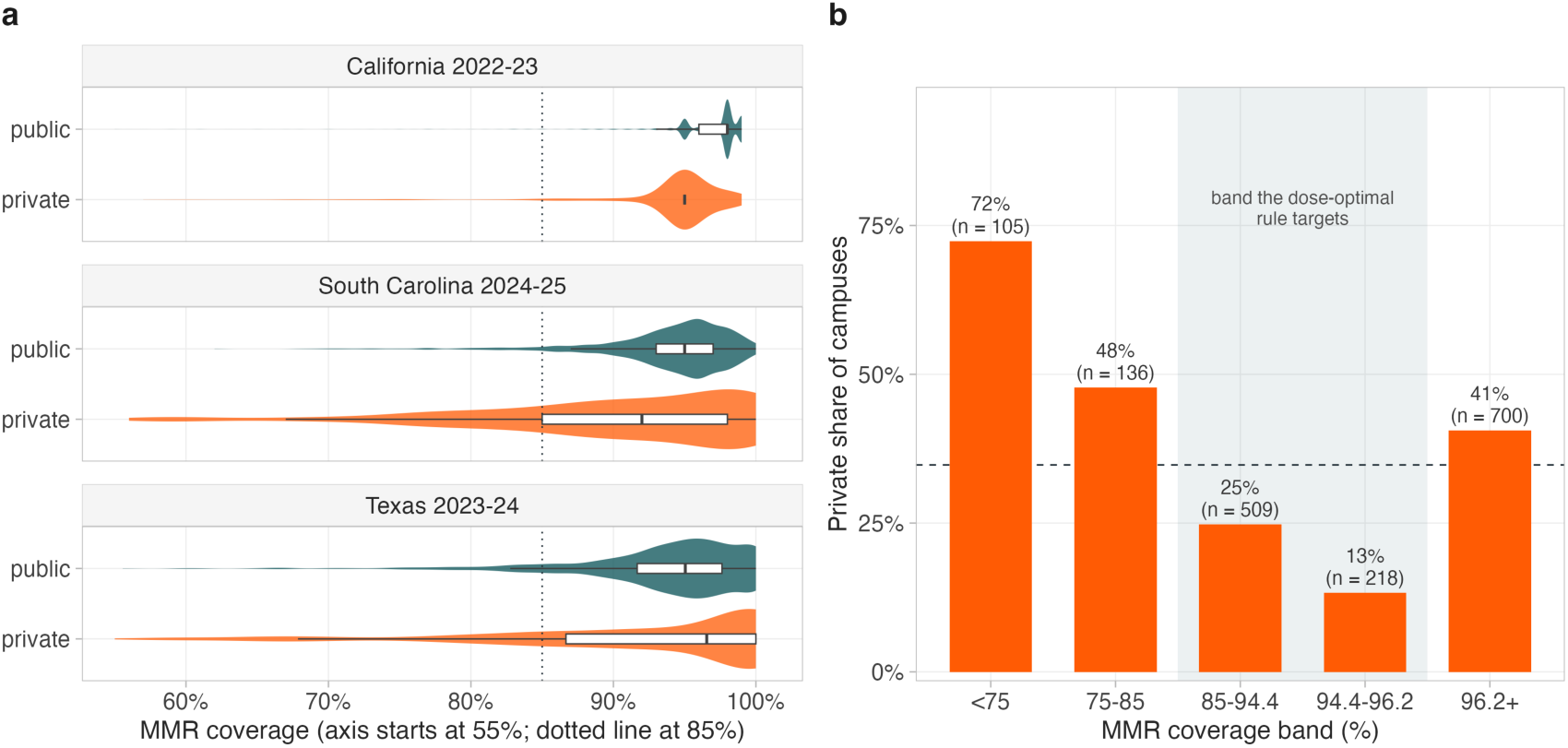
Private-school coverage is more variable than public, and sector composition varies across the coverage distribution. **a**, MMR coverage by reported sector for the three states that flag school sector, shown as violins with interquartile boxes. The axis starts at 55% because private density below that point is negligible in every state, and the dotted line marks 85%. Private coverage is more dispersed in all three states, with standard-deviation ratios of 1.26, 2.28 and 1.96 (all *p* < 0.001, variance-ratio tests) and 5th percentiles of 76.5% against 90.0% in California, 60.0% against 84.0% in South Carolina and 58.8% against 81.1% in Texas. The odds that a campus falls below 85% coverage are 2.18, 6.34 and 3.18 times higher for private schools. Medians differ little and in Texas the private median is 0.8 points *higher* (*p* = 0.15), so the sector difference is one of variance and of the low tail rather than of central tendency (Supplementary Tables 54 and 55). **b**, Private share of Texas campuses by coverage band, with the band the dose-optimal rule targets shaded and the statewide private share of 34.8% dashed. The share falls from 72.4% below 75% coverage to 13.3% in the 94.4 to 96.2% band, then rises to 40.6% in the top band. That rise is a reporting ceiling rather than a reversal: 40.5% of Texas private campuses report exactly 100% coverage against 13.4% of public, and 16.9% against 2.1% in South Carolina (Supplementary Table 4). Private schools are over-represented at both extremes of the distribution, which is the variance result of panel a seen from its upper end. California cannot contribute to this comparison because its 2022-23 release is capped at 99%, with no school reported at 100%.

**Supplementary Fig. 19.**
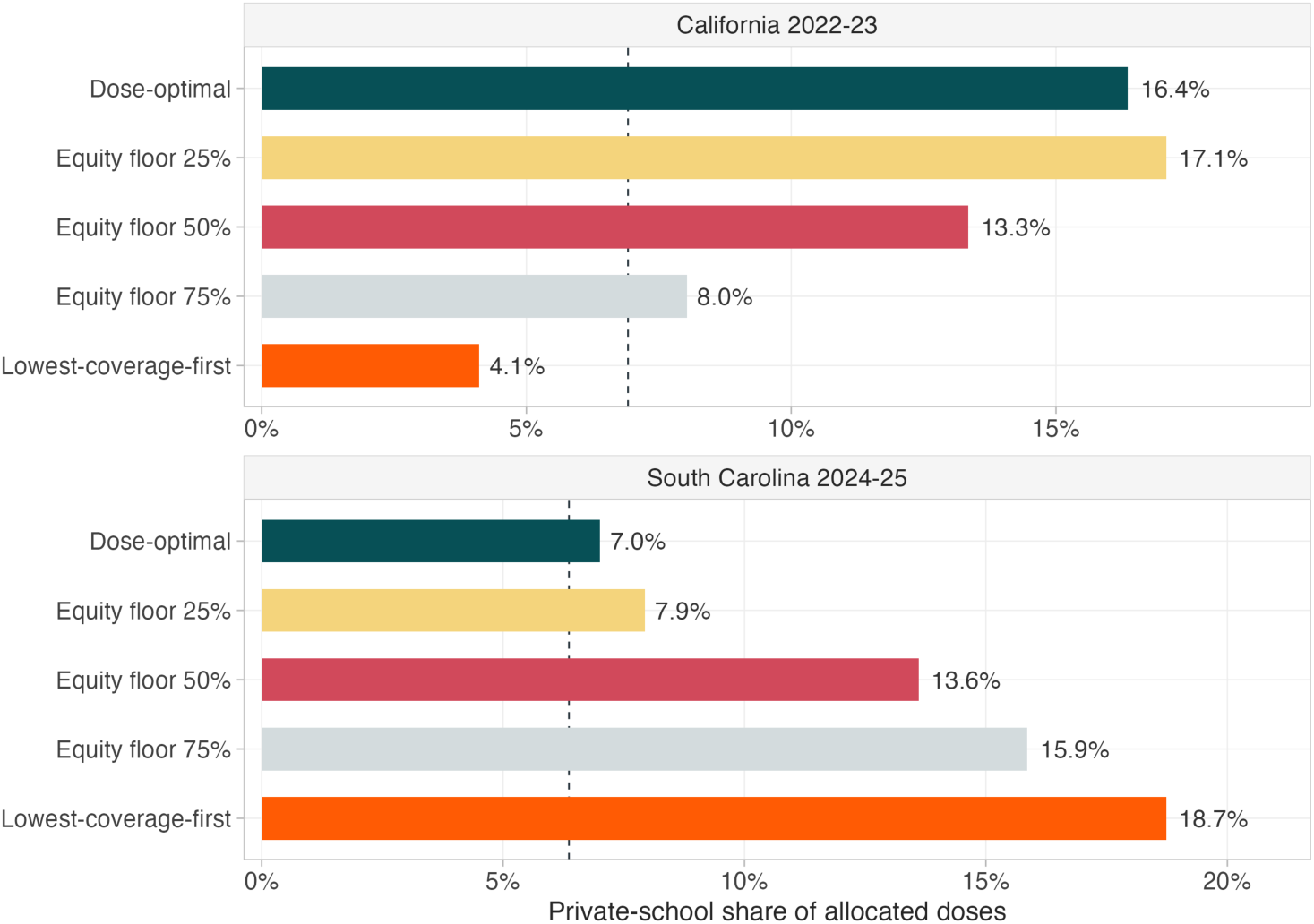
Private-school share of allocated doses, by rule. Share of the doses each rule assigns that goes to private schools, at an identical 25% budget, for the two states that report sector alongside the enrollment needed to form a dose share. The dashed line marks each state’s private share of kindergarten enrollment, 6.9% in California and 6.4% in South Carolina. The direction of over-representation reverses between the two states and the equity floors do not move it consistently, which is why this is reported as a property to check locally rather than as a national estimate.

**Supplementary Fig. 20.**
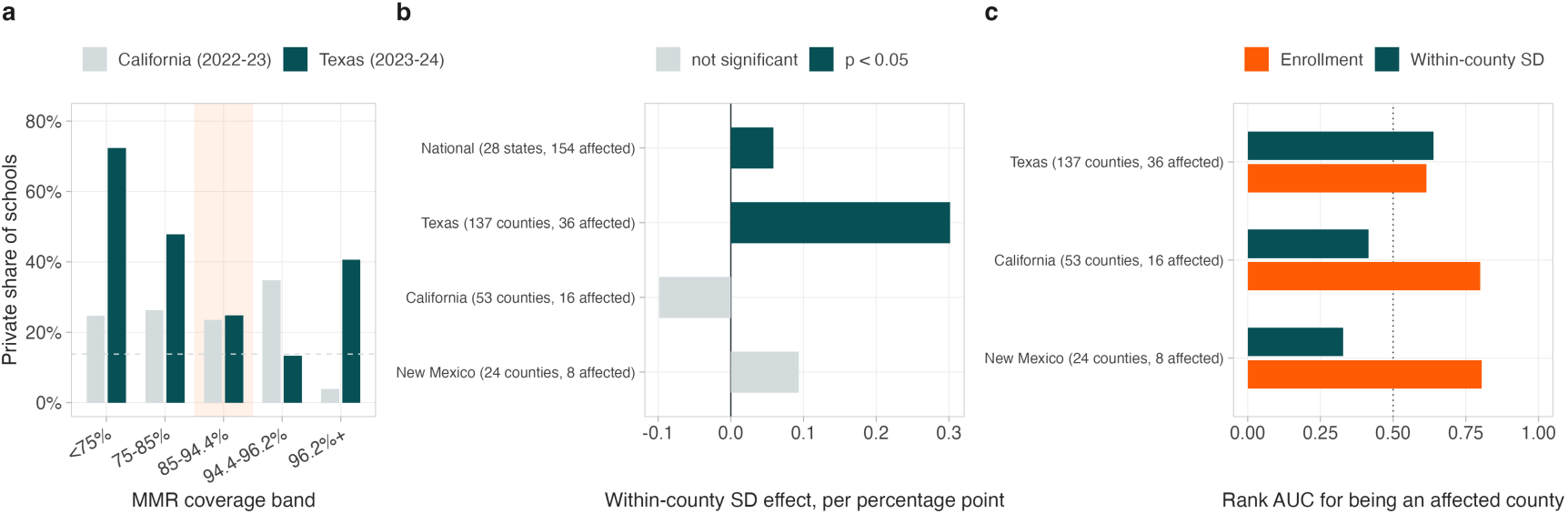
California replication of the county clustering test and the sector position. **a**, Private share of schools by MMR coverage band in California (2022-23) and Texas (2023-24); dashed lines are each state’s overall private share and the shaded column is the band the dose-optimal rule targets. Texas concentrates private schools in the low-coverage tail; California does not (odds ratio 1.09 below 85% against the targeted band, *p* = 0.64). **b**, Within-county coverage-spread coefficient for being an affected county, on one outcome definition across all four panels: at least one case in our own county series. **c**, Rank AUC for being an affected county, log enrollment against within-county spread. Clustering replicates only where size does not already account for reach: enrollment out-predicts spread by 0.48 in New Mexico and 0.38 in California, but the two are level in Texas (−0.02), where the spread coefficient is largest. Values in Supplementary Tables 68, 69, 56 and 57.

**Supplementary Fig. 21.**
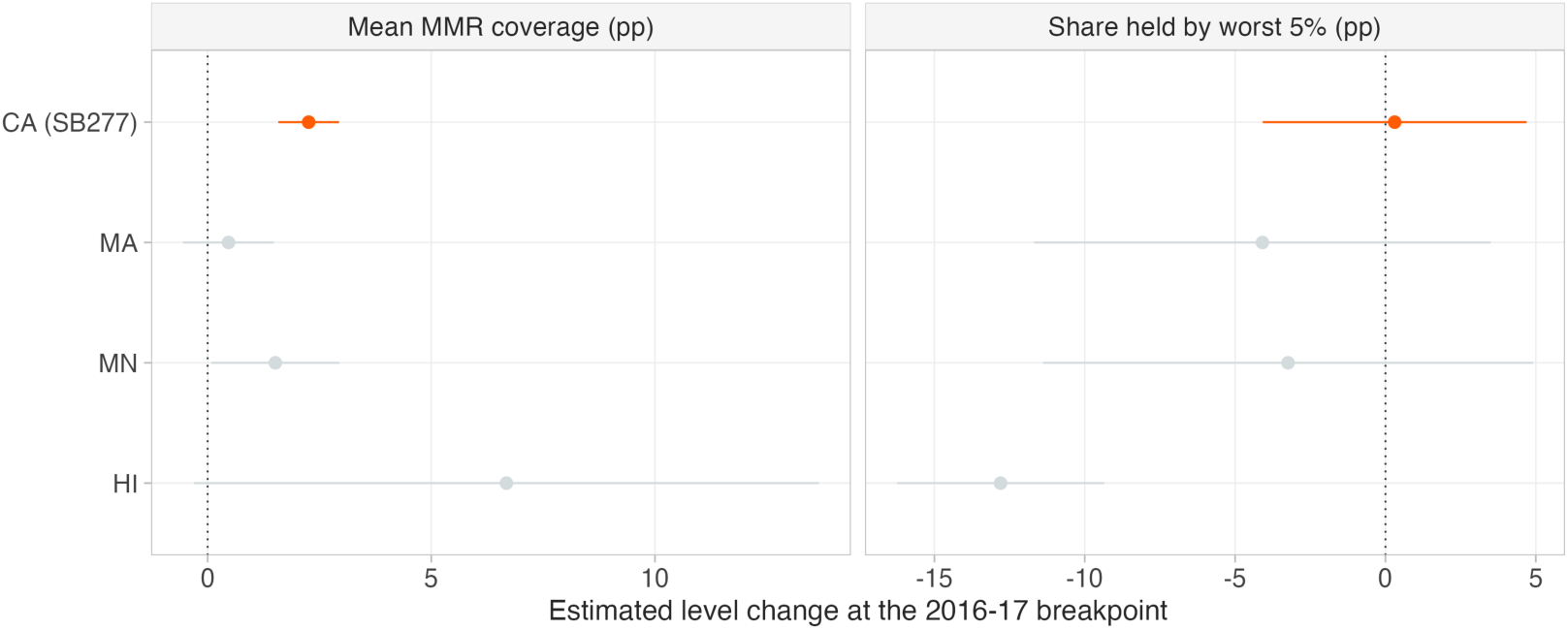
Placebo test. Estimated level change at the 2016-17 breakpoint for California and for the three states with sufficient pre-period data and no mandate change. Error bars are Newey-West 95% confidence intervals.

**Supplementary Fig. 22.**
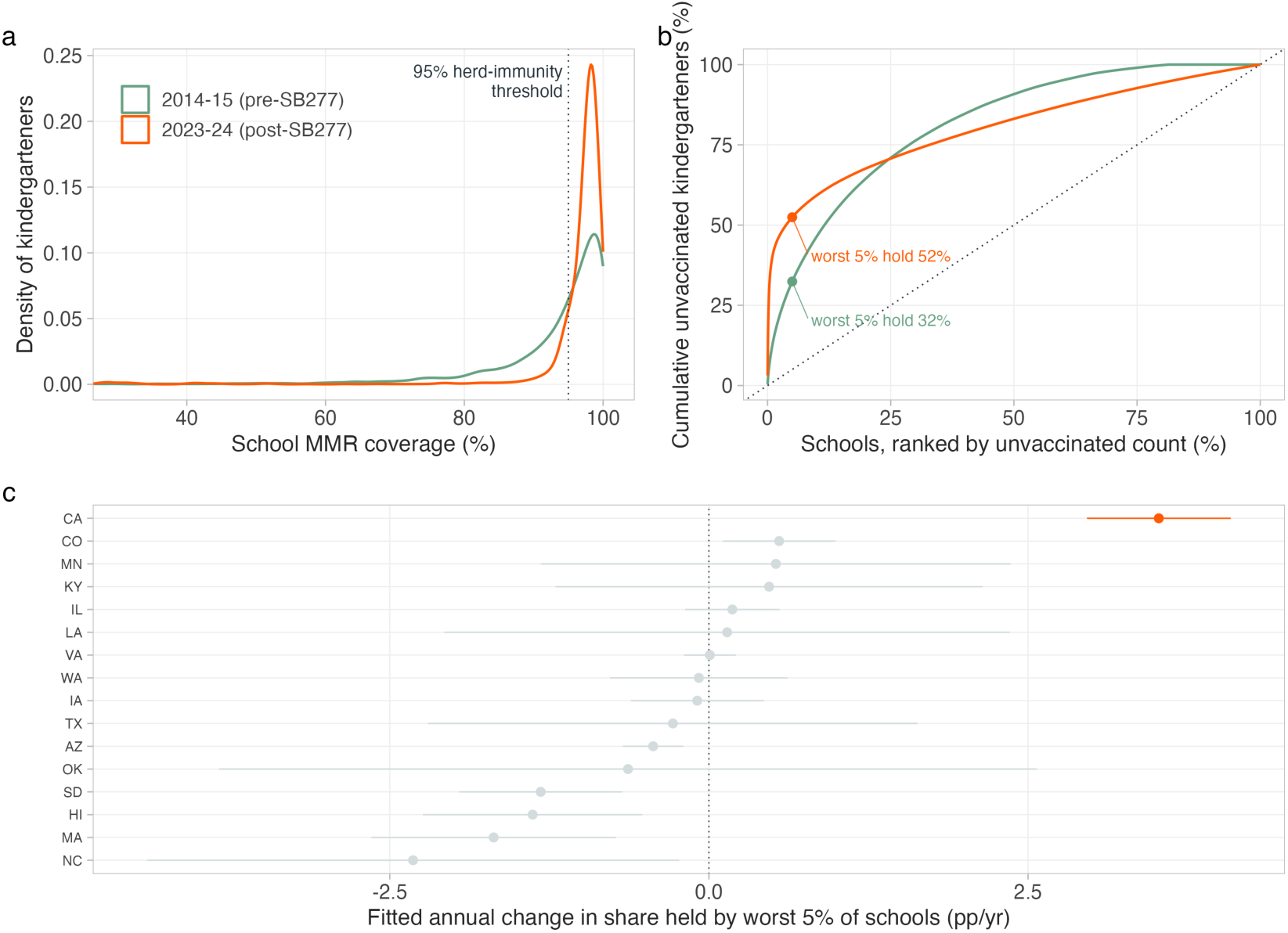
California: a coverage mandate and the concentration it did not change. A ten-point annual series cannot support causal inference about the mandate (Supplementary Table 59), so this figure is descriptive. **a**, Enrollment-weighted distribution of school MMR coverage before and after SB277. The mean rose and the low tail thinned but did not disappear. **b**, Lorenz curves of unvaccinated kindergarteners by school. The bottom5% of schools held 32% of the state’s unvaccinated kindergarteners before the mandate and 52% after. **c**, Fitted annual change in the share held by the bottom 5% of schools, with Newey-West 95% confidence intervals, for the 16 retained states with ≥4 reported years. California’s +3.52 points per year is the steepest and its interval overlaps no other state. This is a descriptive contrast across states, not an inference from the breakpoint.

**Supplementary Fig. 23.**
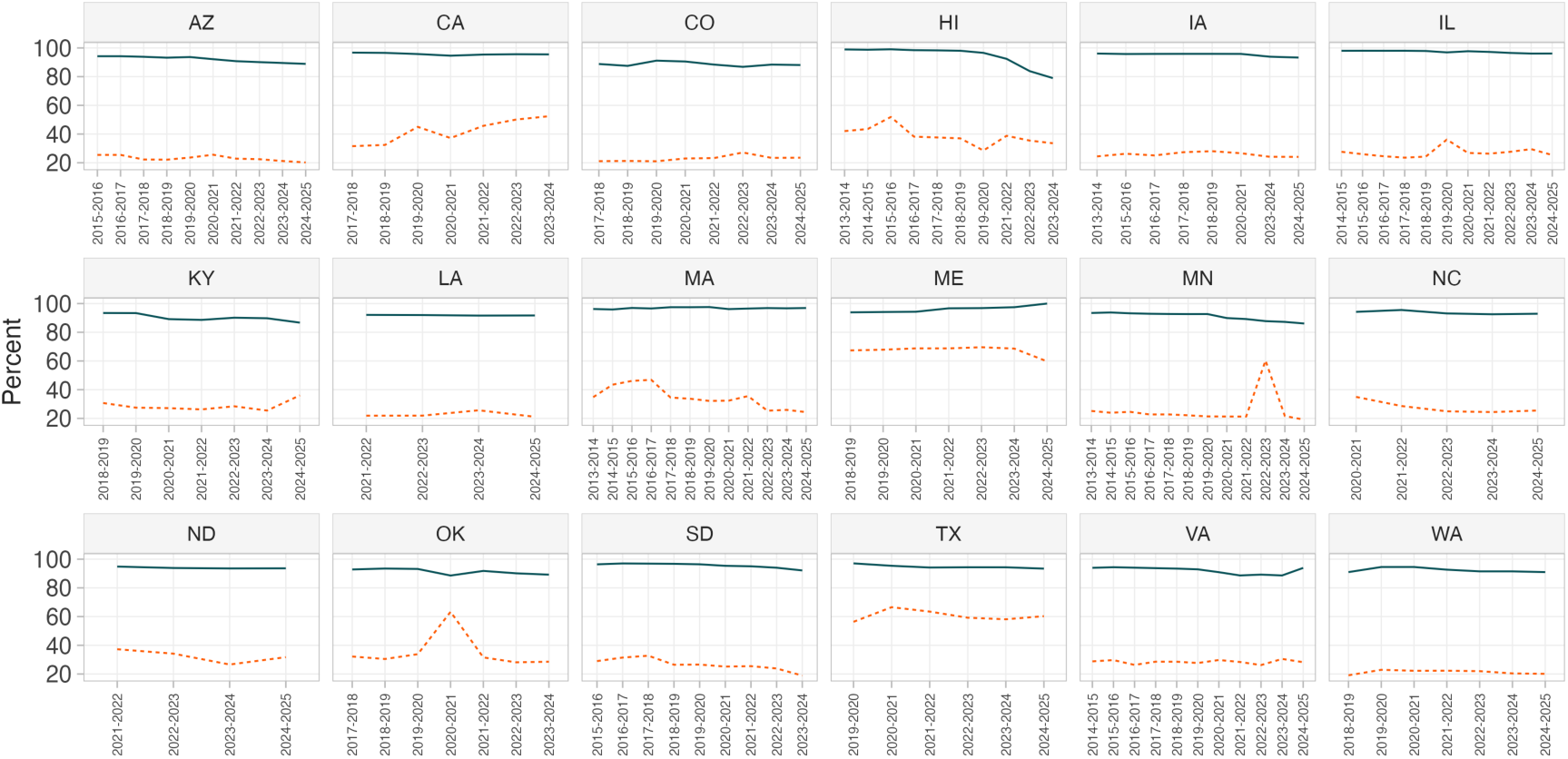
Trend panel. Mean coverage (solid) and share held by the bottom 5% of schools (dashed) for every state with at least four reported years.

**Supplementary Fig. 24.**
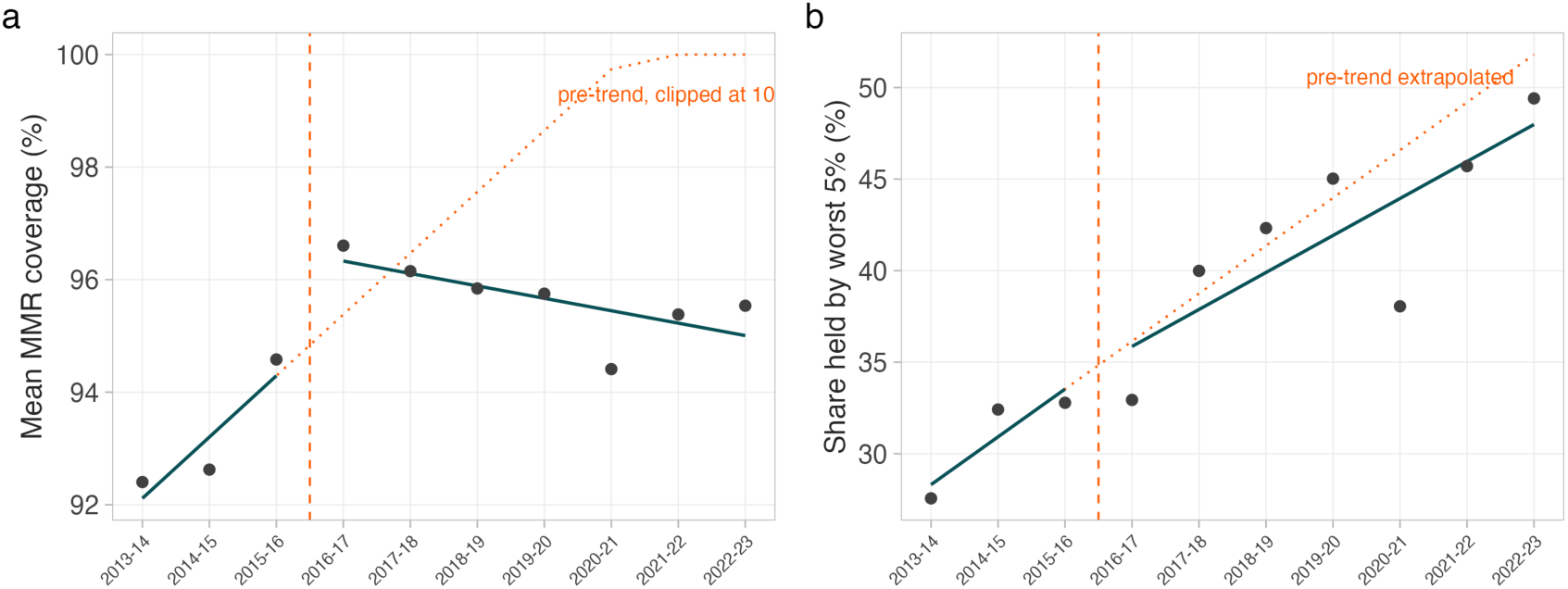
Interrupted time series, SB277. **a**, Mean coverage. **b**, Share held by the bottom 5% of schools. Points are observed annual values; solid lines are the segmented fit; the dotted line is the pre-trend extrapolation (clipped at 100% in **a**); the dashed vertical line marks the 2016-17 interruption.

**Supplementary Fig. 25.**
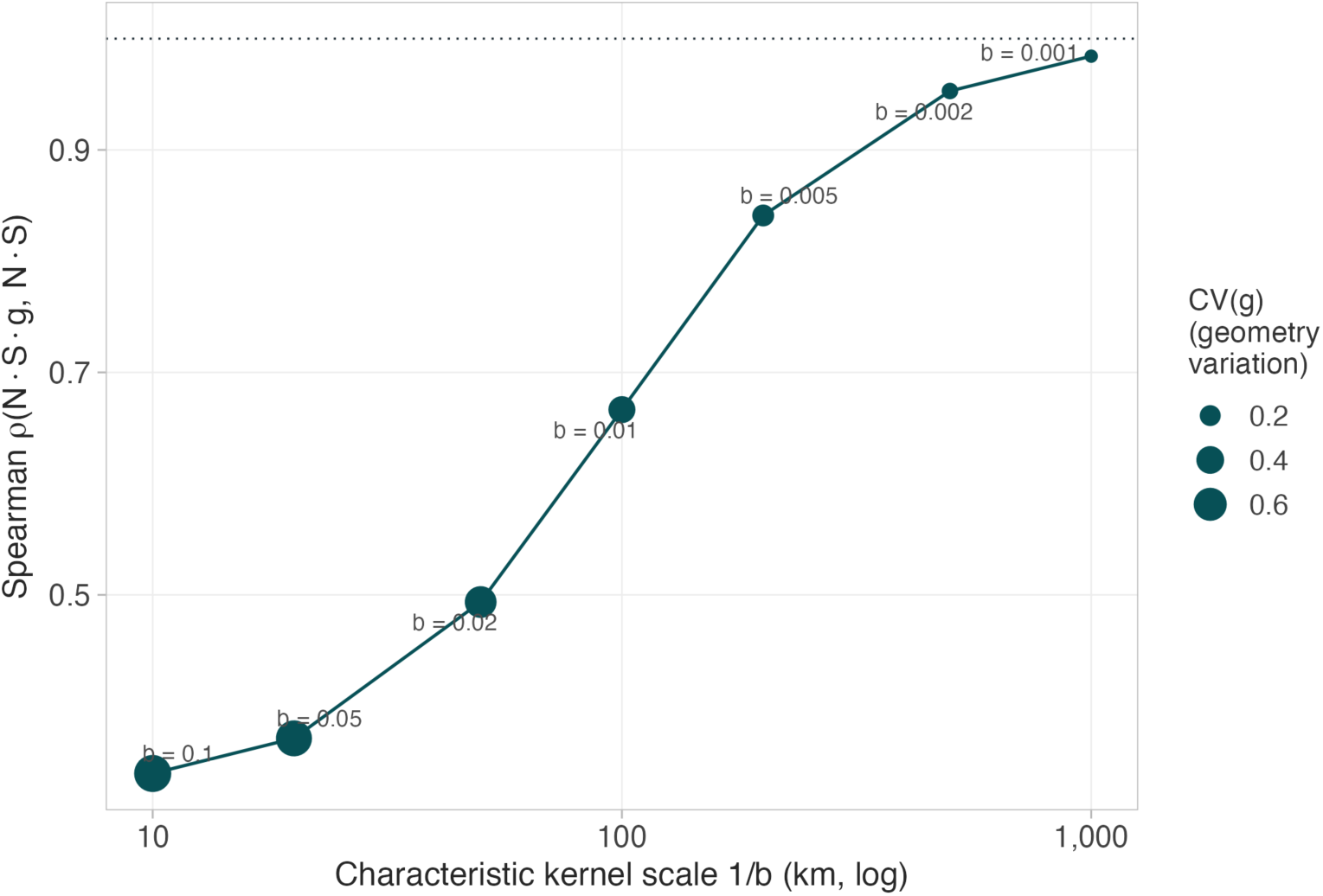
Spatial scope of the distance-decay kernel. Coefficient of variation of the geometry term *g* and Spearman correlation between *R_v_* and the raw susceptible count, across decay parameters from 10^−3^ to 10^−1^ km^−1^. Point area is proportional to *CV(g)*.

**Supplementary Fig. 26.**
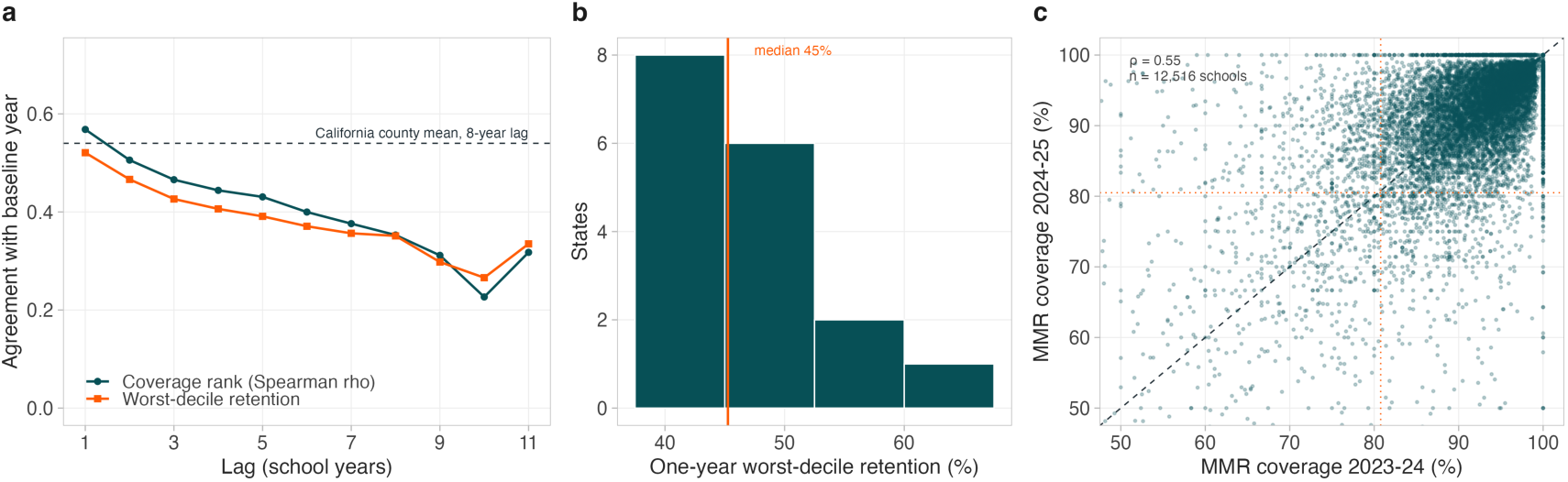
How long a school-level coverage ranking stays valid. **a**, Agreement between a school coverage ranking and the same ranking at a later year, against lag, for the 27 analysis-panel states whose coverage field is not a repeated snapshot. Teal is the Spearman rank correlation, the quantity the dose-optimal rule reads; orange is the share of the bottom decile still in the bottom decile, the set a lowest-coverage-first rule doses. Dashed line marks the California county mean over eight years for comparison (Spearman *ρ* = 0.54, Supplementary Table 58). **b**, Distribution across states of one-year bottom-decile retention; 16 of 17 states with enough consecutive years fall below 60%, median 45% (orange line). **c**, The most recent one-year comparison in the panel, one point per school; dashed line is equality and dotted lines mark the bottom-decile thresholds in each year. Louisiana is excluded throughout because its coverage field repeats across years while its enrollment field does not (Supplementary Table 82); no reported result depends on the exclusion. Values in Supplementary Tables 79 and 80.

**Supplementary Fig. 27.**
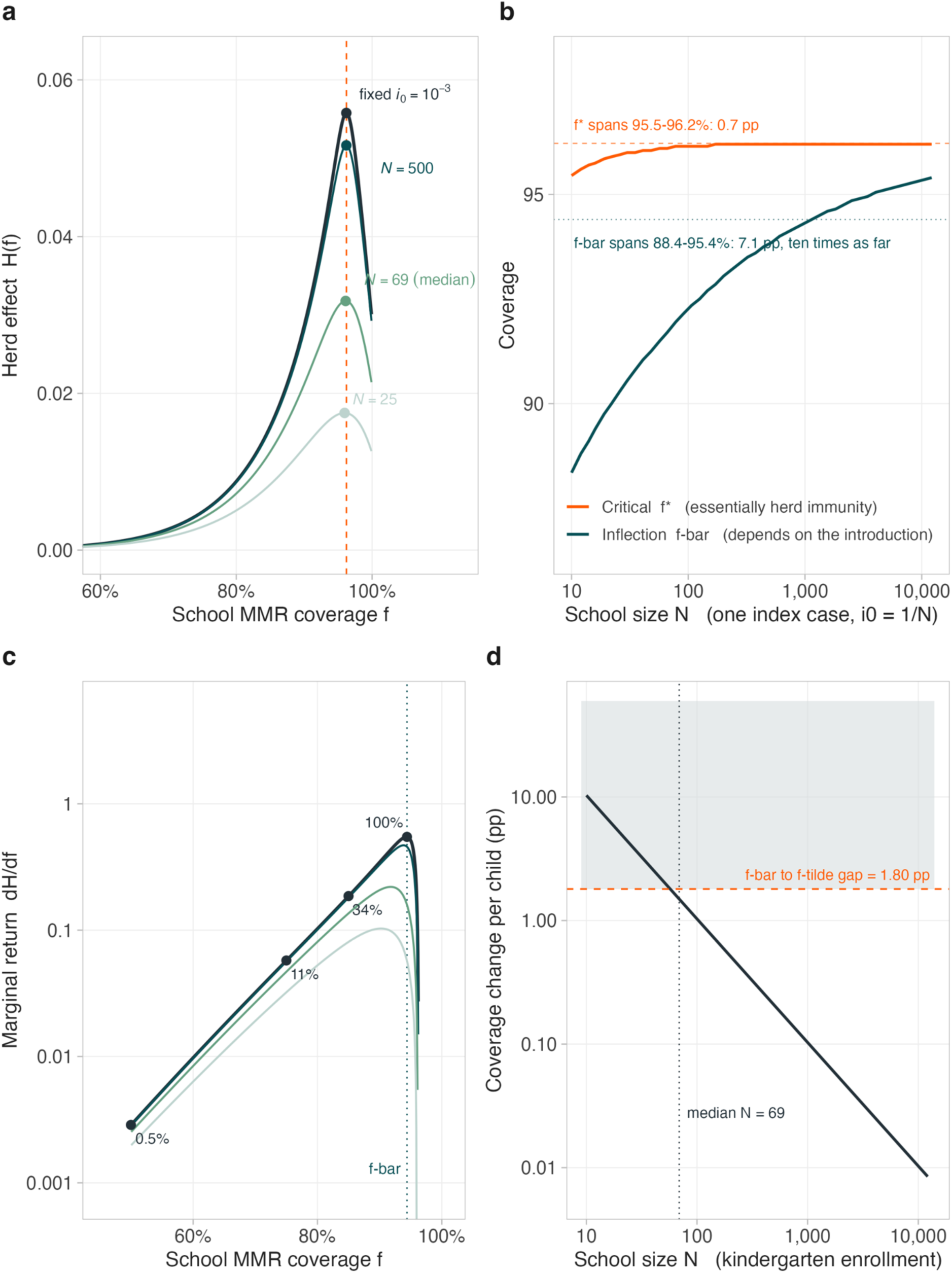
Which threshold is a property of *R*_0_, and which is a property of the introduction. **a,** The herd effect *H*(*f*) under four seeding specifications: the paper’s fixed *i*_0_ = 10^−3^ and one index case in a school of 500, 69 (the median) and 25. All four curves are H: the same deterministic continuous-coverage function evaluated at four different assumed introductions. None is *G*, which is reserved for the integer-lattice and exact stochastic quantities (Supplementary Figs. 27 and 28). A larger assumed introduction lowers the curve and moves its bend far more than its peak: every peak sits at or just below the classical vaccine-adjusted threshold (1 − 1/*R*_0_)/*VE* = 96.2% (dashed), which the peak approaches exactly as *i*_0_ tends to zero. **b**, The two critical fractions against school size under one-index-case seeding. The critical fraction *f*^∗^ spans 95.5-96.2%, a range of 0.7 points, while the inflection *f̄* spans 88.4-95.4%, ten times as far: *f*^∗^ is governed by *R*_0_ and *VE* and moves only slightly with the seed at small *N*, whereas the inflection is a property of the expected return. **c**, Marginal indirect return *dH*/*df*, log axis, with the paper’s fixed specification in ink and the one-index-case family in green. Inside the convex region the return is accelerating but small: 40.3% of all susceptibles sit below 85% coverage, earning under a third of the peak return per dose. **d**, Coverage is an integer lattice. One child represents 100/(*VE* × *N*) percentage points, 1.49 at the median school of 69, so in the shaded region a single child moves coverage further than the entire 1.8-point interval between *f̄* and *f̄*: 39.2% of schools holding 16.5% of susceptibles.

**Supplementary Fig. 28.**
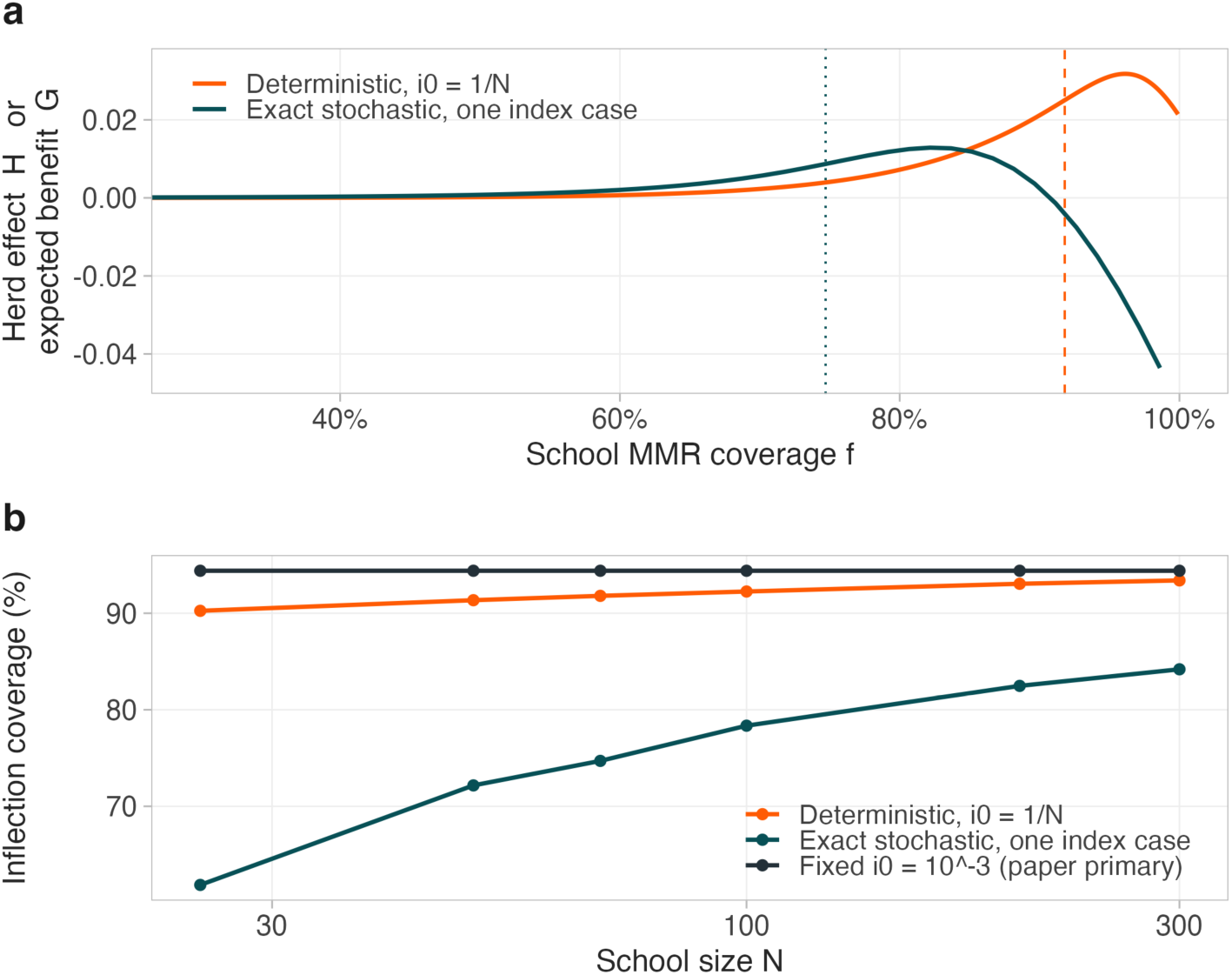
Exact stochastic final size with one index case, and what it does to the inflection. **a,** The expected benefit at the median enrollment of *N* = 69, under two specifications: *G*, the exact stochastic model with a single index case, evaluated on the achievable integer grid of coverages a school of 69 can actually reach, and *H*, the deterministic mean-field final-size relation with the seed fraction set to *i*_0_ = 1/*N*. The dotted vertical line marks the stochastic inflection at 74.7% and the dashed line the deterministic one at 91.8%, a gap of 17.1 percentage points at this enrollment. The vertical axis shows the full range of both curves. Above about 91% coverage the stochastic curve turns negative, reaching −0.0436 at its last achievable point, because the direct credit for every vaccinated child then exceeds total prevented; that trough is 3.4 times the height of the curve’s own peak. It lies well above the inflection and cannot affect it, since subtracting a linear function of coverage leaves a second derivative unchanged. The stochastic curve stops at 98.61% rather than at full coverage because a school of 69 cannot reach it: one child is worth 1.49 percentage points, so 66 vaccinated children give 98.61% and 67 would exceed 100%. **b**, The inflection of *G* for the stochastic series, and of *H* for the two deterministic comparators, against school size, on a logarithmic size axis, for three specifications: exact stochastic seeding with one index case, the deterministic relation at *i*_0_ = 1/*N*, and the paper’s primary fixed *i*_0_ = 10^−3^, which is flat by construction because it does not depend on *N*. The exact stochastic inflection is 61.9% at *N* = 25, 72.2% at *N* = 50, 74.7% at the median *N* = 69, 78.4% at *N* = 100, 82.5% at *N* = 200 and 84.2% at *N* = 300: between 9.2 and 28.4 points below the deterministic school-specific value and further still below the fixed specification, with the gap widening as schools get smaller. At these enrollments the probability that an introduction fails to establish, not the depletion of susceptibles, governs expected benefit; the deterministic relation has no extinction and so cannot represent it. Expectations are solved exactly by first-step analysis on the (*s*, *i*) chain and cross-checked against exhaustive path enumeration at small *N* and a Sellke-construction Monte Carlo.

## Supplementary Tables

**Supplementary Table 1.** Shape of the indirect benefit, total prevented and the direct component.

| quantity | inflection f | max abs d2 | marginal f=.25 | marginal f=.50 | marginal f=.95 |
| --- | --- | --- | --- | --- | --- |
| Indirect benefit H(f) (the allocation objective) | 0.944 | 73.454 | 0.000116 | 0.0029 | 0.5087 |
| Total infections prevented | 0.944 | 73.454 | 0.9691 | 0.9719 | 1.478 |
| Direct component (recipients immunized) | | $5.55 \times 10^{-9}$ | 0.97 | 0.97 | 0.97 |

**Supplementary Table 2.** Replication of published *R_v_* estimates.

| state | year | n schools | max abs diff | max abs diff<br>below 100 | median ratio | correlation | mean<br>deposited | mean<br>reproduced |
| --- | --- | --- | --- | --- | --- | --- | --- | --- |
| AR | 2024-2025 | 471 | 0.0482 | 1.42e-14 | 1. | 0.9999 | 0.2677 | 0.26 |
| AZ | 2021-2022 | 1,112 | 0.0256 | 2.06e-13 | 1. | 1. | 1.372 | 1.372 |
| CA | 2019-2020 | 6,556 | 5.4e-13 | 5.4e-13 | 1. | 1. | 0.6261 | 0.6261 |
| CO | 2020-2021 | 1,368 | 0.033 | 0.0058 | 1. | 1. | 1.411 | 1.41 |
| CT | 2024-2025 | 460 | 0.0355 | 4.88e-15 | 1. | 0.9999 | 0.2546 | 0.2476 |
| DC | 2024-2025 | 153 | 0.0201 | 4.44e-15 | 1. | 1. | 1.04 | 1.039 |
| HI | 2014-2015 | 410 | 0.0895 | 1.24e-14 | 1. | 0.9994 | 0.1811 | 0.1743 |
| IA | 2015-2016 | 1,391 | 0.0106 | 0.000601 | 1. | 1. | 0.6302 | 0.63 |
| ID | 2023-2024 | 465 | 0.0143 | 1.95e-14 | 1. | 1. | 2.712 | 2.712 |
| IN | 2024-2025 | 1,102 | 0.0738 | 0.0059 | 1. | 1. | 1.378 | 1.377 |
| KY | 2018-2019 | 817 | 0.0621 | 5.33e-14 | 1. | 1. | 0.9838 | 0.982 |
| LA | 2021-2022 | 1,053 | 0.033 | 1.42e-14 | 1. | 1. | 1.188 | 1.186 |
| ME | 2019-2020 | 363 | 0.0087 | 8.53e-14 | 1. | 1. | 0.8784 | 0.878 |
| MI | 2024-2025 | 2,138 | 0.0322 | 0.0018 | 1. | 1. | 1.599 | 1.599 |
| MN | 2021-2022 | 1,170 | 0.0278 | 0.0021 | 1. | 1. | 1.606 | 1.605 |
| NC | 2023-2024 | 1,706 | 0.0316 | 0.0034 | 1. | 1. | 1.112 | 1.111 |
| NM | 2024-2025 | 471 | 0.0393 | 1.42e-14 | 1. | 1. | 0.8248 | 0.8216 |
| NV | 2024-2025 | 380 | 0.0293 | 1.42e-14 | 1. | 1. | 1.211 | 1.21 |
| OH | 2024-2025 | 1,901 | 0.0447 | 5.68e-14 | 1. | 1. | 1.701 | 1.7 |
| OK | 2018-2019 | 794 | 0.0449 | 0.0126 | 1. | 1. | 0.9729 | 0.971 |
| OR | 2024-2025 | 851 | 0.0411 | 1.42e-14 | 1. | 1. | 1.35 | 1.349 |
| PA | 2023-2024 | 1,822 | 0.0626 | 1.85e-13 | 1. | 1. | 0.7562 | 0.7544 |
| SC | 2023-2024 | 1,483 | 0.0575 | 0.0575 | 1. | 1. | 0.989 | 0.989 |
| TN | 2024-2025 | 1,319 | 0.0186 | 0.000171 | 1. | 1. | 1.109 | 1.109 |
| WA | 2022-2023 | 1,437 | 0.0335 | 5.68e-14 | 1. | 1. | 1.252 | 1.251 |
| WI | 2024-2025 | 1,320 | 3.91e-14 | 3.91e-14 | 1. | 1. | 2.27 | 2.27 |

\*All 26 rows; the complete table is also TableS9\_replication\_check.csv in the replication package in the GitHub repository<sup>14</sup>.

**Supplementary Table 3.** Reporting-quality exclusion sensitivity.

| threshold | n states | n schools | % convex |
| --- | --- | --- | --- |
| 20. | 26. | 34,773 | 75.927 |
| 30. | 28. | 36,031 | 75.771 |
| 40. | 28. | 36,031 | 75.771 |
| 50. | 29. | 36,259 | 75.793 |
| 60. | 31. | 37,172 | 75.699 |
| 101 | 33. | 38,486 | 75.632 |

**Supplementary Table 4.** Schools reporting exactly full coverage, by sector.

| state | sector | n | at 100 | % at 100 |
| --- | --- | --- | --- | --- |
| California 2022-23 | private | 911 | 0 | 0 |
| California 2022-23 | public | 5,691 | 0 | 0 |
| South Carolina 2024-25 | private | 278 | 47. | 16.906 |
| South Carolina 2024-25 | public | 1,211 | 26. | 2.147 |
| Texas 2023-24 | private | 582 | 236 | 40.55 |
| Texas 2023-24 | public | 1,091 | 146 | 13.382 |

**Supplementary Table 5.**
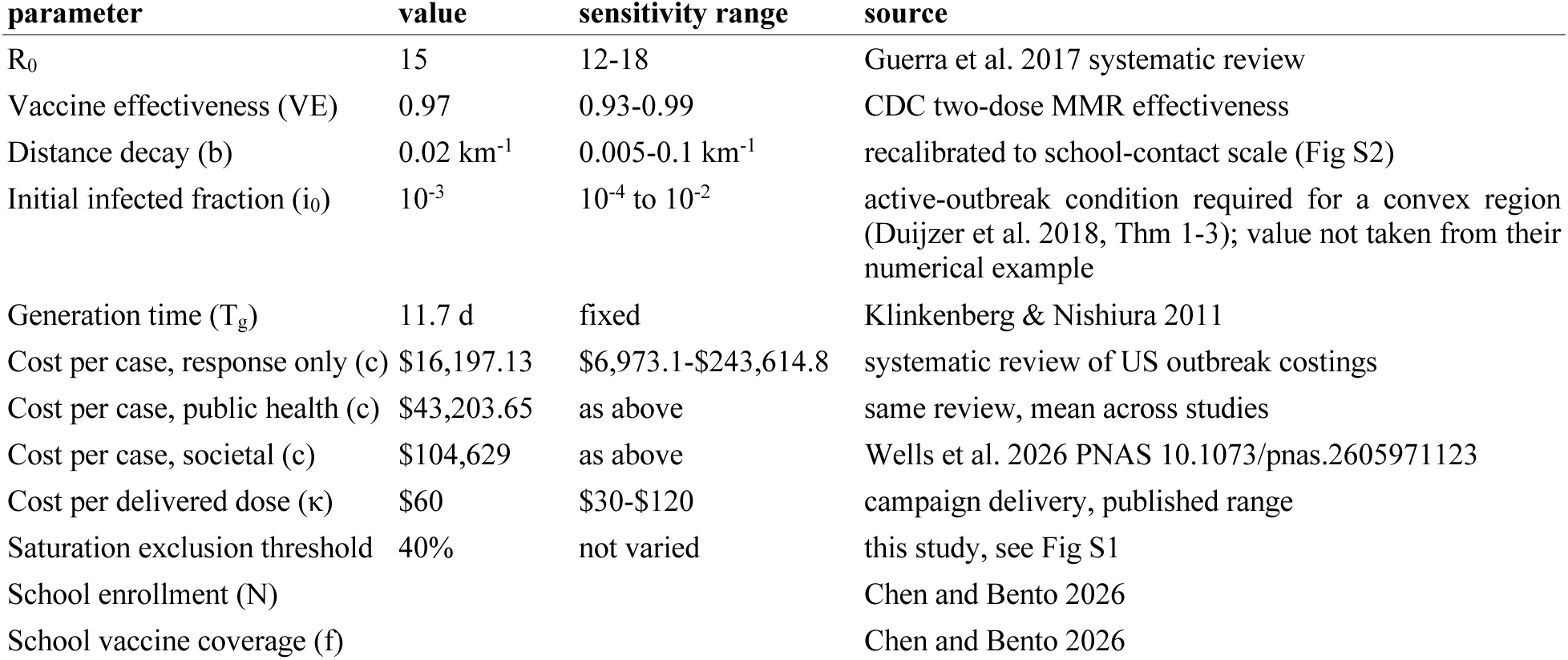
Model parameters and derived, design and unidentified quantities. Supplementary Table 5a. Model parameters and sensitivity ranges

**Supplementary Table 5b.**
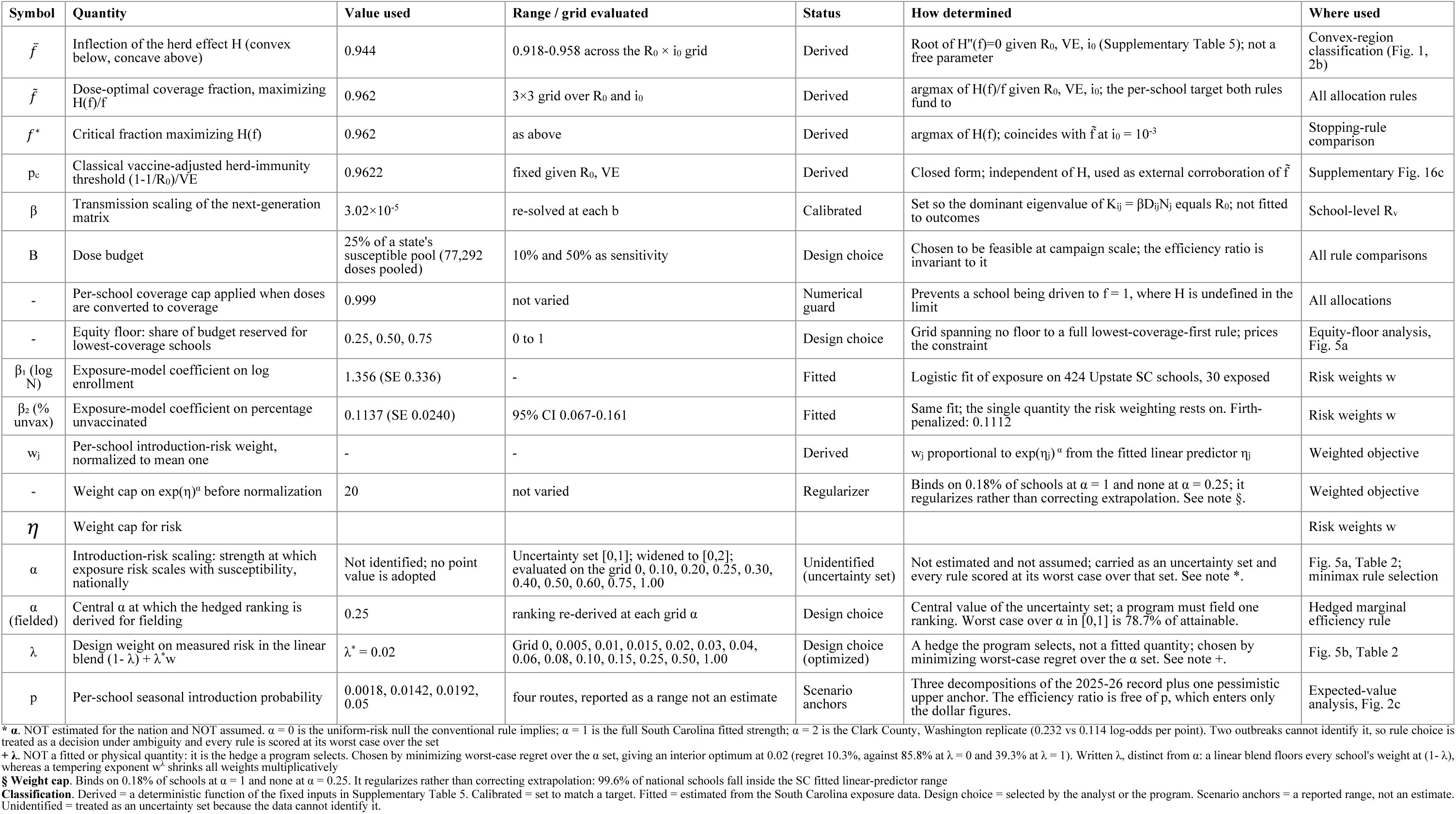
Derived, design and unidentified quantities.

**Supplementary Table 6.** Duijzer critical fractions across the parameter grid.

| $R_0$ | $i_0$ | $\bar{f}$ | $\tilde{f}$ | $f^*$ |
| --- | --- | --- | --- | --- |
| 12. | 0.0001 | 0.935 | 0.9445 | 0.945 |
| 12. | 0.001 | 0.923 | 0.944 | 0.945 |
| 12. | 0.01 | 0.8985 | 0.9405 | 0.9435 |
| 15. | 0.0001 | 0.9535 | 0.962 | 0.962 |
| 15. | 0.001 | 0.9435 | 0.9615 | 0.962 |
| 15. | 0.01 | 0.9225 | 0.959 | 0.961 |
| 18. | 0.0001 | 0.966 | 0.9735 | 0.9735 |
| 18. | 0.001 | 0.957 | 0.973 | 0.9735 |
| 18. | 0.01 | 0.939 | 0.9715 | 0.9725 |

**Supplementary Table 7.** Convex share under fixed and school-specific seeding.

| spec | % susceptibles | % schools |
| --- | --- | --- |
| fixed $i_0 = 1e^{-3}$ | 75.771 | 46.041 |
| school-specific $i_0 = 1/N_j$ | 68.544 | 33.005 |

**Supplementary Table 8.**
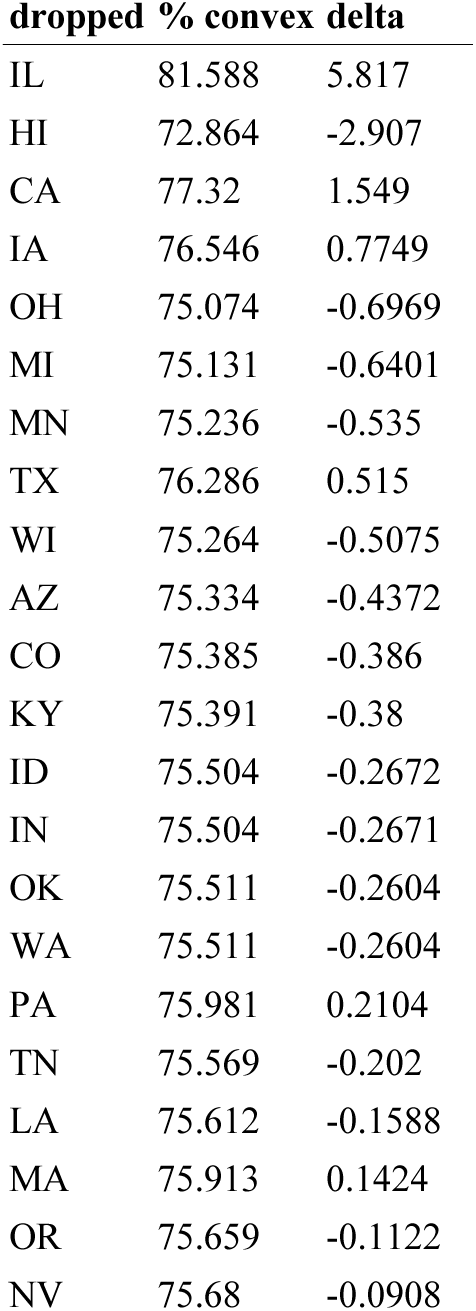
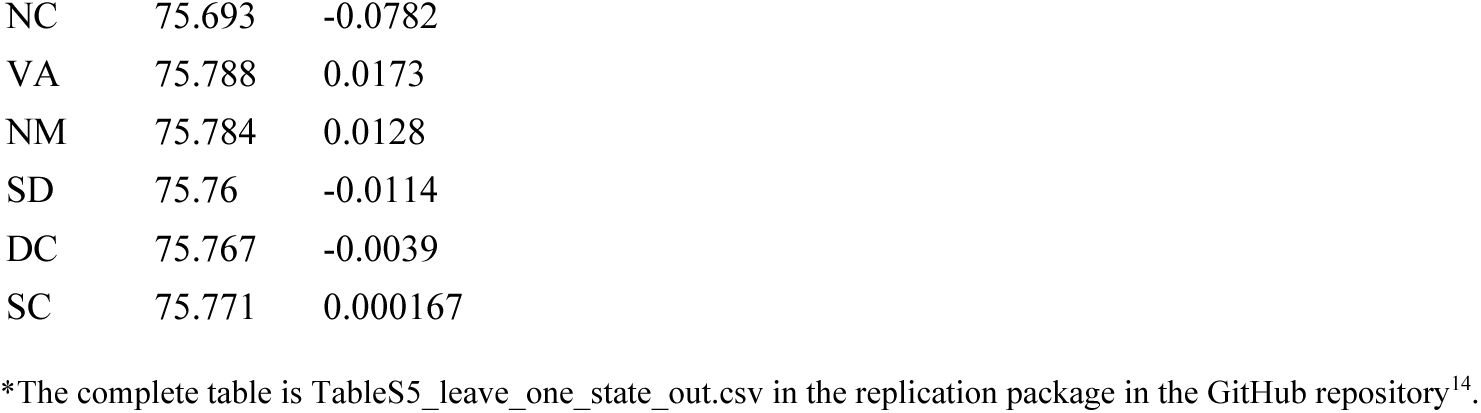
Leave-one-state-out on the convex share.

| dropped | % convex | delta |
| --- | --- | --- |
| IL | 81.588 | 5.817 |
| HI | 72.864 | -2.907 |
| CA | 77.32 | 1.549 |
| IA | 76.546 | 0.7749 |
| OH | 75.074 | -0.6969 |
| MI | 75.131 | -0.6401 |
| MN | 75.236 | -0.535 |
| TX | 76.286 | 0.515 |
| WI | 75.264 | -0.5075 |
| AZ | 75.334 | -0.4372 |
| CO | 75.385 | -0.386 |
| KY | 75.391 | -0.38 |
| ID | 75.504 | -0.2672 |
| IN | 75.504 | -0.2671 |
| OK | 75.511 | -0.2604 |
| WA | 75.511 | -0.2604 |
| PA | 75.981 | 0.2104 |
| TN | 75.569 | -0.202 |
| LA | 75.612 | -0.1588 |
| MA | 75.913 | 0.1424 |
| OR | 75.659 | -0.1122 |
| NV | 75.68 | -0.0908 |
| **dropped % convex delta** |  |  |
| NC | 75.693 | -0.0782 |
| VA | 75.788 | 0.0173 |
| NM | 75.784 | 0.0128 |
| SD | 75.76 | -0.0114 |
| DC | 75.767 | -0.0039 |
| SC | 75.771 | 0.000167 |

\*The complete table is TableS5\_leave\_one\_state\_out.csv in the replication package in the GitHub repository<sup>14</sup>.

**Supplementary Table 9.** Convex share across the *R*_0_ × *i*_0_ grid.

| $R_0$ | $i_0$ | $\bar{f}$ | $\tilde{f}$ | $f^*$ | % schools convex | % unvax convex |
| --- | --- | --- | --- | --- | --- | --- |
| 12. | 0.0001 | 0.935 | 0.9445 | 0.945 | 40.99 | 70.605 |
| 12. | 0.001 | 0.923 | 0.944 | 0.945 | 34.912 | 64.409 |
| 12. | 0.01 | 0.8985 | 0.9405 | 0.9435 | 25.592 | 53.977 |
| 15. | 0.0001 | 0.9535 | 0.962 | 0.962 | 55.375 | 82.247 |
| 15. | 0.001 | 0.9435 | 0.9615 | 0.962 | 45.944 | 75.674 |
| 15. | 0.01 | 0.9225 | 0.959 | 0.961 | 34.798 | 64.183 |
| 18. | 0.0001 | 0.966 | 0.9735 | 0.9735 | 63.82 | 88.979 |
| 18. | 0.001 | 0.957 | 0.973 | 0.9735 | 57.426 | 84.082 |
| 18. | 0.01 | 0.939 | 0.9715 | 0.9725 | 43.049 | 73.033 |

**Supplementary Table 10.**
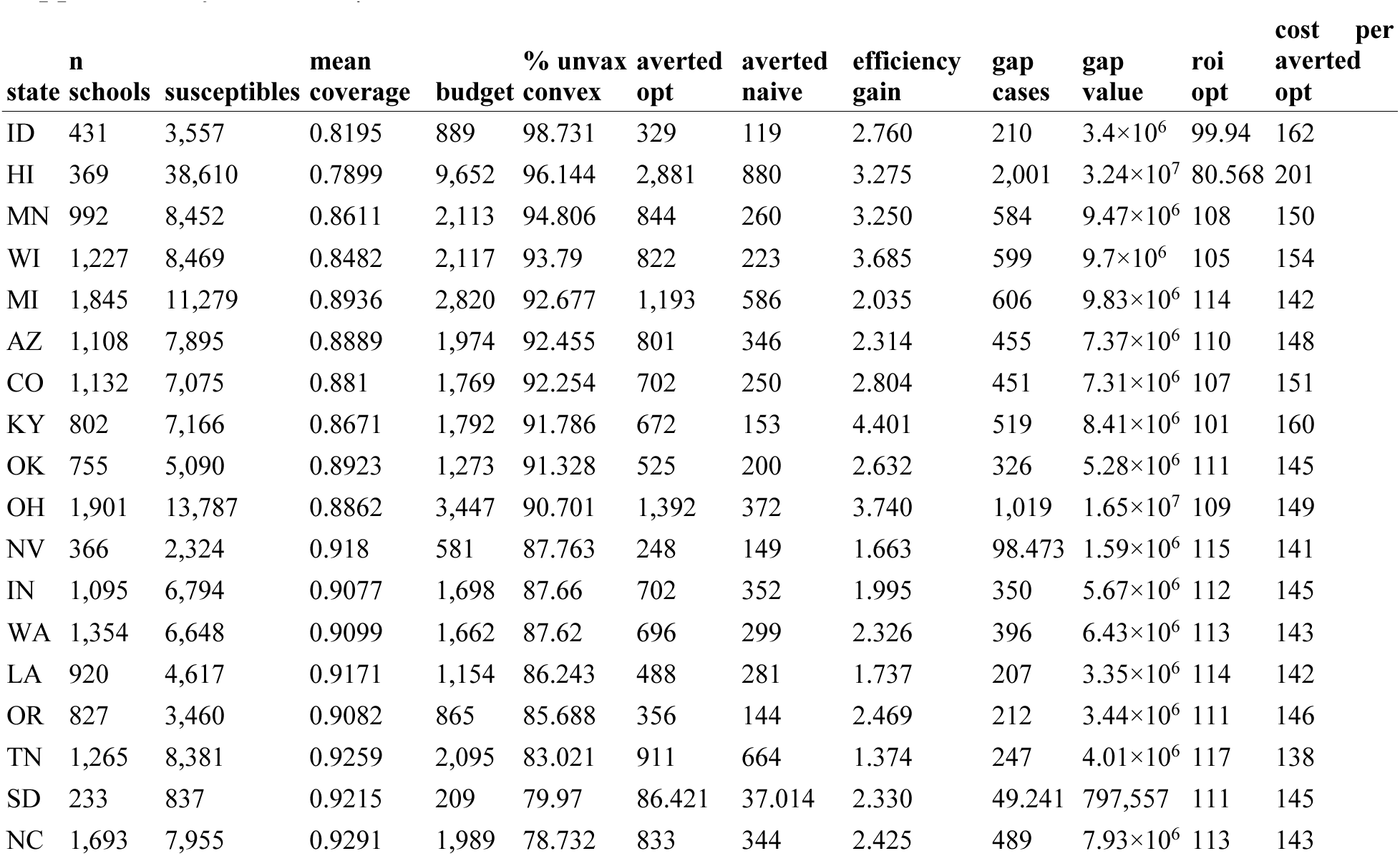
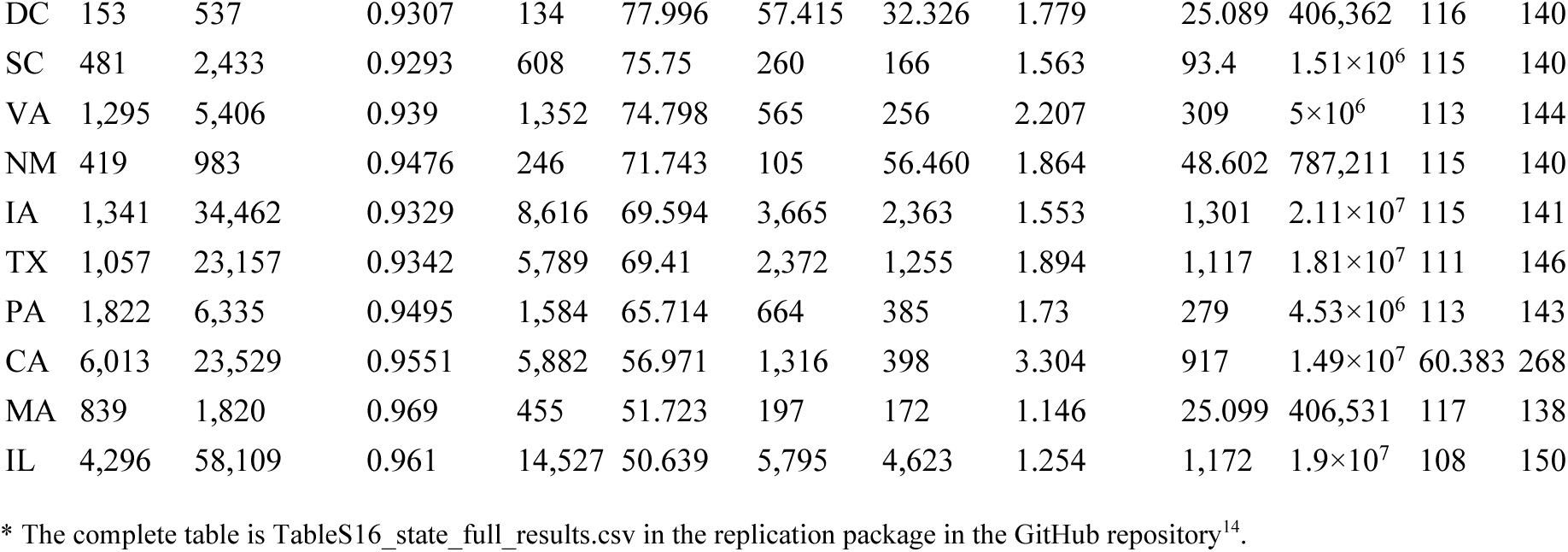
Per-state allocation results.

**Supplementary Table 11.** Per-school dose allocations.

| state | MMR | N | unvax | region | doses to $\tilde{f}$ |
| --- | --- | --- | --- | --- | --- |
| AZ | 0.724 | 170 | 46.92 | Convex | 42. |
| AZ | 0.813 | 209 | 39.083 | Convex | 33. |
| AZ | 0.339 | 59. | 38.999 | Convex | 38. |
| AZ | 0.818 | 209 | 38.038 | Convex | 32. |
| AZ | 0.57 | 86. | 36.98 | Convex | 35. |
| AZ | 0.58 | 88. | 36.96 | Convex | 35. |
| AZ | 0.55 | 80. | 36. | Convex | 34. |
| AZ | 0.077 | 39. | 35.997 | Convex | 36. |
| AZ | 0.5 | 70. | 35. | Convex | 34. |
| AZ | 0.752 | 137 | 33.976 | Convex | 30. |
| AZ | 0.783 | 152 | 32.984 | Convex | 29. |
| AZ | 0.776 | 147 | 32.928 | Convex | 29. |
| AZ | 0.846 | 208 | 32.032 | Convex | 25. |
| AZ | 0.529 | 68. | 32.028 | Convex | 31. |
| AZ | 0.686 | 102 | 32.028 | Convex | 30. |
| AZ | 0.754 | 126 | 30.996 | Convex | 28. |
| AZ | 0.4 | 50. | 30. | Convex | 29. |
| AZ | 0.6 | 75. | 30. | Convex | 28. |
| AZ | 0.762 | 126 | 29.988 | Convex | 26. |
| AZ | 0.804 | 153 | 29.988 | Convex | 25. |
| AZ | 0.707 | 99. | 29.007 | Convex | 27. |
| AZ | 0.592 | 71. | 28.968 | Convex | 28. |
| AZ | 0.826 | 161 | 28.014 | Convex | 23. |
| AZ | 0.783 | 129 | 27.993 | Convex | 24. |
| AZ | 0.81 | 147 | 27.93 | Convex | 24. |
| AZ | 0.816 | 147 | 27.048 | Convex | 23. |
| AZ | 0.571 | 63. | 27.027 | Convex | 26. |
| AZ | 0.738 | 103 | 26.986 | Convex | 24. |
| AZ | 0.62 | 71. | 26.98 | Convex | 26. |
| AZ | 0.645 | 76. | 26.98 | Convex | 25. |

\*First 30 of 1403 rows; the complete table is TableS17\_top\_schools\_by\_state.csv in the replication package in the GitHub repository<sup>14</sup>.

**Supplementary Table 12.**
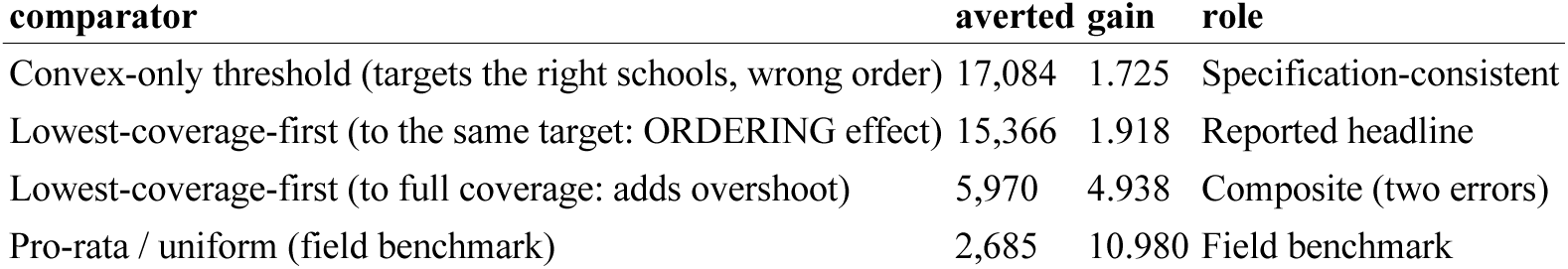
Comparator ladder at per-state budgets.

**Supplementary Table 13.**
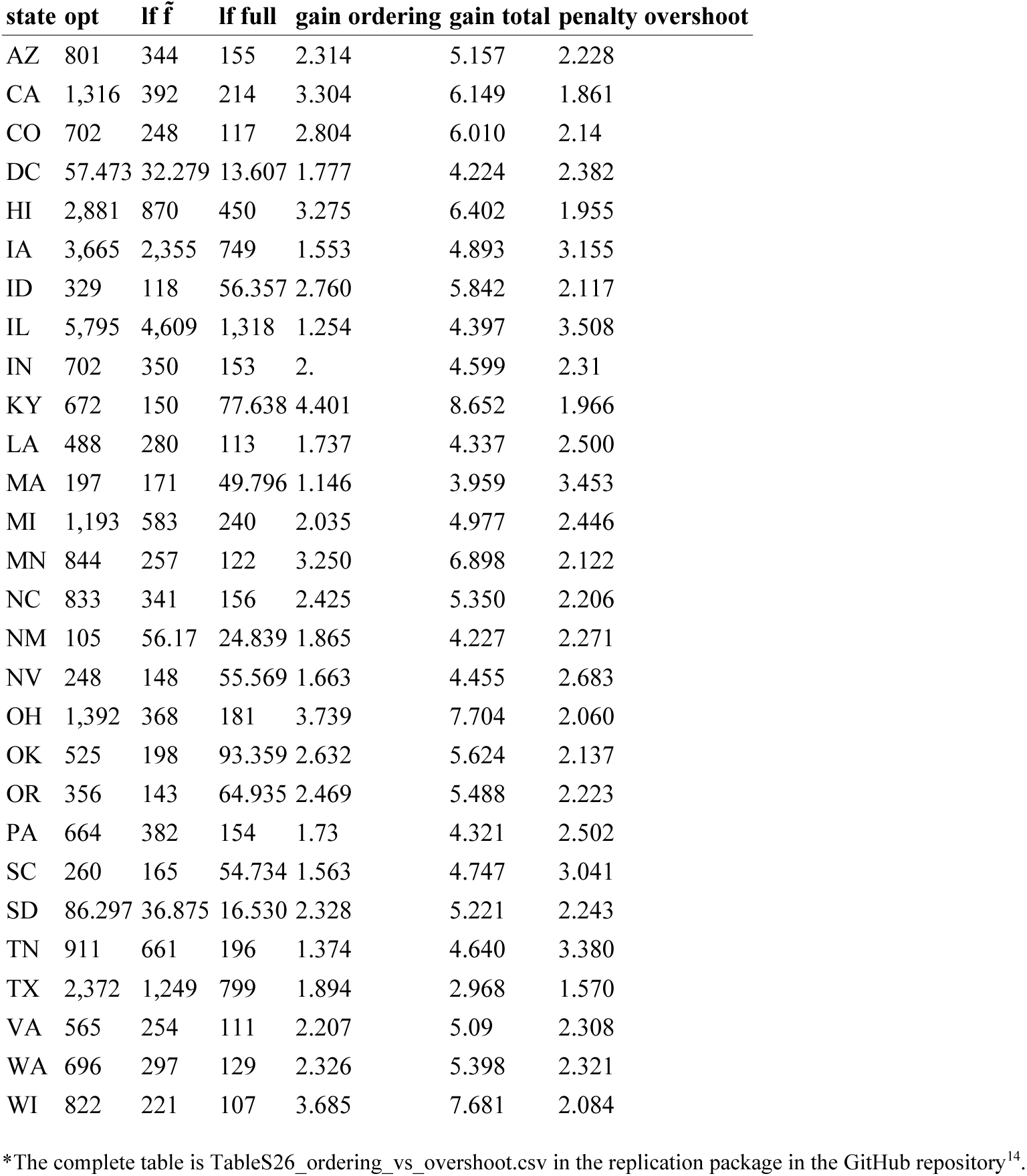
Ordering error versus stopping-rule error, per state.

| state | opt | If $\tilde{f}$ | If full | gain ordering | gain total | penalty overshoot |
| --- | --- | --- | --- | --- | --- | --- |
| AZ | 801 | 344 | 155 | 2.314 | 5.157 | 2.228 |
| CA | 1,316 | 392 | 214 | 3.304 | 6.149 | 1.861 |
| CO | 702 | 248 | 117 | 2.804 | 6.010 | 2.14 |
| DC | 57.473 | 32.279 | 13.607 | 1.777 | 4.224 | 2.382 |
| HI | 2,881 | 870 | 450 | 3.275 | 6.402 | 1.955 |
| IA | 3,665 | 2,355 | 749 | 1.553 | 4.893 | 3.155 |
| ID | 329 | 118 | 56.357 | 2.760 | 5.842 | 2.117 |
| IL | 5,795 | 4,609 | 1,318 | 1.254 | 4.397 | 3.508 |
| IN | 702 | 350 | 153 | 2. | 4.599 | 2.31 |
| KY | 672 | 150 | 77.638 | 4.401 | 8.652 | 1.966 |
| LA | 488 | 280 | 113 | 1.737 | 4.337 | 2.500 |
| MA | 197 | 171 | 49.796 | 1.146 | 3.959 | 3.453 |
| MI | 1,193 | 583 | 240 | 2.035 | 4.977 | 2.446 |
| MN | 844 | 257 | 122 | 3.250 | 6.898 | 2.122 |
| NC | 833 | 341 | 156 | 2.425 | 5.350 | 2.206 |
| NM | 105 | 56.17 | 24.839 | 1.865 | 4.227 | 2.271 |
| NV | 248 | 148 | 55.569 | 1.663 | 4.455 | 2.683 |
| OH | 1,392 | 368 | 181 | 3.739 | 7.704 | 2.060 |
| OK | 525 | 198 | 93.359 | 2.632 | 5.624 | 2.137 |
| OR | 356 | 143 | 64.935 | 2.469 | 5.488 | 2.223 |
| PA | 664 | 382 | 154 | 1.73 | 4.321 | 2.502 |
| SC | 260 | 165 | 54.734 | 1.563 | 4.747 | 3.041 |
| SD | 86.297 | 36.875 | 16.530 | 2.328 | 5.221 | 2.243 |
| TN | 911 | 661 | 196 | 1.374 | 4.640 | 3.380 |
| TX | 2,372 | 1,249 | 799 | 1.894 | 2.968 | 1.570 |
| VA | 565 | 254 | 111 | 2.207 | 5.09 | 2.308 |
| WA | 696 | 297 | 129 | 2.326 | 5.398 | 2.321 |
| WI | 822 | 221 | 107 | 3.685 | 7.681 | 2.084 |

\*The complete table is TableS26\_ordering\_vs\_overshoot.csv in the replication package in the GitHub repository<sup>14</sup>.

**Supplementary Table 14.** Allocation is independent of the risk model.

| state | full opt | nogeo opt | full naive | nogeo naive | abs diff opt | abs diff naive |
| --- | --- | --- | --- | --- | --- | --- |
| AZ | 801 | 801 | 346 | 346 | 0 | 0 |
| CA | 1,316 | 1,316 | 398 | 398 | 0 | 0 |
| CO | 702 | 702 | 250 | 250 | 0 | 0 |
| DC | 57.415 | 57.415 | 32.326 | 32.326 | 0 | 0 |
| HI | 2,881 | 2,881 | 880 | 880 | 0 | 0 |
| IA | 3,665 | 3,665 | 2,363 | 2,363 | 0 | 0 |
| ID | 329 | 329 | 119 | 119 | 0 | 0 |
| IL | 5,795 | 5,795 | 4,623 | 4,623 | 0 | 0 |
| IN | 702 | 702 | 352 | 352 | 0 | 0 |
| KY | 672 | 672 | 153 | 153 | 0 | 0 |
| LA | 488 | 488 | 281 | 281 | 0 | 0 |
| MA | 197 | 197 | 172 | 172 | 0 | 0 |
| MI | 1,193 | 1,193 | 586 | 586 | 0 | 0 |
| MN | 844 | 844 | 260 | 260 | 0 | 0 |
| NC | 833 | 833 | 344 | 344 | 0 | 0 |
| NM | 105 | 105 | 56.460 | 56.460 | 0 | 0 |
| NV | 248 | 248 | 149 | 149 | 0 | 0 |
| OH | 1,392 | 1,392 | 372 | 372 | 0 | 0 |
| OK | 525 | 525 | 200 | 200 | 0 | 0 |
| OR | 356 | 356 | 144 | 144 | 0 | 0 |
| PA | 664 | 664 | 385 | 385 | 0 | 0 |
| SC | 260 | 260 | 166 | 166 | 0 | 0 |
| SD | 86.421 | 86.421 | 37.014 | 37.014 | 0 | 0 |
| TN | 911 | 911 | 664 | 664 | 0 | 0 |
| TX | 2,372 | 2,372 | 1,255 | 1,255 | 0 | 0 |
| VA | 565 | 565 | 256 | 256 | 0 | 0 |
| WA | 696 | 696 | 299 | 299 | 0 | 0 |
| WI | 822 | 822 | 223 | 223 | 0 | 0 |

\*The complete table is TableS18\_riskmodel\_independence.csv in the replication package in the GitHub repository<sup>14</sup>.

**Supplementary Table 15.**
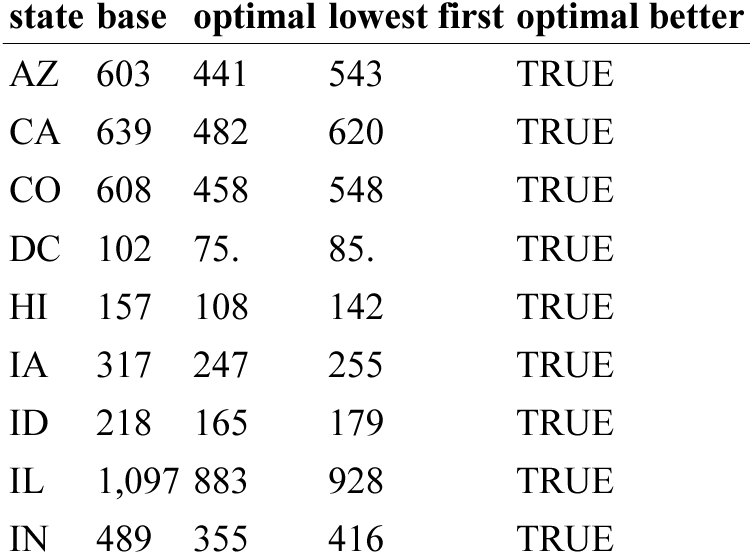
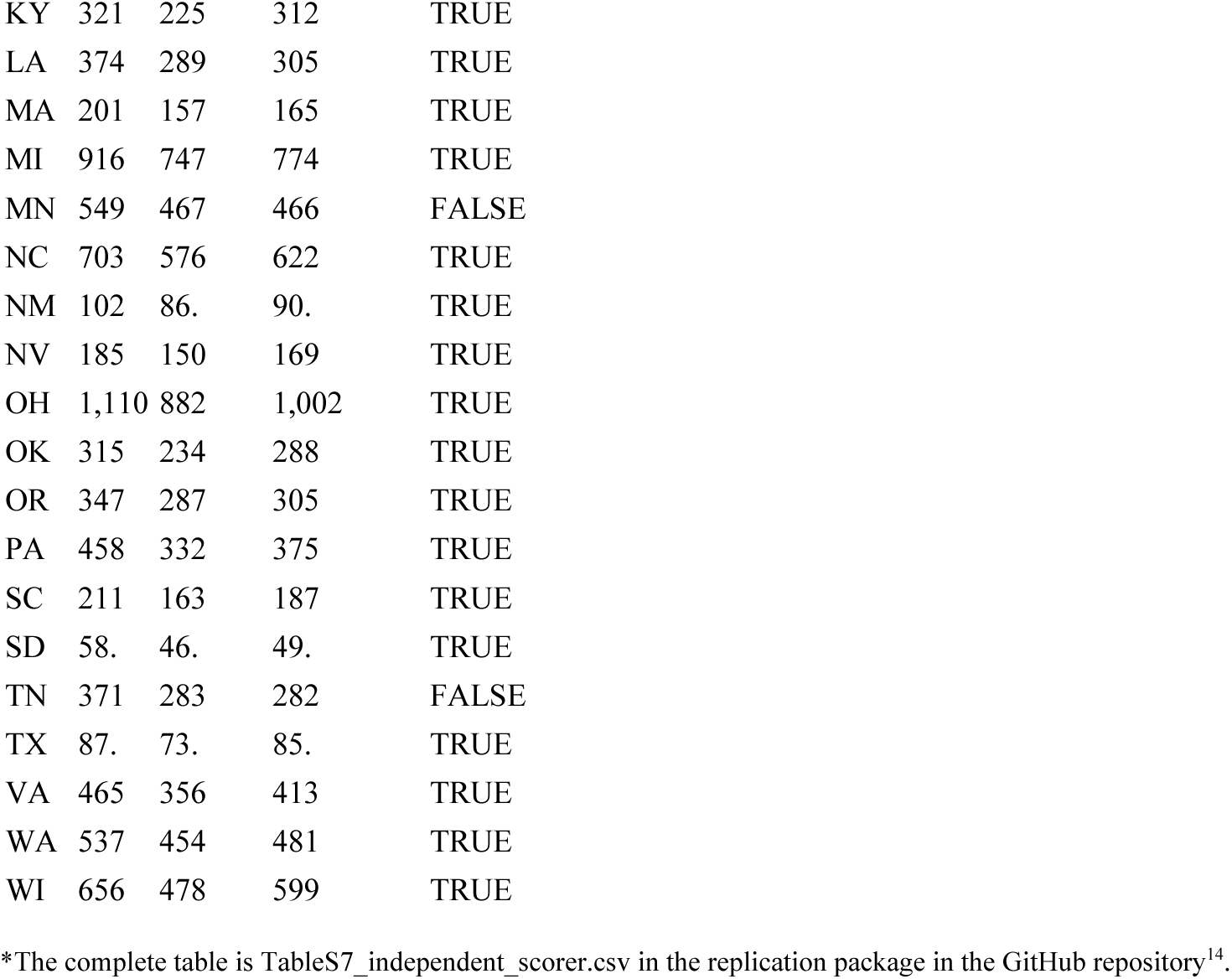
Independent scorer: schools left above *R_v_* = 1.

**Supplementary Table 16.** Allocation rule comparison at a pooled national budget.

| rule | doses | doses to concave schools | herd benefit |
| --- | --- | --- | --- |
| Dose-optimal (Duijzer) | 77,291 | 2,846 | 30,939 |
| Convex-only threshold rule | 77,273 | 0 | 15,875 |
| Lowest-coverage-first | 77,292 | 0 | 10,898 |

**Supplementary Table 17.** Supercritical schools remaining under each rule.

| budget<br>frac | base | opt | naive | enrol<br>base | enrol<br>opt | enrol<br>naive | cleared<br>opt | cleared<br>naive | extra<br>cleared | %<br>opt | %<br>naive | children<br>spared |
| --- | --- | --- | --- | --- | --- | --- | --- | --- | --- | --- | --- | --- |
| 0.1 | 12,196 | 11,010 | 11,693 | $1.97 \times 10^6$ | $1.84 \times 10^6$ | $1.93 \times 10^6$ | 1,186 | 503 | 683 | 9.724 | 4.124 | 89,708 |
| 0.25 | 12,196 | 9,498 | 10,685 | $1.97 \times 10^6$ | $1.69 \times 10^6$ | $1.85 \times 10^6$ | 2,698 | 1,511 | 1,187 | 22.122 | 12.389 | 153,554.5 |
| 0.5 | 12,196 | 7,709 | 8,545 | $1.97 \times 10^6$ | $1.56 \times 10^6$ | $1.64 \times 10^6$ | 4,487 | 3,651 | 836 | 36.791 | 29.936 | 85,518 |

**Supplementary Table 18.**
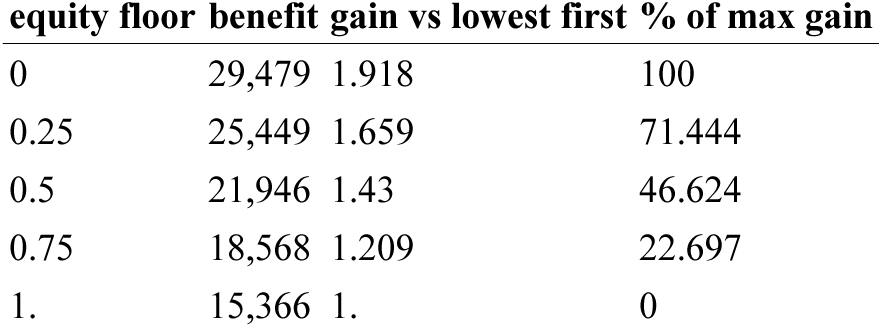
Equity-floor sensitivity.

**Supplementary Table 19.** Indirect and total objectives compared.

| rule | indirect only | total prevented | direct component | gain indirect | gain total |
| --- | --- | --- | --- | --- | --- |
| Dose-optimal | 29,479 | 101,982 | 72,503 | 1.918 | 1.161 |
| Equity floor 50% | 23,889 | 96,392 | 72,503 | 1.557 | 1.097 |
| Lowest-coverage-first | 15,366 | 87,869 | 72,503 | 1. | 1. |

**Supplementary Table 20.** Per-school introduction probability, two decompositions.

| route | factor 1 | factor 2 | p |
| --- | --- | --- | --- |
| A-low: SC attack × introductions per US county | 0.0684 | 0.0258 | 0.0018 |
| A-high: SC attack × introductions per county in affected states | 0.0684 | 0.2072 | 0.0142 |
| B: SC statewide attack × fraction of states affected | 0.0204 | 0.94 | 0.0192 |
| C: pessimistic upper anchor |  |  | 0.05 |

**Supplementary Table 21.**
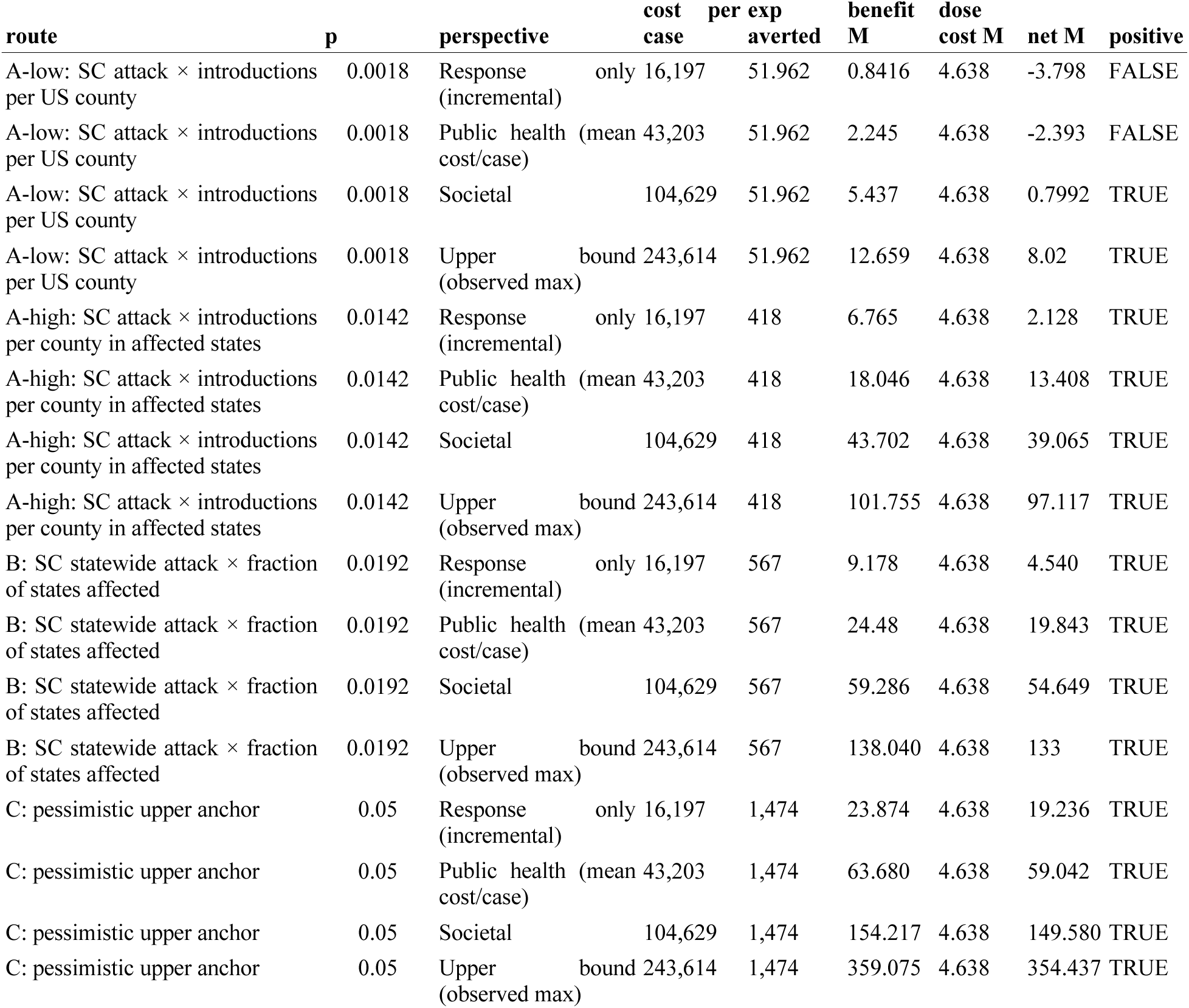
Expected net benefit by probability and perspective.

**Supplementary Table 22.**
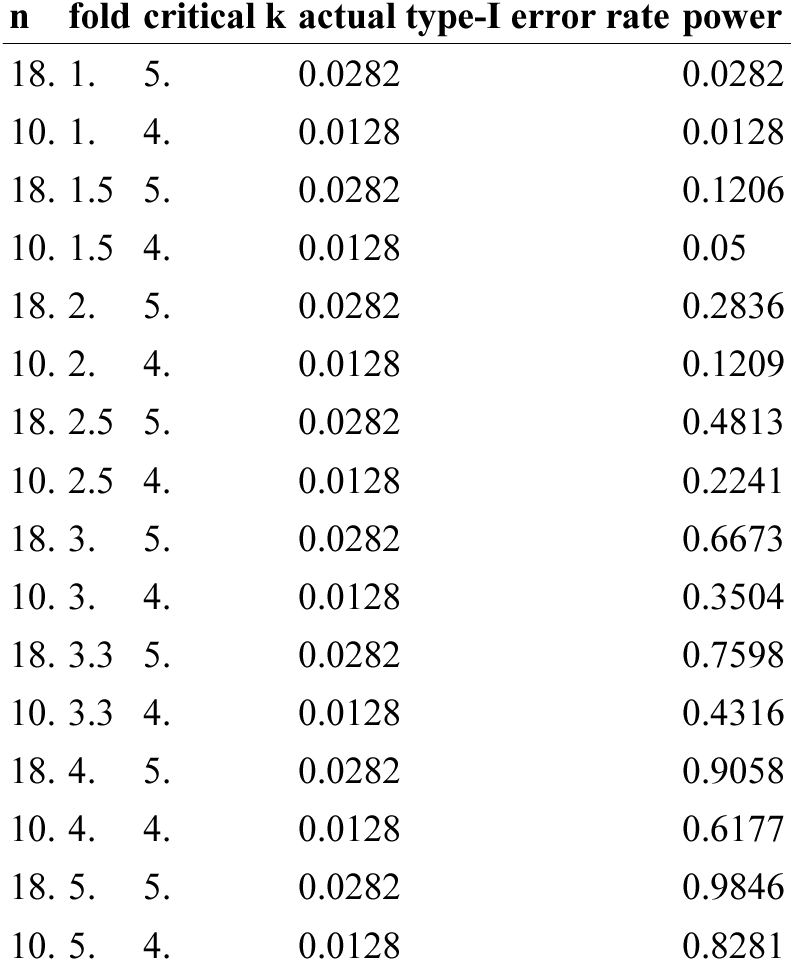
Power of the enrichment design.

**Supplementary Table 23.** Exact intervals on observed enrichment.

| stratum | n amplifying | k top decile | enrichment | CI lo fold | CI hi fold | power vs 2fold |
| --- | --- | --- | --- | --- | --- | --- |
| All counties | 18. | 3. | 1.667 | 0.3579 | 4.142 | 0.2836 |
| Records consistent | 10. | 3. | 3. | 0.6674 | 6.525 | 0.1209 |

**Supplementary Table 24.**
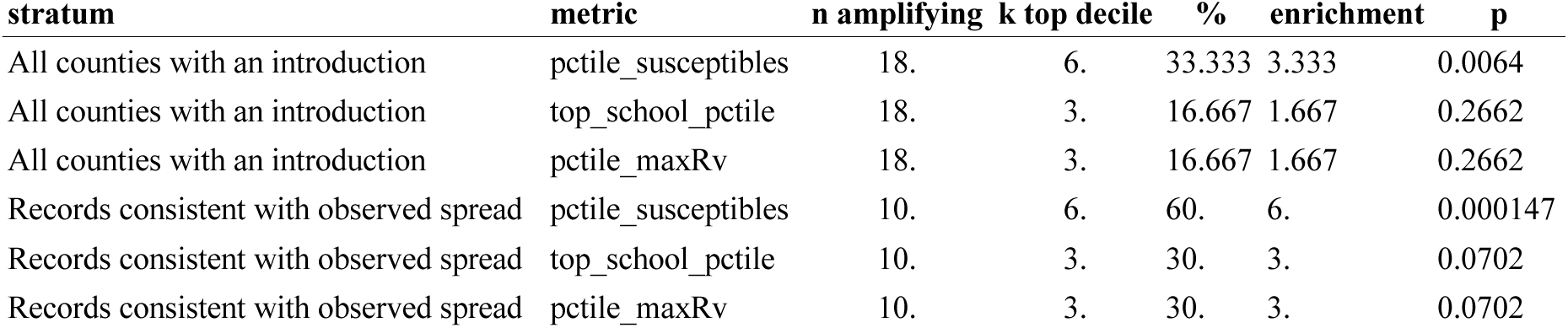
Enrichment stratified by data richness.

**Supplementary Table 25.**
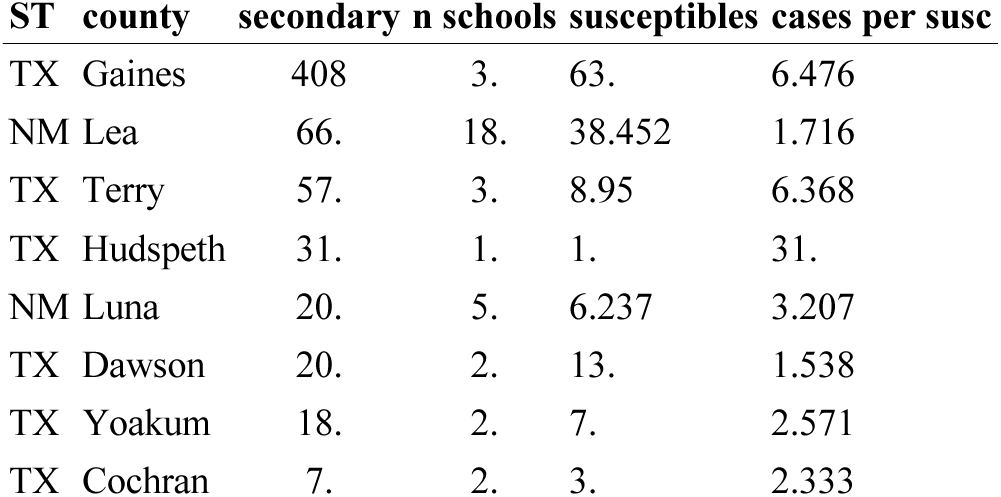
Counties whose records cannot account for observed cases.

**Supplementary Table 26.** Enrichment across richness thresholds.

| max cps | min schools | n counties | n amplifying | enrichment | p |
| --- | --- | --- | --- | --- | --- |
| 0.25 | 0 | 66. | 7. | 4.286 | 0.0257 |
| 0.5 | 0 | 70. | 9. | 3.333 | 0.053 |
| 1. | 0 | 73. | 10. | 3. | 0.0702 |
| 2. | 0 | 75. | 12. | 2.5 | 0.1109 |
| Inf | 0 | 81. | 18. | 1.667 | 0.2662 |
| 0.25 | 10. | 30. | 4. | 5. | 0.0523 |
| 0.5 | 10. | 31. | 5. | 4. | 0.0815 |
| 1. | 10. | 31. | 5. | 4. | 0.0815 |
| 2. | 10. | 32. | 6. | 3.333 | 0.1143 |
| Inf | 10. | 32. | 6. | 3.333 | 0.1143 |
| 0.25 | 20. | 22. | 2. | 5. | 0.19 |
| 0.5 | 20. | 22. | 2. | 5. | 0.19 |
| 1. | 20. | 22. | 2. | 5. | 0.19 |
| 2. | 20. | 22. | 2. | 5. | 0.19 |
| Inf | 20. | 22. | 2. | 5. | 0.19 |

**Supplementary Table 27.** South Carolina data-quality audit.

| quantity | value |
| --- | --- |
| schools in file | 1,564 |
| geocoded | 1,562 |
| coverage 2024-25 present | 1,516 |
| coverage 2025-26 present | 1,512 |
| exposed schools | 31. |
| exposed outside Upstate | 0 |
| at exactly 100% (2024-25) | 75. |
| at exactly 100% (2025-26) | 52. |
| distinct coverage values (2025-26) | 339 |

**Supplementary Table 28.**
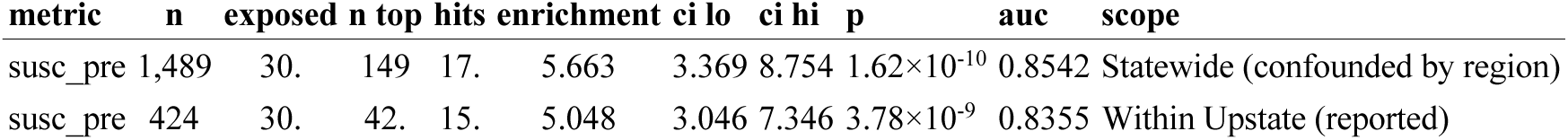
Statewide versus within-region enrichment.

**Supplementary Table 29.** South Carolina validation restricted to kindergarten-containing schools.

| quantity | value | ci lo | ci hi | p | auc | n | n exposed |
| --- | --- | --- | --- | --- | --- | --- | --- |
| All grades (reported) \| Susceptible headcount | 5.048 | 3.046 | 7.346 | $3.78 \times 10^{-9}$ | 0.836 | 424 | 30. |
| Kindergarten-containing only \| Susceptible headcount | 4.719 | 2.339 | 7.64 | $1.27 \times 10^{-5}$ | 0.809 | 269 | 19. |
| All grades (reported) \| Enrollment (negative control) | 1.01 | 0.212 | 2.754 | 0.59 | 0.625 | 424 | 30. |
| Kindergarten-containing only \| Enrollment (negative control) | 0.524 | 0.013 | 2.686 | 0.876 | 0.661 | 269 | 19. |
| Upstate schools, all grades | 424 |  |  |  |  |  |  |
| Upstate schools with a kindergarten grade | 269 |  |  |  |  |  |  |
| Exposed schools, all grades | 30. |  |  |  |  |  |  |
| Exposed schools with a kindergarten grade | 19. |  |  |  |  |  |  |
| Median enrollment, all grades | 511 |  |  |  |  |  |  |
| Median enrollment, kindergarten-containing | 454 |  |  |  |  |  |  |
| Power of the restricted design at the all-grades effect size | 0.85 |  |  |  |  |  |  |

**Supplementary Table 30.** South Carolina logistic model, coverage beyond enrollment.

| term | estimate | se | z | p |
| --- | --- | --- | --- | --- |
| (Intercept) | -12.309 | 2.385 | -5.162 | $2.44 \times 10^{-7}$ |
| log(N) | 1.356 | 0.336 | 4.035 | $5.46 \times 10^{-5}$ |
| I(100 * (1 - MMR)) | 0.1137 | 0.024 | 4.746 | $2.08 \times 10^{-6}$ |
| LRT: adding % unvaccinated to log(enrollment) | 26.671 | | | $2.41 \times 10^{-7}$ |

**Supplementary Table 31.** Flag-completeness bound on top-decile enrichment.

| extra missed | total exposed | non-detection % | enrichment | lo | hi | p |
| --- | --- | --- | --- | --- | --- | --- |
| 0 | 30. | 0 | 5.048 | 3.046 | 7.346 | $8.69 \times 10^{-8}$ |
| 1. | 31. | 3.226 | 4.885 | 2.948 | 7.109 | $1.33 \times 10^{-7}$ |
| 2. | 32. | 6.25 | 4.732 | 2.855 | 6.887 | $2.01 \times 10^{-7}$ |
| 3. | 33. | 9.091 | 4.589 | 2.769 | 6.678 | $3 \times 10^{-7}$ |
| 4. | 34. | 11.765 | 4.454 | 2.688 | 6.481 | $4.4 \times 10^{-7}$ |
| 5. | 35. | 14.286 | 4.327 | 2.611 | 6.296 | $6.38 \times 10^{-7}$ |
| 6. | 36. | 16.667 | 4.206 | 2.538 | 6.121 | $9.13 \times 10^{-7}$ |
| 7. | 37. | 18.919 | 4.093 | 2.47 | 5.956 | $1.29 \times 10^{-6}$ |
| 8. | 38. | 21.053 | 3.985 | 2.405 | 5.799 | $1.81 \times 10^{-6}$ |
| 9. | 39. | 23.077 | 3.883 | 2.343 | 5.651 | $2.5 \times 10^{-6}$ |
| 10. | 40. | 25. | 3.786 | 2.284 | 5.509 | $3.43 \times 10^{-6}$ |
| 11. | 41. | 26.829 | 3.693 | 2.229 | 5.375 | $4.65 \times 10^{-6}$ |
| 12. | 42. | 28.571 | 3.605 | 2.176 | 5.247 | $6.26 \times 10^{-6}$ |
| 13. | 43. | 30.233 | 3.522 | 2.125 | 5.125 | $8.35 \times 10^{-6}$ |
| 14. | 44. | 31.818 | 3.442 | 2.077 | 5.008 | $1.1 \times 10^{-5}$ |
| 15. | 45. | 33.333 | 3.365 | 2.031 | 4.897 | $1.45 \times 10^{-5}$ |
| 16. | 46. | 34.783 | 3.292 | 1.986 | 4.791 | $1.89 \times 10^{-5}$ |
| 17. | 47. | 36.17 | 3.222 | 1.944 | 4.689 | 2.44×10<sup>-5</sup> |
| 18. | 48. | 37.5 | 3.155 | 1.904 | 4.591 | 3.13×10<sup>-5</sup> |
| 19. | 49. | 38.776 | 3.09 | 1.865 | 4.497 | 4×10<sup>-5</sup> |
| 20. | 50. | 40. | 3.029 | 1.828 | 4.407 | 5.07×10<sup>-5</sup> |
| 21. | 51. | 41.176 | 2.969 | 1.792 | 4.321 | 6.38×10<sup>-5</sup> |
| 22. | 52. | 42.308 | 2.912 | 1.757 | 4.238 | 8×10<sup>-5</sup> |
| 23. | 53. | 43.396 | 2.857 | 1.724 | 4.158 | 9.96×10<sup>-5</sup> |
| 24. | 54. | 44.444 | 2.804 | 1.692 | 4.081 | 0.000123 |
| 25. | 55. | 45.455 | 2.753 | 1.661 | 4.007 | 0.000152 |
| 26. | 56. | 46.429 | 2.704 | 1.632 | 3.935 | 0.000186 |
| 27. | 57. | 47.368 | 2.657 | 1.603 | 3.866 | 0.000227 |
| 28. | 58. | 48.276 | 2.611 | 1.575 | 3.799 | 0.000276 |
| 29. | 59. | 49.153 | 2.567 | 1.549 | 3.735 | 0.000333 |

\*First 30 of 91 rows; the complete table is TableS40\_flag\_completeness\_bound.csv in the replication package in the GitHub repository<sup>14</sup>.

**Supplementary Table 32.** Fitted coverage and concentration trends by state.

| state | n years | cov slope | cov lo | cov hi | cov p | top5 slope | top5 lo | top5 hi | top5 p |
| --- | --- | --- | --- | --- | --- | --- | --- | --- | --- |
| CA | 7. | -0.2138 | -0.4231 | -0.0044 | 0.0468 | 3.524 | 2.962 | 4.087 | 1.69×10<sup>-5</sup> |
| CO | 8. | -0.1866 | -0.5343 | 0.1611 | 0.2371 | 0.5502 | 0.108 | 0.9923 | 0.0227 |
| MN | 12. | -0.7157 | -0.9023 | -0.5291 | 6.57×10<sup>-6</sup> | 0.5246 | -1.316 | 2.365 | 0.5397 |
| KY | 7. | -0.9416 | -1.361 | -0.5219 | 0.0022 | 0.4712 | -1.202 | 2.144 | 0.5015 |
| IL | 11. | -0.2164 | -0.2867 | -0.1461 | 6.57×10<sup>-5</sup> | 0.1835 | -0.1872 | 0.5541 | 0.2919 |
| LA | 4. | -0.1582 | -0.2736 | -0.0427 | 0.0276 | 0.1427 | -2.072 | 2.358 | 0.8077 |
| VA | 11. | -0.4594 | -0.8901 | -0.0287 | 0.0391 | 0.0076 | -0.1957 | 0.2109 | 0.9344 |
| WA | 7. | -0.3269 | -0.9801 | 0.3264 | 0.2547 | -0.0789 | -0.774 | 0.6162 | 0.7822 |
| IA | 8. | -0.3453 | -0.6042 | -0.0864 | 0.0172 | -0.0913 | -0.6128 | 0.4303 | 0.6835 |
| TX | 6. | -0.6046 | -0.925 | -0.2843 | 0.0063 | -0.2823 | -2.199 | 1.634 | 0.7035 |
| AZ | 10. | -0.6563 | -0.7789 | -0.5337 | 1.73×10<sup>-6</sup> | -0.4366 | -0.6737 | -0.1994 | 0.0028 |
| OK | 7. | -0.6738 | -0.9267 | -0.4209 | 0.001 | -0.6335 | -3.838 | 2.571 | 0.6329 |
| ME | 7. | 0.9848 | 0.6936 | 1.276 | 0.000333 | -0.7346 | -2.255 | 0.7859 | 0.2693 |
| SD | 9. | -0.5172 | -0.8147 | -0.2198 | 0.0045 | -1.318 | -1.956 | -0.6795 | 0.0018 |
| HI | 10. | -1.958 | -3.24 | -0.6766 | 0.0078 | -1.381 | -2.241 | -0.5218 | 0.006 |
| MA | 12. | 0.0353 | -0.0415 | 0.1121 | 0.3295 | -1.686 | -2.644 | -0.7291 | 0.0028 |
| NC | 5. | -0.5598 | -1.003 | -0.1163 | 0.0277 | -2.318 | -4.402 | -0.233 | 0.0384 |
| ND | 4. | -0.3963 | -0.8115 | 0.019 | 0.0545 | -2.418 | -6.138 | 1.301 | 0.1075 |

**Supplementary Table 33.** South Carolina concentration over two years.

| year | n schools | mean cov | susceptibles | top5 share |
| --- | --- | --- | --- | --- |
| 2024-25 | 1,440 | 0.9356 | 52,925 | 25.038 |
| 2025-26 | 1,440 | 0.9366 | 50,895 | 24.372 |

**Supplementary Table 34.** Exposed-school allocation by coverage band, Upstate South Carolina.

| band | n schools | n exposed | susceptibles | doses optimal | doses lcf |
| --- | --- | --- | --- | --- | --- |
| <75% | 14. | 1. | 1,023 | 0 | 973 |
| 75-85% | 40. | 13. | 2,807 | 0 | 2,319 |
| 85-94.4% | 187 | 12. | 9,825 | 4,510 | 1,219 |
| 94.4-96.2% | 98. | 4. | 3,267 | 0 | 0 |
| >96.2% | 85. | 0 | 1,120 | 0 | 0 |

**Supplementary Table 35.**
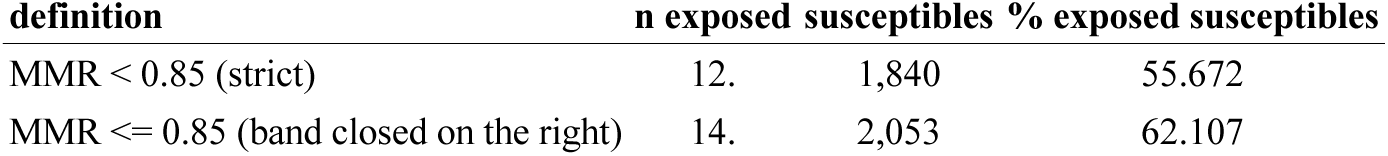
Reconciliation of the two 85% cut points.

**Supplementary Table 36.**
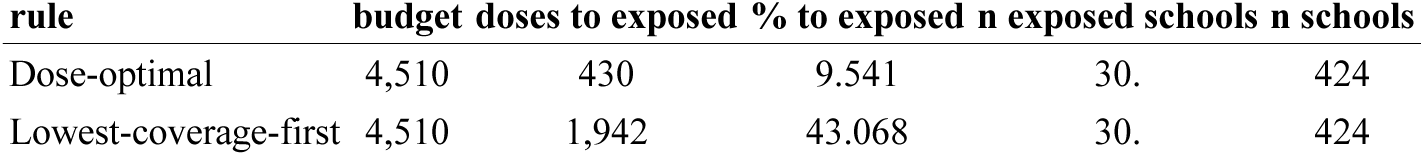
Doses reaching later-exposed schools, Upstate South Carolina.

**Supplementary Table 37.**
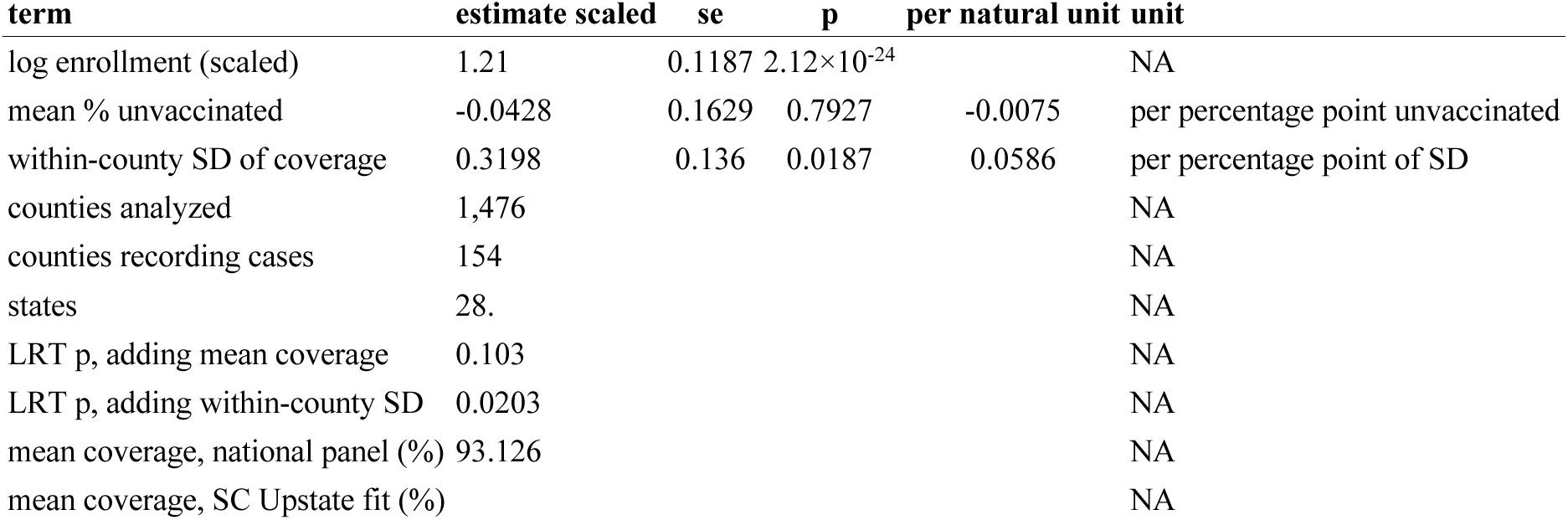
Between-county model of which counties were reached.

**Supplementary Table 38.** Independent replicates of the exposure-risk scaling.

| quantity | n | n exposed | beta % unvaccinated | se | p | lrt 1df | beta log enrollment |
| --- | --- | --- | --- | --- | --- | --- | --- |
| South Carolina, Upstate 2024-25 \| kindergarten-through-12 school rate | 424 | 30. | 0.1137 | 0.024 | $2.08 \times 10^{-6}$ | 26.67 | 1.356 |
| Washington, Clark County 2018-19 \| 1 - MMR exemption rate (only MMR status published at K-12) | 131 | 12. | 0.2315 | 0.0738 | 0.0017 | 14.94 | 0.7367 |
| Washington, Clark County 2018-19 \| all-vaccine Complete (sensitivity) | 131 | 12. | 0.0134 | 0.0163 | 0.41 | 0.62 | 0.1483 |
| Colorado 2024-25 \| MMR Fully Immunized | 126 | 3. | 0.0555 | 0.1118 | 0.62 | 0.2 | 1.379 |
| South Carolina, Upstate 2024-25 \| susceptible headcount | 424 | 30. | 5.048 | 3.046 | $3.78 \times 10^{-9}$ | 7.346 | 0.836 |
| South Carolina, Upstate 2024-25 \| enrollment (negative control) | 424 | 30. | 1.01 | 0.212 | 0.59 | 2.754 | 0.625 |
| Washington, Clark County 2018-19 \| susceptible headcount | 131 | 12. | 2.519 | 0.55 | 0.0994 | 5.875 | 0.755 |
| Washington, Clark County 2018-19 \| enrollment (negative control) | 131 | 12. | 0.84 | 0.021 | 0.731 | 3.933 | 0.499 |
| Colorado 2024-25 \| susceptible headcount | 126 | 3. | 3.231 | 0.082 | 0.281 | 15.132 | 0.753 |
| Utah 2025-26 \| NOT USABLE |  | 20. |  |  |  |  |  |
| New York, Rockland 2018-19 \| NOT USABLE |  | 7. |  |  |  |  |  |
| Washington, Ridgefield 2024-26 \| NOT USABLE |  | 1. |  |  |  |  |  |
| Colorado 2024-25 \| NOT USABLE |  | 3. |  |  |  |  |  |

**Supplementary Table 39.** Risk-weighted allocation gain across tempering exponents.

| alpha | max weight | gain unweighted rule | gain rederived rule | % of attainable |
| --- | --- | --- | --- | --- |
| 0 | 1. | 1.918 | 1.918 | 100 |
| 0.1 | 2.415 | 1.611 | 1.63 | 98.52 |
| 0.2 | 5.706 | 1.328 | 1.455 | 91.191 |
| 0.25 | 8.687 | 1.2 | 1.398 | 85.518 |
| 0.35 | 19.708 | 0.9574 | 1.37 | 70.108 |
| 0.5 | 63.058 | 0.6653 | 1.386 | 48.066 |
| 0.7 | 250.963 | 0.4023 | 1.431 | 28.137 |
| 0.85 | 593 | 0.2802 | 1.47 | 19.072 |
| 1. | 1,173 | 0.200 | 1.504 | 13.327 |

**Supplementary Table 40.**
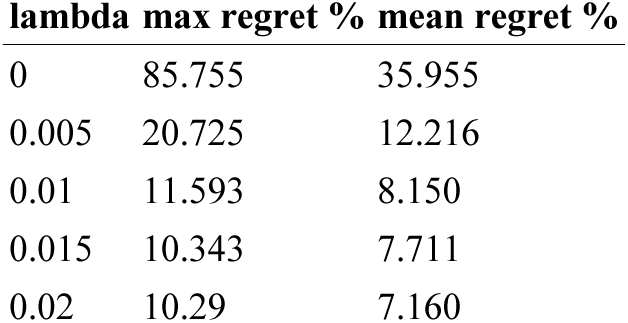
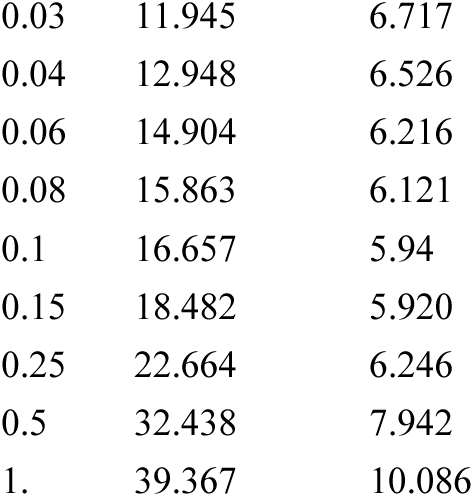
Worst-case regret by design weight.

| lambda | max regret % | mean regret % |
| --- | --- | --- |
| 0 | 85.755 | 35.955 |
| 0.005 | 20.725 | 12.216 |
| 0.01 | 11.593 | 8.150 |
| 0.015 | 10.343 | 7.711 |
| 0.02 | 10.29 | 7.160 |
| **lambda max regret % mean regret %** |  |  |
| 0.03 | 11.945 | 6.717 |
| 0.04 | 12.948 | 6.526 |
| 0.06 | 14.904 | 6.216 |
| 0.08 | 15.863 | 6.121 |
| 0.1 | 16.657 | 5.94 |
| 0.15 | 18.482 | 5.920 |
| 0.25 | 22.664 | 6.246 |
| 0.5 | 32.438 | 7.942 |
| 1. | 39.367 | 10.086 |

**Supplementary Table 41.** Regret ranking of all candidate rules.

| rule | max regret % |
| --- | --- |
| Two-percent hedge (minimax-regret blend, lambda = 0.02) | 10.29 |
| Hedged marginal efficiency | 21.349 |
| Equity floor 75% | 28.755 |
| Equity floor 50% | 31.355 |
| Equity floor 25% | 47.09 |
| Lowest-coverage-first | 47.873 |
| Dose-optimal (unweighted) | 85.755 |

**Supplementary Table 42.** Sensitivity of the rule ranking to the width of the uncertainty set.

| rule | worst over 0-1 | worst over 0-2 | alpha at worst 0-2 | regret 0-1 | regret 0-2 | rank 0-1 | rank 0-2 | rank changed |
| --- | --- | --- | --- | --- | --- | --- | --- | --- |
| Two-percent hedge | 89.706 | 89.706 | 0 | 10.294 | 10.294 | 1. | 1. | FALSE |
| Hedged marginal efficiency | 78.582 | 77.126 | 1.2 | 21.349 | 22.746 | 2. | 2. | FALSE |
| Equity floor 75% | 71.176 | 71.176 | 0 | 28.755 | 28.755 | 3. | 3. | FALSE |
| Equity floor 50% | 68.645 | 59.99 | 2. | 31.355 | 40.01 | 4. | 4. | FALSE |
| Lowest-coverage-first | 52.127 | 52.127 | 0 | 47.873 | 47.873 | 6. | 5. | TRUE |
| Equity floor 25% | 52.91 | 41.987 | 2. | 47.09 | 58.013 | 5. | 6. | TRUE |
| Dose-optimal (unweighted) | 14.206 | 10.633 | 1.5 | 85.755 | 89.339 | 7. | 7. | FALSE |

**Supplementary Table 43.** Minimax worst case by dose budget.

| rule | budget 0.1 | budget 0.25 | budget 0.5 |
| --- | --- | --- | --- |
| Dose-optimal (unweighted) | 9.993 | 14.206 | 71.196 |
| Equity floor 25% | 39.207 | 52.91 | 82.485 |
| Equity floor 50% | 53.133 | 68.645 | 88.66 |
| Equity floor 75% | 57.791 | 71.176 | 90.858 |
| Hedged marginal efficiency | 78.949 | 78.582 | 91.838 |
| Lowest-coverage-first | 32.23 | 52.127 | 81.621 |
| Two-percent hedge | 80.428 | 89.706 | 96.904 |

**Supplementary Table 44.**
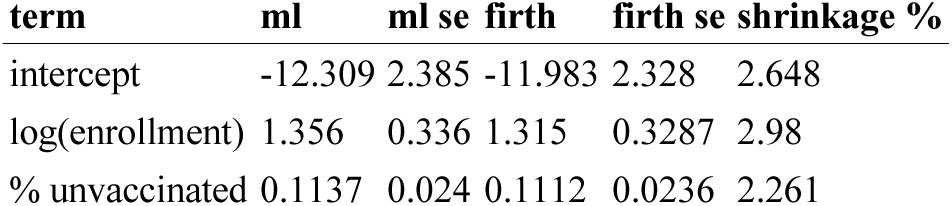
Firth-penalized versus maximum-likelihood risk model.

| term | ml | ml se | firth | firth se | shrinkage % |
| --- | --- | --- | --- | --- | --- |
| intercept | -12.309 | 2.385 | -11.983 | 2.328 | 2.648 |
| log(enrollment) | 1.356 | 0.336 | 1.315 | 0.3287 | 2.98 |
| % unvaccinated | 0.1137 | 0.024 | 0.1112 | 0.0236 | 2.261 |

**Supplementary Table 45.** Rule ranking under three structurally different uncertainty sets.

| set | rule | worst % | worst regret | at scenario |
| --- | --- | --- | --- | --- |
| A: coefficient CI | Lowest-coverage-first | 79.77 | 20.15 | beta = 0.067 |
| A: coefficient CI | Equity floor 75% | 75.24 | 24.76 | beta = 0.161 |
| A: coefficient CI | Hedged marginal efficiency | 73.525 | 26.544 | beta = 0.067 |
| A: coefficient CI | Equity floor 50% | 58.451 | 41.549 | beta = 0.161 |
| A: coefficient CI | Two-percent hedge | 54.972 | 45.028 | beta = 0.161 |
| A: coefficient CI | Equity floor 25% | 43.343 | 56.657 | beta = 0.161 |
| A: coefficient CI | Dose-optimal (unweighted) | 12.131 | 87.826 | beta = 0.114 |
| B: weight functional form | Two-percent hedge | 87.466 | 12.534 | exponential |
| B: weight functional form | Hedged marginal efficiency | 85.314 | 14.686 | exponential |
| B: weight functional form | Equity floor 75% | 71.326 | 28.605 | step |
| B: weight functional form | Equity floor 50% | 61.835 | 38.165 | exponential |
| B: weight functional form | Lowest-coverage-first | 52.173 | 47.827 | step |
| B: weight functional form | Equity floor 25% | 44.502 | 55.498 | exponential |
| B: weight functional form | Dose-optimal (unweighted) | 12.131 | 87.826 | exponential |
| C: covariate set | Lowest-coverage-first | 99.700 | 0.30 | coverage only |
| C: covariate set | Equity floor 75% | 77.615 | 22.385 | coverage only |
| C: covariate set | Equity floor 50% | 64.487 | 35.513 | coverage only |
| C: covariate set | Equity floor 25% | 52.76 | 47.24 | coverage only |
| C: covariate set | Two-percent hedge | 42.331 | 57.669 | coverage only |
| C: covariate set | Hedged marginal efficiency | 34.861 | 65.139 | coverage only |
| C: covariate set | Dose-optimal (unweighted) | 34.11 | 65.89 | coverage only |

**Supplementary Table 46.** Fitted support and implied weight spread.

| quantity | value |
| --- | --- |
| SC linear predictor, min | -8.832 |
| SC linear predictor, max | 5.813 |
| national linear predictor, min | -9.187 |
| national linear predictor, max | 3.374 |
| national schools outside SC support (%) | 0.4413 |
| susceptibles outside SC coverage support (%) | 1.383 |
| relative risk spread within the SC fitted sample (fold) | $2.29 \times 10^6$ |
| relative risk spread across the national panel (fold) | 285,443 |
| same, at the lower 95% bound on the slope | 45,932 |
| same, at the upper 95% bound on the slope | $2.16 \times 10^6$ |

**Supplementary Table 47.**
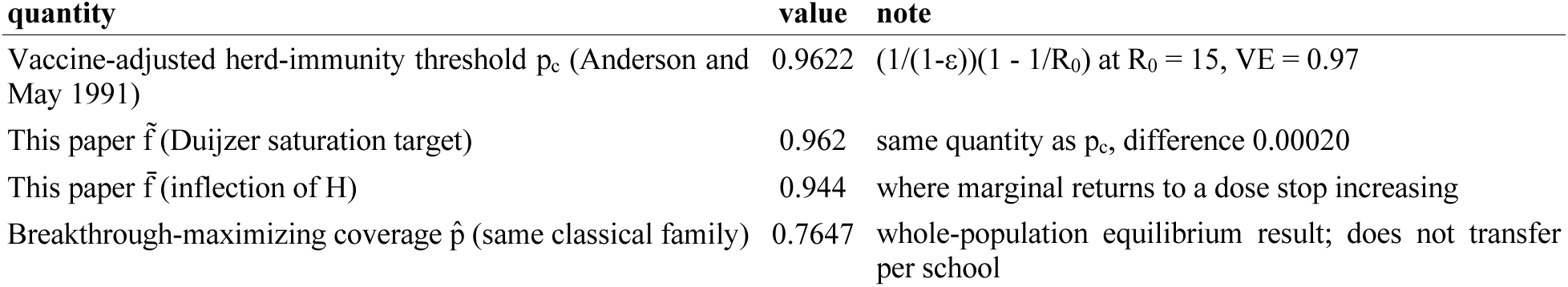
Threshold correspondence with the classical vaccine-adjusted herd-immunity result.

**Supplementary Table 48.** Assortativity by state.

| state | state coverage | $\tilde{\phi}$ (assortativity) | n schools | also, in Harris et al.<sup>10</sup> |
| --- | --- | --- | --- | --- |
| CA | 0.9551 | 0.5069 | 6,013 | TRUE |
| KY | 0.8671 | 0.4895 | 802 | FALSE |
| HI | 0.7899 | 0.4714 | 369 | TRUE |
| NM | 0.9476 | 0.4428 | 419 | TRUE |
| MA | 0.969 | 0.428 | 839 | FALSE |
| VA | 0.939 | 0.405 | 1,295 | FALSE |
| OH | 0.8862 | 0.3912 | 1,901 | FALSE |
| NC | 0.9291 | 0.3907 | 1,693 | FALSE |
| PA | 0.9495 | 0.3863 | 1,822 | FALSE |
| CO | 0.881 | 0.3683 | 1,132 | FALSE |
| OR | 0.9082 | 0.3662 | 827 | FALSE |
| IL | 0.961 | 0.3607 | 4,296 | FALSE |
| WA | 0.9099 | 0.3515 | 1,354 | TRUE |
| OK | 0.8923 | 0.3493 | 755 | TRUE |
| IN | 0.9077 | 0.349 | 1,095 | FALSE |
| SD | 0.9215 | 0.3478 | 233 | FALSE |
| AZ | 0.8889 | 0.3422 | 1,108 | TRUE |
| WI | 0.8482 | 0.3412 | 1,227 | FALSE |
| MN | 0.8611 | 0.336 | 992 | TRUE |
| DC | 0.9307 | 0.3332 | 153 | FALSE |
| LA | 0.9171 | 0.3278 | 920 | FALSE |
| NV | 0.918 | 0.3226 | 366 | FALSE |
| TX | 0.9342 | 0.3125 | 1,057 | TRUE |
| ID | 0.8195 | 0.289 | 431 | FALSE |
| MI | 0.8936 | 0.2681 | 1,845 | TRUE |
| SC | 0.9293 | 0.264 | 481 | TRUE |
| IA | 0.9329 | 0.2582 | 1,341 | TRUE |
| TN | 0.9259 | 0.2374 | 1,265 | FALSE |

\*The complete table is TableS42\_assortativity\_by\_state.csv in the replication package in the GitHub repository<sup>14</sup>.

**Supplementary Table 49.**
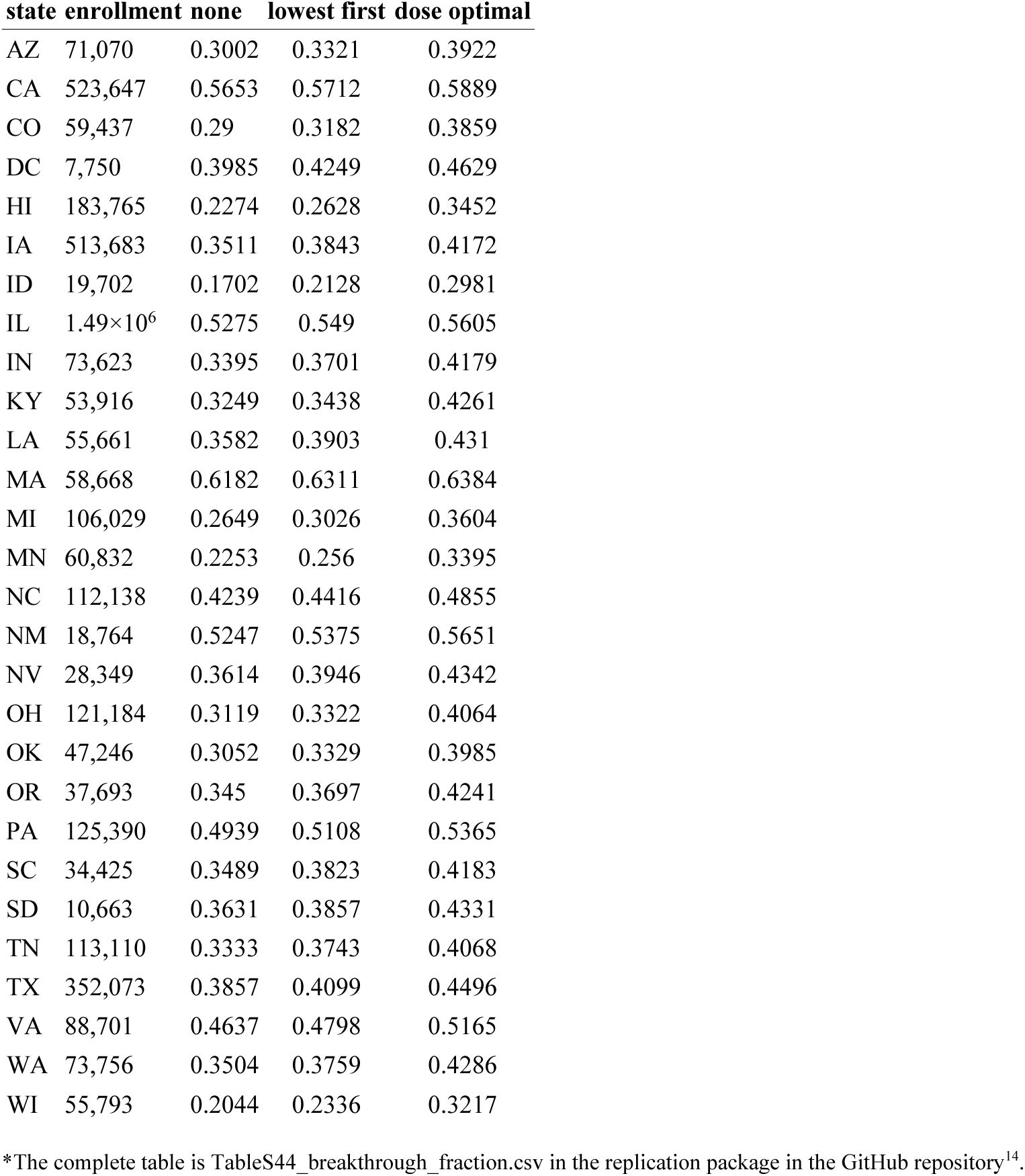
Breakthrough fraction by allocation rule.

**Supplementary Table 50.** Pooled breakthrough fraction by allocation rule.

| quantity | breakthrough fraction | basis |
| --- | --- | --- |
| Baseline, no allocation | 0.4341 | enrollment-weighted mean over the 28 state rows in the per-state table |
| Lowest-coverage-first | 0.4574 | enrollment-weighted mean over the 28 state rows in the per-state table |
| Dose-optimal (unweighted) | 0.4899 | enrollment-weighted mean over the 28 state rows in the per-state table |

**Supplementary Table 51.**
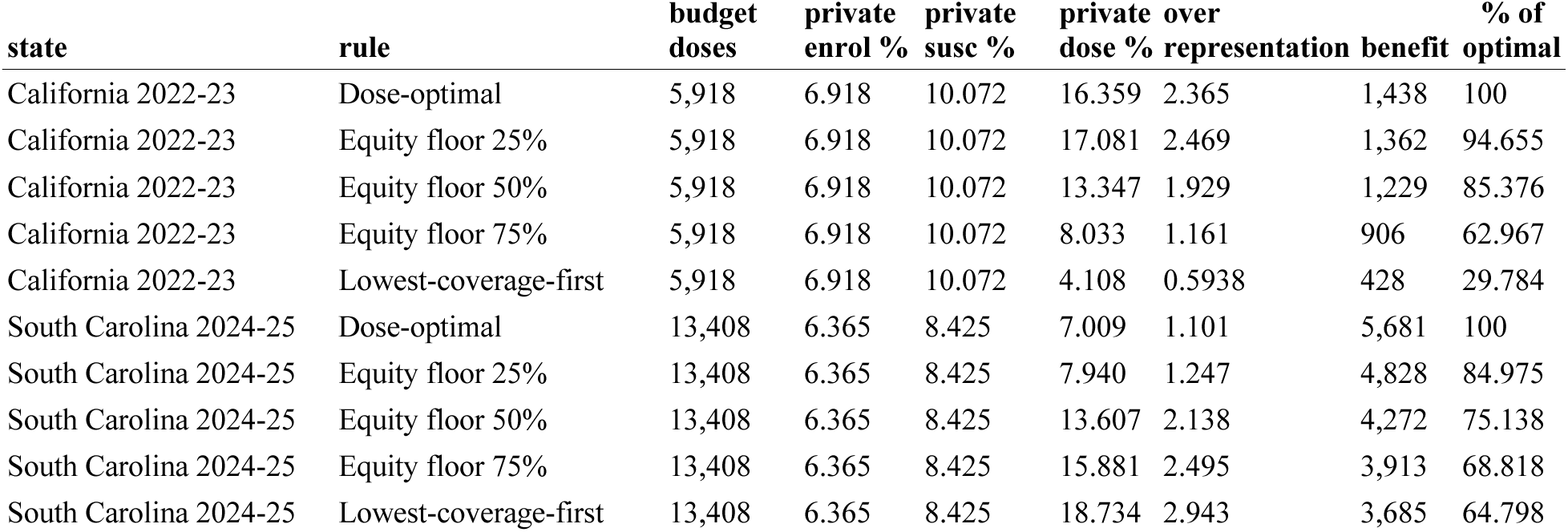
Private-school dose share by allocation rule.

**Supplementary Table 52.**
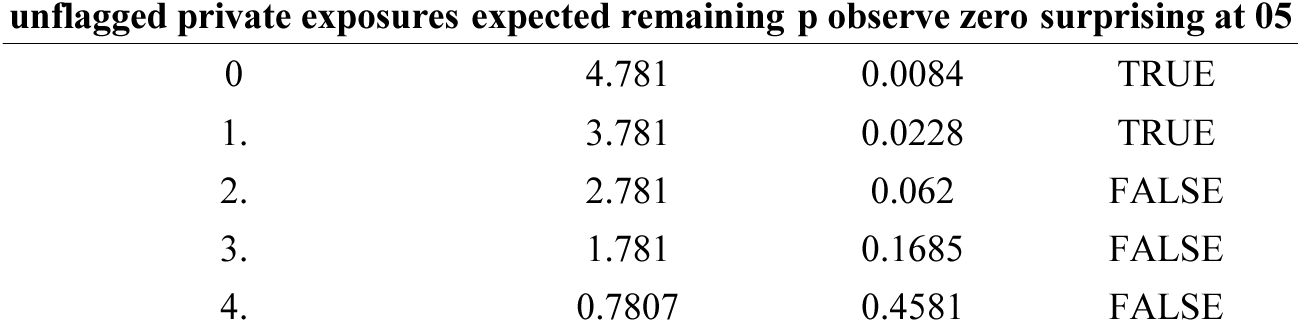
Private-school exposure under assumed non-detection.

**Supplementary Table 53.**
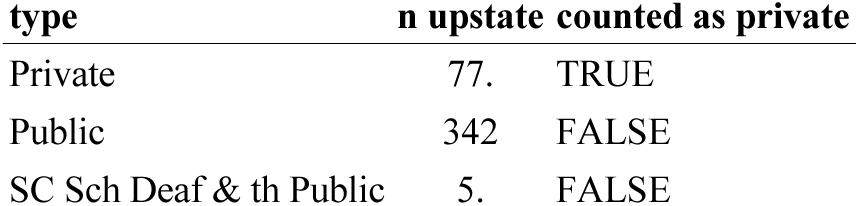
South Carolina school counts by reported sector.

**Supplementary Table 54.** Coverage distribution by school sector.

| state | sector | n | mean | median | sd | p05 | p25 | % below 85 |
| --- | --- | --- | --- | --- | --- | --- | --- | --- |
| California 2022-23 | private | 911 | 92.789 | 95. | 10.682 | 76.5 | 95. | 6.586 |
| California 2022-23 | public | 5,691 | 95.884 | 98. | 8.511 | 90. | 95. | 3.128 |
| South Carolina 2024-25 | private | 278 | 88.532 | 92. | 13.415 | 60. | 84. | 25.54 |
| South Carolina 2024-25 | public | 1,211 | 93.723 | 95. | 5.885 | 84. | 92. | 5.12 |
| Texas 2023-24 | private | 582 | 89.972 | 95.83 | 14.065 | 58.842 | 85.71 | 23.883 |
| Texas 2023-24 | public | 1,091 | 93.359 | 95.02 | 7.161 | 81.14 | 91.67 | 8.983 |

**Supplementary Table 55.** Sector differences in coverage variance and tail.

| state | sd ratio | var p | median gap pp | median p | below85 OR | below85 p |
| --- | --- | --- | --- | --- | --- | --- |
| California 2022-23 | 1.255 | 0 | -3. | $5.71 \times 10^{-176}$ | 2.183 | $1.9 \times 10^{-6}$ |
| South Carolina 2024-25 | 2.28 | 0 | -3. | $1.58 \times 10^{-5}$ | 6.345 | $1.57 \times 10^{-21}$ |
| Texas 2023-24 | 1.964 | 0 | 0.81 | 0.1506 | 3.177 | $5.44 \times 10^{-16}$ |

**Supplementary Table 56.** California private-school share by coverage band.

| band | n private | n public | susc private | susc public | n schools | % private schools | % private susceptibles |
| --- | --- | --- | --- | --- | --- | --- | --- |
| <75% | 39. | 119 | 667 | 8,876 | 158 | 24.684 | 6.996 |
| 75-85% | 21. | 59. | 157 | 926 | 80. | 26.25 | 14.459 |
| 85-94.4% | 109 | 354 | 420 | 2,149 | 463 | 23.542 | 16.353 |
| 94.4-96.2% | 579 | 1,085 | 914 | 2,694 | 1,664 | 34.796 | 25.349 |
| 96.2%+ | 163 | 4,074 | 225 | 6,642 | 4,237 | 3.847 | 3.275 |

**Supplementary Table 57.** California sector position tests.

| quantity | value |
| --- | --- |
| overall private share of schools (%) | 13.799 |
| overall private share of susceptibles (%) | 10.072 |
| private share of schools, <85% (%) | 25.21 |
| private share of schools, targeted band (%) | 23.542 |
| private share of susceptibles, <85% (%) | 7.756 |
| private share of susceptibles, targeted band (%) | 16.353 |
| Fisher OR, private <85% vs targeted band | 1.095 |
| Fisher p | 0.6417 |
| coverage ceiling in this release (censored) | 0.99 |

**Supplementary Table 58.** California persistence of clustering across SB277.

| quantity | spearman rho | p value | value |
| --- | --- | --- | --- |
| Within-county SD, unweighted | 0.313 | 0.044 |  |
| Within-county SD, enrollment-weighted | 0.4103 | 0.0074 |  |
| County mean coverage | 0.5368 | 0.000306 |  |
| Matched schools |  |  | 5,393 |
| Counties with $\geq 10$ matched | | | 42. |
| Schools at exactly 100% in 2014-15 |  |  | 859 |
| Schools at exactly 100% in 2022-23 |  |  | 0 |
| Mean coverage change (pp) |  |  | 3.973 |
| Mean change with 2014-15 capped at 99 (pp) |  |  | 4.133 |
| Censoring contribution (pp) |  |  | 0.1593 |

**Supplementary Table 59.** Permutation test on the SB277 breakpoint.

| outcome | observed level | n breakpoints | n as extreme | perm p | min attainable p |
| --- | --- | --- | --- | --- | --- |
| mean_cov | 2.259 | 7. | 3. | 0.4286 | 0.1429 |
| top5_share | 0.307 | 7. | 7. | 1. | 0.1429 |

**Supplementary Table 60.** California annual series, 2013-14 to 2022-23.

| year | n | mean cov | gini susc | top5 share | % below95 | t | post | t post |
| --- | --- | --- | --- | --- | --- | --- | --- | --- |
| 2013-2014 | 6,982 | 92.404 | 0.5806 | 27.566 | 46.878 | 0 | 0 | 0 |
| 2014-2015 | 7,032 | 92.625 | 0.6126 | 32.418 | 43.231 | 1. | 0 | 0 |
| 2015-2016 | 7,137 | 94.582 | 0.6311 | 32.785 | 33.852 | 2. | 0 | 0 |
| 2016-2017 | 6,469 | 96.605 | 0.5058 | 32.941 | 12.258 | 3. | 1. | 1. |
| 2017-2018 | 6,493 | 96.152 | 0.5534 | 39.988 | 13.384 | 4. | 1. | 2. |
| 2018-2019 | 6,524 | 95.844 | 0.5663 | 42.323 | 13.259 | 5. | 1. | 3. |
| 2019-2020 | 6,565 | 95.751 | 0.5809 | 45.024 | 12.963 | 6. | 1. | 4. |
| 2020-2021 | 6,344 | 94.411 | 0.5521 | 38.052 | 23.66 | 7. | 1. | 5. |
| 2021-2022 | 6,515 | 95.381 | 0.5761 | 45.707 | 12.632 | 8. | 1. | 6. |
| 2022-2023 | 6,602 | 95.537 | 0.6026 | 49.407 | 10.603 | 9. | 1. | 7. |

**Supplementary Table 61.** Interrupted time series estimates.

| outcome | term | estimate | lo | hi | p | dw |
| --- | --- | --- | --- | --- | --- | --- |
| mean_cov | pre_trend_per_yr | 1.089 | 0.5885 | 1.59 | 0.0018 | 2.234 |
| mean_cov | level_change | 2.259 | 1.579 | 2.938 | 0.000185 | 2.234 |
| mean_cov | trend_change_per_yr | -1.31 | -1.868 | -0.7517 | 0.0012 | 2.234 |
| gini_susc | pre_trend_per_yr | 0.0253 | 0.0214 | 0.0292 | 3.94×10<sup>-6</sup> | 2.018 |
| gini_susc | level_change | -0.1169 | -0.1485 | -0.0852 | 0.000103 | 2.018 |
| gini_susc | trend_change_per_yr | -0.0138 | -0.0222 | -0.0054 | 0.0071 | 2.018 |
| top5_share | pre_trend_per_yr | 2.61 | 1.317 | 3.903 | 0.0026 | 2.275 |
| top5_share | level_change | 0.307 | -4.082 | 4.696 | 0.8697 | 2.275 |
| top5_share | trend_change_per_yr | -0.5895 | -2.419 | 1.24 | 0.4604 | 2.275 |
| pct_below95 | pre_trend_per_yr | -6.513 | -8.166 | -4.86 | 7.13×10<sup>-5</sup> | 2.219 |
| pct_below95 | level_change | -21.26 | -25.214 | -17.307 | 1.19×10<sup>-5</sup> | 2.219 |
| pct_below95 | trend_change_per_yr | 6.653 | 4.362 | 8.944 | 0.00039 | 2.219 |

**Supplementary Table 62.**
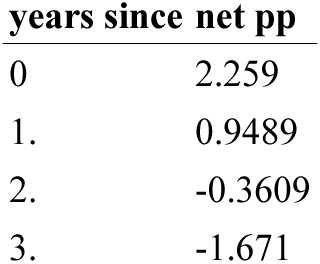
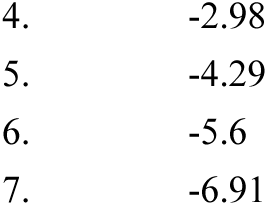
SB277 net trajectory against the pre-trend.

**Supplementary Table 63.** Placebo test at the SB277 breakpoint.

| state | outcome | n pre | level | lo | hi | p |
| --- | --- | --- | --- | --- | --- | --- |
| MA | mean_cov | 3. | 0.4668 | -0.552 | 1.486 | 0.3216 |
| MA | top5_share | 3. | -4.09 | -11.681 | 3.501 | 0.2492 |
| MN | mean_cov | 3. | 1.514 | 0.0817 | 2.947 | 0.0407 |
| MN | top5_share | 3. | -3.239 | -11.387 | 4.91 | 0.3862 |
| HI | mean_cov | 3. | 6.679 | -0.3088 | 13.667 | 0.0579 |
| HI | top5_share | 3. | -12.799 | -16.247 | -9.35 | 0.0001 |

**Supplementary Table 64.**
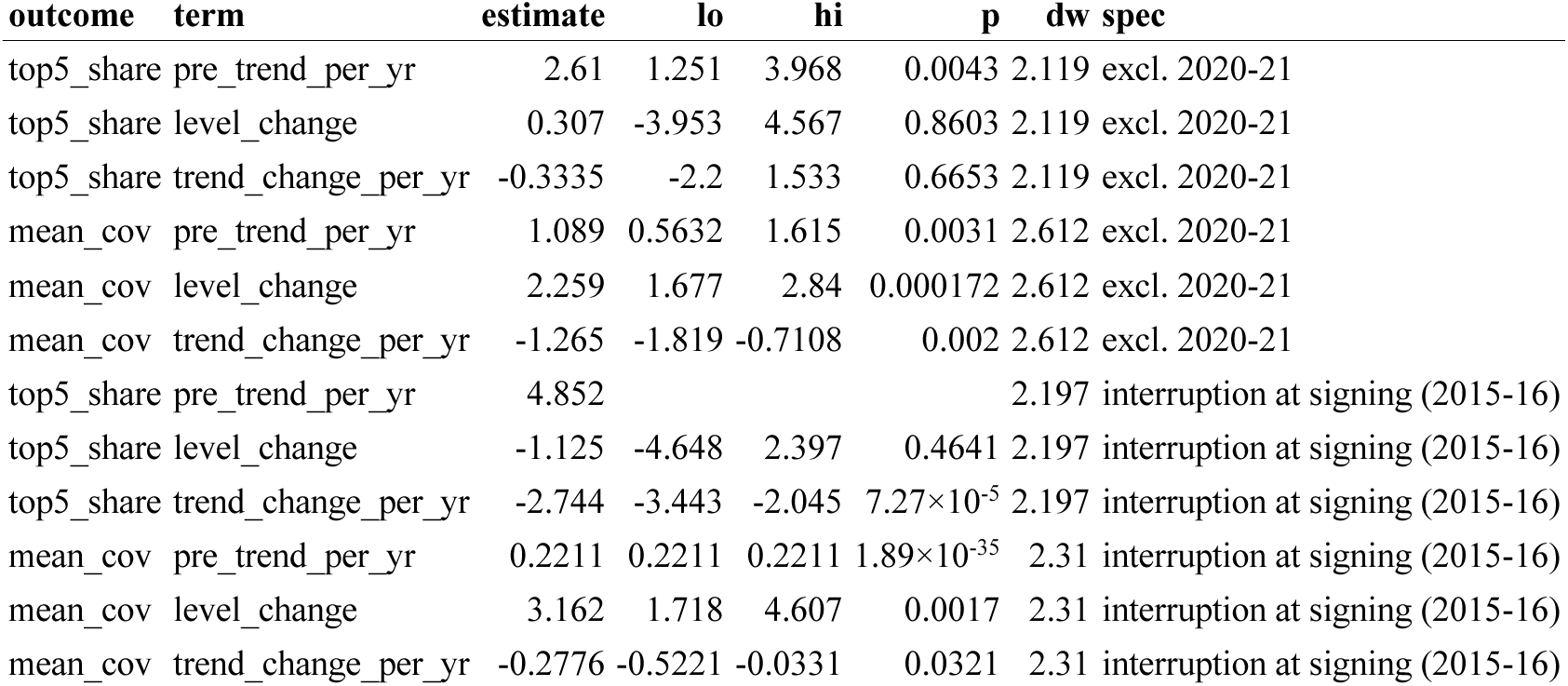
Interrupted time series robustness.

**Supplementary Table 65.**
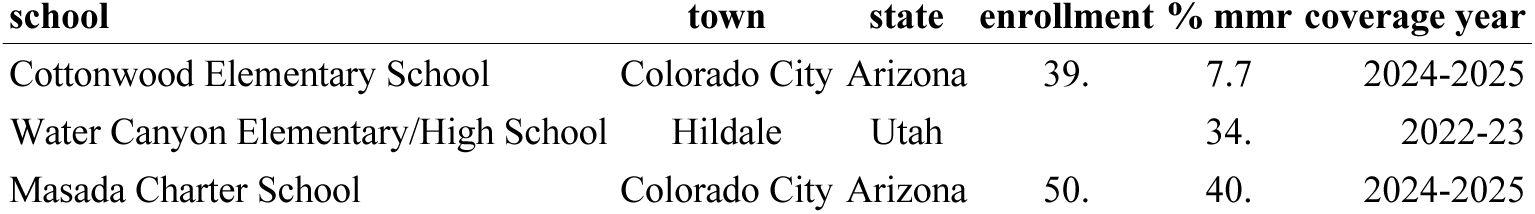
Exposed schools in the Short Creek community across the Utah-Arizona line.

**Supplementary Table 66.** State-level replication of the county model.

| panel | n counties | n affected | sd per pp | sd p | mean per pp | mean p | auc sd | auc size | total susceptibles | outcome |
| --- | --- | --- | --- | --- | --- | --- | --- | --- | --- | --- |
| Texas, school units | 137 | 36. | 0.3015 | 0.0115 | -0.0152 | 0.8511 | 0.6389 | 0.6144 | 21,884 | >= 1 case in the manuscript series |
| Texas, district units | 159 | 40. | 0.0991 | 0.1521 | -0.0297 | 0.664 | 0.6387 | 0.6486 | 1,217 | >= 1 case in the manuscript series |
| New Mexico, school units | 24. | 8. | 0.0936 | 0.7577 | -0.4652 | 0.2719 | 0.3281 | 0.8047 | 969 | >= 1 case in the manuscript series |

**Supplementary Table 67.** Texas outcome-threshold sensitivity.

| panel | n counties | n affected | sd per pp | sd p | mean per pp | mean p | auc sd | auc size | total susceptibles | outcome |
| --- | --- | --- | --- | --- | --- | --- | --- | --- | --- | --- |
| >= 1 cases | 137 | 36. | 0.3015 | 0.0115 | -0.0152 | 0.8511 | 0.6389 | 0.6144 | 21,884 | case-count threshold |
| >= 3 cases | 137 | 18. | 0.2323 | 0.066 | -0.0226 | 0.8176 | 0.6517 | 0.5803 | 21,884 | case-count threshold |
| >= 5 cases | 137 | 15. | 0.2967 | 0.0273 | -0.0621 | 0.557 | 0.6639 | 0.5311 | 21,884 | case-count threshold |
| >= 10 cases | 137 | 5. | 0.2384 | 0.2051 | 0.0808 | 0.5382 | 0.7197 | 0.5545 | 21,884 | case-count threshold |
| >= 20 cases | 137 | 5. | 0.2384 | 0.2051 | 0.0808 | 0.5382 | 0.7197 | 0.5545 | 21,884 | case-count threshold |
| >= 50 cases | 137 | 4. | 0.2402 | 0.2395 | 0.1188 | 0.393 | 0.7331 | 0.5545 | 21,884 | case-count threshold |

**Supplementary Table 68.** California county clustering model.

| n counties | n affected | sd per pp | sd p | mean per pp | mean p | enrol coef | enrol p | auc sd | auc mean | auc enrol | note |
| --- | --- | --- | --- | --- | --- | --- | --- | --- | --- | --- | --- |
| 53. | 16. | -0.0987 | 0.4136 | -0.0443 | 0.832 | 1.34 | 0.0041 | 0.4155 | 0.4172 | 0.799 |  |

**Note:** outcome: >= 1 case in the manuscript county series; coverage is 2022-23, more than two years before the 2025-26 cases

**Supplementary Table 69.**
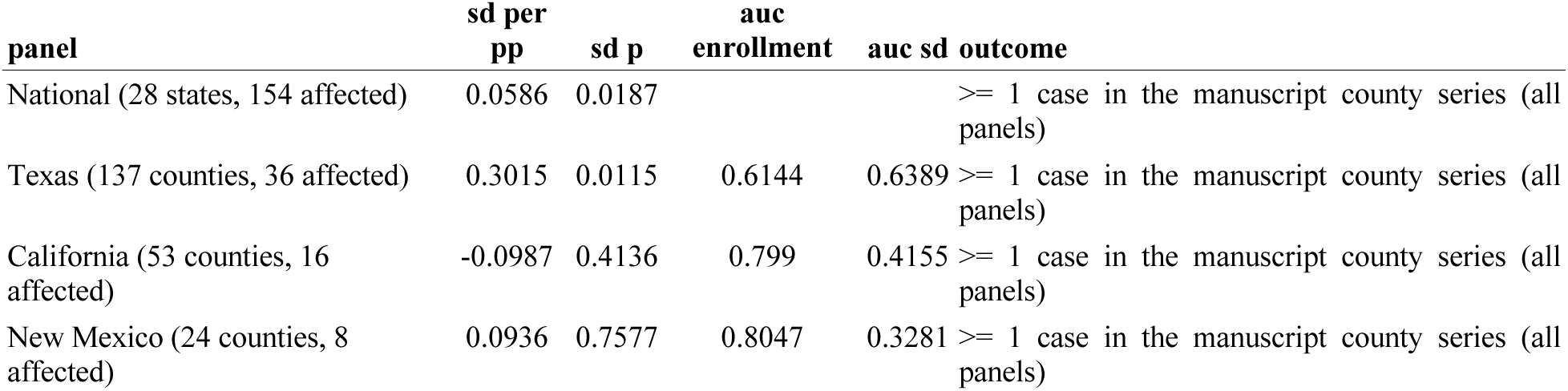
County clustering across four panels on one outcome definition.

**Supplementary Table 70.** Texas effect by coverage year.

| year | n<br>counties | n<br>affected | sd per<br>pp | sd p | mean<br>per pp | mean p | auc sd | auc<br>size | total<br>suscept<br>ibles | outcome | is pre<br>outbrea<br>k | bonferr<br>oni<br>thresho<br>ld | survive<br>s<br>bonferr<br>oni |
| --- | --- | --- | --- | --- | --- | --- | --- | --- | --- | --- | --- | --- | --- |
| 2019-2020 | 135 | 36. | 0.2549 | 0.1182 | 0.0951 | 0.4156 | 0.5522 | 0.6571 | 10,302 | >= 1 case in<br>the<br>manuscript<br>series | FALSE | 0.0083 | FALSE |
| 2020-2021 | 136 | 35. | 0.1596 | 0.1107 | 0.1301 | 0.1688 | 0.6484 | 0.6666 | 16,789 | >= 1 case in<br>the<br>manuscript<br>series | FALSE | 0.0083 | FALSE |
| 2021-2022 | 137 | 35. | 0.1387 | 0.1173 | -0.0342 | 0.7104 | 0.5978 | 0.6738 | 19,880 | >= 1 case in<br>the<br>manuscript<br>series | FALSE | 0.0083 | FALSE |
| 2022-2023 | 139 | 35. | 0.1505 | 0.1974 | -0.0702 | 0.465 | 0.5841 | 0.6459 | 19,138 | >= 1 case in<br>the<br>manuscript<br>series | FALSE | 0.0083 | FALSE |
| 2023-2024 | 147 | 40. | 0.0956 | 0.3385 | -0.042 | 0.5155 | 0.5341 | 0.6257 | 19,896 | >= 1 case in<br>the<br>manuscript<br>series | FALSE | 0.0083 | FALSE |
| 2024-2025 | 137 | 36. | 0.3015 | 0.0115 | -0.0152 | 0.8511 | 0.6389 | 0.6144 | 21,884 | >= 1 case in<br>the<br>manuscript<br>series | TRUE | 0.0083 | FALSE |

**Supplementary Table 71.**
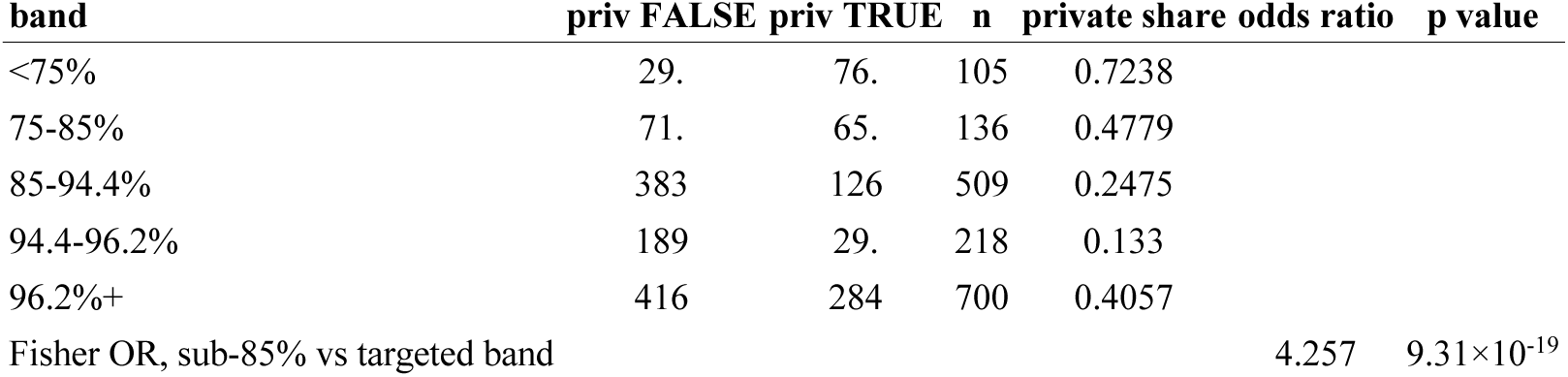
Texas private share by coverage band.

| band | priv FALSE | priv TRUE | n | private share | odds ratio | p value |
| --- | --- | --- | --- | --- | --- | --- |
| <75% | 29. | 76. | 105 | 0.7238 |  |  |
| 75-85% | 71. | 65. | 136 | 0.4779 |  |  |
| 85-94.4% | 383 | 126 | 509 | 0.2475 |  |  |
| 94.4-96.2% | 189 | 29. | 218 | 0.133 |  |  |
| 96.2%+ | 416 | 284 | 700 | 0.4057 |  |  |
| Fisher OR, sub-85% vs targeted band |  |  |  |  | 4.257 | 9.31×10<sup>-19</sup> |

**Supplementary Table 72.** South Carolina missingness by group.

| group | n | exposed | mean cov post |
| --- | --- | --- | --- |
| retained | 424 | 30. | 0.9067 |
| dropped | 20. | 1. | 0.8201 |

**Supplementary Table 73.** Reconstruction of the South Carolina exposure register.

| quantity | value |
| --- | --- |
| settings in the register | 34. |
| post-secondary (excluded) | 2. |
| outside the school reporting frame (excluded) | 1. |
| K-12 reporting units | 31. |
| already flagged in the deposited panel | 30. |
| added by reconciliation | 1. |
| Upstate flags after reconciliation | 31. |

**Supplementary Table 74.** Exposure-coverage linkages considered for an independent risk-scaling estimate.

| state | outbreak | record type | schools | exposed | denominator | usable for $\alpha$ | reason |
| --- | --- | --- | --- | --- | --- | --- | --- |
| South Carolina | Upstate 2025-26 | investigation register | 424 | 30. | yes | yes | school-by-school exposure list in twice-weekly departmental updates; statewide coverage file |
| Washington | Clark County 2018-19 | investigation register | 131 | 12. | yes | yes | county school exposure list with a same-release comparison set |
| Colorado | 2024-25 | visit list | 126 | 3. | yes | no | comparison set present but three exposed schools cannot support a coefficient |
| Utah | 2025-26 | visit list | 662 | 20. | yes | no | visit list, not an investigation register; kindergarten coverage panel obtained August 2026 supplies the comparison set and, in its 11 August release, the enrollment denominator, but no register |
| Arizona | Mohave County 2025-26 | visit list |  | 2. | no | no | two named schools; county roster served only through an interactive query tool, no bulk file |
| New York | Rockland 2018-19 | residence-based exclusion |  | 7. | no | no | exclusion ordered by ZIP code irrespective of case linkage; survey reports percentages without enrollment |
| New Mexico | 2025 | no school transmission |  | 0 | n/a | no | median case age 20 years; no school or childcare outbreaks reported |

**Supplementary Table 75.** Kernel spatial scope diagnostics.

| b | scale km | cv g | rho |
| --- | --- | --- | --- |
| 0.001 | 1,000 | 0.0596 | 0.9843 |
| 0.002 | 500 | 0.1078 | 0.9529 |
| 0.005 | 200 | 0.2238 | 0.841 |
| 0.01 | 100 | 0.3742 | 0.6665 |
| 0.02 | 50. | 0.5476 | 0.4935 |
| 0.05 | 20. | 0.7282 | 0.371 |
| 0.1 | 10. | 0.7793 | 0.3394 |

**Supplementary Table 76.** Cost factorial: conditional net benefit by per-case cost, dose price and budget.

| **budget fraction** | **doses** | **cost per case** | **dose price** | **infections averted optimal** | **infections averted lcf** | **gap infections** | **gap value M** | **dose cost M** | **conditional net benefit M** | **cost per infection averted** |
| --- | --- | --- | --- | --- | --- | --- | --- | --- | --- | --- |
| 0.1 | 30,920 | 6,973 | 30. | 13,289 | 4,283 | 9,005 | 62.798 | 0.9276 | 91.736 | 69.804 |
| 0.1 | 30,920 | 6,973 | 60. | 13,289 | 4,283 | 9,005 | 62.798 | 1.855 | 90.808 | 140 |
| 0.1 | 30,920 | 6,973 | 120 | 13,289 | 4,283 | 9,005 | 62.798 | 3.71 | 88.953 | 279 |
| 0.1 | 30,920 | 16,197 | 30. | 13,289 | 4,283 | 9,005 | 146 | 0.9276 | 214 | 69.804 |
| 0.1 | 30,920 | 16,197 | 60. | 13,289 | 4,283 | 9,005 | 146 | 1.855 | 213 | 140 |
| 0.1 | 30,920 | 16,197 | 120 | 13,289 | 4,283 | 9,005 | 146 | 3.71 | 212 | 279 |
| 0.1 | 30,920 | 43,203 | 30. | 13,289 | 4,283 | 9,005 | 389 | 0.9276 | 573 | 69.804 |
| 0.1 | 30,920 | 43,203 | 60. | 13,289 | 4,283 | 9,005 | 389 | 1.855 | 572 | 140 |
| 0.1 | 30,920 | 43,203 | 120 | 13,289 | 4,283 | 9,005 | 389 | 3.71 | 570 | 279 |
| 0.1 | 30,920 | 104,629 | 30. | 13,289 | 4,283 | 9,005 | 942 | 0.9276 | 1,389 | 69.804 |
| 0.1 | 30,920 | 104,629 | 60. | 13,289 | 4,283 | 9,005 | 942 | 1.855 | 1,389 | 140 |
| 0.1 | 30,920 | 104,629 | 120 | 13,289 | 4,283 | 9,005 | 942 | 3.71 | 1,387 | 279 |
| 0.1 | 30,920 | 243,614 | 30. | 13,289 | 4,283 | 9,005 | 2,193 | 0.9276 | 3,236 | 69.804 |
| 0.1 | 30,920 | 243,614 | 60. | 13,289 | 4,283 | 9,005 | 2,193 | 1.855 | 3,235 | 140 |
| 0.1 | 30,920 | 243,614 | 120 | 13,289 | 4,283 | 9,005 | 2,193 | 3.71 | 3,234 | 279 |
| 0.25 | 77,292 | 6,973 | 30. | 29,479 | 15,366 | 14,112 | 98.409 | 2.319 | 203 | 78.658 |
| 0.25 | 77,292 | 6,973 | 60. | 29,479 | 15,366 | 14,112 | 98.409 | 4.638 | 201 | 157 |
| 0.25 | 77,292 | 6,973 | 120 | 29,479 | 15,366 | 14,112 | 98.409 | 9.275 | 196 | 315 |
| 0.25 | 77,292 | 16,197 | 30. | 29,479 | 15,366 | 14,112 | 229 | 2.319 | 475 | 78.658 |
| 0.25 | 77,292 | 16,197 | 60. | 29,479 | 15,366 | 14,112 | 229 | 4.638 | 473 | 157 |
| 0.25 | 77,292 | 16,197 | 120 | 29,479 | 15,366 | 14,112 | 229 | 9.275 | 468 | 315 |
| 0.25 | 77,292 | 43,203 | 30. | 29,479 | 15,366 | 14,112 | 609 | 2.319 | 1,271 | 78.658 |
| 0.25 | 77,292 | 43,203 | 60. | 29,479 | 15,366 | 14,112 | 609 | 4.638 | 1,269 | 157 |
| 0.25 | 77,292 | 43,203 | 120 | 29,479 | 15,366 | 14,112 | 609 | 9.275 | 1,264 | 315 |
| 0.25 | 77,292 | 104,629 | 30. | 29,479 | 15,366 | 14,112 | 1,476 | 2.319 | 3,082 | 78.658 |
| 0.25 | 77,292 | 104,629 | 60. | 29,479 | 15,366 | 14,112 | 1,476 | 4.638 | 3,080 | 157 |
| 0.25 | 77,292 | 104,629 | 120 | 29,479 | 15,366 | 14,112 | 1,476 | 9.275 | 3,075 | 315 |
| 0.25 | 77,292 | 243,614 | 30. | 29,479 | 15,366 | 14,112 | 3,438 | 2.319 | 7,179 | 78.658 |
| 0.25 | 77,292 | 243,614 | 60. | 29,479 | 15,366 | 14,112 | 3,438 | 4.638 | 7,177 | 157 |
| 0.25 | 77,292 | 243,614 | 120 | 29,479 | 15,366 | 14,112 | 3,438 | 9.275 | 7,172 | 315 |

\*First 30 of 45 rows; the complete table is TableS63\_cost\_factorial.csv in the replication package in the GitHub repository<sup>14</sup>.

**Supplementary Table 77.** Data sources and provenance.

| source | provider | identifier | years | terms |
| --- | --- | --- | --- | --- |
| State kindergarten MMR coverage (33 states) | State health departments, compiled | Zenodo 10.5281/zenodo.19430274, .20174071 | 2013-14 to 2024-25 | CC-BY (Zenodo) |
| School-level $R_v$ estimates (national) | Chen & Bento, Nature Medicine 2026 | Zenodo 10.5281/zenodo.20260835 | 2017-18 to 2024-25 | CC-BY (Zenodo) |
| County-day measles case series | Johns Hopkins measles tracker, compiled | Zenodo 10.5281/zenodo.20260494 | 2025-01 to 2026-03 | CC-BY (Zenodo) |
| California kindergarten coverage, 2014-15 | California Department of Public Health | CDPH kindergarten immunization assessment | 2014-15 | public record |
| County and state boundaries | US Census Bureau TIGER/Line 2023 | cb_2023_us_{county,state}_20m | 2023 | public domain |

**Supplementary Table 78.**
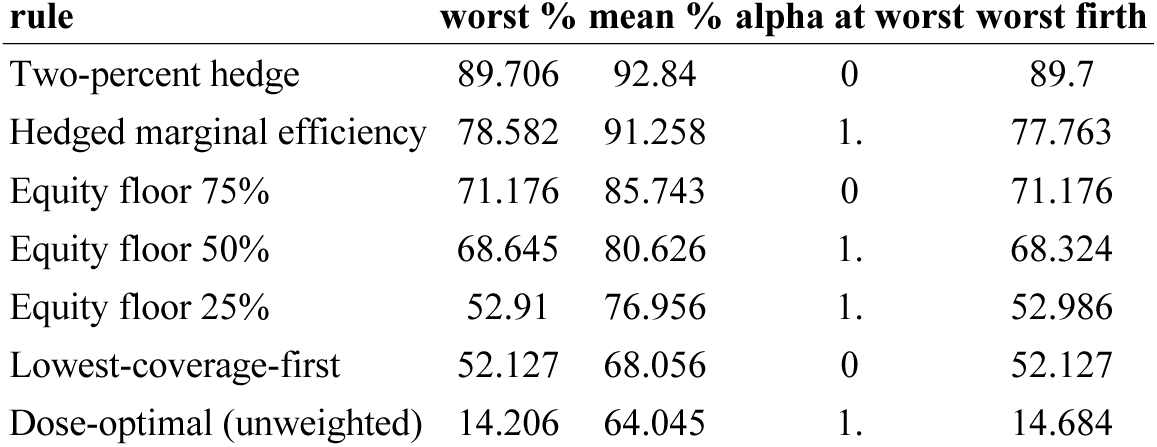
Worst-case performance of each candidate rule.

**Supplementary Table 79.** Persistence of the school coverage ranking by lag.

| lag | n_comparisons | mean_n_schools | spearman_rho | worst_decile_retention |
| --- | --- | --- | --- | --- |
| 1 | 11 | 10,149 | 0.5683 | 0.5209 |
| 2 | 10 | 9,161 | 0.5057 | 0.4663 |
| 3 | 9 | 8,337 | 0.4657 | 0.4264 |
| 4 | 8 | 7,335 | 0.444 | 0.4062 |
| 5 | 7 | 6,220 | 0.4307 | 0.3911 |
| 6 | 6 | 4,872 | 0.3998 | 0.3708 |
| 7 | 5 | 3,653 | 0.3761 | 0.3565 |
| 8 | 4 | 3,262 | 0.3529 | 0.3512 |
| 9 | 3 | 2,799 | 0.3115 | 0.2979 |
| 10 | 2 | 2,360 | 0.2266 | 0.2659 |
| 11 | 1 | 2,090 | 0.3176 | 0.3349 |

**Supplementary Table 80.** One-year persistence of the school coverage ranking by state.

| state | n_comparisons | mean_n_schools | spearman_rho | worst_decile_retention |
| --- | --- | --- | --- | --- |
| NC | 4 | 1,264 | 0.3452 | 0.3995 |
| TN | 1 | 352 | 0.4135 | 0.4857 |
| MA | 11 | 744 | 0.4207 | 0.5134 |
| VA | 10 | 1,005 | 0.4278 | 0.4012 |
| IN | 1 | 934 | 0.4451 | 0.3978 |
| TX | 5 | 952 | 0.4506 | 0.4097 |
| MN | 11 | 803 | 0.4815 | 0.5233 |
| IL | 10 | 188 | 0.4909 | 0.4058 |
| HI | 8 | 267 | 0.4965 | 0.5444 |
| CO | 7 | 760 | 0.4987 | 0.3848 |
| WA | 6 | 1,202 | 0.5133 | 0.4526 |
| MI | 1 | 1,678 | 0.5225 | 0.5179 |
| OK | 6 | 538 | 0.5288 | 0.4597 |
| KY | 6 | 597 | 0.564 | 0.4493 |
| CA | 6 | 5,716 | 0.5767 | 0.444 |
| AZ | 9 | 989 | 0.5905 | 0.592 |
| IA | 4 | 1,076 | 0.6674 | 0.6218 |

**Supplementary Table 81.** . Panel enrollment against NCES Common Core of Data grade-0 counts.

| state | yr | panel_schools | panel_N | ccd_schools | ccd_N | ratio_N | ratio_schools | note |
| --- | --- | --- | --- | --- | --- | --- | --- | --- |
| AR | 2,024 | 463 | 34,657 | 476 | 33,926 | 1.022 | 0.9727 | reporting-quality exclusion, not in analyses |
| AZ | 2,015 | 1,101 | 80,392 | 1,178 | 79,203 | 1.015 | 0.9346 |  |
| AZ | 2,016 | 1,101 | 80,392 | 1,203 | 79,752 | 1.008 | 0.9152 |  |
| AZ | 2,017 | 1,082 | 78,619 | 1,212 | 79,488 | 0.9891 | 0.8927 |  |
| AZ | 2,018 | 1,066 | 77,605 | 1,211 | 79,865 | 0.9717 | 0.8803 |  |
| AZ | 2,019 | 1,071 | 79,856 | 1,213 | 81,494 | 0.9799 | 0.8829 |  |
| AZ | 2,020 | 1,063 | 68,828 | 1,232 | 72,904 | 0.9441 | 0.8628 |  |
| AZ | 2,021 | 1,112 | 78,147 | 1,273 | 79,280 | 0.9857 | 0.8735 |  |
| AZ | 2,022 | 1,106 | 76,309 | 1,254 | 76,147 | 1.002 | 0.882 |  |
| AZ | 2,023 | 1,098 | 72,040 | 1,246 | 72,320 | 0.9961 | 0.8812 |  |
| AZ | 2,024 | 1,108 | 71,070 | 1,238 | 70,774 | 1.004 | 0.895 |  |
| CA | 2,017 | 5,918 | 513,633 | 6,116 | 531,092 | 0.9671 | 0.9676 |  |
| CA | 2,018 | 5,850 | 496,377 | 6,141 | 525,369 | 0.9448 | 0.9526 |  |
| CA | 2,019 | 6,556 | 540,238 | 6,155 | 522,721 | 1.034 | 1.065 |  |
| CA | 2,020 | 6,016 | 456,752 | 6,174 | 458,921 | 0.9953 | 0.9744 |  |
| CA | 2,021 | 6,502 | 489,987 | 6,251 | 469,303 | 1.044 | 1.04 |  |
| CA | 2,022 | 6,211 | 508,302 | 6,227 | 495,430 | 1.026 | 0.9974 |  |
| CA | 2,023 | 6,013 | 523,647 | 6,190 | 521,914 | 1.003 | 0.9714 |  |
| CO | 2,017 | 1,166 | 65,041 | 1,085 | 63,574 | 1.023 | 1.075 |  |
| CO | 2,018 | 1,123 | 63,693 | 1,096 | 63,409 | 1.004 | 1.025 |  |
| CO | 2,019 | 1,177 | 67,151 | 1,096 | 64,009 | 1.049 | 1.074 |  |
| CO | 2,020 | 1,217 | 61,129 | 1,101 | 58,186 | 1.051 | 1.105 |  |
| CO | 2,021 | 1,210 | 64,597 | 1,110 | 61,531 | 1.05 | 1.09 |  |
| CO | 2,022 | 1,215 | 63,193 | 1,099 | 58,675 | 1.077 | 1.106 |  |
| CO | 2,023 | 1,167 | 61,168 | 1,078 | 57,265 | 1.068 | 1.083 |  |
| CO | 2,024 | 1,132 | 59,437 | 1,076 | 57,210 | 1.039 | 1.052 |  |
| CT | 2,024 | 450 | 24,646 | 552 | 30,011 | 0.8212 | 0.8152 | reporting-quality exclusion, not in analyses |
| DC | 2,024 | 153 | 7,750 | 138 | 7,285 | 1.064 | 1.109 |  |
| HI | 2,013 | 400 | 217,053 | 204 | 17,561 | 12.36 | 1.961 | whole-school denominator (Methods) |
| HI | 2,014 | 402 | 208,176 | 205 | 11,661 | 17.852 | 1.961 | whole-school denominator (Methods) |

\*Showing the first 30 of 164 rows; the full table is deposited as TableS81\_ccd\_enrollment\_validation.csv in the replication package in the GitHub repository<sup>14</sup>.

**Supplementary Table 82.**
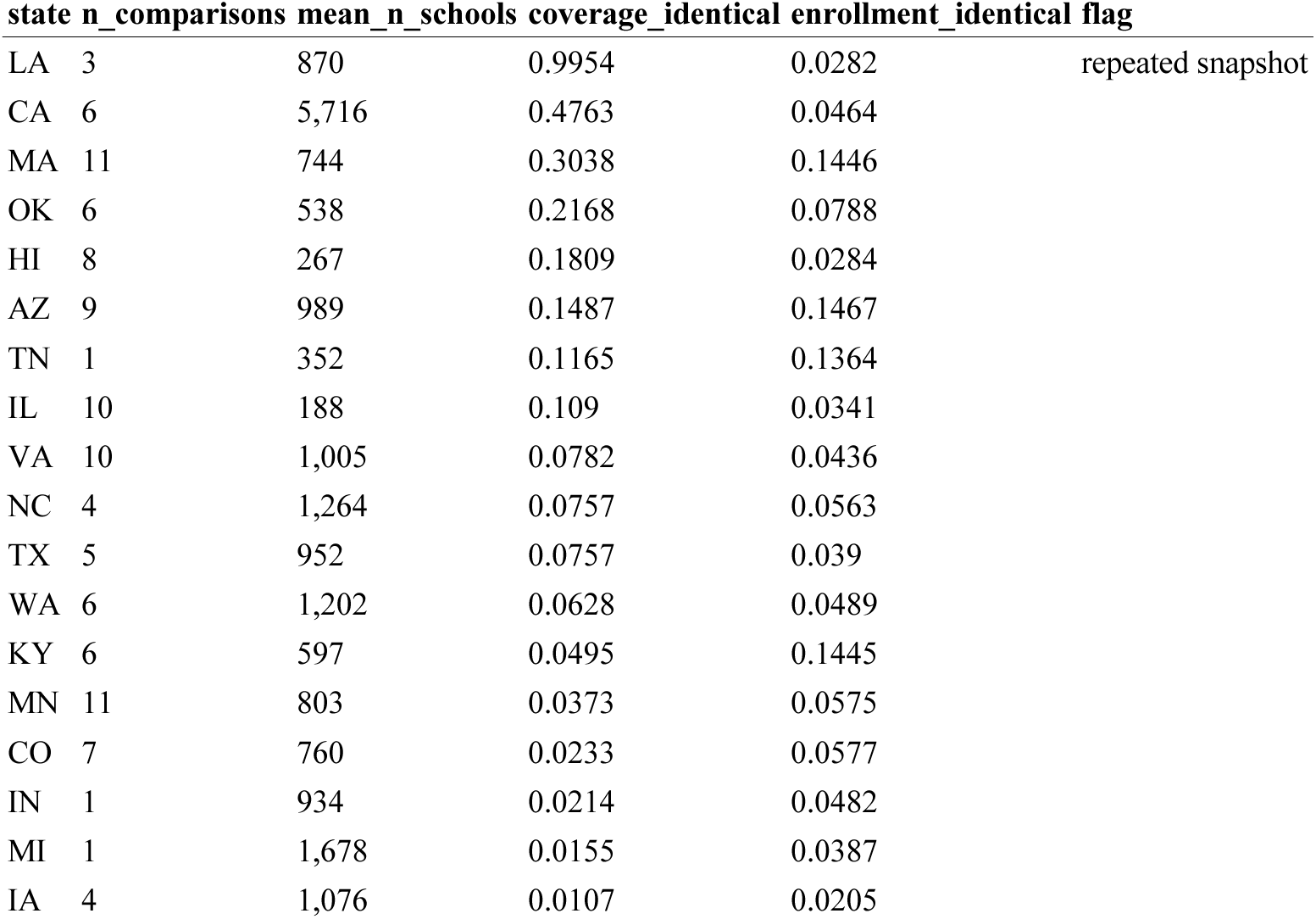
Cross-year repetition audit of the coverage field.

| state | n_comparisons | mean_n_schools | coverage_identical | enrollment_identical | flag |
| --- | --- | --- | --- | --- | --- |
| LA | 3 | 870 | 0.9954 | 0.0282 | repeated snapshot |
| CA | 6 | 5,716 | 0.4763 | 0.0464 |  |
| MA | 11 | 744 | 0.3038 | 0.1446 |  |
| OK | 6 | 538 | 0.2168 | 0.0788 |  |
| HI | 8 | 267 | 0.1809 | 0.0284 |  |
| AZ | 9 | 989 | 0.1487 | 0.1467 |  |
| TN | 1 | 352 | 0.1165 | 0.1364 |  |
| IL | 10 | 188 | 0.109 | 0.0341 |  |
| VA | 10 | 1,005 | 0.0782 | 0.0436 |  |
| NC | 4 | 1,264 | 0.0757 | 0.0563 |  |
| TX | 5 | 952 | 0.0757 | 0.039 |  |
| WA | 6 | 1,202 | 0.0628 | 0.0489 |  |
| KY | 6 | 597 | 0.0495 | 0.1445 |  |
| MN | 11 | 803 | 0.0373 | 0.0575 |  |
| CO | 7 | 760 | 0.0233 | 0.0577 |  |
| IN | 1 | 934 | 0.0214 | 0.0482 |  |
| MI | 1 | 1,678 | 0.0155 | 0.0387 |  |
| IA | 4 | 1,076 | 0.0107 | 0.0205 |  |

